# Immunogenicity and Safety of Biological E’s 14-Valent Pneumococcal Conjugate Vaccine (PNEUBEVAX 14^®^) Administered in a 2p+1 Schedule to Healthy Infants: A Multicenter, Randomized, Active Controlled, Single-Blind, Phase III Trial

**DOI:** 10.1101/2025.08.25.25334360

**Authors:** Subhash Thuluva, Ramesh V. Matur, Subbareddy Gunneri, Siddalingaiah Ningaiah, Vijay Yerroju, Rammohan Reddy Mogulla, Chirag Dhar, Kamal Thammireddy, Anand Kawade, Pradeep N, Jog Pramod, Manish Narang, Chakravarthy B.S., Prashanth M.V.

## Abstract

**Background:** Pneumococcal conjugate vaccines (PCVs) have markedly reduced childhood pneumococcal diseases, yet serotype replacement and regional heterogeneity remain important challenges. The World Health Organization recommends either a 3p + 0 or 2p + 1 schedule for PCV immunization programmes. BE-PCV14, a 14-valent vaccine, has previously been shown to be non-inferior to PCV13 in a 3p + 0 regimen. In this study, we descriptively compared the immunogenicity and safety of BE-PCV14 and PCV13 in a 2p + 1 schedule in Indian infants.

**Methods:** In this randomized, single-blind, multicenter trial, 400 PCV-naïve infants (6–8 weeks old) were randomized 1:1 to receive either BE-PCV14 or PCV13; at 6 and 14 weeks, with a booster at 9 months. Serum IgGs against 14 vaccine serotypes plus cross-protective 6A were measured at post primary (28 days post dose 2), pre booster (at 9 months) and post booster (30 days post dose 3) time points. The primary endpoint was the proportion achieving IgG ≥ 0.35 µg/mL (seroconversion rate) for the 12 serotypes common to both vaccines at post primary, pre booster and post booster time points. Solicited local and systemic reactions were recorded for 7 days after each dose; unsolicited, medically attended, and serious adverse events (SAEs) were captured throughout.

**Results:** Between May 2023 to July 2024, 400 participants were enrolled of which 380 (95%) completed the study. Post primary seroconversion rates in the BE-PCV14 arm for common serotypes ranged from 72.6% (serotype 3) to 100% (14, 19F); PCV13 rates ranged from 71.6% to 100% (3 to 14, 19F, 19A). Post booster rates were 87.6–100% for BE-PCV14 and 85.0–100% for PCV13. BE-PCV14 elicited high responses against the two additional serotypes (22F: 96.8%; 33F: 95.3%) and cross-protective 6A (93.0%). Seroconversion rates and Geometric mean concentrations were similar between groups. Most adverse events were mild or moderate; two unrelated SAEs occurred in the BE-PCV14 arm.

**Interpretation:** Administered in a 2p + 1 schedule, BE-PCV14 was highly immunogenic, well tolerated, and comparable to PCV13 while broadening serotype coverage, supporting its inclusion in routine infant immunisation programmes.

Clinical Trials Registry of India Number: CTRI/2022/11/047366

## 1. Introduction

*Streptococcus pneumoniae* remains a major cause of pneumonia, meningitis and bacteraemia in children below 5 years, with low- and middle-income countries (LMICs) accounting for most of the estimated 300,000 annual deaths.^1, 2^ The introduction of 7-, 10- and 13-valent pneumococcal conjugate vaccines (PCVs) into infant programmes have drastically cut vaccine-type invasive pneumococcal disease (IPD) and pneumonia.^3, 4, 5^ However, this success was accompanied by serotype replacement, with non-vaccine serotypes such as 22F and 33F emerging as significant causes of remaining infections in post-PCV13 settings across multiple regions.^6, 7^ Indian hospital data likewise list 22F and 33F among the ten commonest non vaccine serotypes causing paediatric IPD.^8, 9^ The inclusion of these serotypes in newer PCVs, such as Biological E’s PNEUBEVAX 14^®^(BE-PCV14), PCV15 and PCV20, is aimed at addressing this evolving seroprevalence and preventing diseases related to these additional serotypes.^10, 11^

The World Health Organization recommends either a 3-dose primary series (**3p+0)** or 2 dose primary series and a booster dose (**2p+1)** for PCVs. The 2p+1 schedule consist of primary doses at 6 weeks and 14 weeks followed by a booster dose at 9-15 months of age.^12^ While both schedules are highly effective, a 2p+1 regimen induces higher antibody concentrations in the second year of life that may enhance herd protection while requiring fewer infant visits.^13, 14^ More than 50 countries across the globe have now adopted the 2p+1 schedule in their routine vaccination programme.^15^ BE-PCV14 has undergone extensive clinical development with a phase I trial, a phase II trial, two phase III trials and a phase IV study demonstrating an acceptable safety profile. The two-phase III clinical trials evaluated BE-PCV14 in a 3p+0 schedule, with Pfizer’s PCV13 as a comparator. These studies demonstrated that BE-PCV14 was immunologically non-inferior to PCV13 with a comparable safety profile.^16, 17^ Here, we report the findings of a phase III study that evaluated the immunogenicity and safety of BE-PCV14 when administered to healthy infants in a 2p + 1 dosing regimen.

## 2. Materials and Methods

### 2.1. Study Design and participants

This single-blind, randomised, active controlled phase III trial evaluated the immunogenicity, safety and tolerability of BE-PCV14 (PNEUBEVAX 14^®^) administered in a 2p+1 schedule to healthy Indian infants. The study was conducted at seven sites in India between 15 May 2023 and 5 July 2024 (sites details included as supplementary data, Appendix 1). The protocol complied with the Declaration of Helsinki, Good Clinical Practice and national regulations, and was approved by the ethics committee of each study site. Written informed consent was obtained from parents or legal guardians before any study procedure.

Eligible infants were 6–8 weeks of age, pneumococcal-vaccine naïve and met all inclusion/exclusion criteria (supplementary data, Appendix 2). A total of 400 infants were enrolled.

### 2.2. Randomization and masking

At the screening visit (visit 1), all participants were randomly allocated 1:1 to receive either BE-PCV14 or PCV13 (Prevnar 13™, Pfizer). The randomization was conducted by using an interactive web-response system (IWRS) generated with PROC PLAN (SAS 9.4). The study was single-blind, with participant’s parents/legal representatives unaware of the assigned vaccine. The laboratory personnel involved in serology testing remained blinded to the treatment, and codes were used to link the participant and study (without any link to the treatment to the participant) to each sample.

### 2.3. Procedures

The study comprised five scheduled visits. On Day 0 (Visit 1), participants were screened, randomized, had baseline blood drawn, and received the first dose of either BE-PCV14 or PCV13. The second dose was administered on Day 56 (Visit 2). Post-primary immunogenicity samples were collected on Day 84 (Visit 3). At 9 months of age (Visit 4), pre-booster blood was drawn and a booster dose was administered. Final post-booster blood samples were obtained 30 days post booster (10 months of age) (Visit 5). Safety-follow-up data were recorded on days 56 and 84, and at the post booster time points. Each participant was followed until 30 days post booster with a permissible +7-day window for Visits 2, 3, 4 and 5 to facilitate scheduling compliance.

Each 0.5 mL dose in multi-human dose presentation of PNEUBEVAX 14^®^ (BE-PCV14) contained 3 µg of serotype 1 polysaccharide; 2.2 µg of each of serotypes 3, 4, 5, 7F, 9V, 14, 18C, 19A, 19F, 22F, 23F and 33F; and 4.4 µg of serotype 6B, conjugated to 20–50 µg CRM_197_ and adsorbed on ≤ 0.75 mg aluminium phosphate with 4 mg 2-phenoxyethanol. PCV13 contained 2.2 µg of each of serotypes 1, 3, 4, 5, 6A, 7F, 9V, 14, 18C, 19A, 19F, and 23F saccharides; 4.4 µg of 6B saccharide, conjugated to 34 µg CRM_197_ with 125 µg aluminium phosphate. Both the vaccines were administered intramuscularly in the anterolateral aspect of the thigh.

All the study participants had received birth dose of BCG, Hepatitis B and Oral Polio vaccines prior to enrolment. During the study, concomitant administration of all vaccines recommended in the Universal Immunization Programme (UIP) in the 6-10-14 weeks and 9 months schedule were permitted.

Approximately 5.0 mL of peripheral venous blood was collected by a trained phlebotomist at four time points: visit 1, 3, 4 and 5. Blood samples were centrifuged at approximately 3,500 rpm for 10–15 minutes as per lab manual. The separated serum was transferred into pre-labelled cryovials/serum separation (SS) containers using sterile micro-pipettes with precision. The serum sample containing cryovials were stored below −20 °C and temperature was monitored periodically till transferred to the central laboratory and sponsor facilities. Anti-pneumococcal capsular polysaccharide (PnCPS) IgG antibody concentration estimation against each of the 14 vaccine serotypes and 6A was performed at Biological E’s in-house laboratory as per the World Health Organization (WHO) reference enzyme linked immunosorbent (ELISA) assay^18^ with a minor modification. Cell wall polysaccharide (CWPS) multi was used instead of CWPS and 22F pneumococcal polysaccharide (PnCPS) for preadsorption to neutralize non-specific antibodies and other common contaminants present in the PnCPS coating antigens.

Following each vaccination, infants were observed on-site for a minimum of 60 minutes to monitor for any immediate adverse reactions and to provide medical management if required. During each vaccination visit, the parent(s) or legally acceptable representative (LAR) received a structured diary card to document any solicited local or systemic adverse events (AEs) occurring within the first 7 days post-vaccination. The list of solicited AEs is provided in Supplementary data, Appendix 3. In addition to solicited AEs, all unsolicited adverse events, serious adverse events (SAEs), and medically attended adverse events (MAAEs) were recorded throughout the study duration. The intensity of each adverse event was graded using established criteria, including the Common Terminology Criteria for Adverse Events (CTCAE), version 5.0, and the Division of AIDS (DAIDS) Table for Grading the Severity of Adult and Pediatric Adverse Events, version 2.0. For febrile events, severity was classified according to the Brighton Collaboration case definition scale. Causality assessment was performed by the study investigator, who determined the relationship between each reported event and the study vaccine.

### 2.4. Outcomes

The primary outcome of the study was proportion of infants with serotype-specific anti-PnCPS IgG ≥ 0.35 µg/mL (also referred to as seroconversion rate^19^) with their exact 2-sided 95% CIs at post primary, pre booster and post booster time points at day 84, month 9, and months 10 for each of the 12 serotypes common with PCV13. Other immunogenicity endpoints included geometric mean concentrations (GMCs) of anti-PnCPS IgG for each serotype with their exact 2-sided 95% CIs and the proportion of participants achieving ≥2 fold and ≥4 fold rise in anti-PnCPS IgG from baseline to post-primary vaccination, pre-booster and post-booster time periods. Secondary outcome was the safety, reactogenicity and tolerability of BE-PCV14 in comparison to PCV13, measured by incidence rates of local and systemic adverse events (AEs), medically attended adverse events (MAAEs) and serious adverse events (SAEs).

### 2.5. Sample Size determination

This randomized, phase III study was designed to descriptively compare the immunogenicity and safety of BE-PCV14 administered in a 2p + 1 schedule with that of Pfizer’s PCV13. Although a concurrent comparator arm was included, the trial was not formally powered to demonstrate statistical differences between the test and control arm. Noninferiority of BE-PCV14 had already been established in the earlier BECT051 phase III study.^16^ With 400 participants randomized evenly between the two vaccine groups, the design instead aimed to generate robust descriptive data to support clinical and programmatic decision-making. This number was based on established precedent from prior pivotal PCV trials and current regulatory guidance. ^20, 21^

### 2.6. Statistical Analyses

All demographic and baseline characteristics for each treatment group were summarized in all randomized participants defined as the intention-to-treat (ITT) population. All safety analyses were descriptive and performed for all those who received at least one dose of the vaccine (safety population). Demographics, and baseline characteristics were analyzed by summary statistics. For continuous variables, n, mean, standard deviation, median, and range (minimum and maximum) were presented. For categorical data, frequencies were computed. All reported adverse events during the entire study period were summarized by calculating frequencies and were listed per subject including severity and relationship to the vaccine (causality).

The number and percentage of participants with AEs were presented by system organ class (SOC) and preferred term (PT). All medically attended AEs (MAAEs) reported during the study were listed and analyzed for expectedness and causality. Two-sided 95% exact confidence intervals (CIs) were calculated for all the occurrence rates of reported AEs and SAEs during the study. All AEs were coded using the Medical Dictionary for Regulatory Activities (MedDRA™; version 25.1) coding dictionary, and concomitant medications were coded using the WHO Drug Dictionary, March 2022.

Immunogenicity was evaluated in the per-protocol (PP) population (participants who received all doses, had no protocol deviations and contributed valid immunogenicity data at all protocol defined timepoints) at day 84 (post primary), 9 months (pre booster) and at 10 months (post booster). Seroconversion rates and Geometric mean concentrations (GMCs) were calculated against each of the vaccine serotype along with their two-sided 95% CIs. Wilson score method, without continuity correction was used for seroconversion rates, for GMC calculations, natural log transformation was used, and the results were back-transformed. The proportion of subjects achieving ≥2-fold and ≥4-fold rise in anti-PnCPS IgG antibody concentrations from the baseline against each of the vaccine serotype was also calculated. The fold rise was calculated as the ratio of the post-vaccination concentration value to the pre-vaccination value against each of the vaccine serotypes along with their two-sided 95% CIs. Reverse cumulative distribution (RCD) curves of anti-PnCPS antibody concentrations by serotype for post primary and post booster timepoints were plotted for all common vaccine serotypes in both groups.

The immune response to serotypes 22F and 33F (not present in PCV13 group) after BE-PCV14, were assessed in terms of seroconversion rates and geometric mean concentrations in comparison with the lowest performing serotype in PCV13 group as a reference at respective time points.

## 3. Results

A total of 400 infants aged 6-8 weeks were screened and randomized to receive either BE-PCV14 (n = 200) or PCV13 (n = 200) study vaccines. Safety analyses were performed in all enrolled participants (n = 400), whereas immunogenicity assessment population (per-protocol population) included 380 (95%) participants (186 in the BE-PCV14 group and 194 in the PCV13 group) who completed all the study visits with data available for immunogenicity. Out of 20 participants who did not complete the study procedures, 14 migrated from the study area, 3 withdrew consent, 2 were lost to follow-up, and 1 took another vaccine as booster dose (study flowchart shown in figure 1). During the course of this study, no major protocol deviations were reported at any of the study sites. Few participants reported for their visits out of window period but these deviations were not found to be significant and were duly reported to ethics committees of the respective study sites.

**Figure 1:**
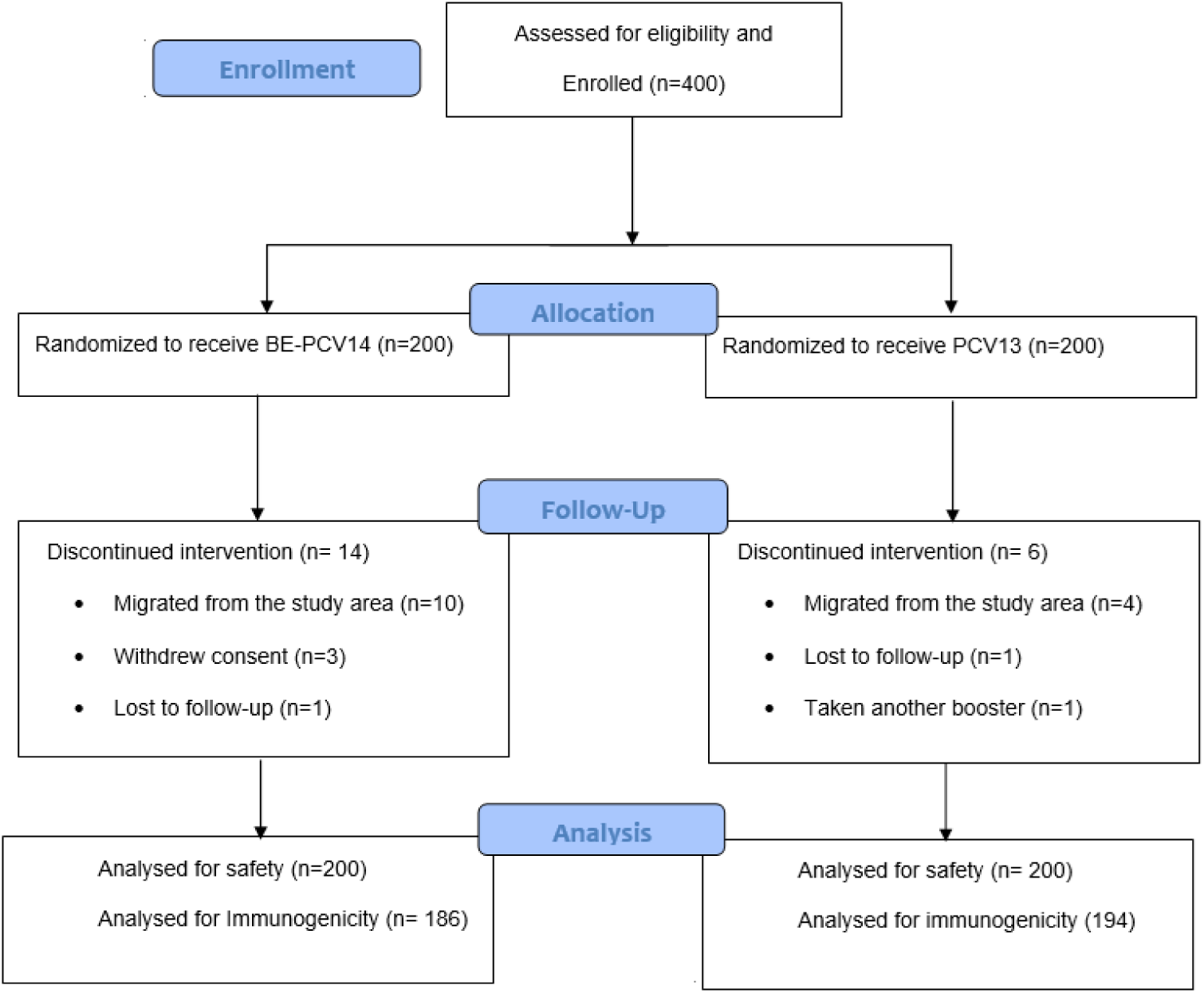
Subject disposition. *n*: number, PCV: Pneumococcal conjugate vaccine, BE-PCV14: Biological E’s 14 valent PCV, PCV13: Pfizer’s 13 valent PCV

Baseline demographic and anthropometric characteristics of the intention-to-treat cohort are summarized in Table 1. Both the BE-PCV14 and PCV13 groups had a mean age at enrolment of 48 days, with comparable mean length (≈ 53 cm) and mean weight (≈ 4.2 kg). Female participants represented 48.0% of the BE-PCV14 arm and 43.5% of the PCV13 arm. Overall, no clinically meaningful imbalances were noted between treatment groups. Details of prior routine immunizations and concomitant medication use are provided in Supplementary data, Appendices 4 and 5. Infants of both the groups have received DTwP-HepB-Hib, Rotavirus vaccine and IPV as part of routine immunization schedule at visit 1, 2 and 3.

**Table 1:**
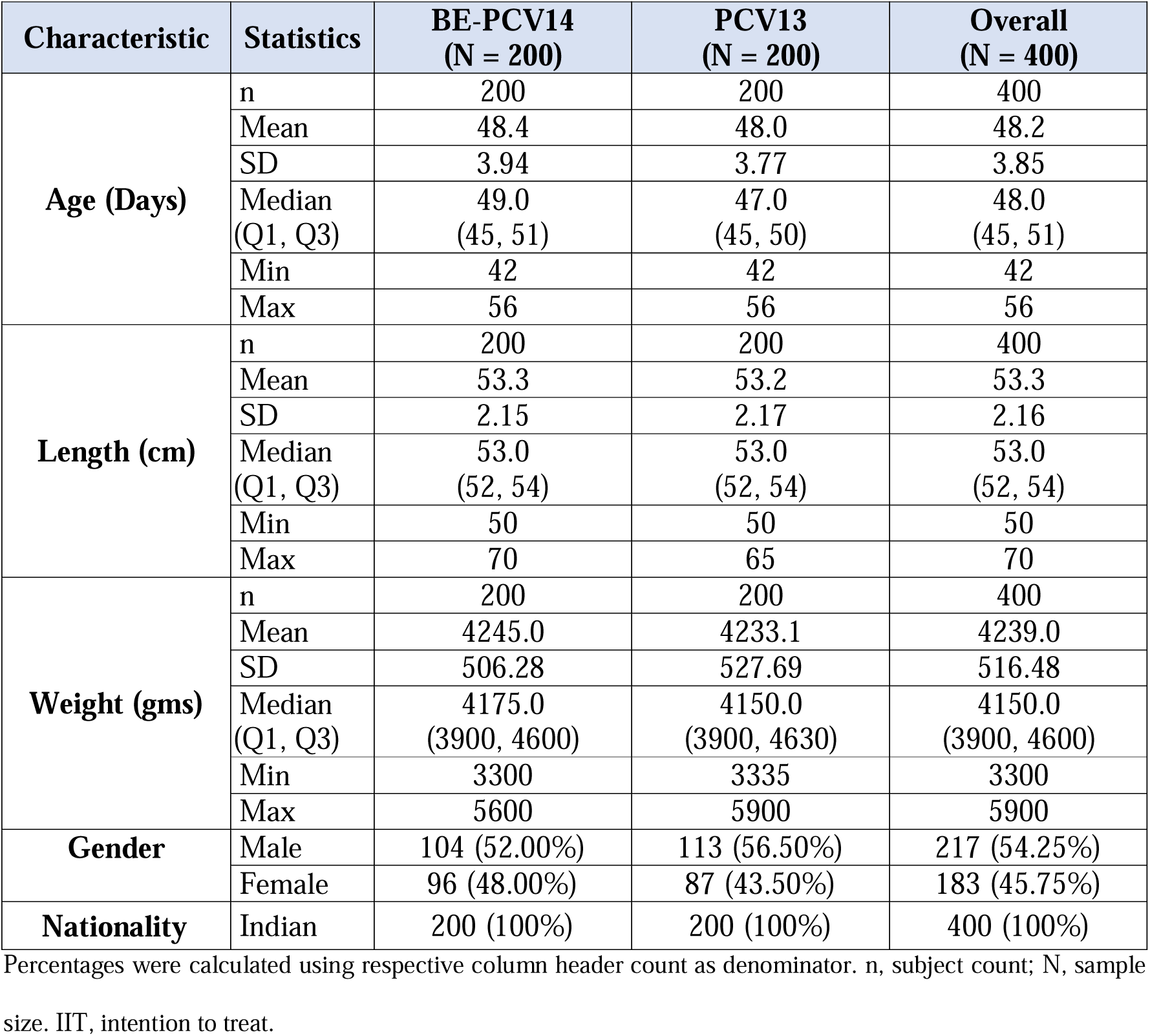
Demographic and Baseline Characteristics – ITT Population (N=400)

### 3.1. Immunogenicity Findings

The primary endpoint was the proportion of infants who achieved seroconversion at three prespecified time points: Day[84 (post-primary series), Month[9 (pre-booster) and Month[10 (one month post-booster) in each vaccine group.

At post-primary time point, seroconversion rates in the BE-PCV14 arm ranged from 72.6% for serotype 3 to 100% for serotypes 14 and 19F; in the PCV13 arm, rates ranged from 71.6% (serotype 3) to 100% (serotypes 14, 19A and 19F). Responses were comparable for 11 of the 12 shared serotypes, with serotype 18C showing a numerically higher rate in the PCV13 group. At pre-booster, BE-PCV14 seroconversion rates ranged between 55.9% (serotype 18C) and 100% (serotype 14), whereas PCV13 rates ranged from 42.8% (serotype 23F) to 99.5% (serotype 14). Notably, BE-PCV14 demonstrated superior persistence for serotypes 3 (57.0% vs 44.8%), 6B (90.9% vs 68.6%), 9V (69.9% vs 59.3%), 18C (55.9% vs 44.8%) and 23F (70.4% vs 42.8%). One month post-booster (Month 10), seroconversion exceeded 88% for every shared serotype in both arms, with virtually identical rates across all common serotypes. Detailed seroconversion data for each time point are illustrated in Figure 2 (bar graphs) and tabulated in Supplementary data Appendix 6. Overall, BE-PCV14 induced seroconversion rates that were comparable to those of PCV13 following both the primary series and booster dose.

**Figure 2:**
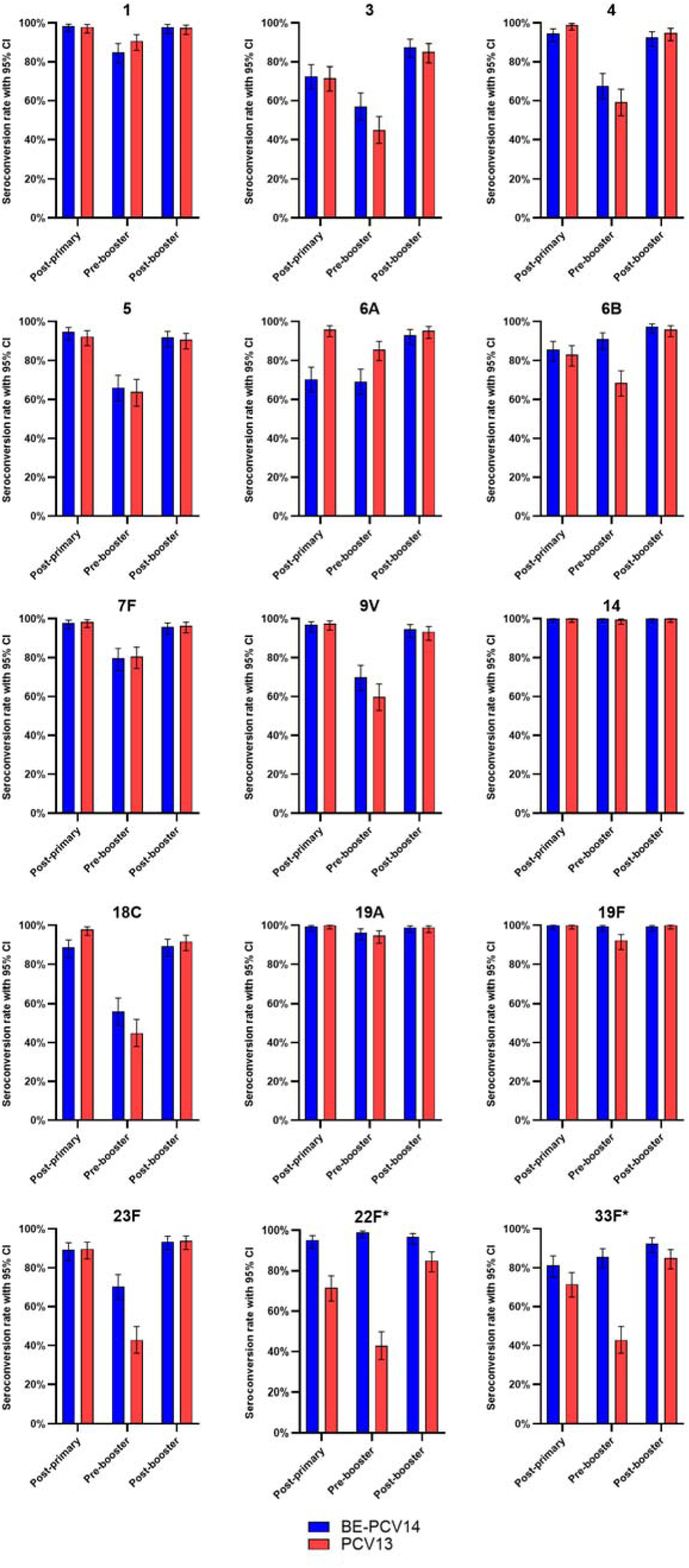
Proportion of subjects with serotype specific seroconversion rate – PP Population (N=380) Data provided for 15 serotypes 1, 3, 4, 5, 6A, 6B, 7F, 9V, 14, 18C, 19A, 19F, 23F, 22F & 33F. BE-PCV14: 14-valent Pneumococcal Polysaccharide Conjugate Vaccine of Biological E. Limited PCV13: Prevenar13® (13-valent Pneumococcal Conjugate Vaccine from Pfizer) *For 22F and 33F, the rates in BE-PCV14 were compared with lowest performing serotype rates in PCV13 Bars shows 95% Confidential Interval

Serotype-specific IgG geometric mean concentrations (GMCs) are shown in Figure 3 and tabulated in supplementary data, Appendix 7. At Day 84 (post-primary series), BE-PCV14 induced GMCs ranging from 0.59 µg/mL (95% CI, 0.52–0.66) for serotype 3 to 8.46 µg/mL (95% CI, 7.02–10.18) for serotype 14; in the PCV13 arm, post-primary GMCs ranged from 0.54 µg/mL (95% CI, 0.48–0.60) for serotype 3 to 6.88 µg/mL for serotype 19F. By Month 9 (pre-booster), BE-PCV14 GMCs varied from 0.42 µg/mL (95% CI, 0.37–0.47) for serotype 3 to 4.89 µg/mL (95% CI, 4.09–5.83) for serotype 14; corresponding PCV13 GMCs ranged from 0.34 µg/mL (95% CI, 0.30–0.38) to 4.31 µg/mL (95% CI, 3.62–5.15). One month post-booster (Month 10), BE-PCV14 elicited GMCs of 0.88 µg/mL (95% CI, 0.78– 0.99) for serotype 3 up to 10.06 µg/mL (95% CI, 8.72–11.62) for serotype 14; PCV13 GMCs ranged from 0.73 µg/mL (95% CI, 0.65–0.81) to 10.60 µg/mL (95% CI, 9.30–12.09) for serotype 14. Overall, GMC profiles were broadly comparable between BE-PCV14 and PCV13 at all assessed time points.

**Figure 3:**
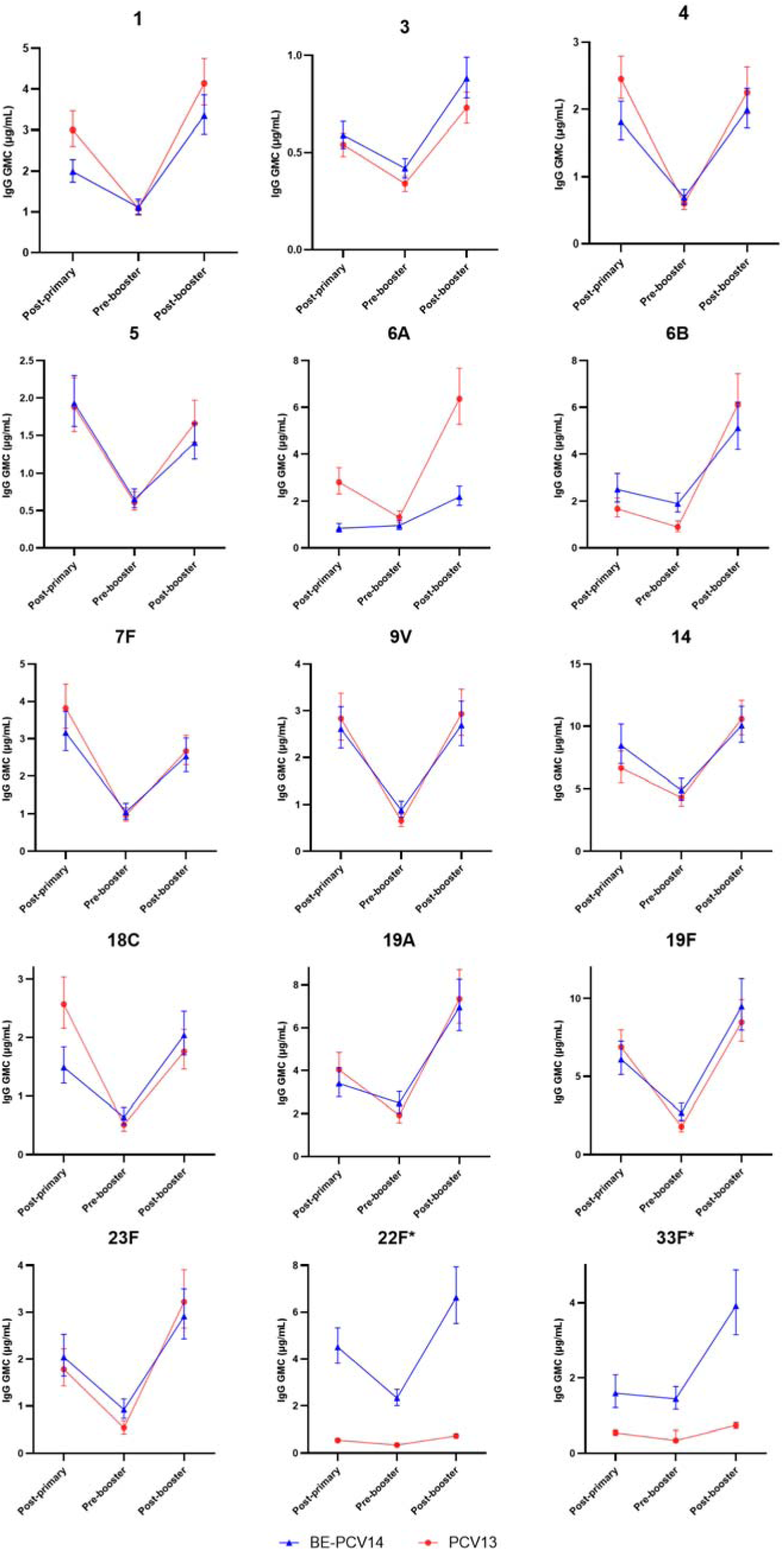
Summary of Geometric Mean Concentrations (GMCs) of serotype specific anti-PnCPS IgG antibodies – PP Population (N=380) *For serotype 22F and 33F, the GMC in BE-PCV14 were compared with lowest performing serotype GMCs in PCV13 at post-primary, pre-booster and post-booster time points

The proportion of participants achieving ≥2-fold and ≥4-fold rises in serotype-specific anti-PnCPS IgG GMCs from baseline at the post-primary, pre-booster and post-booster time points are presented in Supplementary data, Appendix 8. Reverse cumulative distribution curves for all shared serotype IgG responses at the post-primary and post-booster time points were nearly identical between BE-PCV14 and PCV13 (Supplementary data, Appendix 9).

Cross reactive immunogenicity against serotype 6A, not included in the BE PCV14 vaccine, was evaluated to assess potential protection mediated by serotype 6B. Following the two dose primary series, seroconversion rates for 6A were 70.4% (95% CI, 63.5–76.5) at Day 84 (post primary) and 69.3% (95% CI, 62.4–75.5) at Month 9 (pre booster), rising to 93.0% (95% CI, 88.4–95.9) at Month 10 (post booster). Corresponding IgG geometric mean concentrations were 0.84 µg/mL (95% CI, 0.68–1.04) post primary and 0.96 µg/mL (95% CI, 0.78–1.17) pre booster, increasing to 2.19 µg/mL (95% CI, 1.82–2.65) after the booster dose. Comparative immunogenicity data for serotype 6A between BE PCV14 and PCV13 after the three-dose series are summarized in Table 2.

**Table 2:** Seroresponse rate, Geometric mean concentration and proportion of subjects with ≥2-fold GMC rise in serotype 6A – PP population (n=380)

| Parameter | BE-PCV14 (n=186) | PCV-13 (n=194) |
| --- | --- | --- |
| SCR (%) | 93.0 (88.4, 95.9) | 95.4 (91.4, 97.5) |
| IgG GMC ($\mu\text{g/mL}$) | 2.19 (1.82, 2.65) | 6.36 (5.27, 7.68) |
| Proportion of subjects with $\geq 2$-fold rise in GMCs (%) | 62.4 | 82.5 |

### 3.2. Safety findings

Safety was monitored for 28 days after each primary vaccination dose and for 30 days following the booster dose. Solicited adverse events (AEs) were recorded via 7-day patient diary cards after each vaccination, while unsolicited AEs, medically attended AEs, and serious AEs were documented continuously throughout the study period.

Safety evaluations were predefined as secondary objectives. No adverse events (AEs) were observed within the first 60 minutes following any of the three vaccinations. Over the entire study period, 225 AEs were reported among 70 participants (35.0%) in the BE-PCV14 cohort versus 236 AEs in 77 participants (38.5%) in the PCV13 cohort. The vast majority of events (both solicited and unsolicited) occurred within seven days of vaccination (Figure 4; Supplementary Table 10).

**Figure 4:**
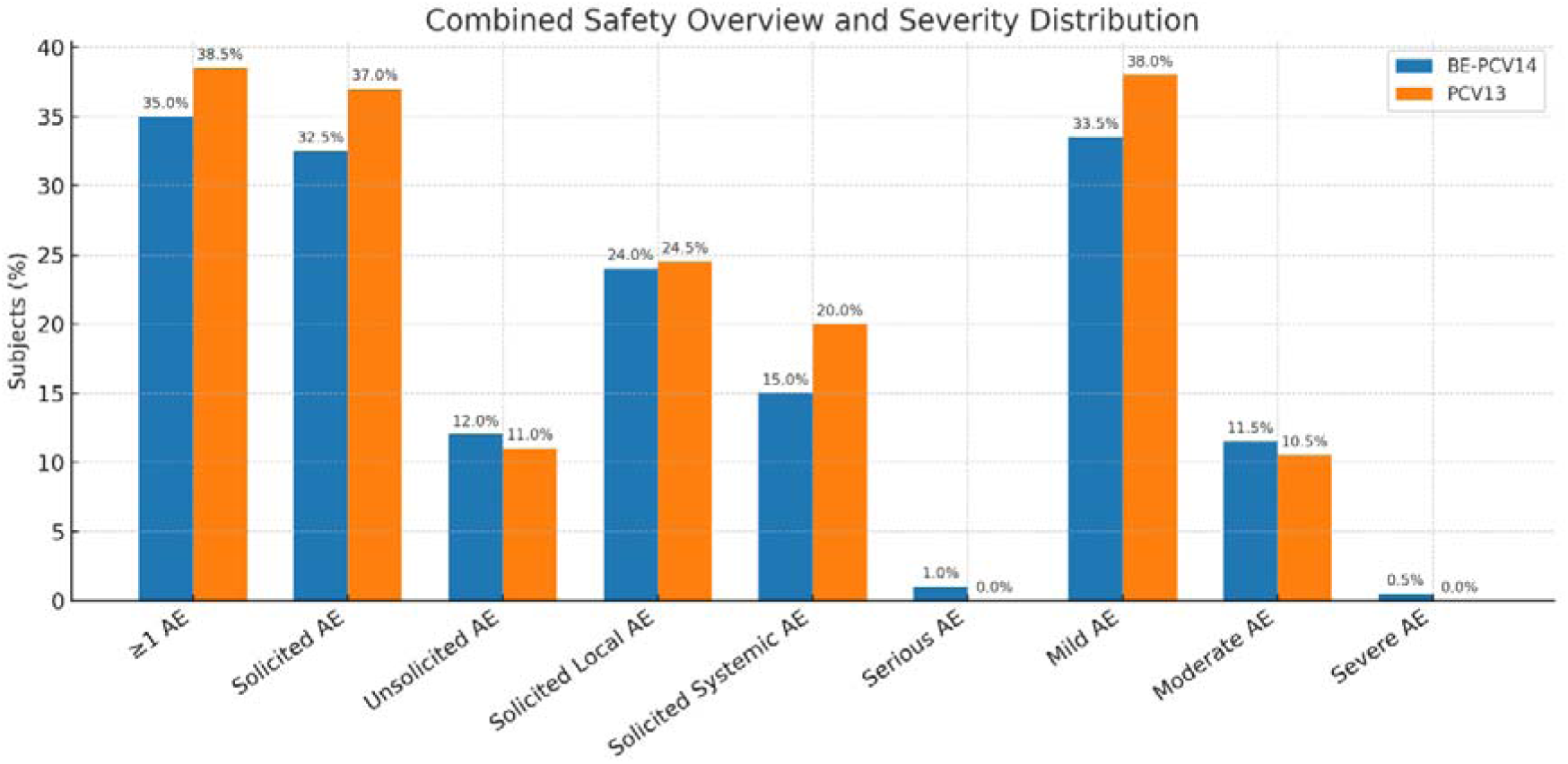
Overview of subjects with Adverse Events after vaccination - Safety Population (N=400)

Solicited reactions in the BE-PCV14 arm were most commonly injection-site pain (35 participants; 17.5%; 40 events), pyrexia (28; 14.0%; 35 events), upper respiratory tract infection (22; 11.0%; 63 events), injection-site swelling (12; 6.0%; 12 events), and erythema (11; 5.5%; 12 events). Among PCV13 recipients, pyrexia (37; 18.5%; 47 events), injection-site pain (34; 17.0%; 42 events), swelling (21; 10.5%; 24 events), upper respiratory tract infection (18; 9.0%; 45 events), and erythema (9; 4.5%; 9 events) were most frequently reported (Figure 5).

**Figure 5:**
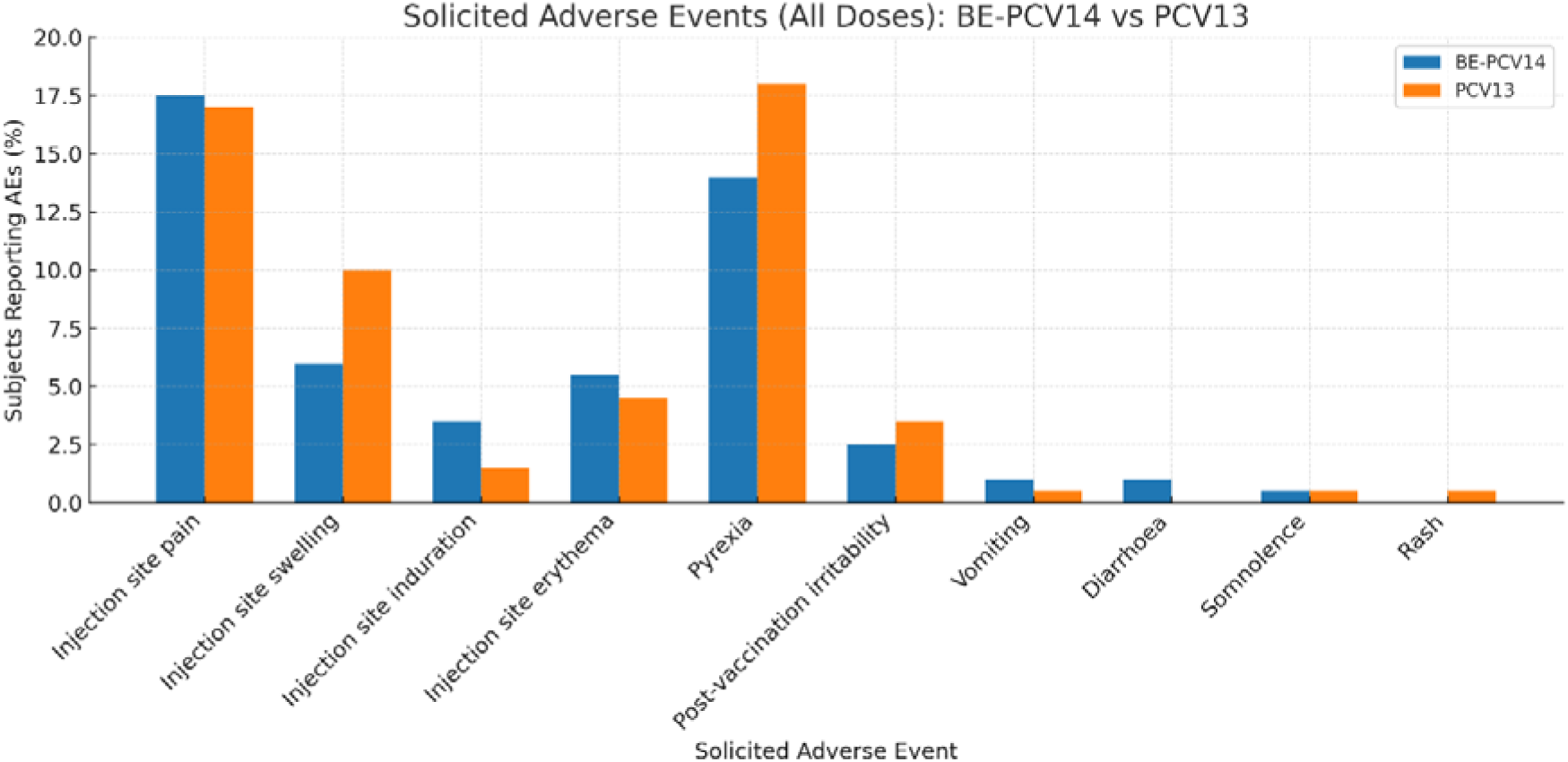
Solicited Adverse events after vaccination - Safety population (N=400)

Two serious adverse events (SAEs) occurred in the BE-PCV14 group: one case of lower respiratory tract infection in an infant with a patent ductus arteriosus, and one case of urinary tract infection. Both events were adjudicated by the principal investigators as unrelated to the study vaccine and resolved fully. No deaths, medically attended AEs, or clinically significant abnormalities in vital signs or physical examinations were reported in either group. Overall, the safety profile of BE-PCV14 was comparable to that of PCV13.

## 4. Discussion

This multicentric, randomized, single-blind, phaselJIII trial was the first trial to compare the BE-PCV14 with PCV13 administered in a 2plJ+lJ1 schedule (two primary doses at 6lJandlJ14lJweeks followed by a booster at 9 months) in Indian infants.

Overall, infants who received BE-PCV14 achieved robust serotype-specific IgG responses that were comparable to those elicited by PCV13. Post primary series at day 84, BE-PCV14 elicited > 90% seroconversion rate for 8 of the 12 common serotypes and >80% for three common serotypes indicating strong priming. At the pre-booster timepoint (month 9), both vaccines demonstrated expected waning of seroconversion rates and Ig G concentration; however, BE-PCV14 exhibited better persistence of immunity for several serotypes. Notably, higher seroconversion rates were observed in the BE-PCV14 group for serotypes 3 (57.0% vs. 44.8%), 6B (90.9% vs. 68.6%), 9V (69.9% vs. 59.3%), 18C (55.9% vs. 44.8%), and 23F (70.4% vs. 42.8%). These findings suggest a more sustained antibody response with BE-PCV14 for specific serotypes, which may have implications for the durability of protection during the period between the primary series and the booster dose.

Following booster dose, the seroconversion rates exceeded 90% for 11 of 12 common serotypes in both vaccine groups. In a post hoc analysis of post booster seroconversion rates, the lower bounds of the two-sided 95% confidence intervals for the difference in seroconversion proportions between BE-PCV14 and PCV13 exceeded –10%, meeting non-inferiority for all 14 serotypes and cross protective serotype 6A (Figure 6A) in accordance with WHO Technical Report Series 977 guidelines.

**Figure 6:**
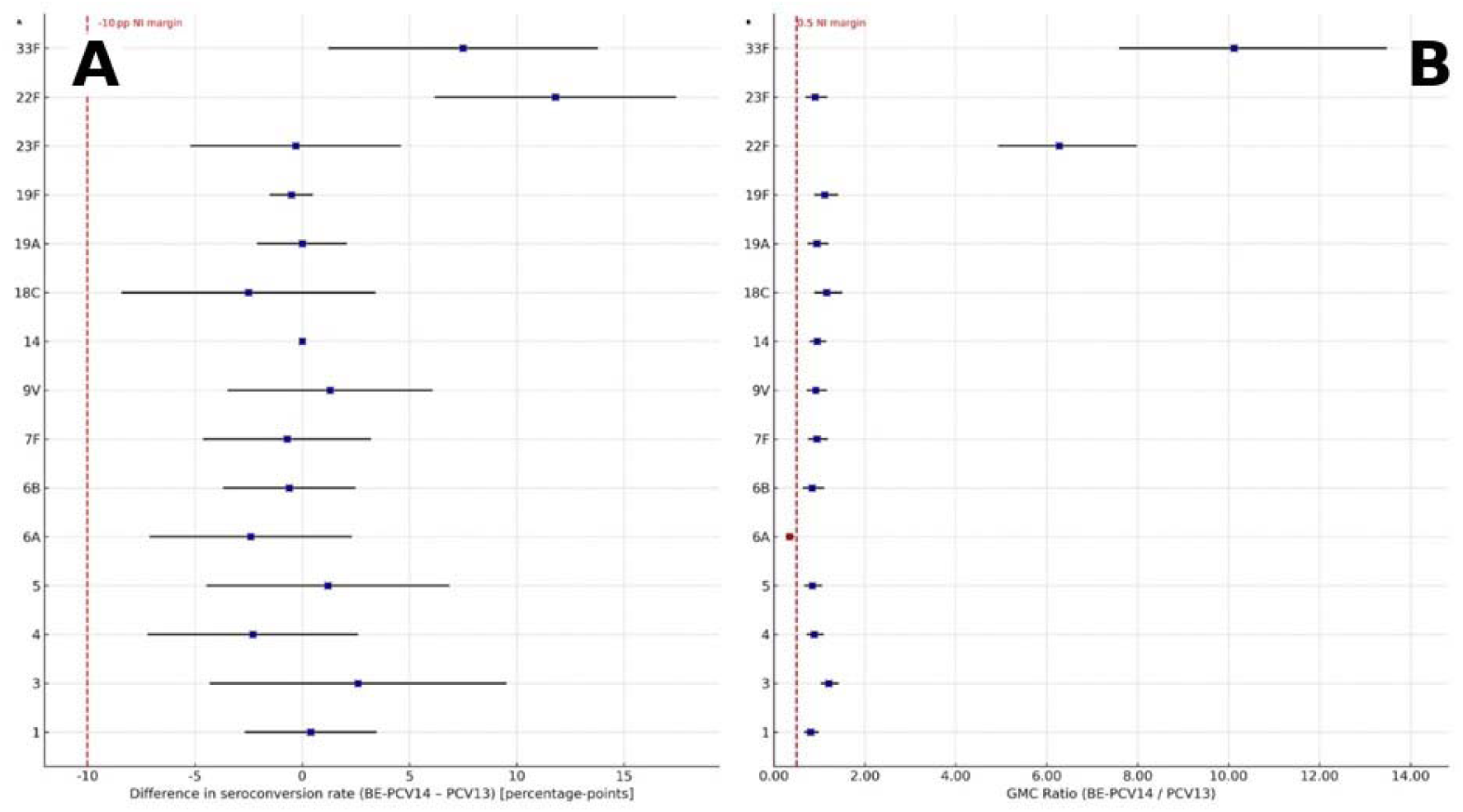
Forest Plot for Non-inferiority between BE-PCV14 and PCV14 Seroconversion rate (A) and IgG GMC at day 300 (Post booster)

The geometric mean concentration (GMC) profiles paralleled those seen with seroconversion. Post-booster GMCs exceeded pre-booster levels for every serotype and surpassed post-primary GMCs for the majority of serotypes. A *post hoc* analysis was performed on the per protocol population post booster GMC ratios. BE-PCV14 met the non-inferiority criteria for all the 14 serotypes with the lower bound 95% CI of GMC ratios exceeding 0.5 (Figure 6B).

BE-PCV14 induced high seroconversion rates and IgG GMCs after the two-dose primary series and elicited robust booster responses against its unique serotypes 22F and 33F. Compared with the lowest-performing PCV13 serotype at the post-booster time point, BE-PCV14 achieved superior seroconversion rates and GMC ratios (Figure 6B). These serotypes are now among the leading non vaccines serotypes, accounting for 9–16 % of invasive pneumococcal disease cases in children and adults in regions with established infant PCV programs^22, 23, 24^ and demonstrate an increased propensity to cause invasive disease following nasopharyngeal colonization in children.^25^ Including 22F and 33F in a conjugate vaccine therefore represents a strategic approach to further reducing the global burden of pneumococcal disease.

In the current study, we also evaluated the cross protective immunogenicity of BE-PCV14 against serotype 6A, attributable to immune responses against serotype 6B, which is included in the vaccine, owing to the high structural similarity between these two serotypes. The seroconversion rate of 70% following 2 dose primary series is consistent with the published 3 dose primary series data for the same vaccine.^16, 17^ Seroconversion rates and IgG GMCs were sustained through the pre-booster time point without evidence of waning. Following the booster dose, seroconversion increased to 93.0%, comparable to 95.4% with PCV13, and IgG GMC rose from 0.84 µg/mL post-primary to 2.19 µg/mL, indicating a robust booster response. These results mirror those of Väkeväinen et al., who reported sustained immunogenicity through the pre-booster period and a robust booster response against cross-reactive serotype 6A following PCV7-CRM vaccination in infants.^26^

Overall, the reverse cumulative distribution curves for all shared serotypes were nearly superimposable between the two vaccines, further supporting the immunological equivalence of BE-PCV14 and PCV13.

From a safety perspective, both vaccines were well tolerated with no safety signals identified. Adverse events were predominantly mild to moderate, solicited in nature, and occurred within the 7-day post-vaccination period. No adverse events were reported within 60 minutes of vaccine administration. The overall incidence of adverse events was comparable between groups: 35.0% in the BE-PCV14 group and 38.5% in the PCV13 group. The most frequently reported adverse events in both groups were pyrexia and injection site pain, consistent with established safety profiles of pneumococcal conjugate vaccines. Importantly, no deaths or medically attended adverse events were reported during the study. Two serious adverse events (SAEs) occurred in the BE-PCV14 group, both assessed by investigators as unrelated to the study vaccine. Clinical examinations, vital signs, and physical assessments throughout the study revealed no vaccine-related concerns. These safety findings align with previously reported data on PCVs and support the favorable safety profile of BE-PCV14.^16, 17^

Multiple randomized trials and meta analyses have assessed immunogenicity and effectiveness of pneumococcal conjugate vaccines in 2p+1 schedule. These studies show that the 2p+1 schedule – two infant doses plus a booster at 9-15 months elicits serotype-specific IgG and opsonophagocytic activity that are comparable with responses after the 3p+0 and 3p+1 schedules. A systematic review of 61 studies reported that, although geometric mean concentrations (GMCs) after the priming phase were usually higher with three infant doses, pre and post booster GMCs were similar regardless of whether infants received two or three priming doses.^27^ In the cluster-randomized FinIP trial of the 10-valent PHiD-CV vaccine conducted in Finland, the 2p+1 immunization regimen conferred 92% effectiveness against invasive pneumococcal disease, closely approximating the 100% effectiveness observed with the 3p+1 schedule.^28^ Population-based surveillance in England following the introduction of PCV7 using a 2p + 1 schedule demonstrated a 90% reduction in vaccine-type invasive pneumococcal disease among children under 5 years and a 50–66% decrease in disease incidence across older age groups.^29^ Following implementation of the 2p + 1 schedule in England, vaccine-type IPD was nearly eliminated in children and substantial herd protection was observed across all age groups^30^, yielding outcomes comparable to those achieved with the 3p + 1 schedule in the United States.^31^ Studies from Netherlands,^32^ and Finland,^33^ showed vaccine effectiveness of ≥90% against vaccine type IPDs in children under 5 years following implementation of 2p+1 schedule. In a retrospective observational study in Australian children, the rate of breakthrough infections in children ≥12 months were significantly lowered (51.7% lower compared to expected IPD) by changing from 3p+0 to 2p+1 schedule for all serotypes of PCV13 excluding serotype 3 indicating superiority of 2p+1 schedule to 3p+0 in overall IPD control.^34^

Immunologically, positioning the third dose in the 9-15 months of life appears advantageous: the systematic review cited earlier showed higher booster induced GMCs when the final dose was deferred beyond six months, supporting longer antibody persistence**Error! Bookmark not defined.**. Nasopharyngeal carriage studies also report equal or greater reductions in nasopharyngeal vaccine type carriage after 2p + 1 compared with 3p + 0, reinforcing the potential for indirect protection.^35^

Taken together, global evidence complements our data and demonstrate that the 2p + 1 strategy provides robust, durable immunity and clinical effectiveness that are non inferior to traditional 3 dose infant schedules. The 2p+1 schedule also affords programmatic flexibility and potential cost savings for national immunisation programmes.

While the results are promising, further post-marketing surveillance will be essential to monitor the long-term safety and real-world effectiveness of BE-PCV14 across diverse populations. Continued evaluation in different geographic regions and in high-risk subpopulations will be critical to fully characterize the vaccine’s performance in routine clinical use.

### Limitations

This study was limited in its modest sample size and descriptive design comparing BE-PCV14 and PCV13 in a 2p+1 schedule. Although the study was not statistically powered to detect between-group differences, post hoc analyses demonstrated that BE-PCV14 achieved non-inferiority to PCV13 for all 14 serotypes with respect to seroconversion rates and IgG geometric mean concentrations following the booster dose. Another limitation was that all immunogenicity analyses were based on pneumococcal capsular polysaccharide (PnCPS) IgG concentrations. Functional antibody activity via opsonophagocytic assay (OPA) - a more direct correlate of protection for PCVs was not assessed in this study. Nonetheless, data from the pivotal Phase III trial of BE-PCV14 using a 3p+0 schedule demonstrated comparable OPA titters with PCV13. This study is limited in its applicability as it was only conducted in India. Studies of BE-PCV14 in other geographies are presently being planned.

## Conclusion

In conclusion, BE-PCV14 demonstrates strong immunogenicity, both post primary with good persistence and post booster vaccination in 2p+1 schedule and an acceptable safety profile in Indian infants. Its inclusion of additional serotypes not present in 10 and 13 valent vaccines offers the potential for enhanced protection against IPD, supporting its consideration as a valuable tool in the global fight against pneumococcal disease. BE-PCV14 also induced robust cross protective immune response with strong booster response against serotype 6A. Future real-world effectiveness studies will be essential to elucidate the performance of BE-PCV14 when administered in a 2p + 1 immunization schedule.

## Data Availability

All data produced in the present study are available upon reasonable request to the authors.

## References

1. Wahl B, O’Brien KL, Greenbaum A, Majumder A, Liu L, Chu Y, et al. Burden of Streptococcus pneumoniae and Haemophilus influenzae type b disease in children in the era of conjugate vaccines: global, regional, and national estimates for 2000-15. Lancet Glob Health. 2018 Jul;6(7):e744–e757.

2. GBD 2016 Lower Respiratory Infections Collaborators. Estimates of the global, regional, and national morbidity, mortality, and aetiologies of lower respiratory infections in 195 countries, 1990–2016: a systematic analysis for the Global Burden of Disease Study 2016. Lancet Infect Dis. 2018;18(11):1191–1210. doi:10.1016/S1473-3099(18)30310-4.

3. Whitney CG, Farley MM, Hadler J, Harrison LH, Bennett NM, Lynfield R, et al. Active Bacterial Core Surveillance of the Emerging Infections Program Network. Decline in invasive pneumococcal disease after the introduction of protein-polysaccharide conjugate vaccine. N Engl J Med. 2003 May 1;348(18):1737–46.

4. Alicino C, Paganino C, Orsi A, Astengo M, Trucchi C, Icardi G, Ansaldi F. The impact of 10-valent and 13-valent pneumococcal conjugate vaccines on hospitalization for pneumonia in children: A systematic review and meta-analysis. Vaccine. 2017 Oct 13;35(43):5776–5785. doi: 10.1016/j.vaccine.2017.09.005. Epub 2017 Sep 11. PMID: 28911902.

5. Bennett JC, Deloria Knoll M, Kagucia EW, Garcia Quesada M, Zeger SL, Hetrich MK, et al. Global impact of ten-valent and 13-valent pneumococcal conjugate vaccines on invasive pneumococcal disease in all ages (the PSERENADE project): a global surveillance analysis. Lancet Infect Dis. 2025 Apr;25(4):457–470. doi: 10.1016/S1473-3099(24)00665-0. Epub 2024 Dec 17. Erratum in: Lancet Infect Dis. 2025 Mar;25(3):e137. doi: 10.1016/S1473-3099(25)00032-5. PMID: 39706204; PMCID: PMC11947069.

6. Metcalf BJ, Gertz RE Jr, Gladstone RA, Walker H, Sherwood LK, Jackson D, et al. Strain features and distributions in pneumococci from children with invasive disease before and after 13-valent conjugate vaccine implementation in the USA. Clin Microbiol Infect. 2016;22(1):60.e9–60.e29. doi:10.1016/j.cmi.2015.07.004.

7. Balsells E, Guillot L, Nair H, Kyaw MH. Serotype distribution of *Streptococcus pneumoniae* causing invasive disease in children in the post-PCV era: A systematic review and meta-analysis. PLoS One. 2017;12(5):e0177113. doi:10.1371/journal.pone.0177113.

8. Jaiswal N, Singh M, Das RR, Jindal I, Agarwal A, Thumburu KK, et al. Distribution of serotypes, vaccine coverage, and antimicrobial susceptibility pattern of Streptococcus pneumoniae in children living in SAARC countries: A systematic review. PLoS One. 2014;9:e108617.

9. John J, Varghese R, Lionell J, Neeravi A, Veeraraghavan B. Non-vaccine Pneumococcal Serotypes Among Children with Invasive Pneumococcal Disease. Indian Pediatr. 2018 Oct 15;55(10):874–876. PMID: 30426954.

10. Kobayashi M, Farrar JL, Gierke R, Leidner AJ, Campos-Outcalt D, Morgan RL, Long SS, Poehling KA, Cohen AL; ACIP Pneumococcal Vaccines Work Group; CDC Contributors. Use of 15-Valent Pneumococcal Conjugate Vaccine Among U.S. Children: Updated Recommendations of the Advisory Committee on Immunization Practices - United States, 2022. MMWR Morb Mortal Wkly Rep. 2022 Sep 16;71(37):1174–1181. doi: 10.15585/mmwr.mm7137a3. PMID: 36107786; PMCID: PMC9484809.

11. ACIP Updates: Recommendations for Use of 20-Valent Pneumococcal Conjugate Vaccine in Children - United States, 2023. MMWR Morb Mortal Wkly Rep. 2023 Sep 29;72(39):1072. doi: 10.15585/mmwr.mm7239a5. PMID: 37768876; PMCID: PMC10545431.

12. World Health Organization. Pneumococcal vaccines: WHO position paper – 2023 update., Wkly Epidemiol Rec. 2023; 98(20): 173–196.

13. Whitney CG, Goldblatt D, O’Brien KL. Dosing schedules for pneumococcal conjugate vaccine: considerations for policy makers. Pediatr Infect Dis J. 2014 Jan;33 Suppl 2(Suppl 2 Optimum Dosing of Pneumococcal Conjugate Vaccine For Infants 0 A Landscape Analysis of Evidence Supportin g Different Schedules):S172-81.

14. Jayasinghe S, Williams PCM, Macartney KK, Crawford NW, Blyth CC. Assessing the Impact of Pneumococcal Conjugate Vaccine Immunization Schedule Change From 3+0 to 2+1 in Australian Children: A Retrospective Observational Study. Clin Infect Dis. 2025 Feb 5;80(1):207–214. doi: 10.1093/cid/ciae377. PMID: 39140767.

15. World Health Organization. Vaccination schedule for pneumococcal disease [Internet]. Geneva: WHO; 2025 [cited 2025 Jun 25]. Available from: https://immunizationdata.who.int/global/wiise-detail-page/vaccination-schedule-for-pneumococcal-disease?ISO_3_CODE=&TARGETPOP_GENERAL=GENERAL

16. Ramesh Matur, Subhash Thuluva, Subbareddy Gunneri, Vijay Yerroju, Rammohanre ddy Mogulla, Kamal Thammireddy, et al. Immunogenicity and Safety of a 14-valent pneumococcal polysaccharide conjugate vaccine (PNEUBEVAX 14TM) administered to 6-8 weeks old healthy Indian Infants: A single blind, randomized, active-controlled, Phase-III study. Vaccine. 2024 May 10;42(13):3157–3165.

17. Thuluva S, Matur RV, Gunneri S, Mogulla RR, Thammireddy K, Peta KK, Paliwal P, Mahantshetti NS, Banala RK, Siddaiah P. Immunogenicity and safety of a multi-human dose formulation of Biological E’s 14-valent pneumococcal polysaccharide conjugate vaccine (PNEUBEVAX 14^®^) administered to 6-8-week-old healthy infants: a phase 3, single-blind, randomized, active-controlled study. Front Immunol. 2025 Apr 7;16:1550227.

18. Wernette CM, Frasch CE, Madore D, Carlone G, Goldblatt D, Plikaytis B, Benjamin W, Quataert SA, Hildreth S, Sikkema DJ, Käyhty H, Jonsdottir I, Nahm MH. Enzyme-linked immunosorbent assay for quantitation of human antibodies to pneumococcal polysaccharides. Clin Diagn Lab Immunol. 2003 Jul;10(4):514–9.

19. Recommendations to assure the quality, safety and efficacy of pneumococcal conjugate vaccines. Replacement of WHO Technical Report Series, No. 927, Annex 2. Available at https://www.who.int/publications/m/item/pneumococcal-conjugate-vaccines-annex3-trs-977. Accessed on June 26, 2025.

20. Snape MD, Klinger CL, Daniels ED, John TM, Layton H, Rollinson L, Pestridge S, Dymond S, Galiza E, Tansey S, Scott DA, Baker SA, Jones TR, Yu LM, Gruber WC, Emini EA, Faust SN, Finn A, Heath PT, Pollard AJ. Immunogenicity and reactogenicity of a 13-valent-pneumococcal conjugate vaccine administered at 2, 4, and 12 months of age: a double-blind randomized active-controlled trial. Pediatr Infect Dis J. 2010 Dec;29(12):e80–90.

21. Clinical Trials RegistrylJ–lJIndia (CTRI). A phase III, multicentre, randomised, double-blind, controlled trial to evaluate the immunogenicity, safety and tolerability of a 10-valent pneumococcal conjugate vaccine (Pneumosil®) administered in a 2 + 1 schedule to healthy Indian infants. CTRI/2019/08/020939*. Registered 14 Aug 2019. Available from:* https://ctri.nic.in. Accessed 24 Jun 2025.

22. Chochua S, Metcalf BJ, Li Z, et al. Pneumococcal serotype evolution and vaccine impact: analysis of invasive pneumococcal disease in the United States, 2010–2018. *Clin Infect Dis*. 2020;71(4):755–763. doi:10.1093/cid/ciz1019.

23. Ladhani SN, Collins S, Djennad A, et al. Effect of the 13-valent pneumococcal conjugate vaccine on invasive pneumococcal disease in England and Wales 4 years after its introduction: an observational cohort study. Lancet Infect Dis. 2018;18(5):539–546. doi:10.1016/S1473-3099(17)30725-7.

24. Tsang RSW, Tyrrell GJ, Vachon J, et al. Epidemiology and serotype distribution of invasive pneumococcal disease in children and adults in Canada, 2010–2016. Can Commun Dis Rep. 2018;44(10):235–244. doi:10.14745/ccdr.v44i10a05.

25. Yildirim M, Keskinocak P, Hinderstein S, Tran K, Dasthagirisaheb YBS, Madoff L, Pelton S, Yildirim I. A comprehensive analysis of serotype-specific invasive capacity, clinical presentations, and mortality trends of invasive pneumococcal disease. Vaccine. 2025 Feb 15;47:126692. doi: 10.1016/j.vaccine.2024.126692. Epub 2025 Jan 7.

26. Väkeväinen M, Eklund C, Eskola J, Käyhty H. Cross-reactivity of antibodies to type 6B and 6A polysaccharides of Streptococcus pneumoniae, evoked by pneumococcal conjugate vaccines, in infants. J Infect Dis. 2001 Sep 15;184(6):789–93.

27. Deloria Knoll M, Park DE, Johnson TS, Chandir S, Nonyane BA, Conklin L, et al. Systematic review of the effect of pneumococcal conjugate vaccine dosing schedules on immunogenicity. Pediatr Infect Dis J. 2014 Jan;33 Suppl 2(Suppl 2 Optimum Dosing of Pneumococcal Conjugate Vaccine For Infants 0 A Landscape Analysis of Evidence Supporting Different Schedules):S119-29.

28. Palmu AA, Jokinen J, Borys D, Nieminen H, Ruokokoski E, Siira L, Puumalainen T, Lommel P, Hezareh M, Moreira M, Schuerman L, Kilpi TM. Effectiveness of the ten-valent pneumococcal Haemophilus influenzae protein D conjugate vaccine (PHiD-CV10) against invasive pneumococcal disease: a cluster randomised trial. Lancet. 2013 Jan 19;381(9862):214-22. doi: 10.1016/S0140-6736(12)61854-6. Epub 2012 Nov 16. Erratum in: Lancet. 2013 May 18;381(9879):1720. Erratum in: Lancet. 2015 May 30;385(9983):2152.

29. Chapman KE, Wilson D, Gorton R. Serotype dynamics of invasive pneumococcal disease post-PCV7 and pre-PCV13 introduction in North East England. Epidemiol Infect. 2013 Feb;141(2):344–52.

30. Miller E, Andrews NJ, Waight PA, Slack MP, George RC. Herd immunity and serotype replacement 4 years after seven-valent pneumococcal conjugate vaccination in England and Wales: an observational cohort study. Lancet Infect Dis. 2011 Oct;11(10):760–8.

31. Centers for Disease Control and Prevention (CDC). Direct and indirect effects of routine vaccination of children with 7-valent pneumococcal conjugate vaccine on incidence of invasive pneumococcal disease--United States, 1998-2003. MMWR Morb Mortal Wkly Rep. 2005 Sep 16;54(36):893–7. PMID: 16163262.

32. Peckeu L, van der Ende A, de Melker HE, Sanders EAM, Knol MJ. Impact and effectiveness of the 10-valent pneumococcal conjugate vaccine on invasive pneumococcal disease among children under 5 years of age in the Netherlands. Vaccine. 2021 Jan 8;39(2):431–437.

33. Jokinen J, Rinta-Kokko H, Siira L, Palmu AA, Virtanen MJ, Nohynek H, et al. Impact of ten-valent pneumococcal conjugate vaccination on invasive pneumococcal disease in Finnish children--a population-based study. PLoS One. 2015 Mar 17;10(3): e0120290.

34. Jayasinghe S, Williams PCM, Macartney KK, Crawford NW, Blyth CC. Assessing the Impact of Pneumococcal Conjugate Vaccine Immunization Schedule Change From 3+0 to 2+1 in Australian Children: A Retrospective Observational Study. Clin Infect Dis. 2025 Feb 5;80(1):207–214.

35. Madhi SA, Moreira M, Koen A, van Niekerk N, de Gouveia L, Jose L, Cutland CL, François N, Schoonbroodt S, Ruiz-Guiñazú J, Yarzabal JP, Borys D, Schuerman L. Impact of HIV status and vaccination schedule on bacterial nasopharyngeal carriage following infant immunisation with the pneumococcal non-typeable Haemophilus influenzae protein D conjugate vaccine in South Africa. Vaccine. 2020 Feb 28;38(10):2350–2360.

