## Supplementary Data for "Immunogenicity and Safety of Biological E’s 14-Valent Pneumococcal Conjugate Vaccine (PNEUBEVAX 14^®^) Administered in a 2p+1 Schedule to Healthy Infants: A Multicenter, Randomized, Active Controlled, Single-Blind, Phase III Trial"

**Appendix 1: Principal Investigators and Clinical study centres**

**INVESTIGATORS & STUDY CENTRE(S):**

The study was conducted at multiple study centers across India.

| **Site ID** | **Principal Investigators and Study Sites** | **Date of EC Approval** |
| --- | --- | --- |
| A | Dr. Manish Narang, M.B.B.S., M.D. (Paediatrics)  UCMS & Guru Teg Bahadur Hospital, Dilshad Garden, Shahdara, Delhi - 110095, India | 09.11.2022 |
| B | Dr. B. S Chakravarthy, M.B.B.S, M.D. (Paediatrics)  King George Hospital, Collectorate Junction, Maharanipeta, Visakhapatnam – 530002, Andhra Pradesh, India. | 31.10.2022 |
| C | Dr. N. Pradeep, M.B.B.S., M.D. (Paediatrics)  Cheluvamba Hospital, Irwin Rd, Devraj Mohalla, Mysuru - 570001, Karnataka, India. | 24.01.2023 |
| E | Dr. Ashish Dhongade, M.B.B.S, M.D. (Paediatrics.)  Sant Dnyaneshwar Medical Education, Research Centre, 695/A, Sadashiv, Peth, Opp. Vijay Talkies, Laxmi Road, Pune-411030, Maharashtra, India. | 12.01.2023 |
| F | Dr. Jog Pramod, M.B.B.S, M.D. (Paediatrics)  Medipoint Hospital, Nagaras Road, DP Road, Near Kumar Padmalaya, Aundh, Pune - 411007, Maharashtra, India | 17.10.2022 |
| G | Dr. Prashanth M. V, M.B.B.S, M.D. (Paediatrics)  Medstar Specialty Hospital, Kodigehalli Main Rd, Sahakar Nagar, Sanjeevini Nagar, Bangalore - 560092, Karnataka, India | 19.06.2023 |
| H | Dr. Anand Kawade, M.B.B.S, M.D. (Paediatrics)  K. E. M Hospital Research Centre, Vadu Rural, Health Program, Post: Vadu (Budruk), Taluka, Shirur, Pune – 412216,India | 13.07.2023 |

**Note: Site D (**Sant Dnyaneshwar Medical Education Research Centre, 695/A, Sadashiv, Peth, Opp. Vijay Talkies, Laxmi Road, Pune-411030, Maharashtra, India; PI – Dr. V. N. Tripathi did not participate in enrollment**)**

**Appendix 2: Inclusion and Exclusion Criteria**

**Inclusion Criteria**

a. Healthy pneumococcal conjugate vaccine-naïve (PCV-naive) infants as established by medical history and clinical assessment before entering into the study. PCV-naïve infants are those who have not been previously vaccinated with any licensed or investigational pneumococcal vaccine.
b. Infants between 6-8 weeks of age (42-56 days, both inclusive), at the time of 1^st^ dose of vaccination.
c. Subjects’ parent(s)/ LAR(s) who, in the opinion of the investigator, can and will comply, with the requirements of the protocol (e.g. completion of the diary cards, return for follow-up visits, with access to a consistent means of telephone contact, either residential land line or mobile).
d. Subject’s parent(s)/LAR(s) willing to provide written or thumb printed informed consent prior to performing any study specific procedure.
e. Infants with a minimal vaccination status for their age at the time of enrolment (“minimal” defined as single dose of BCG, Hepatitis B &/or polio vaccine. at the time of enrolment).

**Exclusion Criteria**

a. Child in care, defined as a child who has been placed under the control or protection of an agency, organisation, institution or entity by the courts, the government or a government body, acting in accordance with powers conferred on them by law or regulation. The definition of a child in care can include a child cared for by foster parents or living in a care home or institution, provided that the arrangement falls within the definition above. The definition of a child in care does not include a child who is adopted or has an appointed legal guardian.
b. Evidence of previous Streptococcus pneumoniae infection or pneumococcal vaccination.
c. Use of any investigational or non-registered product (drug or vaccine) during the period starting 30 days before the administration of study vaccine (Day -29 to Day 0), or planned use during the study period other than the study vaccine.
d. Any medical condition that in the judgment of the investigator would make intramuscular injection unsafe.
e. Chronic administration (defined as more than 14 days in total) of immunosuppressants or other immune-modifying drugs or any blood products during the period starting 30 days prior to the proposed first vaccine dose or planned administration of the same during the study period.
f. Concurrently participating in another clinical study, at any time during the study period, in which the subject has been or will be exposed to an investigational or a non-investigational vaccine/product (pharmaceutical product or device).
g. Any confirmed or suspected immunosuppressive or immunodeficient condition, based on medical history and physical examination (no laboratory testing required).
h. Family history of congenital or hereditary immunodeficiency.
i. History of allergic disease or history of a serious reaction to any prior vaccination or known hypersensitivity likely to be exacerbated by any component of the study vaccines.
j. History of any neurological disorders, meningitis or seizures.
k. Infant who has had a sibling die of sudden infant death syndrome (SIDS) or die suddenly and without apparent other cause or preceding illness in the first year of life.
l. Infant is a direct descendant (child or grand-child) of any person employed by the Sponsor, the Contract Research Organization (CRO) or the Study Site (including the PI and study site personnel).
m. Acute disease and/or fever at the time of vaccination.

Fever is defined as the endogenous elevation of at least one measured body temperature of ≥ 38◦C (≥ 100.4◦F).

n. Acute or chronic, clinically significant pulmonary, cardiovascular, hepatic or renal functional abnormality, as determined by physical examination and Principal investigator judgement.

**Appendix 3: Solicited adverse events**

**Solicited local adverse events**

a. Pain at injection site
b. Redness at injection site
c. Swelling at injection site
d. Induration at injection site

**Solicited systemic (general) adverse events**

a. Decreased/loss of appetite (very common)
b. Irritability (very common)
c. Somnolence; Drowsiness/increased sleep; restless sleep/decreased sleep (very common)
d. Fever-mild (very common)
e. Seizures (including febrile seizures) (uncommon)
f. Hypotonic-hyporesponsive episode (rare)
g. Diarrhoea (common)
h. Vomiting (common)
i. Rash-urticaria like (common)
j. Hypersensitivity reaction including face edema, dyspnoea, bronchospasm (rare)

**Appendix 4: Summary of concomitant vaccination – Safety population (N=400)**

| **Visit** | **Concomitant vaccine administration n (%)** | **BE-PCV14 (N = 200)** | **Prevenar13 (N = 200)** | **Overall (N = 400)** |
| --- | --- | --- | --- | --- |
| Visit 1 | IPV (Inactivated poliovirus vaccine) | 198 (99.00%) | 197 (98.50%) | 395 (98.75%) |
|  | PENTAVALENT (DTWP - RHEPB – Hib) | 198 (99.00%) | 197 (98.50%) | 395 (98.75%) |
|  | ROTAVIRUS VACCINE | 198 (99.00%) | 201 (100.50%) | 399 (99.75%) |
| Visit 2 | IPV (Inactivated poliovirus vaccine) | 192 (96.00%) | 195 (97.50%) | 387 (96.75%) |
|  | PENTAVALENT (DTWP - RHEPB – Hib) | 190 (95.00%) | 190 (95.00%) | 380 (95.00%) |
|  | ROTAVIRUS VACCINE | 192 (96.00%) | 194 (97.00%) | 386 (96.50%) |
| Visit 3 | ROTAVIRUS VACCINE | 0(0%) | 1 (0.50%) | 1 (0.25%) |
| Visit 4 | IPV (Inactivated poliovirus vaccine) | 29 (14.50%) | 32 (16.00%) | 61 (15.25%) |
|  | MEASLES / MR | 40 (20.00%) | 45 (22.50%) | 85 (21.25%) |
|  | ROTAVIRUS VACCINE | 0(0%) | 1 (0.50%) | 1 (0.25%) |
|  | VITAMIN A | 12 (6.00%) | 13 (6.50%) | 25 (6.25%) |
| Visit 5 | JE | 72 (36.00%) | 75 (37.50%) | 147 (36.75%) |
|  | MEASLES / MR | 72 (36.00%) | 75 (37.50%) | 147 (36.75%) |

**Appendix 5:** **Summary of Concomitant Medication by ATC Classification & Medication - Safety Population (N=400)**

| **ATC Level Medicine Name n (%) [95% CI] E** | **BE-PCV14 (N = 200)** | **PCV13 (N = 200)** |
| --- | --- | --- |
| Anilides | 45 (22.50%) | 42 (21.00%) |
|  | [17.26, 28.77] 121 | [15.93, 27.16] 119 |
| Paracetamol | 45 (22.50%) | 42 (21.00%) |
|  | [17.26, 28.77] 121 | [15.93, 27.16] 119 |
| Anticholinergics | 3 (1.50%) | 4 (2.00%) |
|  | [0.51, 4.32] 6 | [0.78, 5.03] 4 |
| Ipratropium | 3 (1.50%) | 3 (1.50%) |
|  | [0.51, 4.32] 6 | [0.51, 4.32] 3 |
| Ipratropium bromide | 0 (0%) | 1 (0.50%) |
|  | NE 0 | [0.09, 2.78] 1 |
| Antidiarrheal microorganisms | 8 (4.00%) | 13 (6.50%) |
|  | [2.04, 7.69] 13 | [3.84, 10.80] 19 |
| Bacillus Clausii | 6 (3.00%) | 9 (4.50%) |
|  | [1.38, 6.39] 7 | [2.39, 8.33] 10 |
| Lactobacillus nos | 2 (1.00%) | 5 (2.50%) |
|  | [0.27, 3.57] 3 | [1.07, 5.72] 6 |
| Lactobacillus rhamnosus | 1 (0.50%) | 1 (0.50%) |
|  | [0.09, 2.78] 1 | [0.09, 2.78] 1 |
| Saccharomyces boulardii | 2 (1.00%) | 2 (1.00%) |
|  | [0.27, 3.57] 2 | [0.27, 3.57] 2 |
| Antiinfectives | 6 (3.00%) | 3 (1.50%) |
|  | [1.38, 6.39] 13 | [0.51, 4.32] 3 |
| Chloramphenicol | 6 (3.00%) | 2 (1.00%) |
|  | [1.38, 6.39] 11 | [0.27, 3.57] 2 |
| Clotrimazole | 2 (1.00%) | 1 (0.50%) |
|  | [0.27, 3.57] 2 | [0.09, 2.78] 1 |
| Beta-lactamase inhibitors | 14 (7.00%) | 14 (7.00%) |
|  | [4.22, 11.41] 30 | [4.22, 11.41] 25 |
| Clavulanic acid | 14 (7.00%) | 13 (6.50%) |
|  | [4.22, 11.41] 28 | [3.84, 10.80] 24 |
| Tazobactam | 2 (1.00%) | 1 (0.50%) |
|  | [0.27, 3.57] 2 | [0.09, 2.78] 1 |
|  | NE 0 | [0.09, 2.78] 1 |
| Expectorants | 21 (10.50%) | 19 (9.50%) |
|  | [6.97, 15.52] 61 | [6.17, 14.36] 50 |
| Ammonium chloride | 0 (0%) | 1 (0.50%) |
|  | NE 0 | [0.09, 2.78] 1 |
| Guaifenesin | 21 (10.50%) | 19 (9.50%) |
|  | [6.97, 15.52] 61 | [6.17, 14.36] 48 |
| Sodium citrate | 0 (0%) | 1 (0.50%) |
|  | NE 0 | [0.09, 2.78] 1 |
| Fenamates | 4 (2.00%) | 6 (3.00%) |
|  | [0.78, 5.03] 6 | [1.38, 6.39] 8 |
| Mefenamic acid | 4 (2.00%) | 6 (3.00%) |
|  | [0.78, 5.03] 6 | [1.38, 6.39] 8 |
| Fluoroquinolones | 7 (3.50%) | 6 (3.00%) |
|  | [1.71, 7.05] 7 | [1.38, 6.39] 10 |
| Levofloxacin | 1 (0.50%) | 0 (0%) |
|  | [0.09, 2.78] 1 | NE 0 |
| Norfloxacin | 1 (0.50%) | 0 (0%) |
|  | [0.09, 2.78] 1 | NE 0 |
| Ofloxacin | 5 (2.50%) | 6 (3.00%) |
|  | [1.07, 5.72] 5 | [1.38, 6.39] 10 |
| Folic acid and derivatives | 3 (1.50%) | 1 (0.50%) |
|  | [0.51, 4.32] 5 | [0.09, 2.78] 1 |
| Folic acid | 3 (1.50%) | 1 (0.50%) |
|  | [0.51, 4.32] 5 | [0.09, 2.78] 1 |
| Glucocorticoids | 14 (7.00%) | 10 (5.00%) |
|  | [4.22, 11.41] 30 | [2.74, 8.96] 21 |
| Beclometasone | 0 (0%) | 1 (0.50%) |
|  | NE 0 | [0.09, 2.78] 1 |
| Beclometasone dipropionate | 1 (0.50%) | 0 (0%) |
|  | [0.09, 2.78] 3 | NE 0 |
| Budesonide | 13 (6.50%) | 9 (4.50%) |
|  | [3.84, 10.80] 20 | [2.39, 8.33] 17 |
| Deflazacort | 1 (0.50%) | 0 (0%) |
|  | [0.09, 2.78] 1 | NE 0 |
| Hydrocortisone sodium succinate | 1 (0.50%) | 0 (0%) |
|  | [0.09, 2.78] 1 | NE 0 |
| Prednisolone | 3 (1.50%) | 2 (1.00%) |
|  | [0.51, 4.32] 5 | [0.27, 3.57] 3 |
| Hedera helix leaf dry extract | 3 (1.50%) | 0 (0%) |
|  | [0.51, 4.32] 3 | NE 0 |
| Imidazole derivatives | 5 (2.50%) | 5 (2.50%) |
|  | [1.07, 5.72] 5 | [1.07, 5.72] 7 |
| Metronidazole | 4 (2.00%) | 5 (2.50%) |
|  | [0.78, 5.03] 4 | [1.07, 5.72] 7 |
| Ornidazole | 1 (0.50%) | 0 (0%) |
|  | [0.09, 2.78] 1 | NE 0 |
| Iron bivalent, oral preparations | 3 (1.50%) | 1 (0.50%) |
|  | [0.51, 4.32] 5 | [0.09, 2.78] 1 |
| Ferrous ascorbate | 3 (1.50%) | 1 (0.50%) |
|  | [0.51, 4.32] 5 | [0.09, 2.78] 1 |
| Leukotriene receptor antagonists | 6 (3.00%) | 4 (2.00%) |
|  | [1.38, 6.39] 10 | [0.78, 5.03] 6 |
| Montelukast | 6 (3.00%) | 4 (2.00%) |
|  | [1.38, 6.39] 10 | [0.78, 5.03] 6 |
| Macrolides | 5 (2.50%) | 7 (3.50%) |
|  | [1.07, 5.72] 7 | [1.71, 7.05] 8 |
| Azithromycin | 5 (2.50%) | 6 (3.00%) |
|  | [1.07, 5.72] 7 | [1.38, 6.39] 7 |
| Erythromycin | 0 (0%) | 1 (0.50%) |
|  | NE 0 | [0.09, 2.78] 1 |
| Mucolytics | 21 (10.50%) | 19 (9.50%) |
|  | [6.97, 15.52] 70 | [6.17, 14.36] 62 |
| Ambroxol | 21 (10.50%) | 19 (9.50%) |
|  | [6.97, 15.52] 70 | [6.17, 14.36] 60 |
| Bromhexine hydrochloride | 0 (0%) | 1 (0.50%) |
|  | NE 0 | [0.09, 2.78] 1 |
| Sodium chloride | 0 (0%) | 1 (0.50%) |
|  | NE 0 | [0.09, 2.78] 1 |
|  | NE 0 | [0.09, 2.78] 1 |
|  | NE 0 | [0.09, 2.78] 1 |
| Other drugs for functional gastrointestinal disorders | 5 (2.50%) | 3 (1.50%) |
|  | [1.07, 5.72] 6 | [0.51, 4.32] 3 |
| Dimeticone | 1 (0.50%) | 0 (0%) |
|  | [0.09, 2.78] 1 | NE 0 |
| Simeticone | 4 (2.00%) | 3 (1.50%) |
|  | [0.78, 5.03] 5 | [0.51, 4.32] 3 |
| Other nasal preparations | 9 (4.50%) | 12 (6.00%) |
|  | [2.39, 8.33] 13 | [3.47, 10.19] 15 |
| Sodium chloride | 9 (4.50%) | 12 (6.00%) |
|  | [2.39, 8.33] 13 | [3.47, 10.19] 15 |
| Penicillins with extended spectrum | 19 (9.50%) | 16 (8.00%) |
|  | [6.17, 14.36] 43 | [4.98, 12.60] 34 |
| Amoxicillin | 19 (9.50%) | 15 (7.50%) |
|  | [6.17, 14.36] 41 | [4.60, 12.00] 33 |
| Piperacillin | 2 (1.00%) | 1 (0.50%) |
|  | [0.27, 3.57] 2 | [0.09, 2.78] 1 |
| Piperazine derivatives | 18 (9.00%) | 14 (7.00%) |
|  | [5.77, 13.78] 28 | [4.22, 11.41] 23 |
| Cetirizine | 6 (3.00%) | 6 (3.00%) |
|  | [1.38, 6.39] 6 | [1.38, 6.39] 6 |
| Levocetirizine | 13 (6.50%) | 12 (6.00%) |
|  | [3.84, 10.80] 22 | [3.47, 10.19] 17 |
| Propionic acid derivatives | 12 (6.00%) | 10 (5.00%) |
|  | [3.47, 10.19] 20 | [2.74, 8.96] 21 |
| Ibuprofen | 12 (6.00%) | 10 (5.00%) |
|  | [3.47, 10.19] 20 | [2.74, 8.96] 21 |
| Selective beta-2-adrenoreceptor agonists | 21 (10.50%) | 19 (9.50%) |
|  | [6.97, 15.52] 96 | [6.17, 14.36] 76 |
| Levosalbutamol | 20 (10.00%) | 19 (9.50%) |
|  | [6.57, 14.94] 82 | [6.17, 14.36] 66 |
| Salbutamol | 3 (1.50%) | 3 (1.50%) |
|  | [0.51, 4.32] 4 | [0.51, 4.32] 4 |
| Terbutaline | 5 (2.50%) | 5 (2.50%) |
|  | [1.07, 5.72] 10 | [1.07, 5.72] 6 |
| Serotonin (5HT3) antagonists | 7 (3.50%) | 5 (2.50%) |
|  | [1.71, 7.05] 8 | [1.07, 5.72] 7 |
| Granisetron | 1 (0.50%) | 1 (0.50%) |
|  | [0.09, 2.78] 1 | [0.09, 2.78] 1 |
| Ondansetron | 5 (2.50%) | 5 (2.50%) |
|  | [1.07, 5.72] 5 | [1.07, 5.72] 5 |
| Ondansetron hydrochloride | 2 (1.00%) | 1 (0.50%) |
|  | [0.27, 3.57] 2 | [0.09, 2.78] 1 |
|  | [0.09, 2.78] 1 | NE 0 |
| Substituted alkylamines | 21 (10.50%) | 18 (9.00%) |
|  | [6.97, 15.52] 65 | [5.77, 13.78] 51 |
| Chlorphenamine maleate | 21 (10.50%) | 18 (9.00%) |
|  | [6.97, 15.52] 65 | [5.77, 13.78] 51 |
| Sympathomimetics | 23 (11.50%) | 18 (9.00%) |
|  | [7.79, 16.66] 68 | [5.77, 13.78] 52 |
| Phenylephrine | 23 (11.50%) | 18 (9.00%) |
|  | [7.79, 16.66] 68 | [5.77, 13.78] 52 |
| Sympathomimetics, plain | 5 (2.50%) | 4 (2.00%) |
|  | [1.07, 5.72] 9 | [0.78, 5.03] 6 |
| Oxymetazoline | 2 (1.00%) | 3 (1.50%) |
|  | [0.27, 3.57] 4 | [0.51, 4.32] 4 |
| Oxymetazoline hydrochloride | 1 (0.50%) | 0 (0%) |
|  | [0.09, 2.78] 1 | NE 0 |
| Xylometazoline | 2 (1.00%) | 1 (0.50%) |
|  | [0.27, 3.57] 3 | [0.09, 2.78] 2 |
| Xylometazoline hydrochloride | 1 (0.50%) | 0 (0%) |
|  | [0.09, 2.78] 1 | NE 0 |
| Third-generation cephalosporins | 11 (5.50%) | 11 (5.50%) |
|  | [3.10, 9.58] 18 | [3.10, 9.58] 21 |
| Cefixime | 5 (2.50%) | 8 (4.00%) |
|  | [1.07, 5.72] 6 | [2.04, 7.69] 12 |
| Cefpodoxime | 7 (3.50%) | 4 (2.00%) |
|  | [1.71, 7.05] 11 | [0.78, 5.03] 6 |
| Cefpodoxime proxetil | 0 (0%) | 1 (0.50%) |
|  | NE 0 | [0.09, 2.78] 2 |
| Ceftriaxone | 1 (0.50%) | 1 (0.50%) |
|  | [0.09, 2.78] 1 | [0.09, 2.78] 1 |
| Zinc | 3 (1.50%) | 6 (3.00%) |
|  | [0.51, 4.32] 3 | [1.38, 6.39] 7 |
| Zinc | 1 (0.50%) | 3 (1.50%) |
|  | [0.09, 2.78] 1 | [0.51, 4.32] 3 |
| Zinc gluconate | 1 (0.50%) | 1 (0.50%) |
|  | [0.09, 2.78] 1 | [0.09, 2.78] 2 |
| Zinc oxide | 1 (0.50%) | 1 (0.50%) |
|  | [0.09, 2.78] 1 | [0.09, 2.78] 1 |
| Zinc sulfate | 0 (0%) | 1 (0.50%) |
|  | NE 0 | [0.09, 2.78] 1 |

**Appendix 6: Summary of serotype specific Seroconversion rates – PP Population (N=380)**

| **Serotype** | **Measurement Day** | **BE-PCV14**  **(n=186)**  **% (95% CI)** | **Prevenar 13**  **(n=194)**  **% (95% CI)** |
| --- | --- | --- | --- |
| 1 | Day 84 (post-primary) | 98.4 (95.4, 99.4) | 97.9 (94.8, 99.2) |
|  | Day 270 (pre-booster) | 84.9 (79.1, 89.4) | 90.7 (85.8, 94.0) |
|  | Day 300 (post-booster) | 97.8 (94.6, 99.2) | 97.4 (94.1, 98.9) |
| 3 | Day 84 | 72.6 (65.8, 78.5) | 71.6 (64.9, 77.5) |
|  | Day 270 | 56.9 (49.8, 63.9) | 44.8 (38.0, 51.9) |
|  | Day 300 | 87.6 (82.1, 91.6) | 85.0 (79.4, 89.4) |
| 4 | Day 84 | 94.6 (90.4, 97.0) | 98.9 (96.3, 99.7) |
|  | Day 270 | 67.7 (60.7, 74.0) | 59.3 (52.2, 65.9) |
|  | Day 300 | 92.5 (87.8, 95.5) | 94.8 (90.8, 97.2) |
| 5 | Day 84 | 94.6 (90.4, 97.0) | 92.3 (87.6, 95.3) |
|  | Day 270 | 66.1 (59.1, 72.5) | 63.9 (56.5, 70.3) |
|  | Day 300 | 91.9 (87.1, 95.0) | 90.7 (85.8, 94.0) |
| 6A | Day 84 | 70.4 (63.5, 76.5) | 95.9 (92.1, 97.9) |
|  | Day 270 | 69.3 (62.4, 75.5) | 85.6 (79.9, 89.8) |
|  | Day 300 | 93.0 (88.4, 95.9) | 95.4 (91.4, 97.5) |
| 6B | Day 84 | 85.5 (79.7, 89.8) | 82.9 (77.1, 87.6) |
|  | Day 270 | 90.9 (85.8, 94.2) | 68.6 (61.7, 74.7) |
|  | Day 300 | 97.3 (93.9, 98.8) | 95.9 (92.1, 97.9) |
| 7F | Day 84 | 97.8 (94.6, 99.2) | 98.4 (95.5, 99.5) |
|  | Day 270 | 79.6 (73.2, 84.7) | 80.4 (74.3, 85.4) |
|  | Day 300 | 95.7 (91.7, 97.8) | 96.4 (92.7, 98.2) |
| 9V | Day 84 | 96.8 (93.1, 98.5) | 97.4 (94.1, 98.9) |
|  | Day 270 | 69.9 (62.9, 76.0) | 59.8 (52.8, 66.4) |
|  | Day 300 | 94.6 (90.4, 97.0) | 93.3 (88.9, 96.0) |
| 14 | Day 84 | 100 (97.9, 100) | 100 (98.1, 100) |
|  | Day 270 | 100 (97.9, 100) | 99.5 (97.1, 99.9) |
|  | Day 300 | 100 (97.9, 100) | 100 (98.1, 100) |
| 18C | Day 84 | 88.7 (83.4, 92.5) | 97.9 (94.8, 99.2) |
|  | Day 270 | 55.9 (48.7, 62.9) | 44.8 (38.0, 51.9) |
|  | Day 300 | 89.2 (83.9, 92.9) | 91.7 (87.0, 94.9) |
| 19A | Day 84 | 99.5 (97.0, 99.9) | 100 (98.1, 100) |
|  | Day 270 | 96.2 (92.4, 98.2) | 94.8 (90.8, 97.2) |
|  | Day 300 | 98.9 (96.2, 99.7) | 98.9 (96.3, 99.7) |
| 19F | Day 84 | 100 (97.9, 100) | 100 (98.1, 100) |
|  | Day 270 | 99.5 (97.0, 99.9) | 92.3 (87.6, 95.3) |
|  | Day 300 | 99.5 (97.0, 99.9) | 100 (98.1, 100) |
| 23F | Day 84 | 89.2 (83.9, 92.9) | 89.7 (84.6, 93.2) |
|  | Day 270 | 70.4 (63.5, 76.5) | 42.8 (36.0, 49.8) |
|  | Day 300 | 93.5 (89.06, 96.3) | 93.8 (89.5, 96.4) |
| 22F | Day 84 | 95.2 (91.1, 97.4) | 71.6 (64.9, 77.5) |
|  | Day 270 | 98.9 (96.2, 99.7) | 42.8 (36.0, 49.8) |
|  | Day 300 | 96.8 (93.1, 98.5) | 85.0 (79.4, 89.4) |
| 33F | Day 84 | 81.2 (74.9, 86.1) | 71.6 (64.9, 77.5) |
|  | Day 270 | 85.5 (79.7, 89.8) | 42.8 (36.0, 49.8) |
|  | Day 300 | 92.5 (87.8, 95.5) | 85.0 (79.4, 89.4) |

**Appendix 7: Summary of Geometric Mean Concentrations (GMCs) of serotype specific anti-PnCPS IgG antibodies – PP Population (N=380)**

| **Serotype** | **Measurement Day** | **BE-PCV14**  **(n=186)**  **µg/mL (95% CI)** | **PCV 13**  **(n=194)**  **µg/mL (95% CI)** |
| --- | --- | --- | --- |
| 1 | Day 84 (post-primary) | 1.99 (1.73, 2.28) | 3.00 (2.60, 3.47) |
|  | Day 270 (pre-booster) | 1.11 (0.94, 1.31) | 1.08 (0.92, 1.26) |
|  | Day 300 (post-booster) | 3.35 (2.90, 3.86) | 4.14 (3.61, 4.75) |
| 3 | Day 84 | 0.59 (0.52, 0.66) | 0.54 (0.48, 0.60) |
|  | Day 270 | 0.42 (0.37, 0.47) | 0.34 (0.30, 0.38) |
|  | Day 300 | 0.88 (0.78, 0.99) | 0.73 (0.65, 0.81) |
| 4 | Day 84 | 1.81 (1.55, 2.12) | 2.45 (2.16, 2.79) |
|  | Day 270 | 0.69 (0.59, 0.81) | 0.60 (0.51, 0.72) |
|  | Day 300 | 1.99 (1.72, 2.31) | 2.25 (1.93, 2.63) |
| 5 | Day 84 | 1.93 (1.62, 2.30) | 1.88 (1.55, 2.27) |
|  | Day 270 | 0.65 (0.54, 0.79) | 0.61 (0.51, 0.75) |
|  | Day 300 | 1.40 (1.19, 1.66) | 1.66 (1.41, 1.97) |
| 6A | Day 84 | 0.84 (0.68, 1.04) | 2.82 (2.31, 3.44) |
|  | Day 270 | 0.96 (0.78, 1.17) | 1.31 (1.08, 1.59) |
|  | Day 300 | 2.19 (1.82, 2.65) | 6.36 (5.27, 7.68) |
| 6B | Day 84 | 2.50 (1.97, 3.19) | 1.68 (1.33, 2.14) |
|  | Day 270 | 1.90 (1.54, 2.36) | 0.89 (0.70, 1.16) |
|  | Day 300 | 5.11 (4.20, 6.22) | 6.11 (5.02, 7.44) |
| 7F | Day 84 | 3.16 (2.68, 3.73) | 3.82 (3.28, 4.46) |
|  | Day 270 | 1.05 (0.86, 1.28) | 0.97 (0.81, 1.16) |
|  | Day 300 | 2.53 (2.12, 3.02) | 2.67 (2.31, 3.09) |
| 9V | Day 84 | 2.61 (2.20, 3.09) | 2.83 (2.37, 3.38) |
|  | Day 270 | 0.88 (0.73, 1.07) | 0.65 (0.53, 0.80) |
|  | Day 300 | 2.69 (2.25, 3.21) | 2.93 (2.47, 3.46) |
| 14 | Day 84 | 8.46 (7.02, 10.18) | 6.65 (5.51, 8.02) |
|  | Day 270 | 4.89 (4.09, 5.83) | 4.31 (3.62, 5.15) |
|  | Day 300 | 10.06 (8.72, 11.62) | 10.60 (9.30, 12.09) |
| 18C | Day 84 | 1.49 (1.22, 1.84) | 2.57 (2.16, 3.04) |
|  | Day 270 | 0.64 (0.51, 0.81) | 0.51 (0.40, 0.64) |
|  | Day 300 | 2.04 (1.70, 2.45) | 1.76 (1.46, 2.14) |
| 19A | Day 84 | 3.39 (2.78, 4.13) | 4.03 (3.36, 4.84) |
|  | Day 270 | 2.48 (2.02, 3.03) | 1.92 (1.56, 2.36) |
|  | Day 300 | 6.96 (5.87, 8.27) | 7.35 (6.21, 8.72) |
| 19F | Day 84 | 6.09 (5.12, 7.26) | 6.88 (5.92, 7.99) |
|  | Day 270 | 2.68 (2.20, 3.28) | 1.78 (1.46, 2.17) |
|  | Day 300 | 9.49 (7.98, 11.28) | 8.48 (7.25, 9.93) |
| 23F | Day 84 | 2.04 (1.64, 2.53) | 1.78 (1.43, 2.22) |
|  | Day 270 | 0.92 (0.74, 1.15) | 0.53 (0.41, 0.69) |
|  | Day 300 | 2.91 (2.43, 3.50) | 3.22 (2.66, 3.91) |
| 22F | Day 84 | 4.51 (3.82, 5.33) | 0.54 (0.48, 0.60) |
|  | Day 270 | 2.33 (2.01, 2.70) | 0.34 (0.30, 0.38) |
|  | Day 300 | 6.62 (5.52, 7.94) | 0.73 (0.65, 0.81) |
| 33F | Day 84 | 1.59 (1.21, 2.08) | 0.54 (0.48, 0.60 |
|  | Day 270 | 1.44 (1.17, 1.77) | 0.34 (0.30, 0.60) |
|  | Day 300 | 3.92 (3.15, 4.88) | 0.73 (0.65, 0.81) |

**Appendix 8:Proportion of subjects achieving ≥2-fold & ≥4-fold rise in anti-PnCPS IgG antibody concentrations – PP Population (N=380)**

| **Serotype**  **N1 (%) [95% CI]** | **Visit** | **Fold Rise** | **BE-PCV14 (N = 186)** | **Prevenar13 (N = 194)** |
| --- | --- | --- | --- | --- |
| 1 | Day 84 | ≥2-fold | 144 (77.42%) | 168 (86.60%) |
|  |  |  | (70.89, 82.84) | (81.09, 90.69) |
|  |  | ≥4-fold | 103 (55.38%) | 131 (67.53%) |
|  |  |  | (48.20, 62.34) | (60.65, 73.72) |
|  | Day 270 (Pre-Booster) | ≥2-fold | 103 (55.38%) | 113 (58.25%) |
|  |  |  | (48.20, 62.34) | (51.21, 64.96) |
|  |  | ≥4-fold | 58 (31.18%) | 68 (35.05%) |
|  |  |  | (24.96, 38.16) | (28.69, 42.00) |
|  | Day 300 (Booster Response) | ≥2-fold | 158 (84.95%) | 170 (87.63%) |
|  |  |  | (79.10, 89.37) | (82.25, 91.54) |
|  |  | ≥4-fold | 134 (72.04%) | 144 (74.23%) |
|  |  |  | (65.20, 78.00) | (67.64, 79.87) |
| 3 | Day 84 | ≥2-fold | 101 (54.30%) | 101 (52.06%) |
|  |  |  | (47.13, 61.30) | (45.06, 58.98) |
|  |  | ≥4-fold | 55 (29.57%) | 42 (21.65%) |
|  |  |  | (23.48, 36.49) | (16.43, 27.97) |
|  | Day 270 (Pre-Booster) | ≥2-fold | 69 (37.10%) | 64 (32.99%) |
|  |  |  | (30.48, 44.23) | (26.76, 39.88) |
|  |  | ≥4-fold | 35 (18.82%) | 31 (15.98%) |
|  |  |  | (13.85, 25.04) | (11.49, 21.79) |
|  | Day 300 (Booster Response) | ≥2-fold | 125 (67.20%) | 120 (61.86%) |
|  |  |  | (60.17, 73.54) | (54.85, 68.40) |
|  |  | ≥4-fold | 82 (44.09%) | 76 (39.18%) |
|  |  |  | (37.14, 51.27) | (32.58, 46.19) |
| 4 | Day 84 | ≥2-fold | 105 (56.45%) | 136 (70.10%) |
|  |  |  | (49.27, 63.38) | (63.32, 76.10) |
|  |  | ≥4-fold | 61 (32.80%) | 93 (47.94%) |
|  |  |  | (26.46, 39.83) | (41.02, 54.94) |
|  | Day 270 (Pre-Booster) | ≥2-fold | 44 (23.66%) | 51 (26.29%) |
|  |  |  | (18.12, 30.26) | (20.60, 32.90) |
|  |  | ≥4-fold | 14 (7.53%) | 13 (6.70%) |
|  |  |  | (4.54, 12.24) | (3.96, 11.13) |
|  | Day 300 (Booster Response) | ≥2-fold | 104 (55.91%) | 135 (69.59%) |
|  |  |  | (48.73, 62.86) | (62.79, 75.63) |
|  |  | ≥4-fold | 65 (34.95%) | 86 (44.33%) |
|  |  |  | (28.46, 42.04) | (37.52, 51.36) |
| 5 | Day 84 | ≥2-fold | 151 (81.18%) | 146 (75.26%) |
|  |  |  | (74.96, 86.15) | (68.73, 80.80) |
|  |  | ≥4-fold | 115 (61.83%) | 107 (55.15%) |
|  |  |  | (54.67, 68.50) | (48.12, 61.99) |
|  | Day 270 (Pre-Booster) | ≥2-fold | 94 (50.54%) | 87 (44.85%) |
|  |  |  | (43.41, 57.64) | (38.01, 51.88) |
|  |  | ≥4-fold | 49 (26.34%) | 45 (23.20%) |
|  |  |  | (20.54, 33.11) | (17.81, 29.62) |
|  | Day 300 (Booster Response) | ≥2-fold | 141 (75.81%) | 144 (74.23%) |
|  |  |  | (69.17, 81.40) | (67.64, 79.87) |
|  |  | ≥4-fold | 92 (49.46%) | 104 (53.61%) |
|  |  |  | (42.36, 56.59) | (46.59, 60.49) |
| 6A | Day 84 | ≥2-fold | 48 (25.81%) | 140 (72.16%) |
|  |  |  | (20.05, 32.54) | (65.47, 77.99) |
|  |  | ≥4-fold | 31 (16.67%) | 93 (47.94%) |
|  |  |  | (12.00, 22.69) | (41.02, 54.94) |
|  | Day 270 (Pre-Booster) | ≥2-fold | 63 (33.87%) | 97 (50.00%) |
|  |  |  | (27.46, 40.94) | (43.03, 56.97) |
|  |  | ≥4-fold | 40 (21.51%) | 51 (26.29%) |
|  |  |  | (16.21, 27.95) | (20.60, 32.90) |
|  | Day 300 (Booster Response) | ≥2-fold | 116 (62.37%) | 160 (82.47%) |
|  |  |  | (55.22, 69.01) | (76.51, 87.18) |
|  |  | ≥4-fold | 69 (37.10%) | 135 (69.59%) |
|  |  |  | (30.48, 44.23) | (62.79, 75.63) |
| 6B | Day 84 | ≥2-fold | 136 (73.12%) | 122 (62.89%) |
|  |  |  | (66.33, 78.97) | (55.90, 69.37) |
|  |  | ≥4-fold | 112 (60.22%) | 88 (45.36%) |
|  |  |  | (53.04, 66.97) | (38.51, 52.39) |
|  | Day 270 (Pre-Booster) | ≥2-fold | 128 (68.82%) | 104 (53.61%) |
|  |  |  | (61.84, 75.04) | (46.59, 60.49) |
|  |  | ≥4-fold | 103 (55.38%) | 70 (36.08%) |
|  |  |  | (48.20, 62.34) | (29.66, 43.05) |
|  | Day 300 (Booster Response) | ≥2-fold | 159 (85.48%) | 172 (88.66%) |
|  |  |  | (79.70, 89.83) | (83.43, 92.39) |
|  |  | ≥4-fold | 141 (75.81%) | 150 (77.32%) |
|  |  |  | (69.17, 81.40) | (70.93, 82.65) |
| 7F | Day 84 | ≥2-fold | 158 (84.95%) | 157 (80.93%) |
|  |  |  | (79.10, 89.37) | (74.82, 85.83) |
|  |  | ≥4-fold | 128 (68.82%) | 136 (70.10%) |
|  |  |  | (61.84, 75.04) | (63.32, 76.10) |
|  | Day 270 (Pre-Booster) | ≥2-fold | 100 (53.76%) | 101 (52.06%) |
|  |  |  | (46.59, 60.78) | (45.06, 58.98) |
|  |  | ≥4-fold | 65 (34.95%) | 61 (31.44%) |
|  |  |  | (28.46, 42.04) | (25.32, 38.28) |
|  | Day 300 (Booster Response) | ≥2-fold | 152 (81.72%) | 151 (77.84%) |
|  |  |  | (75.54, 86.61) | (71.48, 83.11) |
|  |  | ≥4-fold | 110 (59.14%) | 122 (62.89%) |
|  |  |  | (51.96, 65.95) | (55.90, 69.37) |
| 9V | Day 84 | ≥2-fold | 134 (72.04%) | 150 (77.32%) |
|  |  |  | (65.20, 78.00) | (70.93, 82.65) |
|  |  | ≥4-fold | 108 (58.06%) | 118 (60.82%) |
|  |  |  | (50.88, 64.92) | (53.81, 67.42) |
|  | Day 270 (Pre-Booster) | ≥2-fold | 82 (44.09%) | 74 (38.14%) |
|  |  |  | (37.14, 51.27) | (31.60, 45.15) |
|  |  | ≥4-fold | 42 (22.58%) | 36 (18.56%) |
|  |  |  | (17.16, 29.11) | (13.72, 24.62) |
|  | Day 300 (Booster Response) | ≥2-fold | 138 (74.19%) | 150 (77.32%) |
|  |  |  | (67.46, 79.95) | (70.93, 82.65) |
|  |  | ≥4-fold | 106 (56.99%) | 123 (63.40%) |
|  |  |  | (49.80, 63.89) | (56.42, 69.86) |
| 14 | Day 84 | ≥2-fold | 65 (34.95%) | 53 (27.32%) |
|  |  |  | (28.46, 42.04) | (21.54, 33.98) |
|  |  | ≥4-fold | 39 (20.97%) | 23 (11.86%) |
|  |  |  | (15.73, 27.38) | (8.03, 17.16) |
|  | Day 270 (Pre-Booster) | ≥2-fold | 42 (22.58%) | 36 (18.56%) |
|  |  |  | (17.16, 29.11) | (13.72, 24.62) |
|  |  | ≥4-fold | 16 (8.60%) | 9 (4.64%) |
|  |  |  | (5.36, 13.52) | (2.46, 8.58) |
|  | Day 300 (Booster Response) | ≥2-fold | 74 (39.78%) | 84 (43.30%) |
|  |  |  | (33.03, 46.96) | (36.52, 50.33) |
|  |  | ≥4-fold | 34 (18.28%) | 38 (19.59%) |
|  |  |  | (13.39, 24.46) | (14.62, 25.74) |
| 18C | Day 84 | ≥2-fold | 67 (36.02%) | 114 (58.76%) |
|  |  |  | (29.47, 43.14) | (51.73, 65.45) |
|  |  | ≥4-fold | 32 (17.20%) | 59 (30.41%) |
|  |  |  | (12.46, 23.28) | (24.37, 37.21) |
|  | Day 270 (Pre-Booster) | ≥2-fold | 43 (23.12%) | 38 (19.59%) |
|  |  |  | (17.64, 29.68) | (14.62, 25.74) |
|  |  | ≥4-fold | 21 (11.29%) | 15 (7.73%) |
|  |  |  | (7.50, 16.64) | (4.74, 12.36) |
|  | Day 300 (Booster Response) | ≥2-fold | 89 (47.85%) | 95 (48.97%) |
|  |  |  | (40.79, 55.00) | (42.02, 55.95) |
|  |  | ≥4-fold | 50 (26.88%) | 44 (22.68%) |
|  |  |  | (21.03, 33.67) | (17.35, 29.07) |
| 19F | Day 84 | ≥2-fold | 131 (70.43%) | 152 (78.35%) |
|  |  |  | (63.51, 76.52) | (72.03, 83.57) |
|  |  | ≥4-fold | 103 (55.38%) | 111 (57.22%) |
|  |  |  | (48.20, 62.34) | (50.18, 63.97) |
|  | Day 270 (Pre-Booster) | ≥2-fold | 90 (48.39%) | 77 (39.69%) |
|  |  |  | (41.31, 55.53) | (33.07, 46.71) |
|  |  | ≥4-fold | 60 (32.26%) | 38 (19.59%) |
|  |  |  | (25.96, 39.28) | (14.62, 25.74) |
|  | Day 300 (Booster Response) | ≥2-fold | 150 (80.65%) | 156 (80.41%) |
|  |  |  | (74.37, 85.68) | (74.26, 85.38) |
|  |  | ≥4-fold | 123 (66.13%) | 130 (67.01%) |
|  |  |  | (59.06, 72.54) | (60.12, 73.24) |
| 22F | Day 84 | ≥2-fold | 143 (76.88%) | -- |
|  |  |  | (70.32, 82.36) |  |
|  |  | ≥4-fold | 119 (63.98%) | -- |
|  |  |  | (56.86, 70.53) |  |
|  | Day 270 (Pre-Booster) | ≥2-fold | 123 (66.13%) | -- |
|  |  |  | (59.06, 72.54) |  |
|  |  | ≥4-fold | 79 (42.47%) | -- |
|  |  |  | (35.59, 49.66) |  |
|  | Day 300 (Booster Response) | ≥2-fold | 161 (86.56%) | -- |
|  |  |  | (80.91, 90.73) |  |
|  |  | ≥4-fold | 135 (72.58%) | -- |
|  |  |  | (65.76, 78.49) |  |
| 23F | Day 84 | ≥2-fold | 121 (65.05%) | 117 (60.31%) |
|  |  |  | (57.96, 71.54) | (53.29, 66.93) |
|  |  | ≥4-fold | 96 (51.61%) | 81 (41.75%) |
|  |  |  | (44.47, 58.69) | (35.04, 48.79) |
|  | Day 270 (Pre-Booster) | ≥2-fold | 80 (43.01%) | 63 (32.47%) |
|  |  |  | (36.11, 50.20) | (26.28, 39.35) |
|  |  | ≥4-fold | 49 (26.34%) | 41 (21.13%) |
|  |  |  | (20.54, 33.11) | (15.98, 27.41) |
|  | Day 300 (Booster Response) | ≥2-fold | 145 (77.96%) | 153 (78.87%) |
|  |  |  | (71.47, 83.32) | (72.59, 84.02) |
|  |  | ≥4-fold | 117 (62.90%) | 125 (64.43%) |
|  |  |  | (55.77, 69.52) | (57.48, 70.83) |
| 33F | Day 84 | ≥2-fold | 96 (51.61%) | -- |
|  |  |  | (44.47, 58.69) |  |
|  |  | ≥4-fold | 66 (35.48%) | -- |
|  |  |  | (28.97, 42.59) |  |
|  | Day 270 (Pre-Booster) | ≥2-fold | 100 (53.76%) | -- |
|  |  |  | (46.59, 60.78) |  |
|  |  | ≥4-fold | 60 (32.26%) | -- |
|  |  |  | (25.96, 39.28) |  |
|  | Day 300 (Booster Response) | ≥2-fold | 145 (77.96%) | -- |
|  |  |  | (71.47, 83.32) |  |
|  |  | ≥4-fold | 114 (61.29%) | -- |
|  |  |  | (54.13, 67.99) |  |

**Appendix 9**

**Figure: Reverse cumulative distribution (RCD) curves of anti-PnCPS antibody concentrations by serotype for all common vaccine serotypes in both groups. (PP Population)**

BECT056 - RCDC_Serotype 1

|  | ST1-BE-PCV14 (Day 84) |
| --- | --- |
| 47.996 | 0.0052356 |
| 25.506 | 0.0104712 |
| 18.947 | 0.0157068 |
| 18.04 | 0.0209424 |
| 17.446 | 0.026178 |
| 16.311 | 0.0314136 |
| 15.933 | 0.0366492 |
| 14.171 | 0.0418848 |
| 11.846 | 0.0471204 |
| 11.414 | 0.052356 |
| 11.167 | 0.0575916 |
| 10.714 | 0.0628272 |
| 10.423 | 0.0680628 |
| 8.01 | 0.0732984 |
| 7.912 | 0.078534 |
| 7.486 | 0.0837696 |
| 6.595 | 0.0890052 |
| 5.953 | 0.0942408 |
| 5.946 | 0.0994764 |
| 5.923 | 0.104712 |
| 5.909 | 0.109948 |
| 5.58 | 0.115183 |
| 5.498 | 0.120419 |
| 5.133 | 0.125654 |
| 4.972 | 0.13089 |
| 4.964 | 0.136126 |
| 4.865 | 0.141361 |
| 4.812 | 0.146597 |
| 4.685 | 0.151832 |
| 4.674 | 0.157068 |
| 4.546 | 0.162304 |
| 4.505 | 0.167539 |
| 4.404 | 0.172775 |
| 4.382 | 0.17801 |
| 4.319 | 0.183246 |
| 4.311 | 0.188482 |
| 4.274 | 0.193717 |
| 4.11 | 0.198953 |
| 4.077 | 0.204188 |
| 4.047 | 0.209424 |
| 4.044 | 0.21466 |
| 4.024 | 0.219895 |
| 3.996 | 0.225131 |
| 3.902 | 0.230366 |
| 3.891 | 0.235602 |
| 3.875 | 0.240838 |
| 3.856 | 0.246073 |
| 3.795 | 0.251309 |
| 3.719 | 0.256545 |
| 3.516 | 0.26178 |
| 3.483 | 0.267016 |
| 3.463 | 0.272251 |
| 3.345 | 0.277487 |
| 3.334 | 0.282723 |
| 3.306 | 0.287958 |
| 3.27 | 0.293194 |
| 3.23 | 0.298429 |
| 3.204 | 0.303665 |
| 3.193 | 0.308901 |
| 3.155 | 0.314136 |
| 3.072 | 0.319372 |
| 3.035 | 0.324607 |
| 2.956 | 0.329843 |
| 2.916 | 0.335079 |
| 2.911 | 0.340314 |
| 2.907 | 0.34555 |
| 2.798 | 0.350785 |
| 2.78 | 0.356021 |
| 2.723 | 0.361257 |
| 2.675 | 0.366492 |
| 2.589 | 0.371728 |
| 2.577 | 0.376963 |
| 2.57 | 0.382199 |
| 2.565 | 0.387435 |
| 2.455 | 0.39267 |
| 2.402 | 0.397906 |
| 2.305 | 0.403141 |
| 2.201 | 0.408377 |
| 2.13 | 0.413613 |
| 2.12 | 0.418848 |
| 2.112 | 0.424084 |
| 2.079 | 0.429319 |
| 2.073 | 0.434555 |
| 2.052 | 0.439791 |
| 2.042 | 0.445026 |
| 2.018 | 0.450262 |
| 2.001 | 0.455497 |
| 1.971 | 0.460733 |
| 1.971 | 0.465969 |
| 1.958 | 0.471204 |
| 1.944 | 0.47644 |
| 1.889 | 0.481675 |
| 1.886 | 0.486911 |
| 1.877 | 0.492147 |
| 1.872 | 0.497382 |
| 1.853 | 0.502618 |
| 1.811 | 0.507853 |
| 1.795 | 0.513089 |
| 1.768 | 0.518325 |
| 1.749 | 0.52356 |
| 1.707 | 0.528796 |
| 1.704 | 0.534031 |
| 1.702 | 0.539267 |
| 1.649 | 0.544503 |
| 1.643 | 0.549738 |
| 1.621 | 0.554974 |
| 1.613 | 0.560209 |
| 1.61 | 0.565445 |
| 1.608 | 0.570681 |
| 1.563 | 0.575916 |
| 1.559 | 0.581152 |
| 1.556 | 0.586387 |
| 1.541 | 0.591623 |
| 1.5 | 0.596859 |
| 1.457 | 0.602094 |
| 1.456 | 0.60733 |
| 1.443 | 0.612565 |
| 1.423 | 0.617801 |
| 1.416 | 0.623037 |
| 1.398 | 0.628272 |
| 1.392 | 0.633508 |
| 1.335 | 0.638743 |
| 1.33 | 0.643979 |
| 1.31 | 0.649215 |
| 1.303 | 0.65445 |
| 1.288 | 0.659686 |
| 1.268 | 0.664921 |
| 1.257 | 0.670157 |
| 1.247 | 0.675393 |
| 1.227 | 0.680628 |
| 1.221 | 0.685864 |
| 1.216 | 0.691099 |
| 1.192 | 0.696335 |
| 1.189 | 0.701571 |
| 1.13 | 0.706806 |
| 1.106 | 0.712042 |
| 1.055 | 0.717277 |
| 1.02 | 0.722513 |
| 1.015 | 0.727749 |
| 0.992 | 0.732984 |
| 0.99 | 0.73822 |
| 0.988 | 0.743455 |
| 0.972 | 0.748691 |
| 0.891 | 0.753927 |
| 0.886 | 0.759162 |
| 0.884 | 0.764398 |
| 0.878 | 0.769634 |
| 0.875 | 0.774869 |
| 0.868 | 0.780105 |
| 0.863 | 0.78534 |
| 0.854 | 0.790576 |
| 0.85 | 0.795812 |
| 0.842 | 0.801047 |
| 0.823 | 0.806283 |
| 0.81 | 0.811518 |
| 0.797 | 0.816754 |
| 0.776 | 0.82199 |
| 0.756 | 0.827225 |
| 0.751 | 0.832461 |
| 0.746 | 0.837696 |
| 0.745 | 0.842932 |
| 0.739 | 0.848168 |
| 0.727 | 0.853403 |
| 0.689 | 0.858639 |
| 0.686 | 0.863874 |
| 0.658 | 0.86911 |
| 0.655 | 0.874346 |
| 0.653 | 0.879581 |
| 0.65 | 0.884817 |
| 0.647 | 0.890052 |
| 0.642 | 0.895288 |
| 0.623 | 0.900524 |
| 0.607 | 0.905759 |
| 0.59 | 0.910995 |
| 0.575 | 0.91623 |
| 0.563 | 0.921466 |
| 0.56 | 0.926702 |
| 0.558 | 0.931937 |
| 0.545 | 0.937173 |
| 0.507 | 0.942408 |
| 0.477 | 0.947644 |
| 0.474 | 0.95288 |
| 0.46 | 0.958115 |
| 0.455 | 0.963351 |
| 0.448 | 0.968586 |
| 0.375 | 0.973822 |
| 0.365 | 0.979058 |
| 0.359 | 0.984293 |
| 0.34 | 0.989529 |
| 0.299 | 0.994764 |
| 0.22 | 1 |

|  | ST1-Prevenar 13 (Day 84) |
| --- | --- |
| 127.844 | 0.00502513 |
| 118.462 | 0.0100503 |
| 32.973 | 0.0150754 |
| 26.138 | 0.0201005 |
| 24.021 | 0.0251256 |
| 18.014 | 0.0301508 |
| 15.67 | 0.0351759 |
| 14.483 | 0.040201 |
| 13.827 | 0.0452261 |
| 13.443 | 0.0502513 |
| 12.996 | 0.0552764 |
| 12.559 | 0.0603015 |
| 12.538 | 0.0653266 |
| 12.391 | 0.0703518 |
| 10.717 | 0.0753769 |
| 10.244 | 0.080402 |
| 9.792 | 0.0854271 |
| 9.74 | 0.0904523 |
| 9.35 | 0.0954774 |
| 9.169 | 0.100503 |
| 8.596 | 0.105528 |
| 8.459 | 0.110553 |
| 8.025 | 0.115578 |
| 7.904 | 0.120603 |
| 7.904 | 0.125628 |
| 7.872 | 0.130653 |
| 7.78 | 0.135678 |
| 7.718 | 0.140704 |
| 7.477 | 0.145729 |
| 7.323 | 0.150754 |
| 7.268 | 0.155779 |
| 7.257 | 0.160804 |
| 7.223 | 0.165829 |
| 7.219 | 0.170854 |
| 6.759 | 0.175879 |
| 6.734 | 0.180905 |
| 6.729 | 0.18593 |
| 6.579 | 0.190955 |
| 6.538 | 0.19598 |
| 6.508 | 0.201005 |
| 6.394 | 0.20603 |
| 6.342 | 0.211055 |
| 6.111 | 0.21608 |
| 6.09 | 0.221106 |
| 6.027 | 0.226131 |
| 5.981 | 0.231156 |
| 5.978 | 0.236181 |
| 5.871 | 0.241206 |
| 5.796 | 0.246231 |
| 5.721 | 0.251256 |
| 5.643 | 0.256281 |
| 5.622 | 0.261307 |
| 5.606 | 0.266332 |
| 5.574 | 0.271357 |
| 5.44 | 0.276382 |
| 5.381 | 0.281407 |
| 5.242 | 0.286432 |
| 5.223 | 0.291457 |
| 5.18 | 0.296482 |
| 5.161 | 0.301508 |
| 4.885 | 0.306533 |
| 4.87 | 0.311558 |
| 4.781 | 0.316583 |
| 4.778 | 0.321608 |
| 4.736 | 0.326633 |
| 4.722 | 0.331658 |
| 4.711 | 0.336683 |
| 4.655 | 0.341709 |
| 4.622 | 0.346734 |
| 4.596 | 0.351759 |
| 4.359 | 0.356784 |
| 4.359 | 0.361809 |
| 4.354 | 0.366834 |
| 4.339 | 0.371859 |
| 4.187 | 0.376884 |
| 4.15 | 0.38191 |
| 4.147 | 0.386935 |
| 4.113 | 0.39196 |
| 4.074 | 0.396985 |
| 4.03 | 0.40201 |
| 4.027 | 0.407035 |
| 4.021 | 0.41206 |
| 4 | 0.417085 |
| 3.947 | 0.422111 |
| 3.915 | 0.427136 |
| 3.702 | 0.432161 |
| 3.686 | 0.437186 |
| 3.681 | 0.442211 |
| 3.678 | 0.447236 |
| 3.628 | 0.452261 |
| 3.616 | 0.457286 |
| 3.581 | 0.462312 |
| 3.564 | 0.467337 |
| 3.564 | 0.472362 |
| 3.534 | 0.477387 |
| 3.484 | 0.482412 |
| 3.438 | 0.487437 |
| 3.355 | 0.492462 |
| 3.302 | 0.497487 |
| 3.275 | 0.502513 |
| 3.104 | 0.507538 |
| 3.066 | 0.512563 |
| 3.065 | 0.517588 |
| 3.014 | 0.522613 |
| 6.33455 | 0.527638 |
| 2.923 | 0.532663 |
| 2.916 | 0.537688 |
| 2.894 | 0.542714 |
| 2.877 | 0.547739 |
| 2.86 | 0.552764 |
| 2.848 | 0.557789 |
| 2.806 | 0.562814 |
| 2.788 | 0.567839 |
| 2.781 | 0.572864 |
| 2.738 | 0.577889 |
| 2.655 | 0.582915 |
| 2.613 | 0.58794 |
| 2.574 | 0.592965 |
| 2.518 | 0.59799 |
| 2.504 | 0.603015 |
| 2.372 | 0.60804 |
| 2.339 | 0.613065 |
| 2.334 | 0.61809 |
| 2.314 | 0.623116 |
| 2.285 | 0.628141 |
| 2.262 | 0.633166 |
| 2.233 | 0.638191 |
| 2.182 | 0.643216 |
| 2.157 | 0.648241 |
| 2.157 | 0.653266 |
| 2.15 | 0.658291 |
| 2.147 | 0.663317 |
| 2.045 | 0.668342 |
| 2.044 | 0.673367 |
| 2.036 | 0.678392 |
| 1.951 | 0.683417 |
| 1.904 | 0.688442 |
| 1.9 | 0.693467 |
| 1.893 | 0.698492 |
| 1.847 | 0.703518 |
| 1.795 | 0.708543 |
| 1.781 | 0.713568 |
| 1.768 | 0.718593 |
| 1.742 | 0.723618 |
| 1.696 | 0.728643 |
| 1.67 | 0.733668 |
| 1.65 | 0.738693 |
| 1.635 | 0.743719 |
| 1.628 | 0.748744 |
| 1.615 | 0.753769 |
| 1.576 | 0.758794 |
| 1.542 | 0.763819 |
| 1.538 | 0.768844 |
| 1.531 | 0.773869 |
| 1.506 | 0.778894 |
| 1.468 | 0.78392 |
| 1.463 | 0.788945 |
| 1.454 | 0.79397 |
| 1.439 | 0.798995 |
| 1.434 | 0.80402 |
| 1.336 | 0.809045 |
| 1.289 | 0.81407 |
| 1.283 | 0.819095 |
| 1.174 | 0.824121 |
| 1.168 | 0.829146 |
| 1.149 | 0.834171 |
| 1.09 | 0.839196 |
| 1.083 | 0.844221 |
| 1.08 | 0.849246 |
| 1.049 | 0.854271 |
| 1.02 | 0.859296 |
| 1.019 | 0.864322 |
| 0.999 | 0.869347 |
| 0.982 | 0.874372 |
| 0.935 | 0.879397 |
| 0.923 | 0.884422 |
| 0.898 | 0.889447 |
| 0.89 | 0.894472 |
| 0.888 | 0.899497 |
| 0.811 | 0.904523 |
| 0.781 | 0.909548 |
| 0.761 | 0.914573 |
| 0.742 | 0.919598 |
| 0.74 | 0.924623 |
| 0.701 | 0.929648 |
| 0.606 | 0.934673 |
| 0.582 | 0.939698 |
| 0.526 | 0.944724 |
| 0.514 | 0.949749 |
| 0.507 | 0.954774 |
| 0.484 | 0.959799 |
| 0.479 | 0.964824 |
| 0.477 | 0.969849 |
| 0.369 | 0.974874 |
| 0.361 | 0.979899 |
| 0.307 | 0.984925 |
| 0.292 | 0.98995 |
| 0.281 | 0.994975 |
| 0.204 | 1 |

|  | ST1-BE-PCV14 (Day 300) |
| --- | --- |
| 98.443 | 0.00531915 |
| 32.858 | 0.0106383 |
| 31.283 | 0.0159574 |
| 21.838 | 0.0212766 |
| 15.918 | 0.0265957 |
| 14.61 | 0.0319149 |
| 14.551 | 0.037234 |
| 14.272 | 0.0425532 |
| 12.371 | 0.0478723 |
| 12.149 | 0.0531915 |
| 11.863 | 0.0585106 |
| 11.782 | 0.0638298 |
| 11.765 | 0.0691489 |
| 11.693 | 0.0744681 |
| 11.693 | 0.0797872 |
| 11.321 | 0.0851064 |
| 11.275 | 0.0904255 |
| 11.067 | 0.0957447 |
| 10.936 | 0.101064 |
| 10.84 | 0.106383 |
| 10.766 | 0.111702 |
| 10.743 | 0.117021 |
| 10.011 | 0.12234 |
| 9.866 | 0.12766 |
| 9.841 | 0.132979 |
| 9.184 | 0.138298 |
| 9.183 | 0.143617 |
| 9.101 | 0.148936 |
| 8.868 | 0.154255 |
| 8.238 | 0.159574 |
| 7.994 | 0.164894 |
| 7.821 | 0.170213 |
| 7.619 | 0.175532 |
| 7.583 | 0.180851 |
| 7.343 | 0.18617 |
| 7.277 | 0.191489 |
| 7.047 | 0.196809 |
| 6.859 | 0.202128 |
| 6.782 | 0.207447 |
| 6.733 | 0.212766 |
| 6.5 | 0.218085 |
| 6.44 | 0.223404 |
| 6.327 | 0.228723 |
| 6.324 | 0.234043 |
| 6.297 | 0.239362 |
| 6.292 | 0.244681 |
| 6.211 | 0.25 |
| 6.186 | 0.255319 |
| 6.143 | 0.260638 |
| 6.128 | 0.265957 |
| 6.125 | 0.271277 |
| 6.121 | 0.276596 |
| 5.826 | 0.281915 |
| 5.815 | 0.287234 |
| 5.797 | 0.292553 |
| 5.789 | 0.297872 |
| 5.749 | 0.303191 |
| 5.743 | 0.308511 |
| 5.712 | 0.31383 |
| 5.584 | 0.319149 |
| 5.528 | 0.324468 |
| 5.404 | 0.329787 |
| 5.392 | 0.335106 |
| 5.368 | 0.340426 |
| 5.36 | 0.345745 |
| 5.357 | 0.351064 |
| 5.305 | 0.356383 |
| 5.303 | 0.361702 |
| 5.277 | 0.367021 |
| 5.134 | 0.37234 |
| 4.968 | 0.37766 |
| 4.791 | 0.382979 |
| 4.767 | 0.388298 |
| 4.693 | 0.393617 |
| 4.639 | 0.398936 |
| 4.623 | 0.404255 |
| 4.394 | 0.409574 |
| 4.37 | 0.414894 |
| 4.369 | 0.420213 |
| 4.362 | 0.425532 |
| 4.311 | 0.430851 |
| 4.285 | 0.43617 |
| 4.227 | 0.441489 |
| 4.199 | 0.446809 |
| 4.189 | 0.452128 |
| 4.061 | 0.457447 |
| 3.764 | 0.462766 |
| 3.751 | 0.468085 |
| 3.676 | 0.473404 |
| 3.654 | 0.478723 |
| 3.612 | 0.484043 |
| 3.591 | 0.489362 |
| 3.561 | 0.494681 |
| 3.545 | 0.5 |
| 3.539 | 0.505319 |
| 3.52 | 0.510638 |
| 3.49 | 0.515957 |
| 3.456 | 0.521277 |
| 3.354 | 0.526596 |
| 3.339 | 0.531915 |
| 3.304 | 0.537234 |
| 3.257 | 0.542553 |
| 3.245 | 0.547872 |
| 3.243 | 0.553191 |
| 3.216 | 0.558511 |
| 3.193 | 0.56383 |
| 3.191 | 0.569149 |
| 3.138 | 0.574468 |
| 3.087 | 0.579787 |
| 3.07 | 0.585106 |
| 3.066 | 0.590426 |
| 2.92 | 0.595745 |
| 2.901 | 0.601064 |
| 2.856 | 0.606383 |
| 2.846 | 0.611702 |
| 2.804 | 0.617021 |
| 2.767 | 0.62234 |
| 2.767 | 0.62766 |
| 2.666 | 0.632979 |
| 2.638 | 0.638298 |
| 2.626 | 0.643617 |
| 2.558 | 0.648936 |
| 2.54 | 0.654255 |
| 2.525 | 0.659574 |
| 2.467 | 0.664894 |
| 2.448 | 0.670213 |
| 2.331 | 0.675532 |
| 2.324 | 0.680851 |
| 2.308 | 0.68617 |
| 2.287 | 0.691489 |
| 2.271 | 0.696809 |
| 2.25 | 0.702128 |
| 2.204 | 0.707447 |
| 2.188 | 0.712766 |
| 2.169 | 0.718085 |
| 2.118 | 0.723404 |
| 1.958 | 0.728723 |
| 1.915 | 0.734043 |
| 1.892 | 0.739362 |
| 1.87 | 0.744681 |
| 1.814 | 0.75 |
| 1.779 | 0.755319 |
| 1.719 | 0.760638 |
| 1.688 | 0.765957 |
| 1.683 | 0.771277 |
| 1.665 | 0.776596 |
| 1.663 | 0.781915 |
| 1.628 | 0.787234 |
| 1.628 | 0.792553 |
| 1.582 | 0.797872 |
| 1.579 | 0.803191 |
| 1.574 | 0.808511 |
| 1.569 | 0.81383 |
| 1.559 | 0.819149 |
| 1.534 | 0.824468 |
| 1.51 | 0.829787 |
| 1.355 | 0.835106 |
| 1.35 | 0.840426 |
| 1.157 | 0.845745 |
| 1.095 | 0.851064 |
| 1.086 | 0.856383 |
| 1.081 | 0.861702 |
| 1.068 | 0.867021 |
| 1.044 | 0.87234 |
| 1.013 | 0.87766 |
| 1.007 | 0.882979 |
| 0.973 | 0.888298 |
| 0.955 | 0.893617 |
| 0.953 | 0.898936 |
| 0.943 | 0.904255 |
| 0.931 | 0.909574 |
| 0.914 | 0.914894 |
| 0.901 | 0.920213 |
| 0.874 | 0.925532 |
| 0.869 | 0.930851 |
| 0.868 | 0.93617 |
| 0.834 | 0.941489 |
| 0.791 | 0.946809 |
| 0.693 | 0.952128 |
| 0.657 | 0.957447 |
| 0.592 | 0.962766 |
| 0.485 | 0.968085 |
| 0.456 | 0.973404 |
| 0.388 | 0.978723 |
| 0.264 | 0.984043 |
| 0.212 | 0.989362 |
| 0.163 | 0.994681 |
| 0.134 | 1 |

|  | ST1-Prevenar 13 (Day 300) |
| --- | --- |
| 37.422 | 0.00526316 |
| 37.188 | 0.0105263 |
| 36.942 | 0.0157895 |
| 23.895 | 0.0210526 |
| 22.625 | 0.0263158 |
| 20.184 | 0.0315789 |
| 19.526 | 0.0368421 |
| 18.04 | 0.0421053 |
| 17.534 | 0.0473684 |
| 17.24 | 0.0526316 |
| 16.714 | 0.0578947 |
| 16.618 | 0.0631579 |
| 16.291 | 0.0684211 |
| 15.339 | 0.0736842 |
| 14.899 | 0.0789474 |
| 14.837 | 0.0842105 |
| 14.693 | 0.0894737 |
| 14.615 | 0.0947368 |
| 14.575 | 0.1 |
| 14.315 | 0.105263 |
| 14.281 | 0.110526 |
| 14.21 | 0.115789 |
| 13.682 | 0.121053 |
| 13.446 | 0.126316 |
| 12.379 | 0.131579 |
| 12.064 | 0.136842 |
| 11.139 | 0.142105 |
| 10.91 | 0.147368 |
| 10.774 | 0.152632 |
| 10.72 | 0.157895 |
| 10.546 | 0.163158 |
| 10.459 | 0.168421 |
| 10.202 | 0.173684 |
| 9.964 | 0.178947 |
| 9.922 | 0.184211 |
| 9.807 | 0.189474 |
| 9.721 | 0.194737 |
| 9.658 | 0.2 |
| 9.52 | 0.205263 |
| 8.946 | 0.210526 |
| 8.639 | 0.215789 |
| 8.638 | 0.221053 |
| 8.284 | 0.226316 |
| 8.196 | 0.231579 |
| 8.064 | 0.236842 |
| 8.056 | 0.242105 |
| 8.013 | 0.247368 |
| 7.739 | 0.252632 |
| 7.667 | 0.257895 |
| 7.648 | 0.263158 |
| 7.616 | 0.268421 |
| 7.553 | 0.273684 |
| 7.45 | 0.278947 |
| 7.105 | 0.284211 |
| 7.035 | 0.289474 |
| 6.969 | 0.294737 |
| 6.957 | 0.3 |
| 6.58 | 0.305263 |
| 6.567 | 0.310526 |
| 6.452 | 0.315789 |
| 6.294 | 0.321053 |
| 6.289 | 0.326316 |
| 6.243 | 0.331579 |
| 6.235 | 0.336842 |
| 6.126 | 0.342105 |
| 6.06 | 0.347368 |
| 5.852 | 0.352632 |
| 5.781 | 0.357895 |
| 5.712 | 0.363158 |
| 5.637 | 0.368421 |
| 5.616 | 0.373684 |
| 5.584 | 0.378947 |
| 5.512 | 0.384211 |
| 5.455 | 0.389474 |
| 5.349 | 0.394737 |
| 5.346 | 0.4 |
| 5.3 | 0.405263 |
| 5.288 | 0.410526 |
| 5.175 | 0.415789 |
| 5.168 | 0.421053 |
| 5.162 | 0.426316 |
| 5.138 | 0.431579 |
| 5.136 | 0.436842 |
| 5.047 | 0.442105 |
| 4.991 | 0.447368 |
| 4.934 | 0.452632 |
| 4.931 | 0.457895 |
| 4.922 | 0.463158 |
| 4.728 | 0.468421 |
| 4.709 | 0.473684 |
| 4.606 | 0.478947 |
| 4.602 | 0.484211 |
| 4.582 | 0.489474 |
| 4.448 | 0.494737 |
| 4.441 | 0.5 |
| 4.405 | 0.505263 |
| 4.359 | 0.510526 |
| 4.268 | 0.515789 |
| 4.137 | 0.521053 |
| 4.106 | 0.526316 |
| 4.079 | 0.531579 |
| 4.064 | 0.536842 |
| 4.002 | 0.542105 |
| 3.924 | 0.547368 |
| 3.857 | 0.552632 |
| 3.825 | 0.557895 |
| 3.727 | 0.563158 |
| 3.678 | 0.568421 |
| 3.651 | 0.573684 |
| 3.606 | 0.578947 |
| 3.561 | 0.584211 |
| 3.541 | 0.589474 |
| 3.541 | 0.594737 |
| 3.498 | 0.6 |
| 3.424 | 0.605263 |
| 3.418 | 0.610526 |
| 3.409 | 0.615789 |
| 3.365 | 0.621053 |
| 3.355 | 0.626316 |
| 3.326 | 0.631579 |
| 3.313 | 0.636842 |
| 3.297 | 0.642105 |
| 3.247 | 0.647368 |
| 3.214 | 0.652632 |
| 3.201 | 0.657895 |
| 3.199 | 0.663158 |
| 3.115 | 0.668421 |
| 3.108 | 0.673684 |
| 3.098 | 0.678947 |
| 3.076 | 0.684211 |
| 3.055 | 0.689474 |
| 3.044 | 0.694737 |
| 3.026 | 0.7 |
| 2.992 | 0.705263 |
| 2.973 | 0.710526 |
| 2.951 | 0.715789 |
| 2.941 | 0.721053 |
| 2.914 | 0.726316 |
| 2.91 | 0.731579 |
| 2.908 | 0.736842 |
| 2.677 | 0.742105 |
| 2.657 | 0.747368 |
| 2.508 | 0.752632 |
| 2.471 | 0.757895 |
| 2.447 | 0.763158 |
| 2.392 | 0.768421 |
| 2.328 | 0.773684 |
| 2.262 | 0.778947 |
| 2.257 | 0.784211 |
| 2.237 | 0.789474 |
| 2.236 | 0.794737 |
| 2.173 | 0.8 |
| 2.169 | 0.805263 |
| 2.128 | 0.810526 |
| 2.001 | 0.815789 |
| 1.981 | 0.821053 |
| 1.867 | 0.826316 |
| 1.847 | 0.831579 |
| 1.821 | 0.836842 |
| 1.794 | 0.842105 |
| 1.707 | 0.847368 |
| 1.695 | 0.852632 |
| 1.647 | 0.857895 |
| 1.618 | 0.863158 |
| 1.561 | 0.868421 |
| 1.518 | 0.873684 |
| 1.482 | 0.878947 |
| 1.458 | 0.884211 |
| 1.452 | 0.889474 |
| 1.409 | 0.894737 |
| 1.317 | 0.9 |
| 1.307 | 0.905263 |
| 1.304 | 0.910526 |
| 1.253 | 0.915789 |
| 1.251 | 0.921053 |
| 1.184 | 0.926316 |
| 1.117 | 0.931579 |
| 0.856 | 0.936842 |
| 0.847 | 0.942105 |
| 0.819 | 0.947368 |
| 0.57 | 0.952632 |
| 0.5 | 0.957895 |
| 0.474 | 0.963158 |
| 0.422 | 0.968421 |
| 0.378 | 0.973684 |
| 0.27 | 0.978947 |
| 0.254 | 0.984211 |
| 0.246 | 0.989474 |
| 0.21 | 0.994737 |
| 0.192 | 1 |

BECT056 - RCDC_Serotype 3

|  | ST3-BE-PCV14 (Day 84) |
| --- | --- |
| 6.057 | 0.0052356 |
| 4.917 | 0.0104712 |
| 3.697 | 0.0157068 |
| 3.656 | 0.0209424 |
| 2.549 | 0.026178 |
| 2.262 | 0.0314136 |
| 2.188 | 0.0366492 |
| 2.083 | 0.0418848 |
| 2.059 | 0.0471204 |
| 2.023 | 0.052356 |
| 2.011 | 0.0575916 |
| 1.968 | 0.0628272 |
| 1.945 | 0.0680628 |
| 1.943 | 0.0732984 |
| 1.94 | 0.078534 |
| 1.833 | 0.0837696 |
| 1.72 | 0.0890052 |
| 1.699 | 0.0942408 |
| 1.699 | 0.0994764 |
| 1.665 | 0.104712 |
| 1.554 | 0.109948 |
| 1.546 | 0.115183 |
| 1.476 | 0.120419 |
| 1.47 | 0.125654 |
| 1.468 | 0.13089 |
| 1.465 | 0.136126 |
| 1.402 | 0.141361 |
| 1.355 | 0.146597 |
| 1.337 | 0.151832 |
| 1.306 | 0.157068 |
| 1.293 | 0.162304 |
| 1.283 | 0.167539 |
| 1.279 | 0.172775 |
| 1.248 | 0.17801 |
| 1.244 | 0.183246 |
| 1.222 | 0.188482 |
| 1.209 | 0.193717 |
| 1.202 | 0.198953 |
| 1.188 | 0.204188 |
| 1.152 | 0.209424 |
| 1.133 | 0.21466 |
| 1.112 | 0.219895 |
| 1.052 | 0.225131 |
| 1.047 | 0.230366 |
| 1.026 | 0.235602 |
| 1.015 | 0.240838 |
| 0.995 | 0.246073 |
| 0.99 | 0.251309 |
| 0.988 | 0.256545 |
| 0.984 | 0.26178 |
| 0.981 | 0.267016 |
| 0.94 | 0.272251 |
| 0.926 | 0.277487 |
| 0.916 | 0.282723 |
| 0.912 | 0.287958 |
| 0.895 | 0.293194 |
| 0.893 | 0.298429 |
| 0.89 | 0.303665 |
| 0.886 | 0.308901 |
| 0.884 | 0.314136 |
| 0.875 | 0.319372 |
| 0.856 | 0.324607 |
| 0.843 | 0.329843 |
| 0.843 | 0.335079 |
| 0.842 | 0.340314 |
| 0.836 | 0.34555 |
| 0.835 | 0.350785 |
| 0.809 | 0.356021 |
| 0.807 | 0.361257 |
| 0.798 | 0.366492 |
| 0.788 | 0.371728 |
| 0.767 | 0.376963 |
| 0.759 | 0.382199 |
| 0.759 | 0.387435 |
| 0.744 | 0.39267 |
| 0.743 | 0.397906 |
| 0.742 | 0.403141 |
| 0.726 | 0.408377 |
| 0.721 | 0.413613 |
| 0.712 | 0.418848 |
| 0.709 | 0.424084 |
| 0.69 | 0.429319 |
| 0.68 | 0.434555 |
| 0.671 | 0.439791 |
| 0.67 | 0.445026 |
| 0.669 | 0.450262 |
| 0.668 | 0.455497 |
| 0.666 | 0.460733 |
| 0.665 | 0.465969 |
| 0.659 | 0.471204 |
| 0.644 | 0.47644 |
| 0.64 | 0.481675 |
| 0.634 | 0.486911 |
| 0.63 | 0.492147 |
| 0.622 | 0.497382 |
| 0.621 | 0.502618 |
| 0.617 | 0.507853 |
| 0.61 | 0.513089 |
| 0.608 | 0.518325 |
| 0.608 | 0.52356 |
| 0.601 | 0.528796 |
| 0.574 | 0.534031 |
| 0.571 | 0.539267 |
| 0.561 | 0.544503 |
| 0.56 | 0.549738 |
| 0.557 | 0.554974 |
| 0.546 | 0.560209 |
| 0.542 | 0.565445 |
| 0.541 | 0.570681 |
| 0.539 | 0.575916 |
| 0.536 | 0.581152 |
| 0.536 | 0.586387 |
| 0.529 | 0.591623 |
| 0.524 | 0.596859 |
| 0.521 | 0.602094 |
| 0.508 | 0.60733 |
| 0.506 | 0.612565 |
| 0.503 | 0.617801 |
| 0.487 | 0.623037 |
| 0.48 | 0.628272 |
| 0.478 | 0.633508 |
| 0.466 | 0.638743 |
| 0.466 | 0.643979 |
| 0.462 | 0.649215 |
| 0.456 | 0.65445 |
| 0.446 | 0.659686 |
| 0.434 | 0.664921 |
| 0.434 | 0.670157 |
| 0.423 | 0.675393 |
| 0.419 | 0.680628 |
| 0.401 | 0.685864 |
| 0.392 | 0.691099 |
| 0.384 | 0.696335 |
| 0.381 | 0.701571 |
| 0.379 | 0.706806 |
| 0.365 | 0.712042 |
| 0.356 | 0.717277 |
| 0.342 | 0.722513 |
| 0.338 | 0.727749 |
| 0.337 | 0.732984 |
| 0.324 | 0.73822 |
| 0.32 | 0.743455 |
| 0.315 | 0.748691 |
| 0.314 | 0.753927 |
| 0.313 | 0.759162 |
| 0.31 | 0.764398 |
| 0.301 | 0.769634 |
| 0.296 | 0.774869 |
| 0.295 | 0.780105 |
| 0.293 | 0.78534 |
| 0.282 | 0.790576 |
| 0.274 | 0.795812 |
| 0.269 | 0.801047 |
| 0.269 | 0.806283 |
| 0.268 | 0.811518 |
| 0.257 | 0.816754 |
| 0.255 | 0.82199 |
| 0.254 | 0.827225 |
| 0.25 | 0.832461 |
| 0.248 | 0.837696 |
| 0.242 | 0.842932 |
| 0.241 | 0.848168 |
| 0.238 | 0.853403 |
| 0.236 | 0.858639 |
| 0.229 | 0.863874 |
| 0.228 | 0.86911 |
| 0.226 | 0.874346 |
| 0.21 | 0.879581 |
| 0.203 | 0.884817 |
| 0.202 | 0.890052 |
| 0.198 | 0.895288 |
| 0.189 | 0.900524 |
| 0.187 | 0.905759 |
| 0.185 | 0.910995 |
| 0.183 | 0.91623 |
| 0.167 | 0.921466 |
| 0.164 | 0.926702 |
| 0.151 | 0.931937 |
| 0.15 | 0.937173 |
| 0.149 | 0.942408 |
| 0.146 | 0.947644 |
| 0.141 | 0.95288 |
| 0.136 | 0.958115 |
| 0.132 | 0.963351 |
| 0.127 | 0.968586 |
| 0.107 | 0.973822 |
| 0.102 | 0.979058 |
| 0.101 | 0.984293 |
| 0.092 | 0.989529 |
| 0.08 | 0.994764 |
| 0.059 | 1 |

|  | ST3-Prevenar 13 (Day 84) |
| --- | --- |
| 4.703 | 0.00505051 |
| 3.472 | 0.010101 |
| 3.338 | 0.0151515 |
| 3.083 | 0.020202 |
| 3.032 | 0.0252525 |
| 2.522 | 0.030303 |
| 2.421 | 0.0353535 |
| 2.267 | 0.040404 |
| 2.2 | 0.0454545 |
| 2.196 | 0.0505051 |
| 2.132 | 0.0555556 |
| 2.054 | 0.0606061 |
| 1.951 | 0.0656566 |
| 1.741 | 0.0707071 |
| 1.728 | 0.0757576 |
| 1.682 | 0.0808081 |
| 1.597 | 0.0858586 |
| 1.582 | 0.0909091 |
| 1.559 | 0.0959596 |
| 1.552 | 0.10101 |
| 1.519 | 0.106061 |
| 1.514 | 0.111111 |
| 1.505 | 0.116162 |
| 1.496 | 0.121212 |
| 1.423 | 0.126263 |
| 1.397 | 0.131313 |
| 1.34 | 0.136364 |
| 1.336 | 0.141414 |
| 1.268 | 0.146465 |
| 1.236 | 0.151515 |
| 1.207 | 0.156566 |
| 1.204 | 0.161616 |
| 1.202 | 0.166667 |
| 1.157 | 0.171717 |
| 1.103 | 0.176768 |
| 1.047 | 0.181818 |
| 1.038 | 0.186869 |
| 1.036 | 0.191919 |
| 1.023 | 0.19697 |
| 1.007 | 0.20202 |
| 0.939 | 0.207071 |
| 0.924 | 0.212121 |
| 0.917 | 0.217172 |
| 0.914 | 0.222222 |
| 0.887 | 0.227273 |
| 0.883 | 0.232323 |
| 0.868 | 0.237374 |
| 0.858 | 0.242424 |
| 0.855 | 0.247475 |
| 0.852 | 0.252525 |
| 0.841 | 0.257576 |
| 0.833 | 0.262626 |
| 0.805 | 0.267677 |
| 0.789 | 0.272727 |
| 0.779 | 0.277778 |
| 0.776 | 0.282828 |
| 0.77 | 0.287879 |
| 0.755 | 0.292929 |
| 0.746 | 0.29798 |
| 0.734 | 0.30303 |
| 0.715 | 0.308081 |
| 0.685 | 0.313131 |
| 0.684 | 0.318182 |
| 0.683 | 0.323232 |
| 0.679 | 0.328283 |
| 0.672 | 0.333333 |
| 0.672 | 0.338384 |
| 0.667 | 0.343434 |
| 0.658 | 0.348485 |
| 0.648 | 0.353535 |
| 0.645 | 0.358586 |
| 0.639 | 0.363636 |
| 0.625 | 0.368687 |
| 0.624 | 0.373737 |
| 0.609 | 0.378788 |
| 0.606 | 0.383838 |
| 0.604 | 0.388889 |
| 0.591 | 0.393939 |
| 0.585 | 0.39899 |
| 0.583 | 0.40404 |
| 0.581 | 0.409091 |
| 0.559 | 0.414141 |
| 0.557 | 0.419192 |
| 0.553 | 0.424242 |
| 0.551 | 0.429293 |
| 0.544 | 0.434343 |
| 0.54 | 0.439394 |
| 0.538 | 0.444444 |
| 0.534 | 0.449495 |
| 0.533 | 0.454545 |
| 0.52 | 0.459596 |
| 0.518 | 0.464646 |
| 0.518 | 0.469697 |
| 0.517 | 0.474747 |
| 0.517 | 0.479798 |
| 0.516 | 0.484848 |
| 0.511 | 0.489899 |
| 0.507 | 0.494949 |
| 0.499 | 0.5 |
| 0.495 | 0.505051 |
| 0.488 | 0.510101 |
| 0.485 | 0.515152 |
| 0.477 | 0.520202 |
| 0.477 | 0.525253 |
| 0.473 | 0.530303 |
| 0.472 | 0.535354 |
| 0.471 | 0.540404 |
| 0.469 | 0.545455 |
| 0.468 | 0.550505 |
| 0.467 | 0.555556 |
| 0.459 | 0.560606 |
| 0.457 | 0.565657 |
| 0.456 | 0.570707 |
| 0.428 | 0.575758 |
| 0.427 | 0.580808 |
| 0.424 | 0.585859 |
| 0.416 | 0.590909 |
| 0.404 | 0.59596 |
| 0.402 | 0.60101 |
| 0.401 | 0.606061 |
| 0.4 | 0.611111 |
| 0.4 | 0.616162 |
| 0.396 | 0.621212 |
| 0.395 | 0.626263 |
| 0.389 | 0.631313 |
| 0.382 | 0.636364 |
| 0.379 | 0.641414 |
| 0.378 | 0.646465 |
| 0.376 | 0.651515 |
| 0.373 | 0.656566 |
| 0.371 | 0.661616 |
| 0.371 | 0.666667 |
| 0.369 | 0.671717 |
| 0.368 | 0.676768 |
| 0.365 | 0.681818 |
| 0.364 | 0.686869 |
| 0.363 | 0.691919 |
| 0.357 | 0.69697 |
| 0.355 | 0.70202 |
| 0.351 | 0.707071 |
| 0.351 | 0.712121 |
| 0.346 | 0.717172 |
| 0.346 | 0.722222 |
| 0.346 | 0.727273 |
| 0.345 | 0.732323 |
| 0.343 | 0.737374 |
| 0.342 | 0.742424 |
| 0.341 | 0.747475 |
| 0.338 | 0.752525 |
| 0.337 | 0.757576 |
| 0.335 | 0.762626 |
| 0.328 | 0.767677 |
| 0.326 | 0.772727 |
| 0.326 | 0.777778 |
| 0.325 | 0.782828 |
| 0.32 | 0.787879 |
| 0.314 | 0.792929 |
| 0.313 | 0.79798 |
| 0.307 | 0.80303 |
| 0.306 | 0.808081 |
| 0.301 | 0.813131 |
| 0.296 | 0.818182 |
| 0.294 | 0.823232 |
| 0.272 | 0.828283 |
| 0.27 | 0.833333 |
| 0.27 | 0.838384 |
| 0.27 | 0.843434 |
| 0.267 | 0.848485 |
| 0.265 | 0.853535 |
| 0.261 | 0.858586 |
| 0.256 | 0.863636 |
| 0.251 | 0.868687 |
| 0.244 | 0.873737 |
| 0.242 | 0.878788 |
| 0.242 | 0.883838 |
| 0.237 | 0.888889 |
| 0.225 | 0.893939 |
| 0.221 | 0.89899 |
| 0.22 | 0.90404 |
| 0.21 | 0.909091 |
| 0.209 | 0.914141 |
| 0.205 | 0.919192 |
| 0.198 | 0.924242 |
| 0.196 | 0.929293 |
| 0.185 | 0.934343 |
| 0.181 | 0.939394 |
| 0.177 | 0.944444 |
| 0.173 | 0.949495 |
| 0.167 | 0.954545 |
| 0.165 | 0.959596 |
| 0.164 | 0.964646 |
| 0.158 | 0.969697 |
| 0.141 | 0.974747 |
| 0.137 | 0.979798 |
| 0.129 | 0.984848 |
| 0.116 | 0.989899 |
| 0.076 | 0.994949 |
| 0.068 | 1 |

|  | ST3-BE-PCV14 (Day 300) |
| --- | --- |
| 7.136 | 0.00531915 |
| 4.165 | 0.0106383 |
| 3.935 | 0.0159574 |
| 3.861 | 0.0212766 |
| 3.718 | 0.0265957 |
| 3.53 | 0.0319149 |
| 3.388 | 0.037234 |
| 3.289 | 0.0425532 |
| 3.21 | 0.0478723 |
| 3.175 | 0.0531915 |
| 3.097 | 0.0585106 |
| 2.928 | 0.0638298 |
| 2.807 | 0.0691489 |
| 2.661 | 0.0744681 |
| 2.584 | 0.0797872 |
| 2.577 | 0.0851064 |
| 2.504 | 0.0904255 |
| 2.442 | 0.0957447 |
| 2.342 | 0.101064 |
| 2.284 | 0.106383 |
| 2.267 | 0.111702 |
| 2.219 | 0.117021 |
| 1.938 | 0.12234 |
| 1.838 | 0.12766 |
| 1.815 | 0.132979 |
| 1.753 | 0.138298 |
| 1.725 | 0.143617 |
| 1.708 | 0.148936 |
| 1.698 | 0.154255 |
| 1.689 | 0.159574 |
| 1.686 | 0.164894 |
| 1.679 | 0.170213 |
| 1.664 | 0.175532 |
| 1.657 | 0.180851 |
| 1.647 | 0.18617 |
| 1.608 | 0.191489 |
| 1.562 | 0.196809 |
| 1.561 | 0.202128 |
| 1.552 | 0.207447 |
| 1.55 | 0.212766 |
| 1.536 | 0.218085 |
| 1.507 | 0.223404 |
| 1.494 | 0.228723 |
| 1.484 | 0.234043 |
| 1.46 | 0.239362 |
| 1.437 | 0.244681 |
| 1.376 | 0.25 |
| 1.37 | 0.255319 |
| 1.36 | 0.260638 |
| 1.356 | 0.265957 |
| 1.345 | 0.271277 |
| 1.331 | 0.276596 |
| 1.321 | 0.281915 |
| 1.319 | 0.287234 |
| 1.317 | 0.292553 |
| 1.309 | 0.297872 |
| 1.306 | 0.303191 |
| 1.289 | 0.308511 |
| 1.283 | 0.31383 |
| 1.276 | 0.319149 |
| 1.276 | 0.324468 |
| 1.273 | 0.329787 |
| 1.271 | 0.335106 |
| 1.264 | 0.340426 |
| 1.257 | 0.345745 |
| 1.243 | 0.351064 |
| 1.237 | 0.356383 |
| 1.218 | 0.361702 |
| 1.216 | 0.367021 |
| 1.196 | 0.37234 |
| 1.182 | 0.37766 |
| 1.177 | 0.382979 |
| 1.171 | 0.388298 |
| 1.156 | 0.393617 |
| 1.15 | 0.398936 |
| 1.138 | 0.404255 |
| 1.138 | 0.409574 |
| 1.134 | 0.414894 |
| 1.134 | 0.420213 |
| 1.133 | 0.425532 |
| 1.129 | 0.430851 |
| 1.125 | 0.43617 |
| 1.121 | 0.441489 |
| 1.113 | 0.446809 |
| 1.106 | 0.452128 |
| 1.079 | 0.457447 |
| 1.074 | 0.462766 |
| 1.045 | 0.468085 |
| 1.037 | 0.473404 |
| 1.029 | 0.478723 |
| 1.028 | 0.484043 |
| 1.008 | 0.489362 |
| 1.003 | 0.494681 |
| 0.995 | 0.5 |
| 0.986 | 0.505319 |
| 0.979 | 0.510638 |
| 0.962 | 0.515957 |
| 0.959 | 0.521277 |
| 0.952 | 0.526596 |
| 0.935 | 0.531915 |
| 0.923 | 0.537234 |
| 0.922 | 0.542553 |
| 0.921 | 0.547872 |
| 0.896 | 0.553191 |
| 0.895 | 0.558511 |
| 0.888 | 0.56383 |
| 0.881 | 0.569149 |
| 0.853 | 0.574468 |
| 0.842 | 0.579787 |
| 0.841 | 0.585106 |
| 0.82 | 0.590426 |
| 0.802 | 0.595745 |
| 0.793 | 0.601064 |
| 0.791 | 0.606383 |
| 0.789 | 0.611702 |
| 0.777 | 0.617021 |
| 0.776 | 0.62234 |
| 0.753 | 0.62766 |
| 0.749 | 0.632979 |
| 0.727 | 0.638298 |
| 0.711 | 0.643617 |
| 0.697 | 0.648936 |
| 0.694 | 0.654255 |
| 0.691 | 0.659574 |
| 0.673 | 0.664894 |
| 0.654 | 0.670213 |
| 0.641 | 0.675532 |
| 0.638 | 0.680851 |
| 0.631 | 0.68617 |
| 0.626 | 0.691489 |
| 0.618 | 0.696809 |
| 0.606 | 0.702128 |
| 0.603 | 0.707447 |
| 0.599 | 0.712766 |
| 0.587 | 0.718085 |
| 0.577 | 0.723404 |
| 0.574 | 0.728723 |
| 0.567 | 0.734043 |
| 0.566 | 0.739362 |
| 0.558 | 0.744681 |
| 0.545 | 0.75 |
| 0.544 | 0.755319 |
| 0.543 | 0.760638 |
| 0.532 | 0.765957 |
| 0.532 | 0.771277 |
| 0.53 | 0.776596 |
| 0.524 | 0.781915 |
| 0.518 | 0.787234 |
| 0.512 | 0.792553 |
| 0.512 | 0.797872 |
| 0.497 | 0.803191 |
| 0.484 | 0.808511 |
| 0.461 | 0.81383 |
| 0.458 | 0.819149 |
| 0.451 | 0.824468 |
| 0.448 | 0.829787 |
| 0.43 | 0.835106 |
| 0.43 | 0.840426 |
| 0.422 | 0.845745 |
| 0.416 | 0.851064 |
| 0.407 | 0.856383 |
| 0.403 | 0.861702 |
| 0.4 | 0.867021 |
| 0.396 | 0.87234 |
| 0.378 | 0.87766 |
| 0.343 | 0.882979 |
| 0.308 | 0.888298 |
| 0.295 | 0.893617 |
| 0.286 | 0.898936 |
| 0.275 | 0.904255 |
| 0.274 | 0.909574 |
| 0.264 | 0.914894 |
| 0.251 | 0.920213 |
| 0.247 | 0.925532 |
| 0.227 | 0.930851 |
| 0.227 | 0.93617 |
| 0.224 | 0.941489 |
| 0.22 | 0.946809 |
| 0.198 | 0.952128 |
| 0.184 | 0.957447 |
| 0.182 | 0.962766 |
| 0.181 | 0.968085 |
| 0.177 | 0.973404 |
| 0.152 | 0.978723 |
| 0.121 | 0.984043 |
| 0.117 | 0.989362 |
| 0.099 | 0.994681 |
| 0.048 | 1 |

|  | ST3-Prevenar 13 (Day 300) |
| --- | --- |
| 8.159 | 0.00526316 |
| 7.903 | 0.0105263 |
| 3.577 | 0.0157895 |
| 3.507 | 0.0210526 |
| 3.391 | 0.0263158 |
| 3.215 | 0.0315789 |
| 2.952 | 0.0368421 |
| 2.677 | 0.0421053 |
| 2.649 | 0.0473684 |
| 2.371 | 0.0526316 |
| 2.303 | 0.0578947 |
| 2.279 | 0.0631579 |
| 2.216 | 0.0684211 |
| 2.144 | 0.0736842 |
| 2.135 | 0.0789474 |
| 2.078 | 0.0842105 |
| 2.073 | 0.0894737 |
| 2.064 | 0.0947368 |
| 2.062 | 0.1 |
| 2.01 | 0.105263 |
| 1.997 | 0.110526 |
| 1.953 | 0.115789 |
| 1.938 | 0.121053 |
| 1.917 | 0.126316 |
| 1.859 | 0.131579 |
| 1.855 | 0.136842 |
| 1.819 | 0.142105 |
| 1.796 | 0.147368 |
| 1.768 | 0.152632 |
| 1.764 | 0.157895 |
| 1.745 | 0.163158 |
| 1.718 | 0.168421 |
| 1.608 | 0.173684 |
| 1.504 | 0.178947 |
| 1.481 | 0.184211 |
| 1.439 | 0.189474 |
| 1.371 | 0.194737 |
| 1.338 | 0.2 |
| 1.335 | 0.205263 |
| 1.328 | 0.210526 |
| 1.276 | 0.215789 |
| 1.261 | 0.221053 |
| 1.258 | 0.226316 |
| 1.243 | 0.231579 |
| 1.236 | 0.236842 |
| 1.198 | 0.242105 |
| 1.175 | 0.247368 |
| 1.169 | 0.252632 |
| 1.168 | 0.257895 |
| 1.131 | 0.263158 |
| 1.131 | 0.268421 |
| 1.113 | 0.273684 |
| 1.109 | 0.278947 |
| 1.105 | 0.284211 |
| 1.078 | 0.289474 |
| 1.041 | 0.294737 |
| 1.036 | 0.3 |
| 0.999 | 0.305263 |
| 0.989 | 0.310526 |
| 0.968 | 0.315789 |
| 0.965 | 0.321053 |
| 0.959 | 0.326316 |
| 0.957 | 0.331579 |
| 0.956 | 0.336842 |
| 0.949 | 0.342105 |
| 0.938 | 0.347368 |
| 0.929 | 0.352632 |
| 0.91 | 0.357895 |
| 0.907 | 0.363158 |
| 0.907 | 0.368421 |
| 0.905 | 0.373684 |
| 0.903 | 0.378947 |
| 0.882 | 0.384211 |
| 0.865 | 0.389474 |
| 0.859 | 0.394737 |
| 0.857 | 0.4 |
| 0.857 | 0.405263 |
| 0.843 | 0.410526 |
| 0.841 | 0.415789 |
| 0.829 | 0.421053 |
| 0.82 | 0.426316 |
| 0.802 | 0.431579 |
| 0.795 | 0.436842 |
| 0.79 | 0.442105 |
| 0.781 | 0.447368 |
| 0.778 | 0.452632 |
| 0.773 | 0.457895 |
| 0.771 | 0.463158 |
| 0.766 | 0.468421 |
| 0.762 | 0.473684 |
| 0.762 | 0.478947 |
| 0.751 | 0.484211 |
| 0.744 | 0.489474 |
| 0.742 | 0.494737 |
| 0.727 | 0.5 |
| 0.707 | 0.505263 |
| 0.701 | 0.510526 |
| 0.685 | 0.515789 |
| 0.682 | 0.521053 |
| 0.679 | 0.526316 |
| 0.675 | 0.531579 |
| 0.666 | 0.536842 |
| 0.665 | 0.542105 |
| 0.665 | 0.547368 |
| 0.663 | 0.552632 |
| 0.663 | 0.557895 |
| 0.658 | 0.563158 |
| 0.641 | 0.568421 |
| 0.633 | 0.573684 |
| 0.632 | 0.578947 |
| 0.63 | 0.584211 |
| 0.628 | 0.589474 |
| 0.614 | 0.594737 |
| 0.598 | 0.6 |
| 0.597 | 0.605263 |
| 0.585 | 0.610526 |
| 0.584 | 0.615789 |
| 0.582 | 0.621053 |
| 0.572 | 0.626316 |
| 0.569 | 0.631579 |
| 0.567 | 0.636842 |
| 0.563 | 0.642105 |
| 0.56 | 0.647368 |
| 0.552 | 0.652632 |
| 0.543 | 0.657895 |
| 0.542 | 0.663158 |
| 0.538 | 0.668421 |
| 0.532 | 0.673684 |
| 0.531 | 0.678947 |
| 0.525 | 0.684211 |
| 0.523 | 0.689474 |
| 0.515 | 0.694737 |
| 0.511 | 0.7 |
| 0.504 | 0.705263 |
| 0.502 | 0.710526 |
| 0.495 | 0.715789 |
| 0.493 | 0.721053 |
| 0.492 | 0.726316 |
| 0.486 | 0.731579 |
| 0.484 | 0.736842 |
| 0.48 | 0.742105 |
| 0.479 | 0.747368 |
| 0.474 | 0.752632 |
| 0.463 | 0.757895 |
| 0.46 | 0.763158 |
| 0.458 | 0.768421 |
| 0.453 | 0.773684 |
| 0.445 | 0.778947 |
| 0.44 | 0.784211 |
| 0.436 | 0.789474 |
| 0.414 | 0.794737 |
| 0.408 | 0.8 |
| 0.408 | 0.805263 |
| 0.399 | 0.810526 |
| 0.399 | 0.815789 |
| 0.396 | 0.821053 |
| 0.383 | 0.826316 |
| 0.38 | 0.831579 |
| 0.373 | 0.836842 |
| 0.371 | 0.842105 |
| 0.369 | 0.847368 |
| 0.361 | 0.852632 |
| 0.348 | 0.857895 |
| 0.343 | 0.863158 |
| 0.325 | 0.868421 |
| 0.324 | 0.873684 |
| 0.322 | 0.878947 |
| 0.32 | 0.884211 |
| 0.308 | 0.889474 |
| 0.306 | 0.894737 |
| 0.306 | 0.9 |
| 0.3 | 0.905263 |
| 0.299 | 0.910526 |
| 0.292 | 0.915789 |
| 0.288 | 0.921053 |
| 0.274 | 0.926316 |
| 0.266 | 0.931579 |
| 0.257 | 0.936842 |
| 0.256 | 0.942105 |
| 0.215 | 0.947368 |
| 0.176 | 0.952632 |
| 0.174 | 0.957895 |
| 0.163 | 0.963158 |
| 0.139 | 0.968421 |
| 0.139 | 0.973684 |
| 0.138 | 0.978947 |
| 0.13 | 0.984211 |
| 0.108 | 0.989474 |
| 0.071 | 0.994737 |
| 0.035 | 1 |

BECT056 - RCDC_Serotype 4

|  | ST4-BE-PCV14 (Day 84) |
| --- | --- |
| 22.383 | 0.0052356 |
| 20.305 | 0.0104712 |
| 20.271 | 0.0157068 |
| 18.215 | 0.0209424 |
| 17.898 | 0.026178 |
| 16.689 | 0.0314136 |
| 15.534 | 0.0366492 |
| 14.843 | 0.0418848 |
| 14.342 | 0.0471204 |
| 13.691 | 0.052356 |
| 13.624 | 0.0575916 |
| 11.318 | 0.0628272 |
| 10.805 | 0.0680628 |
| 8.506 | 0.0732984 |
| 8.425 | 0.078534 |
| 8.18 | 0.0837696 |
| 7.631 | 0.0890052 |
| 6.958 | 0.0942408 |
| 6.632 | 0.0994764 |
| 6.554 | 0.104712 |
| 6.537 | 0.109948 |
| 5.988 | 0.115183 |
| 5.969 | 0.120419 |
| 5.926 | 0.125654 |
| 5.871 | 0.13089 |
| 5.411 | 0.136126 |
| 5.223 | 0.141361 |
| 5.057 | 0.146597 |
| 5.053 | 0.151832 |
| 4.956 | 0.157068 |
| 4.885 | 0.162304 |
| 4.764 | 0.167539 |
| 4.746 | 0.172775 |
| 4.744 | 0.17801 |
| 4.672 | 0.183246 |
| 4.535 | 0.188482 |
| 4.392 | 0.193717 |
| 4.32 | 0.198953 |
| 4.31 | 0.204188 |
| 4.111 | 0.209424 |
| 4.103 | 0.21466 |
| 4.102 | 0.219895 |
| 4.099 | 0.225131 |
| 4.091 | 0.230366 |
| 4.045 | 0.235602 |
| 3.916 | 0.240838 |
| 3.838 | 0.246073 |
| 3.775 | 0.251309 |
| 3.769 | 0.256545 |
| 3.728 | 0.26178 |
| 3.701 | 0.267016 |
| 3.649 | 0.272251 |
| 3.567 | 0.277487 |
| 3.535 | 0.282723 |
| 3.366 | 0.287958 |
| 3.276 | 0.293194 |
| 3.258 | 0.298429 |
| 3.25 | 0.303665 |
| 3.246 | 0.308901 |
| 3.23 | 0.314136 |
| 3.155 | 0.319372 |
| 3.104 | 0.324607 |
| 2.924 | 0.329843 |
| 2.89 | 0.335079 |
| 2.863 | 0.340314 |
| 2.859 | 0.34555 |
| 2.855 | 0.350785 |
| 2.843 | 0.356021 |
| 2.821 | 0.361257 |
| 2.795 | 0.366492 |
| 2.742 | 0.371728 |
| 2.696 | 0.376963 |
| 2.694 | 0.382199 |
| 2.654 | 0.387435 |
| 2.652 | 0.39267 |
| 2.567 | 0.397906 |
| 2.541 | 0.403141 |
| 2.515 | 0.408377 |
| 2.382 | 0.413613 |
| 2.299 | 0.418848 |
| 2.233 | 0.424084 |
| 2.228 | 0.429319 |
| 2.214 | 0.434555 |
| 2.214 | 0.439791 |
| 2.214 | 0.445026 |
| 2.161 | 0.450262 |
| 2.142 | 0.455497 |
| 2.135 | 0.460733 |
| 2.103 | 0.465969 |
| 2.092 | 0.471204 |
| 2.06 | 0.47644 |
| 2.048 | 0.481675 |
| 1.999 | 0.486911 |
| 1.967 | 0.492147 |
| 1.93 | 0.497382 |
| 1.894 | 0.502618 |
| 1.832 | 0.507853 |
| 1.822 | 0.513089 |
| 1.815 | 0.518325 |
| 1.719 | 0.52356 |
| 1.645 | 0.528796 |
| 1.641 | 0.534031 |
| 1.616 | 0.539267 |
| 1.608 | 0.544503 |
| 1.602 | 0.549738 |
| 1.572 | 0.554974 |
| 1.571 | 0.560209 |
| 1.492 | 0.565445 |
| 1.47 | 0.570681 |
| 1.449 | 0.575916 |
| 1.394 | 0.581152 |
| 1.362 | 0.586387 |
| 1.355 | 0.591623 |
| 1.353 | 0.596859 |
| 1.346 | 0.602094 |
| 1.314 | 0.60733 |
| 1.311 | 0.612565 |
| 1.3 | 0.617801 |
| 1.274 | 0.623037 |
| 1.216 | 0.628272 |
| 1.21 | 0.633508 |
| 1.171 | 0.638743 |
| 1.162 | 0.643979 |
| 1.13 | 0.649215 |
| 1.066 | 0.65445 |
| 1.061 | 0.659686 |
| 1.055 | 0.664921 |
| 1.046 | 0.670157 |
| 1.03 | 0.675393 |
| 0.997 | 0.680628 |
| 0.98 | 0.685864 |
| 0.962 | 0.691099 |
| 0.949 | 0.696335 |
| 0.942 | 0.701571 |
| 0.941 | 0.706806 |
| 0.921 | 0.712042 |
| 0.902 | 0.717277 |
| 0.899 | 0.722513 |
| 0.844 | 0.727749 |
| 0.825 | 0.732984 |
| 0.823 | 0.73822 |
| 0.823 | 0.743455 |
| 0.818 | 0.748691 |
| 0.817 | 0.753927 |
| 0.803 | 0.759162 |
| 0.792 | 0.764398 |
| 0.777 | 0.769634 |
| 0.725 | 0.774869 |
| 0.71 | 0.780105 |
| 0.704 | 0.78534 |
| 0.704 | 0.790576 |
| 0.696 | 0.795812 |
| 0.695 | 0.801047 |
| 0.657 | 0.806283 |
| 0.645 | 0.811518 |
| 0.601 | 0.816754 |
| 0.585 | 0.82199 |
| 0.584 | 0.827225 |
| 0.581 | 0.832461 |
| 0.579 | 0.837696 |
| 0.577 | 0.842932 |
| 0.555 | 0.848168 |
| 0.552 | 0.853403 |
| 0.525 | 0.858639 |
| 0.503 | 0.863874 |
| 0.491 | 0.86911 |
| 0.489 | 0.874346 |
| 0.481 | 0.879581 |
| 0.481 | 0.884817 |
| 0.462 | 0.890052 |
| 0.458 | 0.895288 |
| 0.438 | 0.900524 |
| 0.427 | 0.905759 |
| 0.41 | 0.910995 |
| 0.402 | 0.91623 |
| 0.38 | 0.921466 |
| 0.376 | 0.926702 |
| 0.374 | 0.931937 |
| 0.37 | 0.937173 |
| 0.355 | 0.942408 |
| 0.301 | 0.947644 |
| 0.293 | 0.95288 |
| 0.292 | 0.958115 |
| 0.247 | 0.963351 |
| 0.236 | 0.968586 |
| 0.216 | 0.973822 |
| 0.159 | 0.979058 |
| 0.152 | 0.984293 |
| 0.151 | 0.989529 |
| 0.137 | 0.994764 |
| 0.125 | 1 |

|  | ST4-Prevenar 13 (Day 84) |
| --- | --- |
| 28.468 | 0.00505051 |
| 25 | 0.010101 |
| 15.326 | 0.0151515 |
| 15.151 | 0.020202 |
| 14.917 | 0.0252525 |
| 14.382 | 0.030303 |
| 14.056 | 0.0353535 |
| 12.772 | 0.040404 |
| 12.394 | 0.0454545 |
| 11.264 | 0.0505051 |
| 10.982 | 0.0555556 |
| 9.702 | 0.0606061 |
| 9.646 | 0.0656566 |
| 9.608 | 0.0707071 |
| 9.399 | 0.0757576 |
| 9.394 | 0.0808081 |
| 9.251 | 0.0858586 |
| 8.883 | 0.0909091 |
| 8.696 | 0.0959596 |
| 8.607 | 0.10101 |
| 7.904 | 0.106061 |
| 7.812 | 0.111111 |
| 7.81 | 0.116162 |
| 7.805 | 0.121212 |
| 7.728 | 0.126263 |
| 7.304 | 0.131313 |
| 7.161 | 0.136364 |
| 7.16 | 0.141414 |
| 6.695 | 0.146465 |
| 6.276 | 0.151515 |
| 6.151 | 0.156566 |
| 6.081 | 0.161616 |
| 5.928 | 0.166667 |
| 5.783 | 0.171717 |
| 5.753 | 0.176768 |
| 5.736 | 0.181818 |
| 5.265 | 0.186869 |
| 5.235 | 0.191919 |
| 5.15 | 0.19697 |
| 4.891 | 0.20202 |
| 4.866 | 0.207071 |
| 4.685 | 0.212121 |
| 4.458 | 0.217172 |
| 4.424 | 0.222222 |
| 4.393 | 0.227273 |
| 4.269 | 0.232323 |
| 4.217 | 0.237374 |
| 4.196 | 0.242424 |
| 4.148 | 0.247475 |
| 4.146 | 0.252525 |
| 4.126 | 0.257576 |
| 4.11 | 0.262626 |
| 4.039 | 0.267677 |
| 4.01 | 0.272727 |
| 3.875 | 0.277778 |
| 3.853 | 0.282828 |
| 3.833 | 0.287879 |
| 3.832 | 0.292929 |
| 3.758 | 0.29798 |
| 3.735 | 0.30303 |
| 3.709 | 0.308081 |
| 3.683 | 0.313131 |
| 3.629 | 0.318182 |
| 3.613 | 0.323232 |
| 3.588 | 0.328283 |
| 3.576 | 0.333333 |
| 3.566 | 0.338384 |
| 3.556 | 0.343434 |
| 3.508 | 0.348485 |
| 3.489 | 0.353535 |
| 3.482 | 0.358586 |
| 3.477 | 0.363636 |
| 3.444 | 0.368687 |
| 3.409 | 0.373737 |
| 3.378 | 0.378788 |
| 3.277 | 0.383838 |
| 3.255 | 0.388889 |
| 3.174 | 0.393939 |
| 3.086 | 0.39899 |
| 3.068 | 0.40404 |
| 3.058 | 0.409091 |
| 3.052 | 0.414141 |
| 3.049 | 0.419192 |
| 2.976 | 0.424242 |
| 2.904 | 0.429293 |
| 2.891 | 0.434343 |
| 2.845 | 0.439394 |
| 2.782 | 0.444444 |
| 2.755 | 0.449495 |
| 2.657 | 0.454545 |
| 2.627 | 0.459596 |
| 2.614 | 0.464646 |
| 2.587 | 0.469697 |
| 2.56 | 0.474747 |
| 2.501 | 0.479798 |
| 2.451 | 0.484848 |
| 2.429 | 0.489899 |
| 2.422 | 0.494949 |
| 2.405 | 0.5 |
| 2.364 | 0.505051 |
| 2.355 | 0.510101 |
| 2.347 | 0.515152 |
| 2.336 | 0.520202 |
| 2.336 | 0.525253 |
| 2.327 | 0.530303 |
| 2.326 | 0.535354 |
| 2.242 | 0.540404 |
| 2.224 | 0.545455 |
| 2.189 | 0.550505 |
| 2.184 | 0.555556 |
| 2.181 | 0.560606 |
| 2.139 | 0.565657 |
| 2.138 | 0.570707 |
| 2.128 | 0.575758 |
| 2.123 | 0.580808 |
| 2.108 | 0.585859 |
| 2.064 | 0.590909 |
| 2.043 | 0.59596 |
| 2.041 | 0.60101 |
| 2.033 | 0.606061 |
| 2.021 | 0.611111 |
| 2.003 | 0.616162 |
| 1.984 | 0.621212 |
| 1.97 | 0.626263 |
| 1.862 | 0.631313 |
| 1.848 | 0.636364 |
| 1.787 | 0.641414 |
| 1.774 | 0.646465 |
| 1.768 | 0.651515 |
| 1.742 | 0.656566 |
| 1.74 | 0.661616 |
| 1.694 | 0.666667 |
| 1.69 | 0.671717 |
| 1.644 | 0.676768 |
| 1.642 | 0.681818 |
| 1.606 | 0.686869 |
| 1.6 | 0.691919 |
| 1.525 | 0.69697 |
| 1.513 | 0.70202 |
| 1.506 | 0.707071 |
| 1.452 | 0.712121 |
| 1.409 | 0.717172 |
| 1.396 | 0.722222 |
| 1.374 | 0.727273 |
| 1.367 | 0.732323 |
| 1.338 | 0.737374 |
| 1.334 | 0.742424 |
| 1.314 | 0.747475 |
| 1.263 | 0.752525 |
| 1.258 | 0.757576 |
| 1.228 | 0.762626 |
| 1.216 | 0.767677 |
| 1.207 | 0.772727 |
| 1.197 | 0.777778 |
| 1.154 | 0.782828 |
| 1.147 | 0.787879 |
| 1.133 | 0.792929 |
| 1.132 | 0.79798 |
| 1.129 | 0.80303 |
| 1.123 | 0.808081 |
| 1.122 | 0.813131 |
| 1.105 | 0.818182 |
| 1.097 | 0.823232 |
| 1.048 | 0.828283 |
| 0.993 | 0.833333 |
| 0.992 | 0.838384 |
| 0.898 | 0.843434 |
| 0.853 | 0.848485 |
| 0.85 | 0.853535 |
| 0.838 | 0.858586 |
| 0.826 | 0.863636 |
| 0.81 | 0.868687 |
| 0.807 | 0.873737 |
| 0.752 | 0.878788 |
| 0.745 | 0.883838 |
| 0.741 | 0.888889 |
| 0.713 | 0.893939 |
| 0.708 | 0.89899 |
| 0.706 | 0.90404 |
| 0.7 | 0.909091 |
| 0.679 | 0.914141 |
| 0.675 | 0.919192 |
| 0.657 | 0.924242 |
| 0.638 | 0.929293 |
| 0.635 | 0.934343 |
| 0.634 | 0.939394 |
| 0.633 | 0.944444 |
| 0.629 | 0.949495 |
| 0.593 | 0.954545 |
| 0.553 | 0.959596 |
| 0.544 | 0.964646 |
| 0.446 | 0.969697 |
| 0.436 | 0.974747 |
| 0.433 | 0.979798 |
| 0.424 | 0.984848 |
| 0.408 | 0.989899 |
| 0.322 | 0.994949 |
| 0.3 | 1 |

|  | ST4-BE-PCV14 (Day 300) |
| --- | --- |
| 21.985 | 0.00531915 |
| 18.568 | 0.0106383 |
| 12.741 | 0.0159574 |
| 11.363 | 0.0212766 |
| 11.347 | 0.0265957 |
| 10.14 | 0.0319149 |
| 8.816 | 0.037234 |
| 7.964 | 0.0425532 |
| 7.927 | 0.0478723 |
| 7.798 | 0.0531915 |
| 7.57 | 0.0585106 |
| 7.216 | 0.0638298 |
| 7.146 | 0.0691489 |
| 6.92 | 0.0744681 |
| 6.692 | 0.0797872 |
| 6.672 | 0.0851064 |
| 6.611 | 0.0904255 |
| 6.293 | 0.0957447 |
| 6.17 | 0.101064 |
| 6.158 | 0.106383 |
| 5.978 | 0.111702 |
| 5.882 | 0.117021 |
| 5.87 | 0.12234 |
| 5.85 | 0.12766 |
| 5.717 | 0.132979 |
| 5.367 | 0.138298 |
| 5.247 | 0.143617 |
| 5.133 | 0.148936 |
| 5.121 | 0.154255 |
| 4.973 | 0.159574 |
| 4.953 | 0.164894 |
| 4.898 | 0.170213 |
| 4.718 | 0.175532 |
| 4.566 | 0.180851 |
| 4.519 | 0.18617 |
| 4.515 | 0.191489 |
| 4.507 | 0.196809 |
| 4.361 | 0.202128 |
| 4.341 | 0.207447 |
| 4.24 | 0.212766 |
| 4.221 | 0.218085 |
| 4.197 | 0.223404 |
| 4.149 | 0.228723 |
| 4.053 | 0.234043 |
| 4.049 | 0.239362 |
| 4.02 | 0.244681 |
| 3.999 | 0.25 |
| 3.966 | 0.255319 |
| 3.916 | 0.260638 |
| 3.915 | 0.265957 |
| 3.879 | 0.271277 |
| 3.871 | 0.276596 |
| 3.87 | 0.281915 |
| 3.837 | 0.287234 |
| 3.705 | 0.292553 |
| 3.662 | 0.297872 |
| 3.639 | 0.303191 |
| 3.554 | 0.308511 |
| 3.536 | 0.31383 |
| 3.527 | 0.319149 |
| 3.512 | 0.324468 |
| 3.493 | 0.329787 |
| 3.394 | 0.335106 |
| 3.312 | 0.340426 |
| 3.264 | 0.345745 |
| 3.258 | 0.351064 |
| 3.08 | 0.356383 |
| 3.052 | 0.361702 |
| 3.044 | 0.367021 |
| 3.035 | 0.37234 |
| 3.034 | 0.37766 |
| 3.014 | 0.382979 |
| 2.938 | 0.388298 |
| 2.927 | 0.393617 |
| 2.925 | 0.398936 |
| 2.921 | 0.404255 |
| 2.904 | 0.409574 |
| 2.802 | 0.414894 |
| 2.776 | 0.420213 |
| 2.77 | 0.425532 |
| 2.761 | 0.430851 |
| 2.687 | 0.43617 |
| 2.681 | 0.441489 |
| 2.641 | 0.446809 |
| 2.634 | 0.452128 |
| 2.577 | 0.457447 |
| 2.536 | 0.462766 |
| 2.52 | 0.468085 |
| 2.506 | 0.473404 |
| 2.453 | 0.478723 |
| 2.417 | 0.484043 |
| 2.399 | 0.489362 |
| 2.273 | 0.494681 |
| 2.235 | 0.5 |
| 2.226 | 0.505319 |
| 2.217 | 0.510638 |
| 2.17 | 0.515957 |
| 2.144 | 0.521277 |
| 2.137 | 0.526596 |
| 2.11 | 0.531915 |
| 2.089 | 0.537234 |
| 2.079 | 0.542553 |
| 1.996 | 0.547872 |
| 1.97 | 0.553191 |
| 1.943 | 0.558511 |
| 1.935 | 0.56383 |
| 1.923 | 0.569149 |
| 1.895 | 0.574468 |
| 1.873 | 0.579787 |
| 1.869 | 0.585106 |
| 1.847 | 0.590426 |
| 1.821 | 0.595745 |
| 1.814 | 0.601064 |
| 1.801 | 0.606383 |
| 1.747 | 0.611702 |
| 1.703 | 0.617021 |
| 1.687 | 0.62234 |
| 1.677 | 0.62766 |
| 1.67 | 0.632979 |
| 1.639 | 0.638298 |
| 1.622 | 0.643617 |
| 1.616 | 0.648936 |
| 1.616 | 0.654255 |
| 1.533 | 0.659574 |
| 1.498 | 0.664894 |
| 1.469 | 0.670213 |
| 1.452 | 0.675532 |
| 1.448 | 0.680851 |
| 1.42 | 0.68617 |
| 1.412 | 0.691489 |
| 1.404 | 0.696809 |
| 1.389 | 0.702128 |
| 1.362 | 0.707447 |
| 1.348 | 0.712766 |
| 1.345 | 0.718085 |
| 1.33 | 0.723404 |
| 1.307 | 0.728723 |
| 1.249 | 0.734043 |
| 1.236 | 0.739362 |
| 1.236 | 0.744681 |
| 1.234 | 0.75 |
| 1.217 | 0.755319 |
| 1.189 | 0.760638 |
| 1.162 | 0.765957 |
| 1.108 | 0.771277 |
| 1.1 | 0.776596 |
| 1.078 | 0.781915 |
| 1.02 | 0.787234 |
| 1.019 | 0.792553 |
| 0.998 | 0.797872 |
| 0.988 | 0.803191 |
| 0.966 | 0.808511 |
| 0.955 | 0.81383 |
| 0.944 | 0.819149 |
| 0.915 | 0.824468 |
| 0.852 | 0.829787 |
| 0.791 | 0.835106 |
| 0.78 | 0.840426 |
| 0.765 | 0.845745 |
| 0.743 | 0.851064 |
| 0.741 | 0.856383 |
| 0.74 | 0.861702 |
| 0.722 | 0.867021 |
| 0.662 | 0.87234 |
| 0.432 | 0.87766 |
| 0.432 | 0.882979 |
| 0.426 | 0.888298 |
| 0.423 | 0.893617 |
| 0.421 | 0.898936 |
| 0.398 | 0.904255 |
| 0.393 | 0.909574 |
| 0.382 | 0.914894 |
| 0.38 | 0.920213 |
| 0.372 | 0.925532 |
| 0.346 | 0.930851 |
| 0.34 | 0.93617 |
| 0.326 | 0.941489 |
| 0.32 | 0.946809 |
| 0.318 | 0.952128 |
| 0.27 | 0.957447 |
| 0.253 | 0.962766 |
| 0.205 | 0.968085 |
| 0.197 | 0.973404 |
| 0.173 | 0.978723 |
| 0.167 | 0.984043 |
| 0.167 | 0.989362 |
| 0.155 | 0.994681 |
| 0.12 | 1 |

|  | ST4-Prevenar 13 (Day 300) |
| --- | --- |
| 28.881 | 0.00526316 |
| 27.485 | 0.0105263 |
| 20.115 | 0.0157895 |
| 15.798 | 0.0210526 |
| 14.286 | 0.0263158 |
| 13.753 | 0.0315789 |
| 13.594 | 0.0368421 |
| 13.492 | 0.0421053 |
| 11.991 | 0.0473684 |
| 11.94 | 0.0526316 |
| 11.715 | 0.0578947 |
| 11.567 | 0.0631579 |
| 11.506 | 0.0684211 |
| 11.15 | 0.0736842 |
| 10.725 | 0.0789474 |
| 10.577 | 0.0842105 |
| 10.471 | 0.0894737 |
| 10.333 | 0.0947368 |
| 10.314 | 0.1 |
| 9.2 | 0.105263 |
| 9.025 | 0.110526 |
| 8.46 | 0.115789 |
| 7.831 | 0.121053 |
| 7.67 | 0.126316 |
| 7.466 | 0.131579 |
| 7.273 | 0.136842 |
| 7.083 | 0.142105 |
| 7.044 | 0.147368 |
| 6.909 | 0.152632 |
| 6.521 | 0.157895 |
| 6.039 | 0.163158 |
| 6.028 | 0.168421 |
| 5.875 | 0.173684 |
| 5.753 | 0.178947 |
| 5.635 | 0.184211 |
| 5.6 | 0.189474 |
| 5.489 | 0.194737 |
| 5.342 | 0.2 |
| 5.061 | 0.205263 |
| 4.898 | 0.210526 |
| 4.856 | 0.215789 |
| 4.833 | 0.221053 |
| 4.817 | 0.226316 |
| 4.812 | 0.231579 |
| 4.792 | 0.236842 |
| 4.722 | 0.242105 |
| 4.508 | 0.247368 |
| 4.483 | 0.252632 |
| 4.456 | 0.257895 |
| 4.41 | 0.263158 |
| 4.361 | 0.268421 |
| 4.324 | 0.273684 |
| 4.322 | 0.278947 |
| 4.304 | 0.284211 |
| 4.214 | 0.289474 |
| 4.139 | 0.294737 |
| 4.114 | 0.3 |
| 4.095 | 0.305263 |
| 4.053 | 0.310526 |
| 4.008 | 0.315789 |
| 3.967 | 0.321053 |
| 3.849 | 0.326316 |
| 3.845 | 0.331579 |
| 3.738 | 0.336842 |
| 3.717 | 0.342105 |
| 3.693 | 0.347368 |
| 3.537 | 0.352632 |
| 3.466 | 0.357895 |
| 3.442 | 0.363158 |
| 3.333 | 0.368421 |
| 3.312 | 0.373684 |
| 3.279 | 0.378947 |
| 3.198 | 0.384211 |
| 3.166 | 0.389474 |
| 3.125 | 0.394737 |
| 3.104 | 0.4 |
| 3.102 | 0.405263 |
| 3.095 | 0.410526 |
| 3.033 | 0.415789 |
| 3.027 | 0.421053 |
| 2.948 | 0.426316 |
| 2.946 | 0.431579 |
| 2.94 | 0.436842 |
| 2.932 | 0.442105 |
| 2.893 | 0.447368 |
| 2.697 | 0.452632 |
| 2.662 | 0.457895 |
| 2.656 | 0.463158 |
| 2.645 | 0.468421 |
| 2.592 | 0.473684 |
| 2.572 | 0.478947 |
| 2.525 | 0.484211 |
| 2.518 | 0.489474 |
| 2.471 | 0.494737 |
| 2.387 | 0.5 |
| 2.377 | 0.505263 |
| 2.308 | 0.510526 |
| 2.237 | 0.515789 |
| 2.228 | 0.521053 |
| 2.202 | 0.526316 |
| 2.167 | 0.531579 |
| 2.163 | 0.536842 |
| 2.129 | 0.542105 |
| 2.094 | 0.547368 |
| 2.091 | 0.552632 |
| 2.076 | 0.557895 |
| 2.062 | 0.563158 |
| 2.04 | 0.568421 |
| 2.035 | 0.573684 |
| 1.998 | 0.578947 |
| 1.996 | 0.584211 |
| 1.973 | 0.589474 |
| 1.914 | 0.594737 |
| 1.902 | 0.6 |
| 1.888 | 0.605263 |
| 1.843 | 0.610526 |
| 1.838 | 0.615789 |
| 1.818 | 0.621053 |
| 1.802 | 0.626316 |
| 1.771 | 0.631579 |
| 1.749 | 0.636842 |
| 1.738 | 0.642105 |
| 1.716 | 0.647368 |
| 1.69 | 0.652632 |
| 1.67 | 0.657895 |
| 1.639 | 0.663158 |
| 1.631 | 0.668421 |
| 1.631 | 0.673684 |
| 1.574 | 0.678947 |
| 1.55 | 0.684211 |
| 1.549 | 0.689474 |
| 1.498 | 0.694737 |
| 1.477 | 0.7 |
| 1.429 | 0.705263 |
| 1.419 | 0.710526 |
| 1.418 | 0.715789 |
| 1.394 | 0.721053 |
| 1.369 | 0.726316 |
| 1.364 | 0.731579 |
| 1.325 | 0.736842 |
| 1.303 | 0.742105 |
| 1.192 | 0.747368 |
| 1.188 | 0.752632 |
| 1.16 | 0.757895 |
| 1.132 | 0.763158 |
| 1.089 | 0.768421 |
| 1.07 | 0.773684 |
| 1.063 | 0.778947 |
| 1.053 | 0.784211 |
| 1.052 | 0.789474 |
| 1.05 | 0.794737 |
| 1.028 | 0.8 |
| 1.011 | 0.805263 |
| 0.996 | 0.810526 |
| 0.867 | 0.815789 |
| 0.812 | 0.821053 |
| 0.795 | 0.826316 |
| 0.751 | 0.831579 |
| 0.733 | 0.836842 |
| 0.715 | 0.842105 |
| 0.707 | 0.847368 |
| 0.701 | 0.852632 |
| 0.7 | 0.857895 |
| 0.699 | 0.863158 |
| 0.693 | 0.868421 |
| 0.679 | 0.873684 |
| 0.67 | 0.878947 |
| 0.658 | 0.884211 |
| 0.655 | 0.889474 |
| 0.536 | 0.894737 |
| 0.529 | 0.9 |
| 0.499 | 0.905263 |
| 0.497 | 0.910526 |
| 0.477 | 0.915789 |
| 0.469 | 0.921053 |
| 0.45 | 0.926316 |
| 0.434 | 0.931579 |
| 0.432 | 0.936842 |
| 0.422 | 0.942105 |
| 0.409 | 0.947368 |
| 0.34 | 0.952632 |
| 0.338 | 0.957895 |
| 0.233 | 0.963158 |
| 0.211 | 0.968421 |
| 0.202 | 0.973684 |
| 0.188 | 0.978947 |
| 0.171 | 0.984211 |
| 0.163 | 0.989474 |
| 0.112 | 0.994737 |
| 0.048 | 1 |

BECT056 - RCDC_Serotype 5

|  | ST5-BE-PCV14 (Day 84) |
| --- | --- |
| 143.58 | 0.0052356 |
| 115.186 | 0.0104712 |
| 74.822 | 0.0157068 |
| 48.352 | 0.0209424 |
| 41.495 | 0.026178 |
| 41.422 | 0.0314136 |
| 40.481 | 0.0366492 |
| 39.863 | 0.0418848 |
| 39.831 | 0.0471204 |
| 39.674 | 0.052356 |
| 29.658 | 0.0575916 |
| 15.172 | 0.0628272 |
| 14.416 | 0.0680628 |
| 13.37 | 0.0732984 |
| 12.562 | 0.078534 |
| 12.026 | 0.0837696 |
| 11.324 | 0.0890052 |
| 11.282 | 0.0942408 |
| 10.441 | 0.0994764 |
| 9.242 | 0.104712 |
| 9.157 | 0.109948 |
| 8.72 | 0.115183 |
| 7.721 | 0.120419 |
| 6.054 | 0.125654 |
| 5.835 | 0.13089 |
| 5.808 | 0.136126 |
| 5.594 | 0.141361 |
| 5.227 | 0.146597 |
| 5.126 | 0.151832 |
| 4.715 | 0.157068 |
| 4.375 | 0.162304 |
| 4.094 | 0.167539 |
| 4.072 | 0.172775 |
| 3.957 | 0.17801 |
| 3.829 | 0.183246 |
| 3.786 | 0.188482 |
| 3.647 | 0.193717 |
| 3.533 | 0.198953 |
| 3.488 | 0.204188 |
| 3.253 | 0.209424 |
| 3.219 | 0.21466 |
| 3.074 | 0.219895 |
| 3.023 | 0.225131 |
| 2.924 | 0.230366 |
| 2.907 | 0.235602 |
| 2.875 | 0.240838 |
| 2.843 | 0.246073 |
| 2.826 | 0.251309 |
| 2.669 | 0.256545 |
| 2.645 | 0.26178 |
| 2.623 | 0.267016 |
| 2.616 | 0.272251 |
| 2.538 | 0.277487 |
| 2.445 | 0.282723 |
| 2.424 | 0.287958 |
| 2.422 | 0.293194 |
| 2.402 | 0.298429 |
| 2.374 | 0.303665 |
| 2.3 | 0.308901 |
| 2.288 | 0.314136 |
| 2.286 | 0.319372 |
| 2.285 | 0.324607 |
| 2.215 | 0.329843 |
| 2.211 | 0.335079 |
| 2.2 | 0.340314 |
| 2.185 | 0.34555 |
| 2.179 | 0.350785 |
| 2.123 | 0.356021 |
| 2.055 | 0.361257 |
| 2.055 | 0.366492 |
| 2.026 | 0.371728 |
| 2.011 | 0.376963 |
| 2.01 | 0.382199 |
| 2.002 | 0.387435 |
| 1.994 | 0.39267 |
| 1.993 | 0.397906 |
| 1.989 | 0.403141 |
| 1.969 | 0.408377 |
| 1.962 | 0.413613 |
| 1.943 | 0.418848 |
| 1.937 | 0.424084 |
| 1.888 | 0.429319 |
| 1.881 | 0.434555 |
| 1.88 | 0.439791 |
| 1.849 | 0.445026 |
| 1.832 | 0.450262 |
| 1.83 | 0.455497 |
| 1.817 | 0.460733 |
| 1.739 | 0.465969 |
| 1.738 | 0.471204 |
| 1.737 | 0.47644 |
| 1.704 | 0.481675 |
| 1.688 | 0.486911 |
| 1.681 | 0.492147 |
| 1.675 | 0.497382 |
| 1.652 | 0.502618 |
| 1.645 | 0.507853 |
| 1.638 | 0.513089 |
| 1.62 | 0.518325 |
| 1.601 | 0.52356 |
| 1.561 | 0.528796 |
| 1.56 | 0.534031 |
| 1.548 | 0.539267 |
| 1.535 | 0.544503 |
| 1.532 | 0.549738 |
| 1.53 | 0.554974 |
| 1.515 | 0.560209 |
| 1.513 | 0.565445 |
| 1.513 | 0.570681 |
| 1.451 | 0.575916 |
| 1.445 | 0.581152 |
| 1.438 | 0.586387 |
| 1.423 | 0.591623 |
| 1.422 | 0.596859 |
| 1.416 | 0.602094 |
| 1.38 | 0.60733 |
| 1.369 | 0.612565 |
| 1.36 | 0.617801 |
| 1.346 | 0.623037 |
| 1.332 | 0.628272 |
| 1.331 | 0.633508 |
| 1.329 | 0.638743 |
| 1.327 | 0.643979 |
| 1.31 | 0.649215 |
| 1.285 | 0.65445 |
| 1.284 | 0.659686 |
| 1.256 | 0.664921 |
| 1.215 | 0.670157 |
| 1.155 | 0.675393 |
| 1.146 | 0.680628 |
| 1.117 | 0.685864 |
| 1.107 | 0.691099 |
| 1.106 | 0.696335 |
| 1.09 | 0.701571 |
| 1.062 | 0.706806 |
| 1.037 | 0.712042 |
| 1.033 | 0.717277 |
| 1.032 | 0.722513 |
| 1.012 | 0.727749 |
| 1.011 | 0.732984 |
| 0.995 | 0.73822 |
| 0.978 | 0.743455 |
| 0.945 | 0.748691 |
| 0.915 | 0.753927 |
| 0.894 | 0.759162 |
| 0.887 | 0.764398 |
| 0.884 | 0.769634 |
| 0.846 | 0.774869 |
| 0.841 | 0.780105 |
| 0.837 | 0.78534 |
| 0.82 | 0.790576 |
| 0.82 | 0.795812 |
| 0.778 | 0.801047 |
| 0.769 | 0.806283 |
| 0.755 | 0.811518 |
| 0.747 | 0.816754 |
| 0.727 | 0.82199 |
| 0.725 | 0.827225 |
| 0.723 | 0.832461 |
| 0.712 | 0.837696 |
| 0.69 | 0.842932 |
| 0.674 | 0.848168 |
| 0.655 | 0.853403 |
| 0.648 | 0.858639 |
| 0.639 | 0.863874 |
| 0.637 | 0.86911 |
| 0.598 | 0.874346 |
| 0.594 | 0.879581 |
| 0.589 | 0.884817 |
| 0.574 | 0.890052 |
| 0.539 | 0.895288 |
| 0.528 | 0.900524 |
| 0.514 | 0.905759 |
| 0.51 | 0.910995 |
| 0.508 | 0.91623 |
| 0.456 | 0.921466 |
| 0.439 | 0.926702 |
| 0.392 | 0.931937 |
| 0.392 | 0.937173 |
| 0.384 | 0.942408 |
| 0.35 | 0.947644 |
| 0.348 | 0.95288 |
| 0.333 | 0.958115 |
| 0.31 | 0.963351 |
| 0.307 | 0.968586 |
| 0.274 | 0.973822 |
| 0.256 | 0.979058 |
| 0.244 | 0.984293 |
| 0.222 | 0.989529 |
| 0.201 | 0.994764 |
| 0.136 | 1 |

|  | ST5-Prevenar 13 (Day 84) |
| --- | --- |
| 204.785 | 0.00505051 |
| 165.407 | 0.010101 |
| 122.6 | 0.0151515 |
| 85.633 | 0.020202 |
| 67.528 | 0.0252525 |
| 53.903 | 0.030303 |
| 51.345 | 0.0353535 |
| 41.766 | 0.040404 |
| 31.489 | 0.0454545 |
| 27.377 | 0.0505051 |
| 26.477 | 0.0555556 |
| 25.573 | 0.0606061 |
| 22.199 | 0.0656566 |
| 18.512 | 0.0707071 |
| 17.523 | 0.0757576 |
| 14.603 | 0.0808081 |
| 14.224 | 0.0858586 |
| 13.864 | 0.0909091 |
| 12.568 | 0.0959596 |
| 12.567 | 0.10101 |
| 11.94 | 0.106061 |
| 11.026 | 0.111111 |
| 10.041 | 0.116162 |
| 8.559 | 0.121212 |
| 7.984 | 0.126263 |
| 7.799 | 0.131313 |
| 7.296 | 0.136364 |
| 7.211 | 0.141414 |
| 7.169 | 0.146465 |
| 7.017 | 0.151515 |
| 6.718 | 0.156566 |
| 6.679 | 0.161616 |
| 6.531 | 0.166667 |
| 5.768 | 0.171717 |
| 5.749 | 0.176768 |
| 4.722 | 0.181818 |
| 4.654 | 0.186869 |
| 4.525 | 0.191919 |
| 4.131 | 0.19697 |
| 4.075 | 0.20202 |
| 3.968 | 0.207071 |
| 3.917 | 0.212121 |
| 3.897 | 0.217172 |
| 3.716 | 0.222222 |
| 3.5 | 0.227273 |
| 3.472 | 0.232323 |
| 3.463 | 0.237374 |
| 3.427 | 0.242424 |
| 3.408 | 0.247475 |
| 3.295 | 0.252525 |
| 3.257 | 0.257576 |
| 3.185 | 0.262626 |
| 3.065 | 0.267677 |
| 3.007 | 0.272727 |
| 2.995 | 0.277778 |
| 2.933 | 0.282828 |
| 2.876 | 0.287879 |
| 2.825 | 0.292929 |
| 2.727 | 0.29798 |
| 2.715 | 0.30303 |
| 2.652 | 0.308081 |
| 2.59 | 0.313131 |
| 2.521 | 0.318182 |
| 2.515 | 0.323232 |
| 2.478 | 0.328283 |
| 2.448 | 0.333333 |
| 2.235 | 0.338384 |
| 2.226 | 0.343434 |
| 2.216 | 0.348485 |
| 2.213 | 0.353535 |
| 2.184 | 0.358586 |
| 2.154 | 0.363636 |
| 2.101 | 0.368687 |
| 2.019 | 0.373737 |
| 2.015 | 0.378788 |
| 1.924 | 0.383838 |
| 1.907 | 0.388889 |
| 1.891 | 0.393939 |
| 1.889 | 0.39899 |
| 1.873 | 0.40404 |
| 1.869 | 0.409091 |
| 1.859 | 0.414141 |
| 1.85 | 0.419192 |
| 1.816 | 0.424242 |
| 1.802 | 0.429293 |
| 1.738 | 0.434343 |
| 1.737 | 0.439394 |
| 1.719 | 0.444444 |
| 1.717 | 0.449495 |
| 1.712 | 0.454545 |
| 1.703 | 0.459596 |
| 1.666 | 0.464646 |
| 1.648 | 0.469697 |
| 1.646 | 0.474747 |
| 1.635 | 0.479798 |
| 1.621 | 0.484848 |
| 1.617 | 0.489899 |
| 1.606 | 0.494949 |
| 1.595 | 0.5 |
| 1.561 | 0.505051 |
| 1.554 | 0.510101 |
| 1.511 | 0.515152 |
| 1.508 | 0.520202 |
| 1.501 | 0.525253 |
| 1.465 | 0.530303 |
| 1.44 | 0.535354 |
| 1.418 | 0.540404 |
| 1.415 | 0.545455 |
| 1.412 | 0.550505 |
| 1.404 | 0.555556 |
| 1.383 | 0.560606 |
| 1.373 | 0.565657 |
| 1.364 | 0.570707 |
| 1.361 | 0.575758 |
| 1.347 | 0.580808 |
| 1.321 | 0.585859 |
| 1.312 | 0.590909 |
| 1.26 | 0.59596 |
| 1.26 | 0.60101 |
| 1.249 | 0.606061 |
| 1.208 | 0.611111 |
| 1.165 | 0.616162 |
| 1.109 | 0.621212 |
| 1.082 | 0.626263 |
| 1.053 | 0.631313 |
| 1.047 | 0.636364 |
| 1.041 | 0.641414 |
| 1.028 | 0.646465 |
| 1.014 | 0.651515 |
| 1.006 | 0.656566 |
| 0.995 | 0.661616 |
| 0.987 | 0.666667 |
| 0.983 | 0.671717 |
| 0.958 | 0.676768 |
| 0.957 | 0.681818 |
| 0.907 | 0.686869 |
| 0.891 | 0.691919 |
| 0.886 | 0.69697 |
| 0.87 | 0.70202 |
| 0.861 | 0.707071 |
| 0.842 | 0.712121 |
| 0.834 | 0.717172 |
| 0.817 | 0.722222 |
| 0.811 | 0.727273 |
| 0.799 | 0.732323 |
| 0.774 | 0.737374 |
| 0.765 | 0.742424 |
| 0.761 | 0.747475 |
| 0.761 | 0.752525 |
| 0.746 | 0.757576 |
| 0.727 | 0.762626 |
| 0.725 | 0.767677 |
| 0.725 | 0.772727 |
| 0.698 | 0.777778 |
| 0.696 | 0.782828 |
| 0.691 | 0.787879 |
| 0.682 | 0.792929 |
| 0.682 | 0.79798 |
| 0.659 | 0.80303 |
| 0.637 | 0.808081 |
| 0.634 | 0.813131 |
| 0.628 | 0.818182 |
| 0.615 | 0.823232 |
| 0.607 | 0.828283 |
| 0.603 | 0.833333 |
| 0.595 | 0.838384 |
| 0.594 | 0.843434 |
| 0.583 | 0.848485 |
| 0.556 | 0.853535 |
| 0.488 | 0.858586 |
| 0.483 | 0.863636 |
| 0.474 | 0.868687 |
| 0.471 | 0.873737 |
| 0.466 | 0.878788 |
| 0.42 | 0.883838 |
| 0.417 | 0.888889 |
| 0.414 | 0.893939 |
| 0.412 | 0.89899 |
| 0.409 | 0.90404 |
| 0.396 | 0.909091 |
| 0.388 | 0.914141 |
| 0.351 | 0.919192 |
| 0.349 | 0.924242 |
| 0.345 | 0.929293 |
| 0.319 | 0.934343 |
| 0.303 | 0.939394 |
| 0.287 | 0.944444 |
| 0.274 | 0.949495 |
| 0.263 | 0.954545 |
| 0.248 | 0.959596 |
| 0.245 | 0.964646 |
| 0.233 | 0.969697 |
| 0.221 | 0.974747 |
| 0.218 | 0.979798 |
| 0.196 | 0.984848 |
| 0.182 | 0.989899 |
| 0.175 | 0.994949 |
| 0.168 | 1 |

|  | ST5-BE-PCV14 (Day 300) |
| --- | --- |
| 36.601 | 0.00531915 |
| 34.089 | 0.0106383 |
| 24.927 | 0.0159574 |
| 20.238 | 0.0212766 |
| 19.704 | 0.0265957 |
| 12.465 | 0.0319149 |
| 12.201 | 0.037234 |
| 12.128 | 0.0425532 |
| 11.343 | 0.0478723 |
| 10.838 | 0.0531915 |
| 10.491 | 0.0585106 |
| 9.875 | 0.0638298 |
| 9.728 | 0.0691489 |
| 9.189 | 0.0744681 |
| 9.031 | 0.0797872 |
| 8.315 | 0.0851064 |
| 8.153 | 0.0904255 |
| 7.666 | 0.0957447 |
| 7.485 | 0.101064 |
| 7.247 | 0.106383 |
| 6.849 | 0.111702 |
| 6.763 | 0.117021 |
| 6.532 | 0.12234 |
| 6.509 | 0.12766 |
| 6.323 | 0.132979 |
| 6.134 | 0.138298 |
| 5.831 | 0.143617 |
| 5.664 | 0.148936 |
| 5.344 | 0.154255 |
| 4.499 | 0.159574 |
| 4.463 | 0.164894 |
| 4.105 | 0.170213 |
| 3.814 | 0.175532 |
| 3.731 | 0.180851 |
| 3.675 | 0.18617 |
| 3.61 | 0.191489 |
| 3.451 | 0.196809 |
| 3.419 | 0.202128 |
| 3.376 | 0.207447 |
| 3.355 | 0.212766 |
| 3.341 | 0.218085 |
| 3.181 | 0.223404 |
| 2.984 | 0.228723 |
| 2.982 | 0.234043 |
| 2.936 | 0.239362 |
| 2.931 | 0.244681 |
| 2.918 | 0.25 |
| 2.823 | 0.255319 |
| 2.702 | 0.260638 |
| 2.647 | 0.265957 |
| 2.569 | 0.271277 |
| 2.45 | 0.276596 |
| 2.406 | 0.281915 |
| 2.362 | 0.287234 |
| 2.342 | 0.292553 |
| 2.304 | 0.297872 |
| 2.28 | 0.303191 |
| 2.261 | 0.308511 |
| 2.259 | 0.31383 |
| 2.148 | 0.319149 |
| 2.12 | 0.324468 |
| 2.053 | 0.329787 |
| 2.034 | 0.335106 |
| 2.011 | 0.340426 |
| 1.986 | 0.345745 |
| 1.889 | 0.351064 |
| 1.876 | 0.356383 |
| 1.856 | 0.361702 |
| 1.832 | 0.367021 |
| 1.815 | 0.37234 |
| 1.79 | 0.37766 |
| 1.779 | 0.382979 |
| 1.73 | 0.388298 |
| 1.726 | 0.393617 |
| 1.712 | 0.398936 |
| 1.706 | 0.404255 |
| 1.694 | 0.409574 |
| 1.692 | 0.414894 |
| 1.666 | 0.420213 |
| 1.612 | 0.425532 |
| 1.586 | 0.430851 |
| 1.576 | 0.43617 |
| 1.569 | 0.441489 |
| 1.517 | 0.446809 |
| 1.465 | 0.452128 |
| 1.462 | 0.457447 |
| 1.451 | 0.462766 |
| 1.389 | 0.468085 |
| 1.378 | 0.473404 |
| 1.328 | 0.478723 |
| 1.291 | 0.484043 |
| 1.236 | 0.489362 |
| 1.223 | 0.494681 |
| 1.209 | 0.5 |
| 1.172 | 0.505319 |
| 1.159 | 0.510638 |
| 1.12 | 0.515957 |
| 1.111 | 0.521277 |
| 1.097 | 0.526596 |
| 1.083 | 0.531915 |
| 1.074 | 0.537234 |
| 1.072 | 0.542553 |
| 1.068 | 0.547872 |
| 1.061 | 0.553191 |
| 0.998 | 0.558511 |
| 0.994 | 0.56383 |
| 0.99 | 0.569149 |
| 0.983 | 0.574468 |
| 0.964 | 0.579787 |
| 0.939 | 0.585106 |
| 0.928 | 0.590426 |
| 0.926 | 0.595745 |
| 0.91 | 0.601064 |
| 0.898 | 0.606383 |
| 0.888 | 0.611702 |
| 0.881 | 0.617021 |
| 0.88 | 0.62234 |
| 0.875 | 0.62766 |
| 0.873 | 0.632979 |
| 0.86 | 0.638298 |
| 0.85 | 0.643617 |
| 0.817 | 0.648936 |
| 0.816 | 0.654255 |
| 0.812 | 0.659574 |
| 0.809 | 0.664894 |
| 0.805 | 0.670213 |
| 0.757 | 0.675532 |
| 0.744 | 0.680851 |
| 0.74 | 0.68617 |
| 0.71 | 0.691489 |
| 0.704 | 0.696809 |
| 0.685 | 0.702128 |
| 0.674 | 0.707447 |
| 0.664 | 0.712766 |
| 0.654 | 0.718085 |
| 0.65 | 0.723404 |
| 0.632 | 0.728723 |
| 0.63 | 0.734043 |
| 0.623 | 0.739362 |
| 0.581 | 0.744681 |
| 0.577 | 0.75 |
| 0.576 | 0.755319 |
| 0.572 | 0.760638 |
| 0.566 | 0.765957 |
| 0.558 | 0.771277 |
| 0.549 | 0.776596 |
| 0.548 | 0.781915 |
| 0.538 | 0.787234 |
| 0.53 | 0.792553 |
| 0.527 | 0.797872 |
| 0.492 | 0.803191 |
| 0.486 | 0.808511 |
| 0.482 | 0.81383 |
| 0.48 | 0.819149 |
| 0.477 | 0.824468 |
| 0.477 | 0.829787 |
| 0.456 | 0.835106 |
| 0.437 | 0.840426 |
| 0.435 | 0.845745 |
| 0.434 | 0.851064 |
| 0.419 | 0.856383 |
| 0.416 | 0.861702 |
| 0.411 | 0.867021 |
| 0.41 | 0.87234 |
| 0.405 | 0.87766 |
| 0.399 | 0.882979 |
| 0.398 | 0.888298 |
| 0.395 | 0.893617 |
| 0.387 | 0.898936 |
| 0.378 | 0.904255 |
| 0.365 | 0.909574 |
| 0.365 | 0.914894 |
| 0.35 | 0.920213 |
| 0.346 | 0.925532 |
| 0.331 | 0.930851 |
| 0.322 | 0.93617 |
| 0.308 | 0.941489 |
| 0.307 | 0.946809 |
| 0.297 | 0.952128 |
| 0.289 | 0.957447 |
| 0.284 | 0.962766 |
| 0.265 | 0.968085 |
| 0.237 | 0.973404 |
| 0.204 | 0.978723 |
| 0.203 | 0.984043 |
| 0.18 | 0.989362 |
| 0.152 | 0.994681 |
| 0.048 | 1 |

|  | ST5-Prevenar 13 (Day 300) |
| --- | --- |
| 66.482 | 0.00526316 |
| 61.873 | 0.0105263 |
| 53.452 | 0.0157895 |
| 33.329 | 0.0210526 |
| 30.897 | 0.0263158 |
| 21.344 | 0.0315789 |
| 18.876 | 0.0368421 |
| 18.106 | 0.0421053 |
| 14.147 | 0.0473684 |
| 13.692 | 0.0526316 |
| 10.868 | 0.0578947 |
| 10.702 | 0.0631579 |
| 9.892 | 0.0684211 |
| 9.822 | 0.0736842 |
| 9.67 | 0.0789474 |
| 9.214 | 0.0842105 |
| 8.881 | 0.0894737 |
| 8.87 | 0.0947368 |
| 8.722 | 0.1 |
| 8.193 | 0.105263 |
| 7.845 | 0.110526 |
| 7.574 | 0.115789 |
| 7.088 | 0.121053 |
| 6.693 | 0.126316 |
| 5.566 | 0.131579 |
| 5.514 | 0.136842 |
| 5.492 | 0.142105 |
| 5.458 | 0.147368 |
| 5.366 | 0.152632 |
| 5.194 | 0.157895 |
| 5.126 | 0.163158 |
| 5.04 | 0.168421 |
| 4.94 | 0.173684 |
| 4.739 | 0.178947 |
| 4.58 | 0.184211 |
| 4.524 | 0.189474 |
| 4.179 | 0.194737 |
| 4.161 | 0.2 |
| 4.018 | 0.205263 |
| 3.645 | 0.210526 |
| 3.509 | 0.215789 |
| 3.492 | 0.221053 |
| 3.461 | 0.226316 |
| 3.408 | 0.231579 |
| 3.271 | 0.236842 |
| 3.239 | 0.242105 |
| 3.157 | 0.247368 |
| 3.054 | 0.252632 |
| 2.97 | 0.257895 |
| 2.955 | 0.263158 |
| 2.909 | 0.268421 |
| 2.854 | 0.273684 |
| 2.667 | 0.278947 |
| 2.562 | 0.284211 |
| 2.558 | 0.289474 |
| 2.55 | 0.294737 |
| 2.47 | 0.3 |
| 2.466 | 0.305263 |
| 2.444 | 0.310526 |
| 2.433 | 0.315789 |
| 2.385 | 0.321053 |
| 2.379 | 0.326316 |
| 2.319 | 0.331579 |
| 2.283 | 0.336842 |
| 2.258 | 0.342105 |
| 2.246 | 0.347368 |
| 2.221 | 0.352632 |
| 2.211 | 0.357895 |
| 2.204 | 0.363158 |
| 2.18 | 0.368421 |
| 2.152 | 0.373684 |
| 2.152 | 0.378947 |
| 2.148 | 0.384211 |
| 2.112 | 0.389474 |
| 2.109 | 0.394737 |
| 2.088 | 0.4 |
| 2.076 | 0.405263 |
| 1.992 | 0.410526 |
| 1.86 | 0.415789 |
| 1.846 | 0.421053 |
| 1.817 | 0.426316 |
| 1.769 | 0.431579 |
| 1.762 | 0.436842 |
| 1.762 | 0.442105 |
| 1.727 | 0.447368 |
| 1.718 | 0.452632 |
| 1.707 | 0.457895 |
| 1.697 | 0.463158 |
| 1.685 | 0.468421 |
| 1.669 | 0.473684 |
| 1.641 | 0.478947 |
| 1.615 | 0.484211 |
| 1.611 | 0.489474 |
| 1.596 | 0.494737 |
| 1.59 | 0.5 |
| 1.583 | 0.505263 |
| 1.568 | 0.510526 |
| 1.558 | 0.515789 |
| 1.52 | 0.521053 |
| 1.518 | 0.526316 |
| 1.509 | 0.531579 |
| 1.504 | 0.536842 |
| 1.489 | 0.542105 |
| 1.481 | 0.547368 |
| 1.48 | 0.552632 |
| 1.452 | 0.557895 |
| 1.442 | 0.563158 |
| 1.43 | 0.568421 |
| 1.428 | 0.573684 |
| 1.409 | 0.578947 |
| 1.402 | 0.584211 |
| 1.359 | 0.589474 |
| 1.336 | 0.594737 |
| 1.306 | 0.6 |
| 1.289 | 0.605263 |
| 1.269 | 0.610526 |
| 1.253 | 0.615789 |
| 1.227 | 0.621053 |
| 1.226 | 0.626316 |
| 1.218 | 0.631579 |
| 1.209 | 0.636842 |
| 1.196 | 0.642105 |
| 1.178 | 0.647368 |
| 1.135 | 0.652632 |
| 1.118 | 0.657895 |
| 1.088 | 0.663158 |
| 1.079 | 0.668421 |
| 1.075 | 0.673684 |
| 1.059 | 0.678947 |
| 1.044 | 0.684211 |
| 1.036 | 0.689474 |
| 1.032 | 0.694737 |
| 0.999 | 0.7 |
| 0.986 | 0.705263 |
| 0.981 | 0.710526 |
| 0.971 | 0.715789 |
| 0.954 | 0.721053 |
| 0.942 | 0.726316 |
| 0.899 | 0.731579 |
| 0.861 | 0.736842 |
| 0.838 | 0.742105 |
| 0.828 | 0.747368 |
| 0.806 | 0.752632 |
| 0.793 | 0.757895 |
| 0.745 | 0.763158 |
| 0.741 | 0.768421 |
| 0.729 | 0.773684 |
| 0.726 | 0.778947 |
| 0.689 | 0.784211 |
| 0.684 | 0.789474 |
| 0.681 | 0.794737 |
| 0.647 | 0.8 |
| 0.612 | 0.805263 |
| 0.606 | 0.810526 |
| 0.6 | 0.815789 |
| 0.542 | 0.821053 |
| 0.531 | 0.826316 |
| 0.527 | 0.831579 |
| 0.522 | 0.836842 |
| 0.511 | 0.842105 |
| 0.503 | 0.847368 |
| 0.493 | 0.852632 |
| 0.485 | 0.857895 |
| 0.483 | 0.863158 |
| 0.478 | 0.868421 |
| 0.458 | 0.873684 |
| 0.445 | 0.878947 |
| 0.404 | 0.884211 |
| 0.382 | 0.889474 |
| 0.381 | 0.894737 |
| 0.38 | 0.9 |
| 0.367 | 0.905263 |
| 0.349 | 0.910526 |
| 0.337 | 0.915789 |
| 0.331 | 0.921053 |
| 0.32 | 0.926316 |
| 0.313 | 0.931579 |
| 0.304 | 0.936842 |
| 0.273 | 0.942105 |
| 0.234 | 0.947368 |
| 0.206 | 0.952632 |
| 0.205 | 0.957895 |
| 0.2 | 0.963158 |
| 0.198 | 0.968421 |
| 0.195 | 0.973684 |
| 0.186 | 0.978947 |
| 0.182 | 0.984211 |
| 0.173 | 0.989474 |
| 0.172 | 0.994737 |
| 0.112 | 1 |

BECT056 - RCDC_Serotype 6B

|  | ST 6B-BE-PCV14 (Day 84) |
| --- | --- |
| 157.664 | 0.0052356 |
| 130.636 | 0.0104712 |
| 70.632 | 0.0157068 |
| 66.941 | 0.0209424 |
| 66.383 | 0.026178 |
| 61.194 | 0.0314136 |
| 53.925 | 0.0366492 |
| 48.458 | 0.0418848 |
| 45.834 | 0.0471204 |
| 45.233 | 0.052356 |
| 38.684 | 0.0575916 |
| 36.085 | 0.0628272 |
| 35.959 | 0.0680628 |
| 35.259 | 0.0732984 |
| 32.236 | 0.078534 |
| 28.58 | 0.0837696 |
| 26.902 | 0.0890052 |
| 21.744 | 0.0942408 |
| 21.483 | 0.0994764 |
| 19.914 | 0.104712 |
| 19.435 | 0.109948 |
| 19.428 | 0.115183 |
| 18.629 | 0.120419 |
| 16.144 | 0.125654 |
| 14.573 | 0.13089 |
| 14.271 | 0.136126 |
| 14.165 | 0.141361 |
| 13.664 | 0.146597 |
| 13.592 | 0.151832 |
| 13.218 | 0.157068 |
| 12.363 | 0.162304 |
| 12.064 | 0.167539 |
| 11.423 | 0.172775 |
| 10.802 | 0.17801 |
| 10.747 | 0.183246 |
| 9.94 | 0.188482 |
| 9.294 | 0.193717 |
| 8.94 | 0.198953 |
| 8.697 | 0.204188 |
| 8.618 | 0.209424 |
| 8.583 | 0.21466 |
| 8.41 | 0.219895 |
| 7.506 | 0.225131 |
| 7.195 | 0.230366 |
| 7.067 | 0.235602 |
| 7.032 | 0.240838 |
| 6.925 | 0.246073 |
| 6.883 | 0.251309 |
| 6.82 | 0.256545 |
| 6.811 | 0.26178 |
| 6.791 | 0.267016 |
| 6.736 | 0.272251 |
| 6.526 | 0.277487 |
| 6.373 | 0.282723 |
| 6.194 | 0.287958 |
| 6.064 | 0.293194 |
| 5.75 | 0.298429 |
| 5.64 | 0.303665 |
| 5.585 | 0.308901 |
| 5.567 | 0.314136 |
| 5.394 | 0.319372 |
| 5.33 | 0.324607 |
| 5.193 | 0.329843 |
| 5.058 | 0.335079 |
| 4.921 | 0.340314 |
| 4.914 | 0.34555 |
| 4.881 | 0.350785 |
| 4.781 | 0.356021 |
| 4.574 | 0.361257 |
| 4.557 | 0.366492 |
| 4.425 | 0.371728 |
| 4.248 | 0.376963 |
| 4.151 | 0.382199 |
| 4.144 | 0.387435 |
| 3.891 | 0.39267 |
| 3.795 | 0.397906 |
| 3.747 | 0.403141 |
| 3.677 | 0.408377 |
| 3.652 | 0.413613 |
| 3.463 | 0.418848 |
| 3.432 | 0.424084 |
| 3.198 | 0.429319 |
| 3.181 | 0.434555 |
| 3.174 | 0.439791 |
| 3.011 | 0.445026 |
| 2.973 | 0.450262 |
| 2.928 | 0.455497 |
| 2.899 | 0.460733 |
| 2.88 | 0.465969 |
| 2.88 | 0.471204 |
| 2.815 | 0.47644 |
| 2.811 | 0.481675 |
| 2.8 | 0.486911 |
| 2.734 | 0.492147 |
| 2.601 | 0.497382 |
| 2.599 | 0.502618 |
| 2.541 | 0.507853 |
| 2.534 | 0.513089 |
| 2.329 | 0.518325 |
| 2.324 | 0.52356 |
| 2.278 | 0.528796 |
| 2.225 | 0.534031 |
| 2.204 | 0.539267 |
| 2.139 | 0.544503 |
| 2.098 | 0.549738 |
| 2.096 | 0.554974 |
| 2.026 | 0.560209 |
| 1.982 | 0.565445 |
| 1.952 | 0.570681 |
| 1.926 | 0.575916 |
| 1.895 | 0.581152 |
| 1.794 | 0.586387 |
| 1.73 | 0.591623 |
| 1.697 | 0.596859 |
| 1.662 | 0.602094 |
| 1.645 | 0.60733 |
| 1.626 | 0.612565 |
| 1.597 | 0.617801 |
| 1.589 | 0.623037 |
| 1.55 | 0.628272 |
| 1.547 | 0.633508 |
| 1.522 | 0.638743 |
| 1.495 | 0.643979 |
| 1.48 | 0.649215 |
| 1.441 | 0.65445 |
| 1.415 | 0.659686 |
| 1.327 | 0.664921 |
| 1.31 | 0.670157 |
| 1.256 | 0.675393 |
| 1.252 | 0.680628 |
| 1.24 | 0.685864 |
| 1.184 | 0.691099 |
| 1.155 | 0.696335 |
| 1.091 | 0.701571 |
| 1.066 | 0.706806 |
| 1.044 | 0.712042 |
| 1.016 | 0.717277 |
| 1 | 0.722513 |
| 0.993 | 0.727749 |
| 0.989 | 0.732984 |
| 0.969 | 0.73822 |
| 0.961 | 0.743455 |
| 0.86 | 0.748691 |
| 0.824 | 0.753927 |
| 0.819 | 0.759162 |
| 0.768 | 0.764398 |
| 0.743 | 0.769634 |
| 0.701 | 0.774869 |
| 0.685 | 0.780105 |
| 0.65 | 0.78534 |
| 0.614 | 0.790576 |
| 0.566 | 0.795812 |
| 0.55 | 0.801047 |
| 0.514 | 0.806283 |
| 0.467 | 0.811518 |
| 0.45 | 0.816754 |
| 0.45 | 0.82199 |
| 0.434 | 0.827225 |
| 0.428 | 0.832461 |
| 0.394 | 0.837696 |
| 0.389 | 0.842932 |
| 0.377 | 0.848168 |
| 0.361 | 0.853403 |
| 0.342 | 0.858639 |
| 0.331 | 0.863874 |
| 0.314 | 0.86911 |
| 0.313 | 0.874346 |
| 0.303 | 0.879581 |
| 0.293 | 0.884817 |
| 0.284 | 0.890052 |
| 0.284 | 0.895288 |
| 0.28 | 0.900524 |
| 0.271 | 0.905759 |
| 0.217 | 0.910995 |
| 0.208 | 0.91623 |
| 0.206 | 0.921466 |
| 0.195 | 0.926702 |
| 0.194 | 0.931937 |
| 0.194 | 0.937173 |
| 0.166 | 0.942408 |
| 0.159 | 0.947644 |
| 0.148 | 0.95288 |
| 0.145 | 0.958115 |
| 0.129 | 0.963351 |
| 0.113 | 0.968586 |
| 0.106 | 0.973822 |
| 0.101 | 0.979058 |
| 0.091 | 0.984293 |
| 0.076 | 0.989529 |
| 0.05 | 0.994764 |
| 0.036 | 1 |

|  | ST 6B-Prevenar 13 (Day 84) |
| --- | --- |
| 236.673 | 0.00505051 |
| 122.155 | 0.010101 |
| 121.716 | 0.0151515 |
| 79.566 | 0.020202 |
| 68.037 | 0.0252525 |
| 63.81 | 0.030303 |
| 46.536 | 0.0353535 |
| 41.429 | 0.040404 |
| 38.102 | 0.0454545 |
| 36.979 | 0.0505051 |
| 35.821 | 0.0555556 |
| 32.726 | 0.0606061 |
| 28.06 | 0.0656566 |
| 27.094 | 0.0707071 |
| 25.169 | 0.0757576 |
| 24.714 | 0.0808081 |
| 22.082 | 0.0858586 |
| 21.276 | 0.0909091 |
| 21.198 | 0.0959596 |
| 20.039 | 0.10101 |
| 19.825 | 0.106061 |
| 16.082 | 0.111111 |
| 14.022 | 0.116162 |
| 14.022 | 0.121212 |
| 13.524 | 0.126263 |
| 12.955 | 0.131313 |
| 12.752 | 0.136364 |
| 12.628 | 0.141414 |
| 12.189 | 0.146465 |
| 11.737 | 0.151515 |
| 10.595 | 0.156566 |
| 10.518 | 0.161616 |
| 10.27 | 0.166667 |
| 9.502 | 0.171717 |
| 8.697 | 0.176768 |
| 8.654 | 0.181818 |
| 8.104 | 0.186869 |
| 8.072 | 0.191919 |
| 7.718 | 0.19697 |
| 7.596 | 0.20202 |
| 7.451 | 0.207071 |
| 7.211 | 0.212121 |
| 6.85 | 0.217172 |
| 6.504 | 0.222222 |
| 6.4 | 0.227273 |
| 6.089 | 0.232323 |
| 5.933 | 0.237374 |
| 5.468 | 0.242424 |
| 5.333 | 0.247475 |
| 4.953 | 0.252525 |
| 4.527 | 0.257576 |
| 3.818 | 0.262626 |
| 3.685 | 0.267677 |
| 3.649 | 0.272727 |
| 3.492 | 0.277778 |
| 3.335 | 0.282828 |
| 3.3 | 0.287879 |
| 3.188 | 0.292929 |
| 3.173 | 0.29798 |
| 2.968 | 0.30303 |
| 2.955 | 0.308081 |
| 2.931 | 0.313131 |
| 2.885 | 0.318182 |
| 2.822 | 0.323232 |
| 2.675 | 0.328283 |
| 2.626 | 0.333333 |
| 2.621 | 0.338384 |
| 2.589 | 0.343434 |
| 2.568 | 0.348485 |
| 2.485 | 0.353535 |
| 2.452 | 0.358586 |
| 2.444 | 0.363636 |
| 2.414 | 0.368687 |
| 2.402 | 0.373737 |
| 2.398 | 0.378788 |
| 2.359 | 0.383838 |
| 2.259 | 0.388889 |
| 2.244 | 0.393939 |
| 2.174 | 0.39899 |
| 2.127 | 0.40404 |
| 2.111 | 0.409091 |
| 2.061 | 0.414141 |
| 2.033 | 0.419192 |
| 2.022 | 0.424242 |
| 1.906 | 0.429293 |
| 1.901 | 0.434343 |
| 1.887 | 0.439394 |
| 1.884 | 0.444444 |
| 1.799 | 0.449495 |
| 1.778 | 0.454545 |
| 1.767 | 0.459596 |
| 1.63 | 0.464646 |
| 1.622 | 0.469697 |
| 1.616 | 0.474747 |
| 1.614 | 0.479798 |
| 1.548 | 0.484848 |
| 1.547 | 0.489899 |
| 1.512 | 0.494949 |
| 1.511 | 0.5 |
| 1.482 | 0.505051 |
| 1.445 | 0.510101 |
| 1.421 | 0.515152 |
| 1.374 | 0.520202 |
| 1.35 | 0.525253 |
| 1.32 | 0.530303 |
| 1.308 | 0.535354 |
| 1.306 | 0.540404 |
| 1.283 | 0.545455 |
| 1.27 | 0.550505 |
| 1.254 | 0.555556 |
| 1.235 | 0.560606 |
| 1.177 | 0.565657 |
| 1.169 | 0.570707 |
| 1.164 | 0.575758 |
| 1.135 | 0.580808 |
| 1.127 | 0.585859 |
| 1.052 | 0.590909 |
| 0.98 | 0.59596 |
| 0.929 | 0.60101 |
| 0.927 | 0.606061 |
| 0.903 | 0.611111 |
| 0.874 | 0.616162 |
| 0.874 | 0.621212 |
| 0.864 | 0.626263 |
| 0.834 | 0.631313 |
| 0.813 | 0.636364 |
| 0.784 | 0.641414 |
| 0.757 | 0.646465 |
| 0.748 | 0.651515 |
| 0.741 | 0.656566 |
| 0.73 | 0.661616 |
| 0.729 | 0.666667 |
| 0.723 | 0.671717 |
| 0.673 | 0.676768 |
| 0.658 | 0.681818 |
| 0.641 | 0.686869 |
| 0.617 | 0.691919 |
| 0.606 | 0.69697 |
| 0.603 | 0.70202 |
| 0.596 | 0.707071 |
| 0.567 | 0.712121 |
| 0.55 | 0.717172 |
| 0.531 | 0.722222 |
| 0.516 | 0.727273 |
| 0.516 | 0.732323 |
| 0.504 | 0.737374 |
| 0.502 | 0.742424 |
| 0.459 | 0.747475 |
| 0.459 | 0.752525 |
| 0.453 | 0.757576 |
| 0.423 | 0.762626 |
| 0.421 | 0.767677 |
| 0.412 | 0.772727 |
| 0.411 | 0.777778 |
| 0.403 | 0.782828 |
| 0.394 | 0.787879 |
| 0.389 | 0.792929 |
| 0.384 | 0.79798 |
| 0.381 | 0.80303 |
| 0.372 | 0.808081 |
| 0.364 | 0.813131 |
| 0.36 | 0.818182 |
| 0.359 | 0.823232 |
| 0.357 | 0.828283 |
| 0.343 | 0.833333 |
| 0.341 | 0.838384 |
| 0.337 | 0.843434 |
| 0.335 | 0.848485 |
| 0.327 | 0.853535 |
| 0.309 | 0.858586 |
| 0.279 | 0.863636 |
| 0.25 | 0.868687 |
| 0.229 | 0.873737 |
| 0.218 | 0.878788 |
| 0.214 | 0.883838 |
| 0.21 | 0.888889 |
| 0.208 | 0.893939 |
| 0.206 | 0.89899 |
| 0.205 | 0.90404 |
| 0.203 | 0.909091 |
| 0.185 | 0.914141 |
| 0.18 | 0.919192 |
| 0.177 | 0.924242 |
| 0.172 | 0.929293 |
| 0.17 | 0.934343 |
| 0.168 | 0.939394 |
| 0.167 | 0.944444 |
| 0.167 | 0.949495 |
| 0.148 | 0.954545 |
| 0.136 | 0.959596 |
| 0.118 | 0.964646 |
| 0.118 | 0.969697 |
| 0.118 | 0.974747 |
| 0.108 | 0.979798 |
| 0.101 | 0.984848 |
| 0.083 | 0.989899 |
| 0.055 | 0.994949 |
| 0.033 | 1 |

|  | ST 6B-BE-PCV14 (Day 300) |
| --- | --- |
| 56.269 | 0.00531915 |
| 52.75 | 0.0106383 |
| 41.572 | 0.0159574 |
| 40.305 | 0.0212766 |
| 39.635 | 0.0265957 |
| 39.534 | 0.0319149 |
| 36.846 | 0.037234 |
| 36.314 | 0.0425532 |
| 35.937 | 0.0478723 |
| 34.217 | 0.0531915 |
| 34.163 | 0.0585106 |
| 33.152 | 0.0638298 |
| 31.26 | 0.0691489 |
| 30.565 | 0.0744681 |
| 29.872 | 0.0797872 |
| 27.71 | 0.0851064 |
| 26.905 | 0.0904255 |
| 25.779 | 0.0957447 |
| 25.654 | 0.101064 |
| 25.498 | 0.106383 |
| 25.383 | 0.111702 |
| 24.347 | 0.117021 |
| 23.535 | 0.12234 |
| 23.428 | 0.12766 |
| 22.887 | 0.132979 |
| 22.815 | 0.138298 |
| 22.209 | 0.143617 |
| 21.785 | 0.148936 |
| 21.446 | 0.154255 |
| 20.602 | 0.159574 |
| 19.863 | 0.164894 |
| 19.442 | 0.170213 |
| 19.066 | 0.175532 |
| 18.229 | 0.180851 |
| 18.043 | 0.18617 |
| 17.825 | 0.191489 |
| 17.349 | 0.196809 |
| 17.025 | 0.202128 |
| 16.698 | 0.207447 |
| 16.097 | 0.212766 |
| 15.989 | 0.218085 |
| 15.239 | 0.223404 |
| 15.093 | 0.228723 |
| 14.477 | 0.234043 |
| 14.195 | 0.239362 |
| 14 | 0.244681 |
| 13.909 | 0.25 |
| 13.543 | 0.255319 |
| 13.277 | 0.260638 |
| 13.046 | 0.265957 |
| 12.934 | 0.271277 |
| 12.656 | 0.276596 |
| 11.944 | 0.281915 |
| 11.323 | 0.287234 |
| 11.158 | 0.292553 |
| 10.831 | 0.297872 |
| 10.726 | 0.303191 |
| 10.648 | 0.308511 |
| 10.616 | 0.31383 |
| 10.479 | 0.319149 |
| 10.444 | 0.324468 |
| 10.367 | 0.329787 |
| 9.985 | 0.335106 |
| 9.908 | 0.340426 |
| 9.814 | 0.345745 |
| 9.234 | 0.351064 |
| 9.11 | 0.356383 |
| 8.936 | 0.361702 |
| 8.872 | 0.367021 |
| 8.821 | 0.37234 |
| 8.804 | 0.37766 |
| 8.752 | 0.382979 |
| 8.739 | 0.388298 |
| 8.549 | 0.393617 |
| 8.489 | 0.398936 |
| 8.293 | 0.404255 |
| 8.21 | 0.409574 |
| 8.132 | 0.414894 |
| 8.04 | 0.420213 |
| 8.015 | 0.425532 |
| 8.004 | 0.430851 |
| 7.989 | 0.43617 |
| 7.794 | 0.441489 |
| 7.646 | 0.446809 |
| 7.465 | 0.452128 |
| 7.311 | 0.457447 |
| 7.25 | 0.462766 |
| 7.223 | 0.468085 |
| 7.141 | 0.473404 |
| 7.083 | 0.478723 |
| 7.079 | 0.484043 |
| 6.721 | 0.489362 |
| 6.535 | 0.494681 |
| 6.245 | 0.5 |
| 6.175 | 0.505319 |
| 6.151 | 0.510638 |
| 5.926 | 0.515957 |
| 5.899 | 0.521277 |
| 5.893 | 0.526596 |
| 5.768 | 0.531915 |
| 5.653 | 0.537234 |
| 5.303 | 0.542553 |
| 5.278 | 0.547872 |
| 5.223 | 0.553191 |
| 5.113 | 0.558511 |
| 4.919 | 0.56383 |
| 4.869 | 0.569149 |
| 4.827 | 0.574468 |
| 4.824 | 0.579787 |
| 4.791 | 0.585106 |
| 4.758 | 0.590426 |
| 4.712 | 0.595745 |
| 4.6 | 0.601064 |
| 4.522 | 0.606383 |
| 4.366 | 0.611702 |
| 4.186 | 0.617021 |
| 4.151 | 0.62234 |
| 4.12 | 0.62766 |
| 3.844 | 0.632979 |
| 3.83 | 0.638298 |
| 3.782 | 0.643617 |
| 3.684 | 0.648936 |
| 3.644 | 0.654255 |
| 3.636 | 0.659574 |
| 3.604 | 0.664894 |
| 3.566 | 0.670213 |
| 3.518 | 0.675532 |
| 3.513 | 0.680851 |
| 3.499 | 0.68617 |
| 3.464 | 0.691489 |
| 3.259 | 0.696809 |
| 3.188 | 0.702128 |
| 3.112 | 0.707447 |
| 3.1 | 0.712766 |
| 2.954 | 0.718085 |
| 2.687 | 0.723404 |
| 2.589 | 0.728723 |
| 2.511 | 0.734043 |
| 2.502 | 0.739362 |
| 2.502 | 0.744681 |
| 2.394 | 0.75 |
| 2.37 | 0.755319 |
| 2.334 | 0.760638 |
| 2.228 | 0.765957 |
| 2.086 | 0.771277 |
| 1.848 | 0.776596 |
| 1.838 | 0.781915 |
| 1.806 | 0.787234 |
| 1.607 | 0.792553 |
| 1.594 | 0.797872 |
| 1.48 | 0.803191 |
| 1.414 | 0.808511 |
| 1.366 | 0.81383 |
| 1.351 | 0.819149 |
| 1.348 | 0.824468 |
| 1.258 | 0.829787 |
| 1.058 | 0.835106 |
| 0.974 | 0.840426 |
| 0.916 | 0.845745 |
| 0.857 | 0.851064 |
| 0.852 | 0.856383 |
| 0.84 | 0.861702 |
| 0.837 | 0.867021 |
| 0.824 | 0.87234 |
| 0.816 | 0.87766 |
| 0.803 | 0.882979 |
| 0.788 | 0.888298 |
| 0.775 | 0.893617 |
| 0.774 | 0.898936 |
| 0.741 | 0.904255 |
| 0.734 | 0.909574 |
| 0.724 | 0.914894 |
| 0.687 | 0.920213 |
| 0.646 | 0.925532 |
| 0.585 | 0.930851 |
| 0.579 | 0.93617 |
| 0.53 | 0.941489 |
| 0.514 | 0.946809 |
| 0.482 | 0.952128 |
| 0.391 | 0.957447 |
| 0.389 | 0.962766 |
| 0.373 | 0.968085 |
| 0.352 | 0.973404 |
| 0.257 | 0.978723 |
| 0.211 | 0.984043 |
| 0.203 | 0.989362 |
| 0.156 | 0.994681 |
| 0.051 | 1 |

|  | ST 6B-Prevenar 13 (Day 300) |
| --- | --- |
| 189.052 | 0.00526316 |
| 112.314 | 0.0105263 |
| 107.174 | 0.0157895 |
| 101.263 | 0.0210526 |
| 90.233 | 0.0263158 |
| 75.522 | 0.0315789 |
| 71.176 | 0.0368421 |
| 65.781 | 0.0421053 |
| 63.388 | 0.0473684 |
| 59.519 | 0.0526316 |
| 59.289 | 0.0578947 |
| 57.492 | 0.0631579 |
| 44.264 | 0.0684211 |
| 44.159 | 0.0736842 |
| 42.371 | 0.0789474 |
| 39.154 | 0.0842105 |
| 39.018 | 0.0894737 |
| 38.641 | 0.0947368 |
| 37.092 | 0.1 |
| 36.333 | 0.105263 |
| 34.773 | 0.110526 |
| 31.668 | 0.115789 |
| 31.159 | 0.121053 |
| 30.669 | 0.126316 |
| 29.471 | 0.131579 |
| 28.398 | 0.136842 |
| 26.712 | 0.142105 |
| 25.748 | 0.147368 |
| 23.237 | 0.152632 |
| 21.434 | 0.157895 |
| 19.743 | 0.163158 |
| 19.672 | 0.168421 |
| 19.635 | 0.173684 |
| 19.629 | 0.178947 |
| 19.35 | 0.184211 |
| 16.461 | 0.189474 |
| 15.745 | 0.194737 |
| 15.717 | 0.2 |
| 15.147 | 0.205263 |
| 14.471 | 0.210526 |
| 14.017 | 0.215789 |
| 13.95 | 0.221053 |
| 13.896 | 0.226316 |
| 13.729 | 0.231579 |
| 13.619 | 0.236842 |
| 13.03 | 0.242105 |
| 12.706 | 0.247368 |
| 12.594 | 0.252632 |
| 12.393 | 0.257895 |
| 12.241 | 0.263158 |
| 11.964 | 0.268421 |
| 11.853 | 0.273684 |
| 11.827 | 0.278947 |
| 11.679 | 0.284211 |
| 11.598 | 0.289474 |
| 11.54 | 0.294737 |
| 11.521 | 0.3 |
| 11.506 | 0.305263 |
| 11.353 | 0.310526 |
| 11.155 | 0.315789 |
| 11.05 | 0.321053 |
| 11.034 | 0.326316 |
| 10.979 | 0.331579 |
| 10.839 | 0.336842 |
| 10.817 | 0.342105 |
| 10.462 | 0.347368 |
| 10.11 | 0.352632 |
| 9.834 | 0.357895 |
| 9.591 | 0.363158 |
| 9.539 | 0.368421 |
| 9.531 | 0.373684 |
| 9.249 | 0.378947 |
| 9.241 | 0.384211 |
| 9.236 | 0.389474 |
| 9.2 | 0.394737 |
| 9.182 | 0.4 |
| 8.979 | 0.405263 |
| 8.941 | 0.410526 |
| 8.646 | 0.415789 |
| 8.473 | 0.421053 |
| 8.287 | 0.426316 |
| 8.095 | 0.431579 |
| 8.044 | 0.436842 |
| 8.025 | 0.442105 |
| 7.815 | 0.447368 |
| 7.596 | 0.452632 |
| 7.551 | 0.457895 |
| 7.55 | 0.463158 |
| 7.496 | 0.468421 |
| 7.42 | 0.473684 |
| 7.392 | 0.478947 |
| 7.303 | 0.484211 |
| 7.227 | 0.489474 |
| 7.054 | 0.494737 |
| 7.036 | 0.5 |
| 7.028 | 0.505263 |
| 6.922 | 0.510526 |
| 6.816 | 0.515789 |
| 6.805 | 0.521053 |
| 6.795 | 0.526316 |
| 6.636 | 0.531579 |
| 6.524 | 0.536842 |
| 6.417 | 0.542105 |
| 6.399 | 0.547368 |
| 6.378 | 0.552632 |
| 6.233 | 0.557895 |
| 6.203 | 0.563158 |
| 6.202 | 0.568421 |
| 6.061 | 0.573684 |
| 5.944 | 0.578947 |
| 5.937 | 0.584211 |
| 5.88 | 0.589474 |
| 5.829 | 0.594737 |
| 5.796 | 0.6 |
| 5.76 | 0.605263 |
| 5.706 | 0.610526 |
| 5.574 | 0.615789 |
| 5.41 | 0.621053 |
| 5.382 | 0.626316 |
| 5.23 | 0.631579 |
| 4.913 | 0.636842 |
| 4.866 | 0.642105 |
| 4.86 | 0.647368 |
| 4.659 | 0.652632 |
| 4.566 | 0.657895 |
| 4.561 | 0.663158 |
| 4.422 | 0.668421 |
| 4.409 | 0.673684 |
| 4.403 | 0.678947 |
| 4.224 | 0.684211 |
| 4.131 | 0.689474 |
| 4.122 | 0.694737 |
| 4.091 | 0.7 |
| 3.986 | 0.705263 |
| 3.884 | 0.710526 |
| 3.784 | 0.715789 |
| 3.656 | 0.721053 |
| 3.568 | 0.726316 |
| 3.564 | 0.731579 |
| 3.394 | 0.736842 |
| 3.317 | 0.742105 |
| 3.308 | 0.747368 |
| 3.017 | 0.752632 |
| 3.014 | 0.757895 |
| 2.802 | 0.763158 |
| 2.625 | 0.768421 |
| 2.583 | 0.773684 |
| 2.557 | 0.778947 |
| 2.47 | 0.784211 |
| 2.438 | 0.789474 |
| 2.403 | 0.794737 |
| 2.268 | 0.8 |
| 2.254 | 0.805263 |
| 2.192 | 0.810526 |
| 2.132 | 0.815789 |
| 2.084 | 0.821053 |
| 1.904 | 0.826316 |
| 1.88 | 0.831579 |
| 1.87 | 0.836842 |
| 1.815 | 0.842105 |
| 1.759 | 0.847368 |
| 1.639 | 0.852632 |
| 1.615 | 0.857895 |
| 1.395 | 0.863158 |
| 1.389 | 0.868421 |
| 1.283 | 0.873684 |
| 1.254 | 0.878947 |
| 1.195 | 0.884211 |
| 1.171 | 0.889474 |
| 0.974 | 0.894737 |
| 0.953 | 0.9 |
| 0.847 | 0.905263 |
| 0.774 | 0.910526 |
| 0.746 | 0.915789 |
| 0.711 | 0.921053 |
| 0.637 | 0.926316 |
| 0.624 | 0.931579 |
| 0.611 | 0.936842 |
| 0.58 | 0.942105 |
| 0.451 | 0.947368 |
| 0.404 | 0.952632 |
| 0.387 | 0.957895 |
| 0.345 | 0.963158 |
| 0.336 | 0.968421 |
| 0.308 | 0.973684 |
| 0.271 | 0.978947 |
| 0.238 | 0.984211 |
| 0.196 | 0.989474 |
| 0.154 | 0.994737 |
| 0.045 | 1 |

BECT056 - RCDC_Serotype 7F

|  | ST 7F-BE-PCV14 (Day 84) |
| --- | --- |
| 91.768 | 0.0052356 |
| 64.632 | 0.0104712 |
| 58.938 | 0.0157068 |
| 49.8 | 0.0209424 |
| 45.72 | 0.026178 |
| 39.83 | 0.0314136 |
| 38.539 | 0.0366492 |
| 32.063 | 0.0418848 |
| 27.85 | 0.0471204 |
| 24.407 | 0.052356 |
| 23.027 | 0.0575916 |
| 22.579 | 0.0628272 |
| 22.286 | 0.0680628 |
| 22.159 | 0.0732984 |
| 19.685 | 0.078534 |
| 17.435 | 0.0837696 |
| 16.581 | 0.0890052 |
| 16.224 | 0.0942408 |
| 16.116 | 0.0994764 |
| 15.574 | 0.104712 |
| 14.941 | 0.109948 |
| 14.674 | 0.115183 |
| 14.227 | 0.120419 |
| 13.555 | 0.125654 |
| 13.025 | 0.13089 |
| 11.955 | 0.136126 |
| 11.392 | 0.141361 |
| 10.213 | 0.146597 |
| 10.115 | 0.151832 |
| 9.617 | 0.157068 |
| 9.483 | 0.162304 |
| 9.141 | 0.167539 |
| 8.793 | 0.172775 |
| 8.616 | 0.17801 |
| 8.119 | 0.183246 |
| 7.97 | 0.188482 |
| 7.948 | 0.193717 |
| 7.14 | 0.198953 |
| 6.78 | 0.204188 |
| 6.181 | 0.209424 |
| 6.102 | 0.21466 |
| 5.756 | 0.219895 |
| 5.548 | 0.225131 |
| 5.537 | 0.230366 |
| 5.505 | 0.235602 |
| 5.456 | 0.240838 |
| 5.38 | 0.246073 |
| 5.106 | 0.251309 |
| 5.028 | 0.256545 |
| 4.981 | 0.26178 |
| 4.889 | 0.267016 |
| 4.883 | 0.272251 |
| 4.872 | 0.277487 |
| 4.839 | 0.282723 |
| 4.739 | 0.287958 |
| 4.681 | 0.293194 |
| 4.581 | 0.298429 |
| 4.469 | 0.303665 |
| 4.419 | 0.308901 |
| 4.273 | 0.314136 |
| 4.267 | 0.319372 |
| 4.072 | 0.324607 |
| 4.062 | 0.329843 |
| 4.044 | 0.335079 |
| 4.009 | 0.340314 |
| 3.936 | 0.34555 |
| 3.857 | 0.350785 |
| 3.849 | 0.356021 |
| 3.652 | 0.361257 |
| 3.651 | 0.366492 |
| 3.619 | 0.371728 |
| 3.577 | 0.376963 |
| 3.513 | 0.382199 |
| 3.376 | 0.387435 |
| 3.36 | 0.39267 |
| 3.35 | 0.397906 |
| 3.342 | 0.403141 |
| 3.282 | 0.408377 |
| 3.276 | 0.413613 |
| 3.26 | 0.418848 |
| 3.153 | 0.424084 |
| 3.131 | 0.429319 |
| 3.127 | 0.434555 |
| 3.1 | 0.439791 |
| 3.06 | 0.445026 |
| 3.049 | 0.450262 |
| 2.978 | 0.455497 |
| 2.933 | 0.460733 |
| 2.933 | 0.465969 |
| 2.908 | 0.471204 |
| 2.823 | 0.47644 |
| 2.801 | 0.481675 |
| 2.762 | 0.486911 |
| 2.698 | 0.492147 |
| 2.696 | 0.497382 |
| 2.684 | 0.502618 |
| 2.679 | 0.507853 |
| 2.677 | 0.513089 |
| 2.673 | 0.518325 |
| 2.653 | 0.52356 |
| 2.645 | 0.528796 |
| 2.599 | 0.534031 |
| 2.542 | 0.539267 |
| 2.517 | 0.544503 |
| 2.476 | 0.549738 |
| 2.459 | 0.554974 |
| 2.435 | 0.560209 |
| 2.435 | 0.565445 |
| 2.432 | 0.570681 |
| 2.411 | 0.575916 |
| 2.409 | 0.581152 |
| 2.375 | 0.586387 |
| 2.353 | 0.591623 |
| 2.341 | 0.596859 |
| 2.296 | 0.602094 |
| 2.278 | 0.60733 |
| 2.254 | 0.612565 |
| 2.239 | 0.617801 |
| 2.224 | 0.623037 |
| 2.198 | 0.628272 |
| 2.186 | 0.633508 |
| 2.167 | 0.638743 |
| 2.151 | 0.643979 |
| 2.123 | 0.649215 |
| 2.116 | 0.65445 |
| 2.077 | 0.659686 |
| 2.071 | 0.664921 |
| 1.992 | 0.670157 |
| 1.945 | 0.675393 |
| 1.945 | 0.680628 |
| 1.941 | 0.685864 |
| 1.904 | 0.691099 |
| 1.883 | 0.696335 |
| 1.866 | 0.701571 |
| 1.845 | 0.706806 |
| 1.715 | 0.712042 |
| 1.687 | 0.717277 |
| 1.675 | 0.722513 |
| 1.631 | 0.727749 |
| 1.605 | 0.732984 |
| 1.596 | 0.73822 |
| 1.587 | 0.743455 |
| 1.543 | 0.748691 |
| 1.462 | 0.753927 |
| 1.459 | 0.759162 |
| 1.447 | 0.764398 |
| 1.437 | 0.769634 |
| 1.416 | 0.774869 |
| 1.41 | 0.780105 |
| 1.391 | 0.78534 |
| 1.384 | 0.790576 |
| 1.379 | 0.795812 |
| 1.37 | 0.801047 |
| 1.363 | 0.806283 |
| 1.312 | 0.811518 |
| 1.285 | 0.816754 |
| 1.272 | 0.82199 |
| 1.257 | 0.827225 |
| 1.181 | 0.832461 |
| 1.161 | 0.837696 |
| 1.136 | 0.842932 |
| 1.113 | 0.848168 |
| 1.108 | 0.853403 |
| 1.093 | 0.858639 |
| 1.03 | 0.863874 |
| 1.015 | 0.86911 |
| 0.973 | 0.874346 |
| 0.968 | 0.879581 |
| 0.967 | 0.884817 |
| 0.908 | 0.890052 |
| 0.89 | 0.895288 |
| 0.889 | 0.900524 |
| 0.87 | 0.905759 |
| 0.845 | 0.910995 |
| 0.835 | 0.91623 |
| 0.79 | 0.921466 |
| 0.757 | 0.926702 |
| 0.746 | 0.931937 |
| 0.74 | 0.937173 |
| 0.632 | 0.942408 |
| 0.588 | 0.947644 |
| 0.57 | 0.95288 |
| 0.541 | 0.958115 |
| 0.493 | 0.963351 |
| 0.471 | 0.968586 |
| 0.424 | 0.973822 |
| 0.414 | 0.979058 |
| 0.344 | 0.984293 |
| 0.278 | 0.989529 |
| 0.206 | 0.994764 |
| 0.08 | 1 |

|  | ST 7F-Prevenar 13 (Day 84) |
| --- | --- |
| 86.422 | 0.00505051 |
| 80.771 | 0.010101 |
| 78.03 | 0.0151515 |
| 69.313 | 0.020202 |
| 64.655 | 0.0252525 |
| 49.239 | 0.030303 |
| 37.976 | 0.0353535 |
| 37.347 | 0.040404 |
| 34.34 | 0.0454545 |
| 27.355 | 0.0505051 |
| 26.855 | 0.0555556 |
| 26.377 | 0.0606061 |
| 24.452 | 0.0656566 |
| 23.316 | 0.0707071 |
| 21.705 | 0.0757576 |
| 19.606 | 0.0808081 |
| 19.177 | 0.0858586 |
| 14.853 | 0.0909091 |
| 14.725 | 0.0959596 |
| 14.204 | 0.10101 |
| 11.522 | 0.106061 |
| 10.598 | 0.111111 |
| 10.539 | 0.116162 |
| 10.516 | 0.121212 |
| 10.445 | 0.126263 |
| 10.324 | 0.131313 |
| 10.311 | 0.136364 |
| 10.301 | 0.141414 |
| 9.817 | 0.146465 |
| 9.752 | 0.151515 |
| 9.563 | 0.156566 |
| 9.469 | 0.161616 |
| 9.37 | 0.166667 |
| 9.257 | 0.171717 |
| 9.193 | 0.176768 |
| 9.158 | 0.181818 |
| 8.907 | 0.186869 |
| 8.78 | 0.191919 |
| 8.739 | 0.19697 |
| 8.711 | 0.20202 |
| 8.616 | 0.207071 |
| 8.439 | 0.212121 |
| 8.133 | 0.217172 |
| 7.991 | 0.222222 |
| 7.713 | 0.227273 |
| 7.677 | 0.232323 |
| 7.475 | 0.237374 |
| 6.917 | 0.242424 |
| 6.858 | 0.247475 |
| 6.842 | 0.252525 |
| 6.834 | 0.257576 |
| 6.772 | 0.262626 |
| 6.656 | 0.267677 |
| 6.539 | 0.272727 |
| 6.35 | 0.277778 |
| 6.341 | 0.282828 |
| 6.057 | 0.287879 |
| 5.993 | 0.292929 |
| 5.961 | 0.29798 |
| 5.877 | 0.30303 |
| 5.875 | 0.308081 |
| 5.667 | 0.313131 |
| 5.619 | 0.318182 |
| 5.496 | 0.323232 |
| 5.381 | 0.328283 |
| 5.139 | 0.333333 |
| 5.041 | 0.338384 |
| 5.031 | 0.343434 |
| 4.971 | 0.348485 |
| 4.971 | 0.353535 |
| 4.943 | 0.358586 |
| 4.934 | 0.363636 |
| 4.882 | 0.368687 |
| 4.834 | 0.373737 |
| 4.825 | 0.378788 |
| 4.758 | 0.383838 |
| 4.752 | 0.388889 |
| 4.692 | 0.393939 |
| 4.613 | 0.39899 |
| 4.576 | 0.40404 |
| 4.533 | 0.409091 |
| 4.449 | 0.414141 |
| 4.346 | 0.419192 |
| 4.336 | 0.424242 |
| 4.315 | 0.429293 |
| 4.299 | 0.434343 |
| 4.294 | 0.439394 |
| 4.279 | 0.444444 |
| 4.097 | 0.449495 |
| 4.046 | 0.454545 |
| 4.028 | 0.459596 |
| 4.01 | 0.464646 |
| 3.971 | 0.469697 |
| 3.924 | 0.474747 |
| 3.794 | 0.479798 |
| 3.754 | 0.484848 |
| 3.619 | 0.489899 |
| 3.586 | 0.494949 |
| 3.578 | 0.5 |
| 3.558 | 0.505051 |
| 3.537 | 0.510101 |
| 3.519 | 0.515152 |
| 3.509 | 0.520202 |
| 3.477 | 0.525253 |
| 3.447 | 0.530303 |
| 3.419 | 0.535354 |
| 3.377 | 0.540404 |
| 3.195 | 0.545455 |
| 3.142 | 0.550505 |
| 3.121 | 0.555556 |
| 3.112 | 0.560606 |
| 3.035 | 0.565657 |
| 3.03 | 0.570707 |
| 3.023 | 0.575758 |
| 2.999 | 0.580808 |
| 2.988 | 0.585859 |
| 2.938 | 0.590909 |
| 2.908 | 0.59596 |
| 2.892 | 0.60101 |
| 2.796 | 0.606061 |
| 2.771 | 0.611111 |
| 2.741 | 0.616162 |
| 2.736 | 0.621212 |
| 2.736 | 0.626263 |
| 2.672 | 0.631313 |
| 2.659 | 0.636364 |
| 2.652 | 0.641414 |
| 2.649 | 0.646465 |
| 2.647 | 0.651515 |
| 2.616 | 0.656566 |
| 2.604 | 0.661616 |
| 2.552 | 0.666667 |
| 2.549 | 0.671717 |
| 2.498 | 0.676768 |
| 2.463 | 0.681818 |
| 2.406 | 0.686869 |
| 2.338 | 0.691919 |
| 2.305 | 0.69697 |
| 2.277 | 0.70202 |
| 2.254 | 0.707071 |
| 2.223 | 0.712121 |
| 2.219 | 0.717172 |
| 2.215 | 0.722222 |
| 2.181 | 0.727273 |
| 2.144 | 0.732323 |
| 2.128 | 0.737374 |
| 2.03 | 0.742424 |
| 2.018 | 0.747475 |
| 2.013 | 0.752525 |
| 2 | 0.757576 |
| 1.994 | 0.762626 |
| 1.957 | 0.767677 |
| 1.922 | 0.772727 |
| 1.888 | 0.777778 |
| 1.802 | 0.782828 |
| 1.795 | 0.787879 |
| 1.775 | 0.792929 |
| 1.763 | 0.79798 |
| 1.71 | 0.80303 |
| 1.697 | 0.808081 |
| 1.654 | 0.813131 |
| 1.645 | 0.818182 |
| 1.548 | 0.823232 |
| 1.538 | 0.828283 |
| 1.476 | 0.833333 |
| 1.46 | 0.838384 |
| 1.429 | 0.843434 |
| 1.424 | 0.848485 |
| 1.403 | 0.853535 |
| 1.391 | 0.858586 |
| 1.286 | 0.863636 |
| 1.195 | 0.868687 |
| 1.164 | 0.873737 |
| 1.154 | 0.878788 |
| 1.148 | 0.883838 |
| 1.101 | 0.888889 |
| 1.064 | 0.893939 |
| 1.059 | 0.89899 |
| 1.043 | 0.90404 |
| 1.001 | 0.909091 |
| 0.995 | 0.914141 |
| 0.962 | 0.919192 |
| 0.886 | 0.924242 |
| 0.856 | 0.929293 |
| 0.839 | 0.934343 |
| 0.83 | 0.939394 |
| 0.773 | 0.944444 |
| 0.767 | 0.949495 |
| 0.756 | 0.954545 |
| 0.746 | 0.959596 |
| 0.711 | 0.964646 |
| 0.49 | 0.969697 |
| 0.44 | 0.974747 |
| 0.425 | 0.979798 |
| 0.358 | 0.984848 |
| 0.347 | 0.989899 |
| 0.204 | 0.994949 |
| 0.19 | 1 |

|  | ST 7F-BE-PCV14 (Day 300) |
| --- | --- |
| 55.392 | 0.00531915 |
| 43.357 | 0.0106383 |
| 40.511 | 0.0159574 |
| 22.223 | 0.0212766 |
| 21.679 | 0.0265957 |
| 21.068 | 0.0319149 |
| 19.923 | 0.037234 |
| 19.895 | 0.0425532 |
| 19.759 | 0.0478723 |
| 18.147 | 0.0531915 |
| 15.909 | 0.0585106 |
| 14.792 | 0.0638298 |
| 14.39 | 0.0691489 |
| 13.853 | 0.0744681 |
| 13.237 | 0.0797872 |
| 12.633 | 0.0851064 |
| 11.827 | 0.0904255 |
| 10.081 | 0.0957447 |
| 10.062 | 0.101064 |
| 9.66 | 0.106383 |
| 9.627 | 0.111702 |
| 9.558 | 0.117021 |
| 9.455 | 0.12234 |
| 9.238 | 0.12766 |
| 9.224 | 0.132979 |
| 8.88 | 0.138298 |
| 8.732 | 0.143617 |
| 8.488 | 0.148936 |
| 8.398 | 0.154255 |
| 7.946 | 0.159574 |
| 7.88 | 0.164894 |
| 7.74 | 0.170213 |
| 7.648 | 0.175532 |
| 7.643 | 0.180851 |
| 7.555 | 0.18617 |
| 7.179 | 0.191489 |
| 7.17 | 0.196809 |
| 7.145 | 0.202128 |
| 7.14 | 0.207447 |
| 7.066 | 0.212766 |
| 7.028 | 0.218085 |
| 6.93 | 0.223404 |
| 6.806 | 0.228723 |
| 6.801 | 0.234043 |
| 6.778 | 0.239362 |
| 6.725 | 0.244681 |
| 6.605 | 0.25 |
| 6.533 | 0.255319 |
| 6.414 | 0.260638 |
| 6.399 | 0.265957 |
| 5.75 | 0.271277 |
| 5.729 | 0.276596 |
| 5.689 | 0.281915 |
| 5.598 | 0.287234 |
| 5.127 | 0.292553 |
| 5.113 | 0.297872 |
| 4.845 | 0.303191 |
| 4.776 | 0.308511 |
| 4.576 | 0.31383 |
| 4.495 | 0.319149 |
| 4.483 | 0.324468 |
| 4.447 | 0.329787 |
| 4.298 | 0.335106 |
| 4.293 | 0.340426 |
| 4.114 | 0.345745 |
| 4.081 | 0.351064 |
| 3.999 | 0.356383 |
| 3.939 | 0.361702 |
| 3.896 | 0.367021 |
| 3.893 | 0.37234 |
| 3.693 | 0.37766 |
| 3.678 | 0.382979 |
| 3.658 | 0.388298 |
| 3.446 | 0.393617 |
| 3.39 | 0.398936 |
| 3.388 | 0.404255 |
| 3.37 | 0.409574 |
| 3.252 | 0.414894 |
| 3.192 | 0.420213 |
| 3.104 | 0.425532 |
| 3.09 | 0.430851 |
| 3.076 | 0.43617 |
| 3.026 | 0.441489 |
| 2.973 | 0.446809 |
| 2.875 | 0.452128 |
| 2.806 | 0.457447 |
| 2.705 | 0.462766 |
| 2.674 | 0.468085 |
| 2.67 | 0.473404 |
| 2.651 | 0.478723 |
| 2.646 | 0.484043 |
| 2.645 | 0.489362 |
| 2.62 | 0.494681 |
| 2.585 | 0.5 |
| 2.519 | 0.505319 |
| 2.494 | 0.510638 |
| 2.468 | 0.515957 |
| 2.4 | 0.521277 |
| 2.399 | 0.526596 |
| 2.368 | 0.531915 |
| 2.344 | 0.537234 |
| 2.312 | 0.542553 |
| 2.296 | 0.547872 |
| 2.264 | 0.553191 |
| 2.218 | 0.558511 |
| 2.216 | 0.56383 |
| 2.188 | 0.569149 |
| 2.182 | 0.574468 |
| 2.173 | 0.579787 |
| 2.17 | 0.585106 |
| 2.129 | 0.590426 |
| 2.094 | 0.595745 |
| 2.046 | 0.601064 |
| 2.04 | 0.606383 |
| 2.035 | 0.611702 |
| 2.003 | 0.617021 |
| 1.98 | 0.62234 |
| 1.972 | 0.62766 |
| 1.83 | 0.632979 |
| 1.786 | 0.638298 |
| 1.727 | 0.643617 |
| 1.718 | 0.648936 |
| 1.692 | 0.654255 |
| 1.67 | 0.659574 |
| 1.667 | 0.664894 |
| 1.658 | 0.670213 |
| 1.641 | 0.675532 |
| 1.565 | 0.680851 |
| 1.555 | 0.68617 |
| 1.497 | 0.691489 |
| 1.481 | 0.696809 |
| 1.461 | 0.702128 |
| 1.431 | 0.707447 |
| 1.423 | 0.712766 |
| 1.401 | 0.718085 |
| 1.396 | 0.723404 |
| 1.372 | 0.728723 |
| 1.348 | 0.734043 |
| 1.317 | 0.739362 |
| 1.265 | 0.744681 |
| 1.196 | 0.75 |
| 1.184 | 0.755319 |
| 1.05 | 0.760638 |
| 1.032 | 0.765957 |
| 1.027 | 0.771277 |
| 1.008 | 0.776596 |
| 0.977 | 0.781915 |
| 0.949 | 0.787234 |
| 0.887 | 0.792553 |
| 0.886 | 0.797872 |
| 0.848 | 0.803191 |
| 0.826 | 0.808511 |
| 0.765 | 0.81383 |
| 0.739 | 0.819149 |
| 0.736 | 0.824468 |
| 0.715 | 0.829787 |
| 0.714 | 0.835106 |
| 0.713 | 0.840426 |
| 0.697 | 0.845745 |
| 0.696 | 0.851064 |
| 0.69 | 0.856383 |
| 0.662 | 0.861702 |
| 0.652 | 0.867021 |
| 0.628 | 0.87234 |
| 0.605 | 0.87766 |
| 0.581 | 0.882979 |
| 0.569 | 0.888298 |
| 0.54 | 0.893617 |
| 0.534 | 0.898936 |
| 0.519 | 0.904255 |
| 0.514 | 0.909574 |
| 0.503 | 0.914894 |
| 0.477 | 0.920213 |
| 0.462 | 0.925532 |
| 0.458 | 0.930851 |
| 0.453 | 0.93617 |
| 0.426 | 0.941489 |
| 0.419 | 0.946809 |
| 0.378 | 0.952128 |
| 0.363 | 0.957447 |
| 0.346 | 0.962766 |
| 0.337 | 0.968085 |
| 0.301 | 0.973404 |
| 0.208 | 0.978723 |
| 0.136 | 0.984043 |
| 0.118 | 0.989362 |
| 0.111 | 0.994681 |
| 0.033 | 1 |

|  | ST 7F-Prevenar 13 (Day 300) |
| --- | --- |
| 39.177 | 0.00526316 |
| 28.573 | 0.0105263 |
| 24.336 | 0.0157895 |
| 20.304 | 0.0210526 |
| 19.278 | 0.0263158 |
| 18.822 | 0.0315789 |
| 17.621 | 0.0368421 |
| 16.374 | 0.0421053 |
| 15.053 | 0.0473684 |
| 13.91 | 0.0526316 |
| 13.721 | 0.0578947 |
| 13.084 | 0.0631579 |
| 12.656 | 0.0684211 |
| 12.444 | 0.0736842 |
| 11.841 | 0.0789474 |
| 11.603 | 0.0842105 |
| 11.302 | 0.0894737 |
| 11.046 | 0.0947368 |
| 11.001 | 0.1 |
| 10.249 | 0.105263 |
| 9.957 | 0.110526 |
| 9.925 | 0.115789 |
| 9.813 | 0.121053 |
| 9.502 | 0.126316 |
| 9.45 | 0.131579 |
| 8.958 | 0.136842 |
| 8.469 | 0.142105 |
| 7.735 | 0.147368 |
| 7.536 | 0.152632 |
| 7.431 | 0.157895 |
| 7.261 | 0.163158 |
| 7.231 | 0.168421 |
| 7.226 | 0.173684 |
| 7.201 | 0.178947 |
| 7.131 | 0.184211 |
| 7.075 | 0.189474 |
| 6.968 | 0.194737 |
| 6.459 | 0.2 |
| 6.452 | 0.205263 |
| 6.382 | 0.210526 |
| 6.167 | 0.215789 |
| 6.156 | 0.221053 |
| 5.887 | 0.226316 |
| 5.841 | 0.231579 |
| 5.774 | 0.236842 |
| 5.755 | 0.242105 |
| 5.659 | 0.247368 |
| 5.552 | 0.252632 |
| 5.359 | 0.257895 |
| 5.22 | 0.263158 |
| 5.206 | 0.268421 |
| 5.171 | 0.273684 |
| 5.112 | 0.278947 |
| 5.099 | 0.284211 |
| 4.864 | 0.289474 |
| 4.788 | 0.294737 |
| 4.761 | 0.3 |
| 4.728 | 0.305263 |
| 4.717 | 0.310526 |
| 4.696 | 0.315789 |
| 4.361 | 0.321053 |
| 4.28 | 0.326316 |
| 4.169 | 0.331579 |
| 4.147 | 0.336842 |
| 4.142 | 0.342105 |
| 4.058 | 0.347368 |
| 4.026 | 0.352632 |
| 3.896 | 0.357895 |
| 3.81 | 0.363158 |
| 3.753 | 0.368421 |
| 3.719 | 0.373684 |
| 3.71 | 0.378947 |
| 3.403 | 0.384211 |
| 3.391 | 0.389474 |
| 3.361 | 0.394737 |
| 3.352 | 0.4 |
| 3.233 | 0.405263 |
| 3.206 | 0.410526 |
| 3.056 | 0.415789 |
| 3.052 | 0.421053 |
| 3.025 | 0.426316 |
| 2.971 | 0.431579 |
| 2.948 | 0.436842 |
| 2.929 | 0.442105 |
| 2.91 | 0.447368 |
| 2.851 | 0.452632 |
| 2.847 | 0.457895 |
| 2.835 | 0.463158 |
| 2.768 | 0.468421 |
| 2.707 | 0.473684 |
| 2.702 | 0.478947 |
| 2.701 | 0.484211 |
| 2.663 | 0.489474 |
| 2.653 | 0.494737 |
| 2.542 | 0.5 |
| 2.528 | 0.505263 |
| 2.489 | 0.510526 |
| 2.472 | 0.515789 |
| 2.42 | 0.521053 |
| 2.41 | 0.526316 |
| 2.365 | 0.531579 |
| 2.364 | 0.536842 |
| 2.361 | 0.542105 |
| 2.359 | 0.547368 |
| 2.32 | 0.552632 |
| 2.29 | 0.557895 |
| 2.281 | 0.563158 |
| 2.268 | 0.568421 |
| 2.2 | 0.573684 |
| 2.133 | 0.578947 |
| 2.042 | 0.584211 |
| 2.041 | 0.589474 |
| 2.019 | 0.594737 |
| 1.991 | 0.6 |
| 1.983 | 0.605263 |
| 1.965 | 0.610526 |
| 1.954 | 0.615789 |
| 1.904 | 0.621053 |
| 1.903 | 0.626316 |
| 1.894 | 0.631579 |
| 1.879 | 0.636842 |
| 1.871 | 0.642105 |
| 1.855 | 0.647368 |
| 1.841 | 0.652632 |
| 1.84 | 0.657895 |
| 1.816 | 0.663158 |
| 1.79 | 0.668421 |
| 1.691 | 0.673684 |
| 1.671 | 0.678947 |
| 1.608 | 0.684211 |
| 1.598 | 0.689474 |
| 1.582 | 0.694737 |
| 1.578 | 0.7 |
| 1.573 | 0.705263 |
| 1.561 | 0.710526 |
| 1.554 | 0.715789 |
| 1.534 | 0.721053 |
| 1.532 | 0.726316 |
| 1.506 | 0.731579 |
| 1.488 | 0.736842 |
| 1.473 | 0.742105 |
| 1.44 | 0.747368 |
| 1.42 | 0.752632 |
| 1.406 | 0.757895 |
| 1.398 | 0.763158 |
| 1.366 | 0.768421 |
| 1.345 | 0.773684 |
| 1.34 | 0.778947 |
| 1.33 | 0.784211 |
| 1.254 | 0.789474 |
| 1.206 | 0.794737 |
| 1.202 | 0.8 |
| 1.143 | 0.805263 |
| 1.122 | 0.810526 |
| 1.11 | 0.815789 |
| 1.095 | 0.821053 |
| 1.092 | 0.826316 |
| 1.089 | 0.831579 |
| 1.069 | 0.836842 |
| 1.063 | 0.842105 |
| 1.057 | 0.847368 |
| 1.055 | 0.852632 |
| 1.044 | 0.857895 |
| 1.04 | 0.863158 |
| 1.035 | 0.868421 |
| 1.013 | 0.873684 |
| 1.011 | 0.878947 |
| 1.005 | 0.884211 |
| 0.969 | 0.889474 |
| 0.962 | 0.894737 |
| 0.929 | 0.9 |
| 0.815 | 0.905263 |
| 0.785 | 0.910526 |
| 0.726 | 0.915789 |
| 0.703 | 0.921053 |
| 0.696 | 0.926316 |
| 0.619 | 0.931579 |
| 0.545 | 0.936842 |
| 0.528 | 0.942105 |
| 0.525 | 0.947368 |
| 0.485 | 0.952632 |
| 0.472 | 0.957895 |
| 0.44 | 0.963158 |
| 0.345 | 0.968421 |
| 0.321 | 0.973684 |
| 0.204 | 0.978947 |
| 0.195 | 0.984211 |
| 0.179 | 0.989474 |
| 0.178 | 0.994737 |
| 0.12 | 1 |

BECT056 - RCDC_Serotype 9V

|  | ST 9V-BE-PCV14 (Day 84) |
| --- | --- |
| 61.332 | 0.0052356 |
| 60.227 | 0.0104712 |
| 43.124 | 0.0157068 |
| 35.766 | 0.0209424 |
| 33.401 | 0.026178 |
| 32.313 | 0.0314136 |
| 28.292 | 0.0366492 |
| 27.579 | 0.0418848 |
| 21.712 | 0.0471204 |
| 21.249 | 0.052356 |
| 17.942 | 0.0575916 |
| 17.774 | 0.0628272 |
| 16.417 | 0.0680628 |
| 16.406 | 0.0732984 |
| 16.221 | 0.078534 |
| 15.471 | 0.0837696 |
| 12.884 | 0.0890052 |
| 12.668 | 0.0942408 |
| 11.924 | 0.0994764 |
| 11.868 | 0.104712 |
| 11.291 | 0.109948 |
| 10.375 | 0.115183 |
| 10.107 | 0.120419 |
| 10.073 | 0.125654 |
| 8.95 | 0.13089 |
| 8.905 | 0.136126 |
| 8.886 | 0.141361 |
| 7.975 | 0.146597 |
| 7.798 | 0.151832 |
| 7.696 | 0.157068 |
| 7.504 | 0.162304 |
| 7.484 | 0.167539 |
| 7.364 | 0.172775 |
| 6.996 | 0.17801 |
| 6.611 | 0.183246 |
| 6.339 | 0.188482 |
| 6.215 | 0.193717 |
| 5.957 | 0.198953 |
| 5.933 | 0.204188 |
| 5.769 | 0.209424 |
| 5.278 | 0.21466 |
| 5.268 | 0.219895 |
| 5.211 | 0.225131 |
| 5.198 | 0.230366 |
| 5.064 | 0.235602 |
| 5.005 | 0.240838 |
| 4.993 | 0.246073 |
| 4.629 | 0.251309 |
| 4.603 | 0.256545 |
| 4.564 | 0.26178 |
| 4.429 | 0.267016 |
| 4.363 | 0.272251 |
| 4.242 | 0.277487 |
| 4.224 | 0.282723 |
| 4.178 | 0.287958 |
| 4.109 | 0.293194 |
| 4.065 | 0.298429 |
| 4.055 | 0.303665 |
| 4.026 | 0.308901 |
| 4.004 | 0.314136 |
| 3.978 | 0.319372 |
| 3.945 | 0.324607 |
| 3.929 | 0.329843 |
| 3.899 | 0.335079 |
| 3.836 | 0.340314 |
| 3.713 | 0.34555 |
| 3.708 | 0.350785 |
| 3.648 | 0.356021 |
| 3.646 | 0.361257 |
| 3.612 | 0.366492 |
| 3.61 | 0.371728 |
| 3.566 | 0.376963 |
| 3.524 | 0.382199 |
| 3.516 | 0.387435 |
| 3.476 | 0.39267 |
| 3.405 | 0.397906 |
| 3.401 | 0.403141 |
| 3.37 | 0.408377 |
| 3.351 | 0.413613 |
| 3.346 | 0.418848 |
| 3.123 | 0.424084 |
| 2.969 | 0.429319 |
| 2.908 | 0.434555 |
| 2.907 | 0.439791 |
| 2.901 | 0.445026 |
| 2.888 | 0.450262 |
| 2.831 | 0.455497 |
| 2.796 | 0.460733 |
| 2.793 | 0.465969 |
| 2.777 | 0.471204 |
| 2.776 | 0.47644 |
| 2.765 | 0.481675 |
| 2.757 | 0.486911 |
| 2.702 | 0.492147 |
| 2.695 | 0.497382 |
| 2.661 | 0.502618 |
| 2.652 | 0.507853 |
| 2.611 | 0.513089 |
| 2.606 | 0.518325 |
| 2.563 | 0.52356 |
| 2.553 | 0.528796 |
| 2.54 | 0.534031 |
| 2.522 | 0.539267 |
| 2.49 | 0.544503 |
| 2.459 | 0.549738 |
| 2.409 | 0.554974 |
| 2.389 | 0.560209 |
| 2.376 | 0.565445 |
| 2.366 | 0.570681 |
| 2.3 | 0.575916 |
| 2.293 | 0.581152 |
| 2.276 | 0.586387 |
| 2.23 | 0.591623 |
| 2.214 | 0.596859 |
| 2.205 | 0.602094 |
| 2.153 | 0.60733 |
| 2.05 | 0.612565 |
| 2.045 | 0.617801 |
| 1.986 | 0.623037 |
| 1.927 | 0.628272 |
| 1.904 | 0.633508 |
| 1.861 | 0.638743 |
| 1.722 | 0.643979 |
| 1.692 | 0.649215 |
| 1.69 | 0.65445 |
| 1.643 | 0.659686 |
| 1.557 | 0.664921 |
| 1.552 | 0.670157 |
| 1.523 | 0.675393 |
| 1.52 | 0.680628 |
| 1.497 | 0.685864 |
| 1.495 | 0.691099 |
| 1.401 | 0.696335 |
| 1.384 | 0.701571 |
| 1.366 | 0.706806 |
| 1.339 | 0.712042 |
| 1.294 | 0.717277 |
| 1.289 | 0.722513 |
| 1.27 | 0.727749 |
| 1.258 | 0.732984 |
| 1.25 | 0.73822 |
| 1.212 | 0.743455 |
| 1.197 | 0.748691 |
| 1.177 | 0.753927 |
| 1.148 | 0.759162 |
| 1.14 | 0.764398 |
| 1.095 | 0.769634 |
| 1.051 | 0.774869 |
| 0.969 | 0.780105 |
| 0.926 | 0.78534 |
| 0.912 | 0.790576 |
| 0.912 | 0.795812 |
| 0.881 | 0.801047 |
| 0.872 | 0.806283 |
| 0.836 | 0.811518 |
| 0.833 | 0.816754 |
| 0.825 | 0.82199 |
| 0.793 | 0.827225 |
| 0.767 | 0.832461 |
| 0.758 | 0.837696 |
| 0.753 | 0.842932 |
| 0.752 | 0.848168 |
| 0.751 | 0.853403 |
| 0.715 | 0.858639 |
| 0.713 | 0.863874 |
| 0.677 | 0.86911 |
| 0.671 | 0.874346 |
| 0.633 | 0.879581 |
| 0.626 | 0.884817 |
| 0.612 | 0.890052 |
| 0.588 | 0.895288 |
| 0.583 | 0.900524 |
| 0.578 | 0.905759 |
| 0.575 | 0.910995 |
| 0.52 | 0.91623 |
| 0.451 | 0.921466 |
| 0.43 | 0.926702 |
| 0.419 | 0.931937 |
| 0.416 | 0.937173 |
| 0.414 | 0.942408 |
| 0.412 | 0.947644 |
| 0.408 | 0.95288 |
| 0.407 | 0.958115 |
| 0.368 | 0.963351 |
| 0.328 | 0.968586 |
| 0.284 | 0.973822 |
| 0.233 | 0.979058 |
| 0.205 | 0.984293 |
| 0.202 | 0.989529 |
| 0.107 | 0.994764 |
| 0.088 | 1 |

|  | ST 9V-Prevenar 13 (Day 84) |
| --- | --- |
| 267.11 | 0.00505051 |
| 93.064 | 0.010101 |
| 92.654 | 0.0151515 |
| 47.557 | 0.020202 |
| 41.972 | 0.0252525 |
| 39.315 | 0.030303 |
| 36.131 | 0.0353535 |
| 35.836 | 0.040404 |
| 32.351 | 0.0454545 |
| 27.227 | 0.0505051 |
| 26.254 | 0.0555556 |
| 24.907 | 0.0606061 |
| 24.597 | 0.0656566 |
| 23.481 | 0.0707071 |
| 22.895 | 0.0757576 |
| 19.216 | 0.0808081 |
| 18.262 | 0.0858586 |
| 17.874 | 0.0909091 |
| 17.455 | 0.0959596 |
| 15.849 | 0.10101 |
| 15.429 | 0.106061 |
| 15.034 | 0.111111 |
| 14.675 | 0.116162 |
| 13.269 | 0.121212 |
| 12.258 | 0.126263 |
| 11.412 | 0.131313 |
| 11.309 | 0.136364 |
| 10.354 | 0.141414 |
| 10.34 | 0.146465 |
| 9.971 | 0.151515 |
| 9.309 | 0.156566 |
| 9.179 | 0.161616 |
| 8.52 | 0.166667 |
| 8.329 | 0.171717 |
| 7.942 | 0.176768 |
| 7.753 | 0.181818 |
| 7.682 | 0.186869 |
| 7.398 | 0.191919 |
| 7.226 | 0.19697 |
| 7.066 | 0.20202 |
| 6.984 | 0.207071 |
| 6.846 | 0.212121 |
| 6.765 | 0.217172 |
| 6.701 | 0.222222 |
| 6.336 | 0.227273 |
| 6.109 | 0.232323 |
| 6.08 | 0.237374 |
| 5.856 | 0.242424 |
| 5.618 | 0.247475 |
| 5.413 | 0.252525 |
| 5.17 | 0.257576 |
| 5.073 | 0.262626 |
| 4.941 | 0.267677 |
| 4.833 | 0.272727 |
| 4.824 | 0.277778 |
| 4.822 | 0.282828 |
| 4.658 | 0.287879 |
| 4.605 | 0.292929 |
| 4.57 | 0.29798 |
| 4.439 | 0.30303 |
| 4.338 | 0.308081 |
| 4.313 | 0.313131 |
| 4.169 | 0.318182 |
| 4.058 | 0.323232 |
| 4.05 | 0.328283 |
| 3.995 | 0.333333 |
| 3.91 | 0.338384 |
| 3.871 | 0.343434 |
| 3.723 | 0.348485 |
| 3.66 | 0.353535 |
| 3.5 | 0.358586 |
| 3.477 | 0.363636 |
| 3.462 | 0.368687 |
| 3.413 | 0.373737 |
| 3.364 | 0.378788 |
| 3.297 | 0.383838 |
| 3.249 | 0.388889 |
| 3.205 | 0.393939 |
| 3.122 | 0.39899 |
| 3.051 | 0.40404 |
| 2.985 | 0.409091 |
| 2.981 | 0.414141 |
| 2.967 | 0.419192 |
| 2.96 | 0.424242 |
| 2.858 | 0.429293 |
| 2.828 | 0.434343 |
| 2.815 | 0.439394 |
| 2.814 | 0.444444 |
| 2.793 | 0.449495 |
| 2.765 | 0.454545 |
| 2.759 | 0.459596 |
| 2.748 | 0.464646 |
| 2.723 | 0.469697 |
| 2.675 | 0.474747 |
| 2.669 | 0.479798 |
| 2.619 | 0.484848 |
| 2.617 | 0.489899 |
| 2.581 | 0.494949 |
| 2.547 | 0.5 |
| 2.486 | 0.505051 |
| 2.45 | 0.510101 |
| 2.401 | 0.515152 |
| 2.324 | 0.520202 |
| 2.306 | 0.525253 |
| 2.289 | 0.530303 |
| 2.239 | 0.535354 |
| 2.21 | 0.540404 |
| 2.197 | 0.545455 |
| 2.166 | 0.550505 |
| 2.146 | 0.555556 |
| 2.134 | 0.560606 |
| 2.115 | 0.565657 |
| 2.088 | 0.570707 |
| 2.062 | 0.575758 |
| 2.058 | 0.580808 |
| 2.023 | 0.585859 |
| 2.02 | 0.590909 |
| 1.957 | 0.59596 |
| 1.929 | 0.60101 |
| 1.919 | 0.606061 |
| 1.896 | 0.611111 |
| 1.89 | 0.616162 |
| 1.855 | 0.621212 |
| 1.844 | 0.626263 |
| 1.809 | 0.631313 |
| 1.806 | 0.636364 |
| 1.791 | 0.641414 |
| 1.783 | 0.646465 |
| 1.774 | 0.651515 |
| 1.728 | 0.656566 |
| 1.709 | 0.661616 |
| 1.69 | 0.666667 |
| 1.664 | 0.671717 |
| 1.659 | 0.676768 |
| 1.645 | 0.681818 |
| 1.625 | 0.686869 |
| 1.612 | 0.691919 |
| 1.608 | 0.69697 |
| 1.584 | 0.70202 |
| 1.554 | 0.707071 |
| 1.507 | 0.712121 |
| 1.497 | 0.717172 |
| 1.418 | 0.722222 |
| 1.315 | 0.727273 |
| 1.306 | 0.732323 |
| 1.297 | 0.737374 |
| 1.292 | 0.742424 |
| 1.284 | 0.747475 |
| 1.264 | 0.752525 |
| 1.235 | 0.757576 |
| 1.199 | 0.762626 |
| 1.16 | 0.767677 |
| 1.143 | 0.772727 |
| 1.12 | 0.777778 |
| 1.106 | 0.782828 |
| 1.104 | 0.787879 |
| 1.062 | 0.792929 |
| 1.042 | 0.79798 |
| 1.027 | 0.80303 |
| 1.027 | 0.808081 |
| 1.02 | 0.813131 |
| 1.014 | 0.818182 |
| 1.002 | 0.823232 |
| 0.98 | 0.828283 |
| 0.961 | 0.833333 |
| 0.889 | 0.838384 |
| 0.88 | 0.843434 |
| 0.858 | 0.848485 |
| 0.845 | 0.853535 |
| 0.843 | 0.858586 |
| 0.835 | 0.863636 |
| 0.833 | 0.868687 |
| 0.797 | 0.873737 |
| 0.792 | 0.878788 |
| 0.789 | 0.883838 |
| 0.746 | 0.888889 |
| 0.72 | 0.893939 |
| 0.713 | 0.89899 |
| 0.71 | 0.90404 |
| 0.653 | 0.909091 |
| 0.653 | 0.914141 |
| 0.62 | 0.919192 |
| 0.618 | 0.924242 |
| 0.615 | 0.929293 |
| 0.598 | 0.934343 |
| 0.585 | 0.939394 |
| 0.561 | 0.944444 |
| 0.486 | 0.949495 |
| 0.474 | 0.954545 |
| 0.39 | 0.959596 |
| 0.37 | 0.964646 |
| 0.357 | 0.969697 |
| 0.21 | 0.974747 |
| 0.185 | 0.979798 |
| 0.162 | 0.984848 |
| 0.153 | 0.989899 |
| 0.15 | 0.994949 |
| 0.126 | 1 |

|  | ST 9V-BE-PCV14 (Day 300) |
| --- | --- |
| 65.799 | 0.00531915 |
| 52.706 | 0.0106383 |
| 43.483 | 0.0159574 |
| 22.449 | 0.0212766 |
| 21.885 | 0.0265957 |
| 18.01 | 0.0319149 |
| 17.473 | 0.037234 |
| 16.726 | 0.0425532 |
| 15.977 | 0.0478723 |
| 13.671 | 0.0531915 |
| 13.63 | 0.0585106 |
| 10.944 | 0.0638298 |
| 10.683 | 0.0691489 |
| 10.581 | 0.0744681 |
| 10.546 | 0.0797872 |
| 10.292 | 0.0851064 |
| 10.218 | 0.0904255 |
| 10.192 | 0.0957447 |
| 10.107 | 0.101064 |
| 10.105 | 0.106383 |
| 10.038 | 0.111702 |
| 9.504 | 0.117021 |
| 9.464 | 0.12234 |
| 9.329 | 0.12766 |
| 9.092 | 0.132979 |
| 9.057 | 0.138298 |
| 9.004 | 0.143617 |
| 8.815 | 0.148936 |
| 8.809 | 0.154255 |
| 8.502 | 0.159574 |
| 8.48 | 0.164894 |
| 8.434 | 0.170213 |
| 8.397 | 0.175532 |
| 8.334 | 0.180851 |
| 8.309 | 0.18617 |
| 7.994 | 0.191489 |
| 7.945 | 0.196809 |
| 7.943 | 0.202128 |
| 7.66 | 0.207447 |
| 7.421 | 0.212766 |
| 7.292 | 0.218085 |
| 7.282 | 0.223404 |
| 7.181 | 0.228723 |
| 6.884 | 0.234043 |
| 6.86 | 0.239362 |
| 6.689 | 0.244681 |
| 6.672 | 0.25 |
| 6.622 | 0.255319 |
| 6.538 | 0.260638 |
| 6.342 | 0.265957 |
| 6.278 | 0.271277 |
| 6.27 | 0.276596 |
| 5.986 | 0.281915 |
| 5.981 | 0.287234 |
| 5.949 | 0.292553 |
| 5.582 | 0.297872 |
| 5.529 | 0.303191 |
| 5.501 | 0.308511 |
| 5.331 | 0.31383 |
| 5.212 | 0.319149 |
| 5.195 | 0.324468 |
| 5.007 | 0.329787 |
| 4.974 | 0.335106 |
| 4.762 | 0.340426 |
| 4.721 | 0.345745 |
| 4.715 | 0.351064 |
| 4.654 | 0.356383 |
| 4.623 | 0.361702 |
| 4.613 | 0.367021 |
| 4.57 | 0.37234 |
| 4.488 | 0.37766 |
| 4.48 | 0.382979 |
| 4.184 | 0.388298 |
| 4.136 | 0.393617 |
| 4.087 | 0.398936 |
| 4.003 | 0.404255 |
| 3.993 | 0.409574 |
| 3.989 | 0.414894 |
| 3.901 | 0.420213 |
| 3.827 | 0.425532 |
| 3.815 | 0.430851 |
| 3.809 | 0.43617 |
| 3.691 | 0.441489 |
| 3.63 | 0.446809 |
| 3.576 | 0.452128 |
| 3.524 | 0.457447 |
| 3.498 | 0.462766 |
| 3.387 | 0.468085 |
| 3.314 | 0.473404 |
| 3.226 | 0.478723 |
| 3.18 | 0.484043 |
| 3.17 | 0.489362 |
| 3.12 | 0.494681 |
| 3.076 | 0.5 |
| 3.064 | 0.505319 |
| 3.03 | 0.510638 |
| 2.997 | 0.515957 |
| 2.991 | 0.521277 |
| 2.947 | 0.526596 |
| 2.89 | 0.531915 |
| 2.873 | 0.537234 |
| 2.872 | 0.542553 |
| 2.871 | 0.547872 |
| 2.823 | 0.553191 |
| 2.718 | 0.558511 |
| 2.699 | 0.56383 |
| 2.617 | 0.569149 |
| 2.566 | 0.574468 |
| 2.488 | 0.579787 |
| 2.455 | 0.585106 |
| 2.35 | 0.590426 |
| 2.313 | 0.595745 |
| 2.25 | 0.601064 |
| 2.231 | 0.606383 |
| 2.205 | 0.611702 |
| 2.137 | 0.617021 |
| 2.057 | 0.62234 |
| 2.033 | 0.62766 |
| 1.976 | 0.632979 |
| 1.967 | 0.638298 |
| 1.915 | 0.643617 |
| 1.876 | 0.648936 |
| 1.873 | 0.654255 |
| 1.821 | 0.659574 |
| 1.821 | 0.664894 |
| 1.781 | 0.670213 |
| 1.744 | 0.675532 |
| 1.741 | 0.680851 |
| 1.716 | 0.68617 |
| 1.7 | 0.691489 |
| 1.699 | 0.696809 |
| 1.667 | 0.702128 |
| 1.661 | 0.707447 |
| 1.658 | 0.712766 |
| 1.655 | 0.718085 |
| 1.642 | 0.723404 |
| 1.64 | 0.728723 |
| 1.601 | 0.734043 |
| 1.519 | 0.739362 |
| 1.487 | 0.744681 |
| 1.48 | 0.75 |
| 1.449 | 0.755319 |
| 1.409 | 0.760638 |
| 1.408 | 0.765957 |
| 1.392 | 0.771277 |
| 1.288 | 0.776596 |
| 1.273 | 0.781915 |
| 1.23 | 0.787234 |
| 1.19 | 0.792553 |
| 1.146 | 0.797872 |
| 1.115 | 0.803191 |
| 1.021 | 0.808511 |
| 0.968 | 0.81383 |
| 0.953 | 0.819149 |
| 0.813 | 0.824468 |
| 0.662 | 0.829787 |
| 0.624 | 0.835106 |
| 0.598 | 0.840426 |
| 0.563 | 0.845745 |
| 0.558 | 0.851064 |
| 0.553 | 0.856383 |
| 0.552 | 0.861702 |
| 0.531 | 0.867021 |
| 0.515 | 0.87234 |
| 0.51 | 0.87766 |
| 0.507 | 0.882979 |
| 0.496 | 0.888298 |
| 0.486 | 0.893617 |
| 0.47 | 0.898936 |
| 0.454 | 0.904255 |
| 0.443 | 0.909574 |
| 0.431 | 0.914894 |
| 0.41 | 0.920213 |
| 0.4 | 0.925532 |
| 0.384 | 0.930851 |
| 0.374 | 0.93617 |
| 0.37 | 0.941489 |
| 0.369 | 0.946809 |
| 0.337 | 0.952128 |
| 0.322 | 0.957447 |
| 0.255 | 0.962766 |
| 0.227 | 0.968085 |
| 0.176 | 0.973404 |
| 0.15 | 0.978723 |
| 0.149 | 0.984043 |
| 0.132 | 0.989362 |
| 0.119 | 0.994681 |
| 0.077 | 1 |

|  | ST 9V-Prevenar 13 (Day 300) |
| --- | --- |
| 35.463 | 0.00526316 |
| 30.514 | 0.0105263 |
| 27.38 | 0.0157895 |
| 26.856 | 0.0210526 |
| 26.191 | 0.0263158 |
| 24.288 | 0.0315789 |
| 23.985 | 0.0368421 |
| 23.952 | 0.0421053 |
| 21.868 | 0.0473684 |
| 21.339 | 0.0526316 |
| 20.404 | 0.0578947 |
| 20.024 | 0.0631579 |
| 19.015 | 0.0684211 |
| 18.729 | 0.0736842 |
| 18.197 | 0.0789474 |
| 17.852 | 0.0842105 |
| 17.227 | 0.0894737 |
| 16.082 | 0.0947368 |
| 15.106 | 0.1 |
| 12.976 | 0.105263 |
| 12.76 | 0.110526 |
| 12.437 | 0.115789 |
| 11.76 | 0.121053 |
| 11.493 | 0.126316 |
| 11.105 | 0.131579 |
| 10.499 | 0.136842 |
| 10.347 | 0.142105 |
| 10.339 | 0.147368 |
| 10.282 | 0.152632 |
| 9.429 | 0.157895 |
| 9.08 | 0.163158 |
| 8.712 | 0.168421 |
| 8.286 | 0.173684 |
| 8.231 | 0.178947 |
| 7.86 | 0.184211 |
| 7.827 | 0.189474 |
| 7.667 | 0.194737 |
| 7.506 | 0.2 |
| 7.494 | 0.205263 |
| 7.298 | 0.210526 |
| 7.042 | 0.215789 |
| 6.995 | 0.221053 |
| 6.982 | 0.226316 |
| 6.977 | 0.231579 |
| 6.959 | 0.236842 |
| 6.676 | 0.242105 |
| 6.45 | 0.247368 |
| 6.294 | 0.252632 |
| 6.131 | 0.257895 |
| 6.069 | 0.263158 |
| 6.013 | 0.268421 |
| 5.909 | 0.273684 |
| 5.893 | 0.278947 |
| 5.822 | 0.284211 |
| 5.781 | 0.289474 |
| 5.623 | 0.294737 |
| 5.598 | 0.3 |
| 5.493 | 0.305263 |
| 5.468 | 0.310526 |
| 5.447 | 0.315789 |
| 5.437 | 0.321053 |
| 5.336 | 0.326316 |
| 5.098 | 0.331579 |
| 5.017 | 0.336842 |
| 4.995 | 0.342105 |
| 4.913 | 0.347368 |
| 4.713 | 0.352632 |
| 4.627 | 0.357895 |
| 4.534 | 0.363158 |
| 4.459 | 0.368421 |
| 4.211 | 0.373684 |
| 4.195 | 0.378947 |
| 4.098 | 0.384211 |
| 4.05 | 0.389474 |
| 3.965 | 0.394737 |
| 3.945 | 0.4 |
| 3.814 | 0.405263 |
| 3.688 | 0.410526 |
| 3.614 | 0.415789 |
| 3.476 | 0.421053 |
| 3.455 | 0.426316 |
| 3.412 | 0.431579 |
| 3.405 | 0.436842 |
| 3.233 | 0.442105 |
| 3.221 | 0.447368 |
| 3.165 | 0.452632 |
| 3.138 | 0.457895 |
| 3.129 | 0.463158 |
| 3.125 | 0.468421 |
| 3.092 | 0.473684 |
| 3.087 | 0.478947 |
| 2.99 | 0.484211 |
| 2.943 | 0.489474 |
| 2.924 | 0.494737 |
| 2.921 | 0.5 |
| 2.901 | 0.505263 |
| 2.893 | 0.510526 |
| 2.852 | 0.515789 |
| 2.827 | 0.521053 |
| 2.821 | 0.526316 |
| 2.781 | 0.531579 |
| 2.743 | 0.536842 |
| 2.697 | 0.542105 |
| 2.622 | 0.547368 |
| 2.55 | 0.552632 |
| 2.535 | 0.557895 |
| 2.53 | 0.563158 |
| 2.505 | 0.568421 |
| 2.419 | 0.573684 |
| 2.32 | 0.578947 |
| 2.312 | 0.584211 |
| 2.28 | 0.589474 |
| 2.221 | 0.594737 |
| 2.162 | 0.6 |
| 2.145 | 0.605263 |
| 2.141 | 0.610526 |
| 2.139 | 0.615789 |
| 2.036 | 0.621053 |
| 2.004 | 0.626316 |
| 1.994 | 0.631579 |
| 1.938 | 0.636842 |
| 1.926 | 0.642105 |
| 1.926 | 0.647368 |
| 1.894 | 0.652632 |
| 1.875 | 0.657895 |
| 1.858 | 0.663158 |
| 1.858 | 0.668421 |
| 1.836 | 0.673684 |
| 1.832 | 0.678947 |
| 1.796 | 0.684211 |
| 1.794 | 0.689474 |
| 1.793 | 0.694737 |
| 1.771 | 0.7 |
| 1.766 | 0.705263 |
| 1.756 | 0.710526 |
| 1.722 | 0.715789 |
| 1.719 | 0.721053 |
| 1.713 | 0.726316 |
| 1.7 | 0.731579 |
| 1.699 | 0.736842 |
| 1.675 | 0.742105 |
| 1.669 | 0.747368 |
| 1.663 | 0.752632 |
| 1.662 | 0.757895 |
| 1.653 | 0.763158 |
| 1.643 | 0.768421 |
| 1.614 | 0.773684 |
| 1.605 | 0.778947 |
| 1.59 | 0.784211 |
| 1.544 | 0.789474 |
| 1.521 | 0.794737 |
| 1.377 | 0.8 |
| 1.351 | 0.805263 |
| 1.314 | 0.810526 |
| 1.304 | 0.815789 |
| 1.295 | 0.821053 |
| 1.267 | 0.826316 |
| 1.2 | 0.831579 |
| 1.163 | 0.836842 |
| 1.14 | 0.842105 |
| 1.136 | 0.847368 |
| 1.13 | 0.852632 |
| 1.129 | 0.857895 |
| 1.118 | 0.863158 |
| 1.05 | 0.868421 |
| 1.006 | 0.873684 |
| 0.981 | 0.878947 |
| 0.926 | 0.884211 |
| 0.858 | 0.889474 |
| 0.841 | 0.894737 |
| 0.735 | 0.9 |
| 0.724 | 0.905263 |
| 0.653 | 0.910526 |
| 0.618 | 0.915789 |
| 0.565 | 0.921053 |
| 0.504 | 0.926316 |
| 0.371 | 0.931579 |
| 0.345 | 0.936842 |
| 0.332 | 0.942105 |
| 0.297 | 0.947368 |
| 0.27 | 0.952632 |
| 0.247 | 0.957895 |
| 0.231 | 0.963158 |
| 0.176 | 0.968421 |
| 0.139 | 0.973684 |
| 0.134 | 0.978947 |
| 0.125 | 0.984211 |
| 0.118 | 0.989474 |
| 0.116 | 0.994737 |
| 0.078 | 1 |

BECT056 - RCDC_Serotype 14

|  | ST 14-BE-PCV14 (Day 84) |
| --- | --- |
| 557.781 | 0.0052356 |
| 157.819 | 0.0104712 |
| 154.356 | 0.0157068 |
| 146.679 | 0.0209424 |
| 113.443 | 0.026178 |
| 106.272 | 0.0314136 |
| 98.362 | 0.0366492 |
| 87.747 | 0.0418848 |
| 86.816 | 0.0471204 |
| 83.17 | 0.052356 |
| 76.796 | 0.0575916 |
| 72.652 | 0.0628272 |
| 71.567 | 0.0680628 |
| 69.352 | 0.0732984 |
| 61.263 | 0.078534 |
| 60.6 | 0.0837696 |
| 56.678 | 0.0890052 |
| 55.487 | 0.0942408 |
| 53.73 | 0.0994764 |
| 53 | 0.104712 |
| 46.975 | 0.109948 |
| 45.211 | 0.115183 |
| 41.076 | 0.120419 |
| 39.976 | 0.125654 |
| 37.847 | 0.13089 |
| 37.826 | 0.136126 |
| 37.478 | 0.141361 |
| 36.967 | 0.146597 |
| 36.321 | 0.151832 |
| 36.027 | 0.157068 |
| 34.177 | 0.162304 |
| 33.306 | 0.167539 |
| 32.138 | 0.172775 |
| 31.154 | 0.17801 |
| 29.712 | 0.183246 |
| 29.575 | 0.188482 |
| 29.354 | 0.193717 |
| 28.096 | 0.198953 |
| 27.83 | 0.204188 |
| 27.196 | 0.209424 |
| 26.464 | 0.21466 |
| 24.045 | 0.219895 |
| 23.632 | 0.225131 |
| 22.828 | 0.230366 |
| 22.473 | 0.235602 |
| 22.063 | 0.240838 |
| 20.207 | 0.246073 |
| 19.44 | 0.251309 |
| 18.416 | 0.256545 |
| 17.978 | 0.26178 |
| 17.843 | 0.267016 |
| 17.656 | 0.272251 |
| 16 | 0.277487 |
| 15.616 | 0.282723 |
| 15.203 | 0.287958 |
| 14.649 | 0.293194 |
| 14.639 | 0.298429 |
| 14.332 | 0.303665 |
| 14.146 | 0.308901 |
| 13.983 | 0.314136 |
| 13.262 | 0.319372 |
| 12.997 | 0.324607 |
| 12.858 | 0.329843 |
| 12.817 | 0.335079 |
| 12.348 | 0.340314 |
| 11.998 | 0.34555 |
| 11.79 | 0.350785 |
| 11.556 | 0.356021 |
| 11.245 | 0.361257 |
| 11.11 | 0.366492 |
| 10.657 | 0.371728 |
| 10.655 | 0.376963 |
| 10.387 | 0.382199 |
| 10.164 | 0.387435 |
| 9.872 | 0.39267 |
| 9.867 | 0.397906 |
| 9.815 | 0.403141 |
| 9.646 | 0.408377 |
| 9.625 | 0.413613 |
| 9.244 | 0.418848 |
| 8.869 | 0.424084 |
| 8.833 | 0.429319 |
| 8.537 | 0.434555 |
| 8.501 | 0.439791 |
| 8.48 | 0.445026 |
| 8.377 | 0.450262 |
| 8.37 | 0.455497 |
| 8.3 | 0.460733 |
| 8.284 | 0.465969 |
| 8.279 | 0.471204 |
| 8.113 | 0.47644 |
| 8.084 | 0.481675 |
| 7.94 | 0.486911 |
| 7.888 | 0.492147 |
| 7.84 | 0.497382 |
| 7.525 | 0.502618 |
| 7.445 | 0.507853 |
| 7.257 | 0.513089 |
| 7.187 | 0.518325 |
| 7.081 | 0.52356 |
| 7.052 | 0.528796 |
| 6.998 | 0.534031 |
| 6.993 | 0.539267 |
| 6.936 | 0.544503 |
| 6.897 | 0.549738 |
| 6.303 | 0.554974 |
| 6.3 | 0.560209 |
| 6.192 | 0.565445 |
| 6.176 | 0.570681 |
| 6.051 | 0.575916 |
| 5.864 | 0.581152 |
| 5.812 | 0.586387 |
| 5.664 | 0.591623 |
| 5.578 | 0.596859 |
| 5.561 | 0.602094 |
| 5.519 | 0.60733 |
| 5.474 | 0.612565 |
| 5.385 | 0.617801 |
| 5.375 | 0.623037 |
| 5.246 | 0.628272 |
| 5.245 | 0.633508 |
| 5.239 | 0.638743 |
| 5.169 | 0.643979 |
| 5.131 | 0.649215 |
| 4.898 | 0.65445 |
| 4.417 | 0.659686 |
| 4.339 | 0.664921 |
| 4.33 | 0.670157 |
| 4.21 | 0.675393 |
| 4.195 | 0.680628 |
| 4.117 | 0.685864 |
| 4.115 | 0.691099 |
| 3.99 | 0.696335 |
| 3.96 | 0.701571 |
| 3.78 | 0.706806 |
| 3.477 | 0.712042 |
| 3.391 | 0.717277 |
| 3.347 | 0.722513 |
| 3.29 | 0.727749 |
| 3.277 | 0.732984 |
| 3.22 | 0.73822 |
| 3.128 | 0.743455 |
| 3.087 | 0.748691 |
| 2.992 | 0.753927 |
| 2.819 | 0.759162 |
| 2.769 | 0.764398 |
| 2.765 | 0.769634 |
| 2.702 | 0.774869 |
| 2.601 | 0.780105 |
| 2.529 | 0.78534 |
| 2.482 | 0.790576 |
| 2.482 | 0.795812 |
| 2.455 | 0.801047 |
| 2.423 | 0.806283 |
| 2.391 | 0.811518 |
| 2.389 | 0.816754 |
| 2.324 | 0.82199 |
| 2.321 | 0.827225 |
| 2.297 | 0.832461 |
| 2.285 | 0.837696 |
| 2.284 | 0.842932 |
| 2.205 | 0.848168 |
| 2.187 | 0.853403 |
| 2.179 | 0.858639 |
| 2.051 | 0.863874 |
| 2.022 | 0.86911 |
| 2.018 | 0.874346 |
| 2.013 | 0.879581 |
| 1.977 | 0.884817 |
| 1.963 | 0.890052 |
| 1.935 | 0.895288 |
| 1.889 | 0.900524 |
| 1.863 | 0.905759 |
| 1.835 | 0.910995 |
| 1.802 | 0.91623 |
| 1.764 | 0.921466 |
| 1.745 | 0.926702 |
| 1.623 | 0.931937 |
| 1.619 | 0.937173 |
| 1.562 | 0.942408 |
| 1.358 | 0.947644 |
| 1.339 | 0.95288 |
| 1.247 | 0.958115 |
| 1.221 | 0.963351 |
| 1.179 | 0.968586 |
| 1.105 | 0.973822 |
| 0.91 | 0.979058 |
| 0.89 | 0.984293 |
| 0.829 | 0.989529 |
| 0.74 | 0.994764 |
| 0.479 | 1 |

|  | ST 14-Prevenar 13 (Day 84) |
| --- | --- |
| 176.695 | 0.00505051 |
| 168.969 | 0.010101 |
| 146.377 | 0.0151515 |
| 144.9 | 0.020202 |
| 143.489 | 0.0252525 |
| 141.727 | 0.030303 |
| 107.035 | 0.0353535 |
| 105.273 | 0.040404 |
| 84.683 | 0.0454545 |
| 66.644 | 0.0505051 |
| 65.184 | 0.0555556 |
| 59.051 | 0.0606061 |
| 57.423 | 0.0656566 |
| 42.582 | 0.0707071 |
| 42.567 | 0.0757576 |
| 42.098 | 0.0808081 |
| 42.027 | 0.0858586 |
| 41.47 | 0.0909091 |
| 40.356 | 0.0959596 |
| 40.125 | 0.10101 |
| 37.719 | 0.106061 |
| 37.57 | 0.111111 |
| 34.256 | 0.116162 |
| 33.55 | 0.121212 |
| 31.62 | 0.126263 |
| 28.125 | 0.131313 |
| 28.095 | 0.136364 |
| 26.269 | 0.141414 |
| 26.16 | 0.146465 |
| 26.062 | 0.151515 |
| 25.461 | 0.156566 |
| 24.973 | 0.161616 |
| 24.57 | 0.166667 |
| 24.287 | 0.171717 |
| 24.133 | 0.176768 |
| 23.521 | 0.181818 |
| 23.447 | 0.186869 |
| 22.331 | 0.191919 |
| 22.113 | 0.19697 |
| 22.083 | 0.20202 |
| 21.833 | 0.207071 |
| 21.624 | 0.212121 |
| 21.583 | 0.217172 |
| 20.596 | 0.222222 |
| 20.451 | 0.227273 |
| 20.413 | 0.232323 |
| 20.028 | 0.237374 |
| 19.75 | 0.242424 |
| 19.464 | 0.247475 |
| 18.893 | 0.252525 |
| 18.394 | 0.257576 |
| 17.63 | 0.262626 |
| 17.163 | 0.267677 |
| 17.107 | 0.272727 |
| 16.506 | 0.277778 |
| 15.363 | 0.282828 |
| 15.178 | 0.287879 |
| 14.238 | 0.292929 |
| 14.2 | 0.29798 |
| 13.651 | 0.30303 |
| 13.624 | 0.308081 |
| 13.337 | 0.313131 |
| 13.267 | 0.318182 |
| 13.179 | 0.323232 |
| 12.848 | 0.328283 |
| 12.251 | 0.333333 |
| 11.987 | 0.338384 |
| 11.926 | 0.343434 |
| 11.464 | 0.348485 |
| 11.462 | 0.353535 |
| 11.008 | 0.358586 |
| 10.75 | 0.363636 |
| 10.545 | 0.368687 |
| 9.666 | 0.373737 |
| 9.472 | 0.378788 |
| 8.965 | 0.383838 |
| 8.531 | 0.388889 |
| 8.152 | 0.393939 |
| 8.148 | 0.39899 |
| 7.992 | 0.40404 |
| 7.982 | 0.409091 |
| 7.858 | 0.414141 |
| 7.785 | 0.419192 |
| 7.682 | 0.424242 |
| 7.553 | 0.429293 |
| 7.506 | 0.434343 |
| 7.4 | 0.439394 |
| 6.898 | 0.444444 |
| 6.828 | 0.449495 |
| 6.715 | 0.454545 |
| 6.66 | 0.459596 |
| 6.63 | 0.464646 |
| 6.618 | 0.469697 |
| 6.616 | 0.474747 |
| 6.542 | 0.479798 |
| 6.5 | 0.484848 |
| 6.381 | 0.489899 |
| 6.341 | 0.494949 |
| 6.305 | 0.5 |
| 6.21 | 0.505051 |
| 5.873 | 0.510101 |
| 5.845 | 0.515152 |
| 5.655 | 0.520202 |
| 5.578 | 0.525253 |
| 5.499 | 0.530303 |
| 5.253 | 0.535354 |
| 5.234 | 0.540404 |
| 5.126 | 0.545455 |
| 5.061 | 0.550505 |
| 5.021 | 0.555556 |
| 4.861 | 0.560606 |
| 4.733 | 0.565657 |
| 4.73 | 0.570707 |
| 4.573 | 0.575758 |
| 4.468 | 0.580808 |
| 4.364 | 0.585859 |
| 4.33 | 0.590909 |
| 4.266 | 0.59596 |
| 4.073 | 0.60101 |
| 4.001 | 0.606061 |
| 3.951 | 0.611111 |
| 3.834 | 0.616162 |
| 3.653 | 0.621212 |
| 3.555 | 0.626263 |
| 3.289 | 0.631313 |
| 3.289 | 0.636364 |
| 3.187 | 0.641414 |
| 3.175 | 0.646465 |
| 3.141 | 0.651515 |
| 3.044 | 0.656566 |
| 3.014 | 0.661616 |
| 3.013 | 0.666667 |
| 2.995 | 0.671717 |
| 2.986 | 0.676768 |
| 2.974 | 0.681818 |
| 2.888 | 0.686869 |
| 2.86 | 0.691919 |
| 2.813 | 0.69697 |
| 2.757 | 0.70202 |
| 2.726 | 0.707071 |
| 2.655 | 0.712121 |
| 2.654 | 0.717172 |
| 2.579 | 0.722222 |
| 2.541 | 0.727273 |
| 2.521 | 0.732323 |
| 2.472 | 0.737374 |
| 2.469 | 0.742424 |
| 2.463 | 0.747475 |
| 2.441 | 0.752525 |
| 2.391 | 0.757576 |
| 2.304 | 0.762626 |
| 2.274 | 0.767677 |
| 2.143 | 0.772727 |
| 2.115 | 0.777778 |
| 2.112 | 0.782828 |
| 2.105 | 0.787879 |
| 2.059 | 0.792929 |
| 1.971 | 0.79798 |
| 1.962 | 0.80303 |
| 1.924 | 0.808081 |
| 1.85 | 0.813131 |
| 1.736 | 0.818182 |
| 1.736 | 0.823232 |
| 1.706 | 0.828283 |
| 1.705 | 0.833333 |
| 1.598 | 0.838384 |
| 1.592 | 0.843434 |
| 1.586 | 0.848485 |
| 1.542 | 0.853535 |
| 1.542 | 0.858586 |
| 1.479 | 0.863636 |
| 1.471 | 0.868687 |
| 1.462 | 0.873737 |
| 1.453 | 0.878788 |
| 1.407 | 0.883838 |
| 1.4 | 0.888889 |
| 1.398 | 0.893939 |
| 1.395 | 0.89899 |
| 1.345 | 0.90404 |
| 1.321 | 0.909091 |
| 1.318 | 0.914141 |
| 1.268 | 0.919192 |
| 1.254 | 0.924242 |
| 1.248 | 0.929293 |
| 1.2 | 0.934343 |
| 1.185 | 0.939394 |
| 1.124 | 0.944444 |
| 1.069 | 0.949495 |
| 1.059 | 0.954545 |
| 0.95 | 0.959596 |
| 0.875 | 0.964646 |
| 0.79 | 0.969697 |
| 0.778 | 0.974747 |
| 0.773 | 0.979798 |
| 0.727 | 0.984848 |
| 0.715 | 0.989899 |
| 0.476 | 0.994949 |
| 0.452 | 1 |

|  | ST 14-BE-PCV14 (Day 300) |
| --- | --- |
| 82.989 | 0.00531915 |
| 79.603 | 0.0106383 |
| 74.885 | 0.0159574 |
| 72.872 | 0.0212766 |
| 71.25 | 0.0265957 |
| 67.858 | 0.0319149 |
| 61.971 | 0.037234 |
| 61.479 | 0.0425532 |
| 57.07 | 0.0478723 |
| 52.069 | 0.0531915 |
| 51.987 | 0.0585106 |
| 45.921 | 0.0638298 |
| 45.361 | 0.0691489 |
| 42.934 | 0.0744681 |
| 39.059 | 0.0797872 |
| 38.565 | 0.0851064 |
| 36.435 | 0.0904255 |
| 34.942 | 0.0957447 |
| 34.915 | 0.101064 |
| 33.728 | 0.106383 |
| 33.706 | 0.111702 |
| 33.604 | 0.117021 |
| 33.599 | 0.12234 |
| 33.076 | 0.12766 |
| 31.371 | 0.132979 |
| 30.225 | 0.138298 |
| 29.872 | 0.143617 |
| 29.505 | 0.148936 |
| 28.731 | 0.154255 |
| 28.71 | 0.159574 |
| 28.139 | 0.164894 |
| 27.648 | 0.170213 |
| 27 | 0.175532 |
| 26.585 | 0.180851 |
| 25.189 | 0.18617 |
| 23.238 | 0.191489 |
| 22.909 | 0.196809 |
| 22.527 | 0.202128 |
| 22.133 | 0.207447 |
| 22.057 | 0.212766 |
| 21.869 | 0.218085 |
| 20.877 | 0.223404 |
| 20.766 | 0.228723 |
| 20.171 | 0.234043 |
| 19.9 | 0.239362 |
| 19.558 | 0.244681 |
| 19.424 | 0.25 |
| 19.325 | 0.255319 |
| 19.319 | 0.260638 |
| 19.271 | 0.265957 |
| 18.884 | 0.271277 |
| 18.675 | 0.276596 |
| 18.42 | 0.281915 |
| 18.195 | 0.287234 |
| 18.007 | 0.292553 |
| 17.704 | 0.297872 |
| 17.632 | 0.303191 |
| 17.468 | 0.308511 |
| 17.239 | 0.31383 |
| 17.1 | 0.319149 |
| 16.933 | 0.324468 |
| 16.866 | 0.329787 |
| 16.748 | 0.335106 |
| 16.711 | 0.340426 |
| 16.299 | 0.345745 |
| 16.036 | 0.351064 |
| 15.871 | 0.356383 |
| 15.781 | 0.361702 |
| 15.638 | 0.367021 |
| 15.579 | 0.37234 |
| 15.416 | 0.37766 |
| 15.314 | 0.382979 |
| 15.219 | 0.388298 |
| 15.074 | 0.393617 |
| 15.049 | 0.398936 |
| 14.955 | 0.404255 |
| 14.807 | 0.409574 |
| 14.034 | 0.414894 |
| 13.948 | 0.420213 |
| 13.873 | 0.425532 |
| 13.747 | 0.430851 |
| 13.544 | 0.43617 |
| 13.504 | 0.441489 |
| 12.933 | 0.446809 |
| 12.906 | 0.452128 |
| 12.64 | 0.457447 |
| 12.05 | 0.462766 |
| 11.996 | 0.468085 |
| 11.858 | 0.473404 |
| 11.525 | 0.478723 |
| 11.412 | 0.484043 |
| 11.411 | 0.489362 |
| 11.231 | 0.494681 |
| 10.69 | 0.5 |
| 10.28 | 0.505319 |
| 10.225 | 0.510638 |
| 10.22 | 0.515957 |
| 10.009 | 0.521277 |
| 9.931 | 0.526596 |
| 9.481 | 0.531915 |
| 9.392 | 0.537234 |
| 9.107 | 0.542553 |
| 8.775 | 0.547872 |
| 8.566 | 0.553191 |
| 8.538 | 0.558511 |
| 8.242 | 0.56383 |
| 8.178 | 0.569149 |
| 8.042 | 0.574468 |
| 7.906 | 0.579787 |
| 7.886 | 0.585106 |
| 7.681 | 0.590426 |
| 7.445 | 0.595745 |
| 7.343 | 0.601064 |
| 7.299 | 0.606383 |
| 7.151 | 0.611702 |
| 7.014 | 0.617021 |
| 7.013 | 0.62234 |
| 6.998 | 0.62766 |
| 6.932 | 0.632979 |
| 6.874 | 0.638298 |
| 6.804 | 0.643617 |
| 6.791 | 0.648936 |
| 6.777 | 0.654255 |
| 6.613 | 0.659574 |
| 6.285 | 0.664894 |
| 6.275 | 0.670213 |
| 6.257 | 0.675532 |
| 6.246 | 0.680851 |
| 6.153 | 0.68617 |
| 6.134 | 0.691489 |
| 6.093 | 0.696809 |
| 6.071 | 0.702128 |
| 5.63 | 0.707447 |
| 5.574 | 0.712766 |
| 5.554 | 0.718085 |
| 5.407 | 0.723404 |
| 5.339 | 0.728723 |
| 5.282 | 0.734043 |
| 5.207 | 0.739362 |
| 5.125 | 0.744681 |
| 5.119 | 0.75 |
| 5.082 | 0.755319 |
| 5.057 | 0.760638 |
| 4.9 | 0.765957 |
| 4.87 | 0.771277 |
| 4.842 | 0.776596 |
| 4.808 | 0.781915 |
| 4.736 | 0.787234 |
| 4.684 | 0.792553 |
| 4.46 | 0.797872 |
| 4.354 | 0.803191 |
| 4.146 | 0.808511 |
| 4.123 | 0.81383 |
| 3.992 | 0.819149 |
| 3.905 | 0.824468 |
| 3.852 | 0.829787 |
| 3.791 | 0.835106 |
| 3.748 | 0.840426 |
| 3.693 | 0.845745 |
| 3.67 | 0.851064 |
| 3.635 | 0.856383 |
| 3.287 | 0.861702 |
| 3.135 | 0.867021 |
| 3.079 | 0.87234 |
| 2.731 | 0.87766 |
| 2.706 | 0.882979 |
| 2.637 | 0.888298 |
| 2.619 | 0.893617 |
| 2.614 | 0.898936 |
| 2.557 | 0.904255 |
| 2.499 | 0.909574 |
| 2.493 | 0.914894 |
| 2.417 | 0.920213 |
| 2.39 | 0.925532 |
| 2.149 | 0.930851 |
| 2.126 | 0.93617 |
| 2.066 | 0.941489 |
| 2.01 | 0.946809 |
| 1.531 | 0.952128 |
| 1.495 | 0.957447 |
| 1.471 | 0.962766 |
| 1.413 | 0.968085 |
| 1.387 | 0.973404 |
| 1.376 | 0.978723 |
| 1.359 | 0.984043 |
| 1.349 | 0.989362 |
| 1.308 | 0.994681 |
| 1.255 | 1 |

|  | ST 14-Prevenar 13 (Day 300) |
| --- | --- |
| 91.961 | 0.00526316 |
| 83.006 | 0.0105263 |
| 81.055 | 0.0157895 |
| 75.354 | 0.0210526 |
| 74.886 | 0.0263158 |
| 68.767 | 0.0315789 |
| 68.38 | 0.0368421 |
| 64.564 | 0.0421053 |
| 56.347 | 0.0473684 |
| 54.177 | 0.0526316 |
| 53.134 | 0.0578947 |
| 52.16 | 0.0631579 |
| 48.323 | 0.0684211 |
| 47.628 | 0.0736842 |
| 47.217 | 0.0789474 |
| 47.026 | 0.0842105 |
| 46.361 | 0.0894737 |
| 39.921 | 0.0947368 |
| 37.509 | 0.1 |
| 36.083 | 0.105263 |
| 35.482 | 0.110526 |
| 33.869 | 0.115789 |
| 33.779 | 0.121053 |
| 33.33 | 0.126316 |
| 32.453 | 0.131579 |
| 32.284 | 0.136842 |
| 30.241 | 0.142105 |
| 29.723 | 0.147368 |
| 29.616 | 0.152632 |
| 29.419 | 0.157895 |
| 26.703 | 0.163158 |
| 25.32 | 0.168421 |
| 24.457 | 0.173684 |
| 23.845 | 0.178947 |
| 23.761 | 0.184211 |
| 23.411 | 0.189474 |
| 22.881 | 0.194737 |
| 22.83 | 0.2 |
| 21.795 | 0.205263 |
| 21.502 | 0.210526 |
| 21.316 | 0.215789 |
| 20.919 | 0.221053 |
| 20.894 | 0.226316 |
| 20.727 | 0.231579 |
| 20.661 | 0.236842 |
| 20.655 | 0.242105 |
| 20.35 | 0.247368 |
| 19.415 | 0.252632 |
| 18.789 | 0.257895 |
| 18.687 | 0.263158 |
| 18.646 | 0.268421 |
| 17.559 | 0.273684 |
| 17.46 | 0.278947 |
| 16.871 | 0.284211 |
| 16.787 | 0.289474 |
| 16.325 | 0.294737 |
| 16.24 | 0.3 |
| 16.072 | 0.305263 |
| 15.542 | 0.310526 |
| 15.038 | 0.315789 |
| 14.985 | 0.321053 |
| 14.925 | 0.326316 |
| 14.837 | 0.331579 |
| 14.76 | 0.336842 |
| 14.729 | 0.342105 |
| 14.294 | 0.347368 |
| 14.208 | 0.352632 |
| 14.12 | 0.357895 |
| 13.52 | 0.363158 |
| 13.446 | 0.368421 |
| 13.137 | 0.373684 |
| 13.078 | 0.378947 |
| 12.819 | 0.384211 |
| 12.811 | 0.389474 |
| 12.768 | 0.394737 |
| 12.56 | 0.4 |
| 12.499 | 0.405263 |
| 12.422 | 0.410526 |
| 12.328 | 0.415789 |
| 12.292 | 0.421053 |
| 12.25 | 0.426316 |
| 11.821 | 0.431579 |
| 11.736 | 0.436842 |
| 11.652 | 0.442105 |
| 11.52 | 0.447368 |
| 11.362 | 0.452632 |
| 11.306 | 0.457895 |
| 11.273 | 0.463158 |
| 11.119 | 0.468421 |
| 11.078 | 0.473684 |
| 11.039 | 0.478947 |
| 10.826 | 0.484211 |
| 10.441 | 0.489474 |
| 10.256 | 0.494737 |
| 10.208 | 0.5 |
| 10.189 | 0.505263 |
| 10.071 | 0.510526 |
| 9.999 | 0.515789 |
| 9.986 | 0.521053 |
| 9.959 | 0.526316 |
| 9.802 | 0.531579 |
| 9.668 | 0.536842 |
| 9.521 | 0.542105 |
| 9.277 | 0.547368 |
| 9.165 | 0.552632 |
| 9.085 | 0.557895 |
| 8.999 | 0.563158 |
| 8.776 | 0.568421 |
| 8.574 | 0.573684 |
| 8.542 | 0.578947 |
| 8.492 | 0.584211 |
| 8.437 | 0.589474 |
| 8.397 | 0.594737 |
| 8.319 | 0.6 |
| 8.315 | 0.605263 |
| 8.092 | 0.610526 |
| 8.077 | 0.615789 |
| 7.933 | 0.621053 |
| 7.881 | 0.626316 |
| 7.8 | 0.631579 |
| 7.719 | 0.636842 |
| 7.595 | 0.642105 |
| 7.589 | 0.647368 |
| 7.549 | 0.652632 |
| 7.419 | 0.657895 |
| 7.408 | 0.663158 |
| 7.385 | 0.668421 |
| 7.351 | 0.673684 |
| 7.187 | 0.678947 |
| 6.977 | 0.684211 |
| 6.943 | 0.689474 |
| 6.887 | 0.694737 |
| 6.87 | 0.7 |
| 6.809 | 0.705263 |
| 6.421 | 0.710526 |
| 6.359 | 0.715789 |
| 6.242 | 0.721053 |
| 6.052 | 0.726316 |
| 5.887 | 0.731579 |
| 5.581 | 0.736842 |
| 5.544 | 0.742105 |
| 5.337 | 0.747368 |
| 5.305 | 0.752632 |
| 5.24 | 0.757895 |
| 5.194 | 0.763158 |
| 5.14 | 0.768421 |
| 5.132 | 0.773684 |
| 5.132 | 0.778947 |
| 5.086 | 0.784211 |
| 5.045 | 0.789474 |
| 5.023 | 0.794737 |
| 4.993 | 0.8 |
| 4.936 | 0.805263 |
| 4.89 | 0.810526 |
| 4.882 | 0.815789 |
| 4.88 | 0.821053 |
| 4.866 | 0.826316 |
| 4.821 | 0.831579 |
| 4.765 | 0.836842 |
| 4.62 | 0.842105 |
| 4.566 | 0.847368 |
| 4.558 | 0.852632 |
| 4.392 | 0.857895 |
| 4.339 | 0.863158 |
| 4.329 | 0.868421 |
| 4.266 | 0.873684 |
| 4.104 | 0.878947 |
| 4.099 | 0.884211 |
| 4.054 | 0.889474 |
| 3.987 | 0.894737 |
| 3.935 | 0.9 |
| 3.718 | 0.905263 |
| 3.683 | 0.910526 |
| 3.629 | 0.915789 |
| 3.519 | 0.921053 |
| 3.232 | 0.926316 |
| 3.15 | 0.931579 |
| 2.996 | 0.936842 |
| 2.917 | 0.942105 |
| 2.893 | 0.947368 |
| 2.889 | 0.952632 |
| 2.8 | 0.957895 |
| 2.773 | 0.963158 |
| 2.706 | 0.968421 |
| 2.593 | 0.973684 |
| 1.083 | 0.978947 |
| 1.021 | 0.984211 |
| 0.893 | 0.989474 |
| 0.681 | 0.994737 |
| 0.677 | 1 |

BECT056 - RCDC_Serotype 18C

|  | ST 18C-BE-PCV14 (Day 84) |
| --- | --- |
| 68.07 | 0.0052356 |
| 59.936 | 0.0104712 |
| 35.771 | 0.0157068 |
| 35.445 | 0.0209424 |
| 31.087 | 0.026178 |
| 28.903 | 0.0314136 |
| 28.235 | 0.0366492 |
| 25.776 | 0.0418848 |
| 24.629 | 0.0471204 |
| 24.018 | 0.052356 |
| 22.923 | 0.0575916 |
| 21.742 | 0.0628272 |
| 21.581 | 0.0680628 |
| 19.514 | 0.0732984 |
| 19.25 | 0.078534 |
| 18.869 | 0.0837696 |
| 16.991 | 0.0890052 |
| 16.847 | 0.0942408 |
| 16.569 | 0.0994764 |
| 15.208 | 0.104712 |
| 14.958 | 0.109948 |
| 14.667 | 0.115183 |
| 12.069 | 0.120419 |
| 11.63 | 0.125654 |
| 11.566 | 0.13089 |
| 11.406 | 0.136126 |
| 11.186 | 0.141361 |
| 11.14 | 0.146597 |
| 10.554 | 0.151832 |
| 10.395 | 0.157068 |
| 9.258 | 0.162304 |
| 8.205 | 0.167539 |
| 8.03 | 0.172775 |
| 7.919 | 0.17801 |
| 7.559 | 0.183246 |
| 6.35 | 0.188482 |
| 5.621 | 0.193717 |
| 5.272 | 0.198953 |
| 4.873 | 0.204188 |
| 4.515 | 0.209424 |
| 4.133 | 0.21466 |
| 3.569 | 0.219895 |
| 3.453 | 0.225131 |
| 3.313 | 0.230366 |
| 3.293 | 0.235602 |
| 3.149 | 0.240838 |
| 3.136 | 0.246073 |
| 3.093 | 0.251309 |
| 2.723 | 0.256545 |
| 2.719 | 0.26178 |
| 2.696 | 0.267016 |
| 2.504 | 0.272251 |
| 2.226 | 0.277487 |
| 2.124 | 0.282723 |
| 2.11 | 0.287958 |
| 2.097 | 0.293194 |
| 2.001 | 0.298429 |
| 1.875 | 0.303665 |
| 1.825 | 0.308901 |
| 1.782 | 0.314136 |
| 1.742 | 0.319372 |
| 1.713 | 0.324607 |
| 1.704 | 0.329843 |
| 1.698 | 0.335079 |
| 1.654 | 0.340314 |
| 1.648 | 0.34555 |
| 1.647 | 0.350785 |
| 1.608 | 0.356021 |
| 1.582 | 0.361257 |
| 1.581 | 0.366492 |
| 1.58 | 0.371728 |
| 1.569 | 0.376963 |
| 1.565 | 0.382199 |
| 1.564 | 0.387435 |
| 1.552 | 0.39267 |
| 1.535 | 0.397906 |
| 1.5 | 0.403141 |
| 1.489 | 0.408377 |
| 1.445 | 0.413613 |
| 1.386 | 0.418848 |
| 1.377 | 0.424084 |
| 1.373 | 0.429319 |
| 1.364 | 0.434555 |
| 1.356 | 0.439791 |
| 1.352 | 0.445026 |
| 1.308 | 0.450262 |
| 1.261 | 0.455497 |
| 1.257 | 0.460733 |
| 1.226 | 0.465969 |
| 1.191 | 0.471204 |
| 1.19 | 0.47644 |
| 1.178 | 0.481675 |
| 1.177 | 0.486911 |
| 1.17 | 0.492147 |
| 1.165 | 0.497382 |
| 1.142 | 0.502618 |
| 1.113 | 0.507853 |
| 1.096 | 0.513089 |
| 1.09 | 0.518325 |
| 1.067 | 0.52356 |
| 1.041 | 0.528796 |
| 1.039 | 0.534031 |
| 1.015 | 0.539267 |
| 0.943 | 0.544503 |
| 0.936 | 0.549738 |
| 0.909 | 0.554974 |
| 0.907 | 0.560209 |
| 0.897 | 0.565445 |
| 0.896 | 0.570681 |
| 0.888 | 0.575916 |
| 0.854 | 0.581152 |
| 0.845 | 0.586387 |
| 0.8 | 0.591623 |
| 0.792 | 0.596859 |
| 0.777 | 0.602094 |
| 0.767 | 0.60733 |
| 0.755 | 0.612565 |
| 0.742 | 0.617801 |
| 0.727 | 0.623037 |
| 0.724 | 0.628272 |
| 0.701 | 0.633508 |
| 0.698 | 0.638743 |
| 0.691 | 0.643979 |
| 0.682 | 0.649215 |
| 0.664 | 0.65445 |
| 0.663 | 0.659686 |
| 0.661 | 0.664921 |
| 0.657 | 0.670157 |
| 0.656 | 0.675393 |
| 0.648 | 0.680628 |
| 0.628 | 0.685864 |
| 0.619 | 0.691099 |
| 0.605 | 0.696335 |
| 0.601 | 0.701571 |
| 0.59 | 0.706806 |
| 0.583 | 0.712042 |
| 0.568 | 0.717277 |
| 0.566 | 0.722513 |
| 0.551 | 0.727749 |
| 0.549 | 0.732984 |
| 0.544 | 0.73822 |
| 0.539 | 0.743455 |
| 0.533 | 0.748691 |
| 0.53 | 0.753927 |
| 0.516 | 0.759162 |
| 0.481 | 0.764398 |
| 0.479 | 0.769634 |
| 0.478 | 0.774869 |
| 0.477 | 0.780105 |
| 0.47 | 0.78534 |
| 0.465 | 0.790576 |
| 0.462 | 0.795812 |
| 0.46 | 0.801047 |
| 0.457 | 0.806283 |
| 0.441 | 0.811518 |
| 0.435 | 0.816754 |
| 0.424 | 0.82199 |
| 0.422 | 0.827225 |
| 0.406 | 0.832461 |
| 0.405 | 0.837696 |
| 0.395 | 0.842932 |
| 0.394 | 0.848168 |
| 0.388 | 0.853403 |
| 0.388 | 0.858639 |
| 0.385 | 0.863874 |
| 0.384 | 0.86911 |
| 0.384 | 0.874346 |
| 0.381 | 0.879581 |
| 0.375 | 0.884817 |
| 0.371 | 0.890052 |
| 0.349 | 0.895288 |
| 0.343 | 0.900524 |
| 0.342 | 0.905759 |
| 0.34 | 0.910995 |
| 0.339 | 0.91623 |
| 0.335 | 0.921466 |
| 0.331 | 0.926702 |
| 0.319 | 0.931937 |
| 0.302 | 0.937173 |
| 0.298 | 0.942408 |
| 0.286 | 0.947644 |
| 0.284 | 0.95288 |
| 0.276 | 0.958115 |
| 0.27 | 0.963351 |
| 0.227 | 0.968586 |
| 0.217 | 0.973822 |
| 0.192 | 0.979058 |
| 0.188 | 0.984293 |
| 0.188 | 0.989529 |
| 0.182 | 0.994764 |
| 0.088 | 1 |

|  | ST 18C-Prevenar 13 (Day 84) |
| --- | --- |
| 80.804 | 0.00505051 |
| 69.408 | 0.010101 |
| 51.709 | 0.0151515 |
| 50.994 | 0.020202 |
| 47.995 | 0.0252525 |
| 47.013 | 0.030303 |
| 40.574 | 0.0353535 |
| 39.305 | 0.040404 |
| 32.09 | 0.0454545 |
| 29.248 | 0.0505051 |
| 28.089 | 0.0555556 |
| 26.377 | 0.0606061 |
| 24.277 | 0.0656566 |
| 21.63 | 0.0707071 |
| 21.281 | 0.0757576 |
| 21.074 | 0.0808081 |
| 20.685 | 0.0858586 |
| 20.442 | 0.0909091 |
| 20.177 | 0.0959596 |
| 19.239 | 0.10101 |
| 18.417 | 0.106061 |
| 17.31 | 0.111111 |
| 14.266 | 0.116162 |
| 14.126 | 0.121212 |
| 11.915 | 0.126263 |
| 11.654 | 0.131313 |
| 10.971 | 0.136364 |
| 9.946 | 0.141414 |
| 9.753 | 0.146465 |
| 9.511 | 0.151515 |
| 9.326 | 0.156566 |
| 8.445 | 0.161616 |
| 8.381 | 0.166667 |
| 8.211 | 0.171717 |
| 7.46 | 0.176768 |
| 7.328 | 0.181818 |
| 7.019 | 0.186869 |
| 6.862 | 0.191919 |
| 6.755 | 0.19697 |
| 6.327 | 0.20202 |
| 5.048 | 0.207071 |
| 4.9 | 0.212121 |
| 4.582 | 0.217172 |
| 4.382 | 0.222222 |
| 4.371 | 0.227273 |
| 4.347 | 0.232323 |
| 4.159 | 0.237374 |
| 4.101 | 0.242424 |
| 4.017 | 0.247475 |
| 3.977 | 0.252525 |
| 3.973 | 0.257576 |
| 3.936 | 0.262626 |
| 3.912 | 0.267677 |
| 3.729 | 0.272727 |
| 3.714 | 0.277778 |
| 3.665 | 0.282828 |
| 3.655 | 0.287879 |
| 3.651 | 0.292929 |
| 3.545 | 0.29798 |
| 3.529 | 0.30303 |
| 3.426 | 0.308081 |
| 3.373 | 0.313131 |
| 3.327 | 0.318182 |
| 3.325 | 0.323232 |
| 3.256 | 0.328283 |
| 3.245 | 0.333333 |
| 3.236 | 0.338384 |
| 3.202 | 0.343434 |
| 3.194 | 0.348485 |
| 3.184 | 0.353535 |
| 3.161 | 0.358586 |
| 3.098 | 0.363636 |
| 3.035 | 0.368687 |
| 2.948 | 0.373737 |
| 2.916 | 0.378788 |
| 2.899 | 0.383838 |
| 2.867 | 0.388889 |
| 2.854 | 0.393939 |
| 2.793 | 0.39899 |
| 2.763 | 0.40404 |
| 2.757 | 0.409091 |
| 2.751 | 0.414141 |
| 2.652 | 0.419192 |
| 2.64 | 0.424242 |
| 2.601 | 0.429293 |
| 2.532 | 0.434343 |
| 2.476 | 0.439394 |
| 2.439 | 0.444444 |
| 2.436 | 0.449495 |
| 2.419 | 0.454545 |
| 2.408 | 0.459596 |
| 2.372 | 0.464646 |
| 2.348 | 0.469697 |
| 2.314 | 0.474747 |
| 2.253 | 0.479798 |
| 2.235 | 0.484848 |
| 2.233 | 0.489899 |
| 2.209 | 0.494949 |
| 2.111 | 0.5 |
| 2.098 | 0.505051 |
| 2.032 | 0.510101 |
| 2.008 | 0.515152 |
| 1.953 | 0.520202 |
| 1.946 | 0.525253 |
| 1.886 | 0.530303 |
| 1.875 | 0.535354 |
| 1.852 | 0.540404 |
| 1.843 | 0.545455 |
| 1.824 | 0.550505 |
| 1.809 | 0.555556 |
| 1.786 | 0.560606 |
| 1.785 | 0.565657 |
| 1.767 | 0.570707 |
| 1.762 | 0.575758 |
| 1.75 | 0.580808 |
| 1.738 | 0.585859 |
| 1.735 | 0.590909 |
| 1.731 | 0.59596 |
| 1.716 | 0.60101 |
| 1.698 | 0.606061 |
| 1.659 | 0.611111 |
| 1.658 | 0.616162 |
| 1.642 | 0.621212 |
| 1.633 | 0.626263 |
| 1.629 | 0.631313 |
| 1.621 | 0.636364 |
| 1.569 | 0.641414 |
| 1.561 | 0.646465 |
| 1.555 | 0.651515 |
| 1.54 | 0.656566 |
| 1.527 | 0.661616 |
| 1.519 | 0.666667 |
| 1.499 | 0.671717 |
| 1.484 | 0.676768 |
| 1.453 | 0.681818 |
| 1.415 | 0.686869 |
| 1.407 | 0.691919 |
| 1.365 | 0.69697 |
| 1.346 | 0.70202 |
| 1.336 | 0.707071 |
| 1.307 | 0.712121 |
| 1.3 | 0.717172 |
| 1.27 | 0.722222 |
| 1.269 | 0.727273 |
| 1.261 | 0.732323 |
| 1.257 | 0.737374 |
| 1.245 | 0.742424 |
| 1.185 | 0.747475 |
| 1.185 | 0.752525 |
| 1.171 | 0.757576 |
| 1.168 | 0.762626 |
| 1.159 | 0.767677 |
| 1.098 | 0.772727 |
| 1.094 | 0.777778 |
| 1.085 | 0.782828 |
| 1.059 | 0.787879 |
| 1.039 | 0.792929 |
| 1.007 | 0.79798 |
| 0.971 | 0.80303 |
| 0.968 | 0.808081 |
| 0.949 | 0.813131 |
| 0.926 | 0.818182 |
| 0.902 | 0.823232 |
| 0.867 | 0.828283 |
| 0.846 | 0.833333 |
| 0.842 | 0.838384 |
| 0.816 | 0.843434 |
| 0.792 | 0.848485 |
| 0.779 | 0.853535 |
| 0.776 | 0.858586 |
| 0.773 | 0.863636 |
| 0.771 | 0.868687 |
| 0.722 | 0.873737 |
| 0.718 | 0.878788 |
| 0.718 | 0.883838 |
| 0.702 | 0.888889 |
| 0.696 | 0.893939 |
| 0.665 | 0.89899 |
| 0.657 | 0.90404 |
| 0.657 | 0.909091 |
| 0.654 | 0.914141 |
| 0.616 | 0.919192 |
| 0.611 | 0.924242 |
| 0.589 | 0.929293 |
| 0.579 | 0.934343 |
| 0.563 | 0.939394 |
| 0.558 | 0.944444 |
| 0.54 | 0.949495 |
| 0.534 | 0.954545 |
| 0.503 | 0.959596 |
| 0.39 | 0.964646 |
| 0.372 | 0.969697 |
| 0.361 | 0.974747 |
| 0.359 | 0.979798 |
| 0.33 | 0.984848 |
| 0.305 | 0.989899 |
| 0.302 | 0.994949 |
| 0.232 | 1 |

|  | ST 18C-BE-PCV14 (Day 300) |
| --- | --- |
| 20.792 | 0.00531915 |
| 19.02 | 0.0106383 |
| 18.155 | 0.0159574 |
| 16.253 | 0.0212766 |
| 15.452 | 0.0265957 |
| 15.149 | 0.0319149 |
| 13.532 | 0.037234 |
| 13.503 | 0.0425532 |
| 12.708 | 0.0478723 |
| 12.666 | 0.0531915 |
| 12.183 | 0.0585106 |
| 11.973 | 0.0638298 |
| 11.644 | 0.0691489 |
| 11.592 | 0.0744681 |
| 11.526 | 0.0797872 |
| 11.427 | 0.0851064 |
| 11.301 | 0.0904255 |
| 11.26 | 0.0957447 |
| 11.252 | 0.101064 |
| 11.197 | 0.106383 |
| 11.159 | 0.111702 |
| 10.801 | 0.117021 |
| 10.655 | 0.12234 |
| 10.602 | 0.12766 |
| 10.317 | 0.132979 |
| 10.274 | 0.138298 |
| 9.793 | 0.143617 |
| 9.352 | 0.148936 |
| 9.087 | 0.154255 |
| 8.789 | 0.159574 |
| 8.341 | 0.164894 |
| 8.236 | 0.170213 |
| 7.93 | 0.175532 |
| 7.761 | 0.180851 |
| 6.187 | 0.18617 |
| 5.284 | 0.191489 |
| 5.262 | 0.196809 |
| 5.172 | 0.202128 |
| 4.967 | 0.207447 |
| 4.931 | 0.212766 |
| 4.793 | 0.218085 |
| 4.766 | 0.223404 |
| 4.718 | 0.228723 |
| 4.591 | 0.234043 |
| 4.588 | 0.239362 |
| 4.5 | 0.244681 |
| 4.472 | 0.25 |
| 4.117 | 0.255319 |
| 4.086 | 0.260638 |
| 3.949 | 0.265957 |
| 3.885 | 0.271277 |
| 3.859 | 0.276596 |
| 3.826 | 0.281915 |
| 3.823 | 0.287234 |
| 3.81 | 0.292553 |
| 3.799 | 0.297872 |
| 3.767 | 0.303191 |
| 3.758 | 0.308511 |
| 3.753 | 0.31383 |
| 3.642 | 0.319149 |
| 3.567 | 0.324468 |
| 3.531 | 0.329787 |
| 3.525 | 0.335106 |
| 3.486 | 0.340426 |
| 3.463 | 0.345745 |
| 3.456 | 0.351064 |
| 3.446 | 0.356383 |
| 3.432 | 0.361702 |
| 3.421 | 0.367021 |
| 3.397 | 0.37234 |
| 3.354 | 0.37766 |
| 3.351 | 0.382979 |
| 3.26 | 0.388298 |
| 3.121 | 0.393617 |
| 3.112 | 0.398936 |
| 3.078 | 0.404255 |
| 3.073 | 0.409574 |
| 3.044 | 0.414894 |
| 3.021 | 0.420213 |
| 3.017 | 0.425532 |
| 2.957 | 0.430851 |
| 2.916 | 0.43617 |
| 2.909 | 0.441489 |
| 2.879 | 0.446809 |
| 2.814 | 0.452128 |
| 2.782 | 0.457447 |
| 2.761 | 0.462766 |
| 2.752 | 0.468085 |
| 2.677 | 0.473404 |
| 2.592 | 0.478723 |
| 2.436 | 0.484043 |
| 2.432 | 0.489362 |
| 2.262 | 0.494681 |
| 2.202 | 0.5 |
| 2.182 | 0.505319 |
| 2.137 | 0.510638 |
| 2.13 | 0.515957 |
| 2.103 | 0.521277 |
| 2.078 | 0.526596 |
| 2.048 | 0.531915 |
| 2.044 | 0.537234 |
| 2.02 | 0.542553 |
| 2.011 | 0.547872 |
| 2.006 | 0.553191 |
| 1.982 | 0.558511 |
| 1.966 | 0.56383 |
| 1.958 | 0.569149 |
| 1.913 | 0.574468 |
| 1.89 | 0.579787 |
| 1.789 | 0.585106 |
| 1.756 | 0.590426 |
| 1.729 | 0.595745 |
| 1.697 | 0.601064 |
| 1.665 | 0.606383 |
| 1.634 | 0.611702 |
| 1.622 | 0.617021 |
| 1.599 | 0.62234 |
| 1.572 | 0.62766 |
| 1.567 | 0.632979 |
| 1.546 | 0.638298 |
| 1.54 | 0.643617 |
| 1.536 | 0.648936 |
| 1.511 | 0.654255 |
| 1.501 | 0.659574 |
| 1.495 | 0.664894 |
| 1.466 | 0.670213 |
| 1.375 | 0.675532 |
| 1.37 | 0.680851 |
| 1.367 | 0.68617 |
| 1.356 | 0.691489 |
| 1.355 | 0.696809 |
| 1.333 | 0.702128 |
| 1.318 | 0.707447 |
| 1.293 | 0.712766 |
| 1.29 | 0.718085 |
| 1.288 | 0.723404 |
| 1.284 | 0.728723 |
| 1.254 | 0.734043 |
| 1.23 | 0.739362 |
| 1.173 | 0.744681 |
| 1.168 | 0.75 |
| 1.148 | 0.755319 |
| 1.128 | 0.760638 |
| 1.104 | 0.765957 |
| 1.074 | 0.771277 |
| 1.005 | 0.776596 |
| 0.989 | 0.781915 |
| 0.935 | 0.787234 |
| 0.826 | 0.792553 |
| 0.751 | 0.797872 |
| 0.699 | 0.803191 |
| 0.665 | 0.808511 |
| 0.595 | 0.81383 |
| 0.563 | 0.819149 |
| 0.541 | 0.824468 |
| 0.541 | 0.829787 |
| 0.539 | 0.835106 |
| 0.507 | 0.840426 |
| 0.48 | 0.845745 |
| 0.448 | 0.851064 |
| 0.431 | 0.856383 |
| 0.414 | 0.861702 |
| 0.409 | 0.867021 |
| 0.409 | 0.87234 |
| 0.408 | 0.87766 |
| 0.368 | 0.882979 |
| 0.365 | 0.888298 |
| 0.362 | 0.893617 |
| 0.349 | 0.898936 |
| 0.287 | 0.904255 |
| 0.258 | 0.909574 |
| 0.232 | 0.914894 |
| 0.223 | 0.920213 |
| 0.222 | 0.925532 |
| 0.22 | 0.930851 |
| 0.212 | 0.93617 |
| 0.209 | 0.941489 |
| 0.207 | 0.946809 |
| 0.204 | 0.952128 |
| 0.192 | 0.957447 |
| 0.191 | 0.962766 |
| 0.189 | 0.968085 |
| 0.181 | 0.973404 |
| 0.167 | 0.978723 |
| 0.089 | 0.984043 |
| 0.087 | 0.989362 |
| 0.085 | 0.994681 |
| 0.082 | 1 |

|  | ST 18C-Prevenar 13 (Day 300) |
| --- | --- |
| 47.416 | 0.00526316 |
| 29.338 | 0.0105263 |
| 29.201 | 0.0157895 |
| 28.651 | 0.0210526 |
| 28.141 | 0.0263158 |
| 27.013 | 0.0315789 |
| 26.02 | 0.0368421 |
| 25.184 | 0.0421053 |
| 22.297 | 0.0473684 |
| 20.523 | 0.0526316 |
| 20.389 | 0.0578947 |
| 20.231 | 0.0631579 |
| 19.784 | 0.0684211 |
| 19.774 | 0.0736842 |
| 19.753 | 0.0789474 |
| 18.64 | 0.0842105 |
| 18.601 | 0.0894737 |
| 17.965 | 0.0947368 |
| 13.793 | 0.1 |
| 12.757 | 0.105263 |
| 12.682 | 0.110526 |
| 11.754 | 0.115789 |
| 9.872 | 0.121053 |
| 9.833 | 0.126316 |
| 9.621 | 0.131579 |
| 8.701 | 0.136842 |
| 8.493 | 0.142105 |
| 7.833 | 0.147368 |
| 7.819 | 0.152632 |
| 7.79 | 0.157895 |
| 6.457 | 0.163158 |
| 5.41 | 0.168421 |
| 5.35 | 0.173684 |
| 4.701 | 0.178947 |
| 4.682 | 0.184211 |
| 4.613 | 0.189474 |
| 4.547 | 0.194737 |
| 4.529 | 0.2 |
| 4.396 | 0.205263 |
| 4.167 | 0.210526 |
| 4.103 | 0.215789 |
| 4.044 | 0.221053 |
| 4.03 | 0.226316 |
| 3.958 | 0.231579 |
| 3.918 | 0.236842 |
| 3.613 | 0.242105 |
| 3.606 | 0.247368 |
| 3.516 | 0.252632 |
| 3.473 | 0.257895 |
| 3.367 | 0.263158 |
| 3.344 | 0.268421 |
| 3.265 | 0.273684 |
| 3.145 | 0.278947 |
| 3.078 | 0.284211 |
| 2.993 | 0.289474 |
| 2.946 | 0.294737 |
| 2.942 | 0.3 |
| 2.877 | 0.305263 |
| 2.719 | 0.310526 |
| 2.712 | 0.315789 |
| 2.691 | 0.321053 |
| 2.688 | 0.326316 |
| 2.651 | 0.331579 |
| 2.639 | 0.336842 |
| 2.607 | 0.342105 |
| 2.585 | 0.347368 |
| 2.539 | 0.352632 |
| 2.51 | 0.357895 |
| 2.451 | 0.363158 |
| 2.451 | 0.368421 |
| 2.413 | 0.373684 |
| 2.371 | 0.378947 |
| 2.343 | 0.384211 |
| 2.338 | 0.389474 |
| 2.236 | 0.394737 |
| 2.223 | 0.4 |
| 2.158 | 0.405263 |
| 2.151 | 0.410526 |
| 2.115 | 0.415789 |
| 2.101 | 0.421053 |
| 2.041 | 0.426316 |
| 2.04 | 0.431579 |
| 1.965 | 0.436842 |
| 1.917 | 0.442105 |
| 1.908 | 0.447368 |
| 1.87 | 0.452632 |
| 1.859 | 0.457895 |
| 1.856 | 0.463158 |
| 1.847 | 0.468421 |
| 1.824 | 0.473684 |
| 1.74 | 0.478947 |
| 1.71 | 0.484211 |
| 1.626 | 0.489474 |
| 1.584 | 0.494737 |
| 1.56 | 0.5 |
| 1.551 | 0.505263 |
| 1.541 | 0.510526 |
| 1.505 | 0.515789 |
| 1.503 | 0.521053 |
| 1.501 | 0.526316 |
| 1.498 | 0.531579 |
| 1.489 | 0.536842 |
| 1.483 | 0.542105 |
| 1.46 | 0.547368 |
| 1.457 | 0.552632 |
| 1.442 | 0.557895 |
| 1.436 | 0.563158 |
| 1.428 | 0.568421 |
| 1.42 | 0.573684 |
| 1.387 | 0.578947 |
| 1.379 | 0.584211 |
| 1.378 | 0.589474 |
| 1.36 | 0.594737 |
| 1.318 | 0.6 |
| 1.31 | 0.605263 |
| 1.276 | 0.610526 |
| 1.274 | 0.615789 |
| 1.221 | 0.621053 |
| 1.208 | 0.626316 |
| 1.196 | 0.631579 |
| 1.169 | 0.636842 |
| 1.159 | 0.642105 |
| 1.152 | 0.647368 |
| 1.137 | 0.652632 |
| 1.124 | 0.657895 |
| 1.121 | 0.663158 |
| 1.106 | 0.668421 |
| 1.084 | 0.673684 |
| 1.064 | 0.678947 |
| 1.046 | 0.684211 |
| 1.037 | 0.689474 |
| 1.011 | 0.694737 |
| 1.004 | 0.7 |
| 1.002 | 0.705263 |
| 0.972 | 0.710526 |
| 0.969 | 0.715789 |
| 0.944 | 0.721053 |
| 0.925 | 0.726316 |
| 0.924 | 0.731579 |
| 0.915 | 0.736842 |
| 0.911 | 0.742105 |
| 0.891 | 0.747368 |
| 0.889 | 0.752632 |
| 0.885 | 0.757895 |
| 0.845 | 0.763158 |
| 0.839 | 0.768421 |
| 0.815 | 0.773684 |
| 0.777 | 0.778947 |
| 0.772 | 0.784211 |
| 0.768 | 0.789474 |
| 0.754 | 0.794737 |
| 0.744 | 0.8 |
| 0.72 | 0.805263 |
| 0.673 | 0.810526 |
| 0.657 | 0.815789 |
| 0.618 | 0.821053 |
| 0.598 | 0.826316 |
| 0.568 | 0.831579 |
| 0.564 | 0.836842 |
| 0.559 | 0.842105 |
| 0.556 | 0.847368 |
| 0.549 | 0.852632 |
| 0.524 | 0.857895 |
| 0.505 | 0.863158 |
| 0.504 | 0.868421 |
| 0.503 | 0.873684 |
| 0.502 | 0.878947 |
| 0.487 | 0.884211 |
| 0.48 | 0.889474 |
| 0.476 | 0.894737 |
| 0.423 | 0.9 |
| 0.414 | 0.905263 |
| 0.411 | 0.910526 |
| 0.369 | 0.915789 |
| 0.331 | 0.921053 |
| 0.308 | 0.926316 |
| 0.226 | 0.931579 |
| 0.186 | 0.936842 |
| 0.183 | 0.942105 |
| 0.171 | 0.947368 |
| 0.158 | 0.952632 |
| 0.132 | 0.957895 |
| 0.121 | 0.963158 |
| 0.112 | 0.968421 |
| 0.102 | 0.973684 |
| 0.086 | 0.978947 |
| 0.085 | 0.984211 |
| 0.077 | 0.989474 |
| 0.022 | 0.994737 |
| 0.022 | 1 |

BECT056 - RCDC_Serotype 19A

|  | ST 19A-BE-PCV14 (Day 84) |
| --- | --- |
| 288.715 | 0.0052356 |
| 182.394 | 0.0104712 |
| 143.969 | 0.0157068 |
| 140.088 | 0.0209424 |
| 136.558 | 0.026178 |
| 91.561 | 0.0314136 |
| 78.086 | 0.0366492 |
| 76.522 | 0.0418848 |
| 51.996 | 0.0471204 |
| 48.227 | 0.052356 |
| 44.675 | 0.0575916 |
| 42.743 | 0.0628272 |
| 40.636 | 0.0680628 |
| 37.428 | 0.0732984 |
| 37.208 | 0.078534 |
| 31.647 | 0.0837696 |
| 28.848 | 0.0890052 |
| 28.39 | 0.0942408 |
| 25.789 | 0.0994764 |
| 25.661 | 0.104712 |
| 24.654 | 0.109948 |
| 22.56 | 0.115183 |
| 22.511 | 0.120419 |
| 21.823 | 0.125654 |
| 21.764 | 0.13089 |
| 20.322 | 0.136126 |
| 20.321 | 0.141361 |
| 20.242 | 0.146597 |
| 18.092 | 0.151832 |
| 17.662 | 0.157068 |
| 14.805 | 0.162304 |
| 14.678 | 0.167539 |
| 13.639 | 0.172775 |
| 12.549 | 0.17801 |
| 12.015 | 0.183246 |
| 11.156 | 0.188482 |
| 9.682 | 0.193717 |
| 8.928 | 0.198953 |
| 8.836 | 0.204188 |
| 8.039 | 0.209424 |
| 7.968 | 0.21466 |
| 7.935 | 0.219895 |
| 7.321 | 0.225131 |
| 7.191 | 0.230366 |
| 7.035 | 0.235602 |
| 6.976 | 0.240838 |
| 6.922 | 0.246073 |
| 6.849 | 0.251309 |
| 6.742 | 0.256545 |
| 6.235 | 0.26178 |
| 6.137 | 0.267016 |
| 5.856 | 0.272251 |
| 5.527 | 0.277487 |
| 5.243 | 0.282723 |
| 5.226 | 0.287958 |
| 4.461 | 0.293194 |
| 4.343 | 0.298429 |
| 4.194 | 0.303665 |
| 4.181 | 0.308901 |
| 4.112 | 0.314136 |
| 3.997 | 0.319372 |
| 3.982 | 0.324607 |
| 3.776 | 0.329843 |
| 3.746 | 0.335079 |
| 3.734 | 0.340314 |
| 3.68 | 0.34555 |
| 3.611 | 0.350785 |
| 3.428 | 0.356021 |
| 3.383 | 0.361257 |
| 3.297 | 0.366492 |
| 3.237 | 0.371728 |
| 3.169 | 0.376963 |
| 3.136 | 0.382199 |
| 3.042 | 0.387435 |
| 2.977 | 0.39267 |
| 2.967 | 0.397906 |
| 2.947 | 0.403141 |
| 2.926 | 0.408377 |
| 2.87 | 0.413613 |
| 2.851 | 0.418848 |
| 2.814 | 0.424084 |
| 2.806 | 0.429319 |
| 2.681 | 0.434555 |
| 2.597 | 0.439791 |
| 2.482 | 0.445026 |
| 2.449 | 0.450262 |
| 2.444 | 0.455497 |
| 2.442 | 0.460733 |
| 2.425 | 0.465969 |
| 2.422 | 0.471204 |
| 2.391 | 0.47644 |
| 2.339 | 0.481675 |
| 2.328 | 0.486911 |
| 2.273 | 0.492147 |
| 2.267 | 0.497382 |
| 2.258 | 0.502618 |
| 2.223 | 0.507853 |
| 2.221 | 0.513089 |
| 2.218 | 0.518325 |
| 2.197 | 0.52356 |
| 2.18 | 0.528796 |
| 2.177 | 0.534031 |
| 2.096 | 0.539267 |
| 2.049 | 0.544503 |
| 2.001 | 0.549738 |
| 1.986 | 0.554974 |
| 1.98 | 0.560209 |
| 1.976 | 0.565445 |
| 1.952 | 0.570681 |
| 1.92 | 0.575916 |
| 1.916 | 0.581152 |
| 1.907 | 0.586387 |
| 1.903 | 0.591623 |
| 1.892 | 0.596859 |
| 1.89 | 0.602094 |
| 1.87 | 0.60733 |
| 1.856 | 0.612565 |
| 1.814 | 0.617801 |
| 1.8 | 0.623037 |
| 1.754 | 0.628272 |
| 1.744 | 0.633508 |
| 1.735 | 0.638743 |
| 1.714 | 0.643979 |
| 1.709 | 0.649215 |
| 1.702 | 0.65445 |
| 1.691 | 0.659686 |
| 1.672 | 0.664921 |
| 1.661 | 0.670157 |
| 1.642 | 0.675393 |
| 1.632 | 0.680628 |
| 1.604 | 0.685864 |
| 1.576 | 0.691099 |
| 1.556 | 0.696335 |
| 1.52 | 0.701571 |
| 1.516 | 0.706806 |
| 1.471 | 0.712042 |
| 1.448 | 0.717277 |
| 1.434 | 0.722513 |
| 1.432 | 0.727749 |
| 1.41 | 0.732984 |
| 1.403 | 0.73822 |
| 1.368 | 0.743455 |
| 1.349 | 0.748691 |
| 1.337 | 0.753927 |
| 1.32 | 0.759162 |
| 1.259 | 0.764398 |
| 1.236 | 0.769634 |
| 1.217 | 0.774869 |
| 1.215 | 0.780105 |
| 1.185 | 0.78534 |
| 1.156 | 0.790576 |
| 1.142 | 0.795812 |
| 1.133 | 0.801047 |
| 1.105 | 0.806283 |
| 1.095 | 0.811518 |
| 1.094 | 0.816754 |
| 1.059 | 0.82199 |
| 1.054 | 0.827225 |
| 1.042 | 0.832461 |
| 1.034 | 0.837696 |
| 1.032 | 0.842932 |
| 1.016 | 0.848168 |
| 0.991 | 0.853403 |
| 0.988 | 0.858639 |
| 0.969 | 0.863874 |
| 0.964 | 0.86911 |
| 0.949 | 0.874346 |
| 0.905 | 0.879581 |
| 0.902 | 0.884817 |
| 0.9 | 0.890052 |
| 0.885 | 0.895288 |
| 0.879 | 0.900524 |
| 0.848 | 0.905759 |
| 0.832 | 0.910995 |
| 0.804 | 0.91623 |
| 0.765 | 0.921466 |
| 0.717 | 0.926702 |
| 0.666 | 0.931937 |
| 0.644 | 0.937173 |
| 0.641 | 0.942408 |
| 0.623 | 0.947644 |
| 0.6 | 0.95288 |
| 0.549 | 0.958115 |
| 0.545 | 0.963351 |
| 0.51 | 0.968586 |
| 0.48 | 0.973822 |
| 0.465 | 0.979058 |
| 0.449 | 0.984293 |
| 0.431 | 0.989529 |
| 0.413 | 0.994764 |
| 0.313 | 1 |

|  | ST 19A-Prevenar 13 (Day 84) |
| --- | --- |
| 255.528 | 0.00505051 |
| 241.293 | 0.010101 |
| 159.24 | 0.0151515 |
| 128.831 | 0.020202 |
| 106.658 | 0.0252525 |
| 103.335 | 0.030303 |
| 59.851 | 0.0353535 |
| 58.668 | 0.040404 |
| 53.722 | 0.0454545 |
| 53.222 | 0.0505051 |
| 48.982 | 0.0555556 |
| 48.273 | 0.0606061 |
| 47.764 | 0.0656566 |
| 45.336 | 0.0707071 |
| 43.586 | 0.0757576 |
| 41.571 | 0.0808081 |
| 40.052 | 0.0858586 |
| 35.058 | 0.0909091 |
| 29.505 | 0.0959596 |
| 22.546 | 0.10101 |
| 21.869 | 0.106061 |
| 20.473 | 0.111111 |
| 20.061 | 0.116162 |
| 19.754 | 0.121212 |
| 19.736 | 0.126263 |
| 18.122 | 0.131313 |
| 17.596 | 0.136364 |
| 16.822 | 0.141414 |
| 16.433 | 0.146465 |
| 16.273 | 0.151515 |
| 15.783 | 0.156566 |
| 15.526 | 0.161616 |
| 14.589 | 0.166667 |
| 14.15 | 0.171717 |
| 13.951 | 0.176768 |
| 13.188 | 0.181818 |
| 11.779 | 0.186869 |
| 11.763 | 0.191919 |
| 11.743 | 0.19697 |
| 10.945 | 0.20202 |
| 10.158 | 0.207071 |
| 9.994 | 0.212121 |
| 9.533 | 0.217172 |
| 8.975 | 0.222222 |
| 8.686 | 0.227273 |
| 7.656 | 0.232323 |
| 7.491 | 0.237374 |
| 7.336 | 0.242424 |
| 7.016 | 0.247475 |
| 6.988 | 0.252525 |
| 6.768 | 0.257576 |
| 6.714 | 0.262626 |
| 6.675 | 0.267677 |
| 6.586 | 0.272727 |
| 6.186 | 0.277778 |
| 6.101 | 0.282828 |
| 6.095 | 0.287879 |
| 6.017 | 0.292929 |
| 5.969 | 0.29798 |
| 5.935 | 0.30303 |
| 5.895 | 0.308081 |
| 5.831 | 0.313131 |
| 5.744 | 0.318182 |
| 5.679 | 0.323232 |
| 5.505 | 0.328283 |
| 5.347 | 0.333333 |
| 5.091 | 0.338384 |
| 4.869 | 0.343434 |
| 4.836 | 0.348485 |
| 4.833 | 0.353535 |
| 4.781 | 0.358586 |
| 4.675 | 0.363636 |
| 4.632 | 0.368687 |
| 4.618 | 0.373737 |
| 4.617 | 0.378788 |
| 4.466 | 0.383838 |
| 4.413 | 0.388889 |
| 4.374 | 0.393939 |
| 4.373 | 0.39899 |
| 4.211 | 0.40404 |
| 4.07 | 0.409091 |
| 4.06 | 0.414141 |
| 3.996 | 0.419192 |
| 3.882 | 0.424242 |
| 3.821 | 0.429293 |
| 3.82 | 0.434343 |
| 3.81 | 0.439394 |
| 3.785 | 0.444444 |
| 3.778 | 0.449495 |
| 3.756 | 0.454545 |
| 3.713 | 0.459596 |
| 3.704 | 0.464646 |
| 3.585 | 0.469697 |
| 3.53 | 0.474747 |
| 3.527 | 0.479798 |
| 3.375 | 0.484848 |
| 3.186 | 0.489899 |
| 3.114 | 0.494949 |
| 3.113 | 0.5 |
| 3.083 | 0.505051 |
| 2.903 | 0.510101 |
| 2.892 | 0.515152 |
| 2.864 | 0.520202 |
| 2.858 | 0.525253 |
| 2.848 | 0.530303 |
| 2.834 | 0.535354 |
| 2.797 | 0.540404 |
| 2.774 | 0.545455 |
| 2.745 | 0.550505 |
| 2.74 | 0.555556 |
| 2.737 | 0.560606 |
| 2.693 | 0.565657 |
| 2.639 | 0.570707 |
| 2.63 | 0.575758 |
| 2.608 | 0.580808 |
| 2.591 | 0.585859 |
| 2.584 | 0.590909 |
| 2.584 | 0.59596 |
| 2.556 | 0.60101 |
| 2.54 | 0.606061 |
| 2.472 | 0.611111 |
| 2.442 | 0.616162 |
| 2.439 | 0.621212 |
| 2.358 | 0.626263 |
| 2.3 | 0.631313 |
| 2.196 | 0.636364 |
| 2.121 | 0.641414 |
| 2.117 | 0.646465 |
| 2.094 | 0.651515 |
| 2.092 | 0.656566 |
| 2.073 | 0.661616 |
| 2.055 | 0.666667 |
| 2.012 | 0.671717 |
| 1.956 | 0.676768 |
| 1.934 | 0.681818 |
| 1.924 | 0.686869 |
| 1.841 | 0.691919 |
| 1.832 | 0.69697 |
| 1.813 | 0.70202 |
| 1.789 | 0.707071 |
| 1.75 | 0.712121 |
| 1.74 | 0.717172 |
| 1.731 | 0.722222 |
| 1.691 | 0.727273 |
| 1.675 | 0.732323 |
| 1.673 | 0.737374 |
| 1.662 | 0.742424 |
| 1.656 | 0.747475 |
| 1.64 | 0.752525 |
| 1.622 | 0.757576 |
| 1.591 | 0.762626 |
| 1.585 | 0.767677 |
| 1.58 | 0.772727 |
| 1.565 | 0.777778 |
| 1.549 | 0.782828 |
| 1.548 | 0.787879 |
| 1.523 | 0.792929 |
| 1.46 | 0.79798 |
| 1.443 | 0.80303 |
| 1.441 | 0.808081 |
| 1.42 | 0.813131 |
| 1.372 | 0.818182 |
| 1.354 | 0.823232 |
| 1.312 | 0.828283 |
| 1.288 | 0.833333 |
| 1.274 | 0.838384 |
| 1.253 | 0.843434 |
| 1.249 | 0.848485 |
| 1.228 | 0.853535 |
| 1.209 | 0.858586 |
| 1.173 | 0.863636 |
| 1.149 | 0.868687 |
| 1.109 | 0.873737 |
| 1.091 | 0.878788 |
| 1.08 | 0.883838 |
| 1.059 | 0.888889 |
| 1.005 | 0.893939 |
| 0.997 | 0.89899 |
| 0.996 | 0.90404 |
| 0.968 | 0.909091 |
| 0.907 | 0.914141 |
| 0.906 | 0.919192 |
| 0.899 | 0.924242 |
| 0.858 | 0.929293 |
| 0.851 | 0.934343 |
| 0.803 | 0.939394 |
| 0.796 | 0.944444 |
| 0.784 | 0.949495 |
| 0.769 | 0.954545 |
| 0.731 | 0.959596 |
| 0.719 | 0.964646 |
| 0.63 | 0.969697 |
| 0.621 | 0.974747 |
| 0.607 | 0.979798 |
| 0.595 | 0.984848 |
| 0.557 | 0.989899 |
| 0.53 | 0.994949 |
| 0.504 | 1 |

|  | ST 19A-BE-PCV14 (Day 300) |
| --- | --- |
| 108.203 | 0.00531915 |
| 101.792 | 0.0106383 |
| 92.635 | 0.0159574 |
| 75.51 | 0.0212766 |
| 74.727 | 0.0265957 |
| 72.222 | 0.0319149 |
| 71.153 | 0.037234 |
| 62.075 | 0.0425532 |
| 52.316 | 0.0478723 |
| 44.428 | 0.0531915 |
| 41.111 | 0.0585106 |
| 40.765 | 0.0638298 |
| 40.627 | 0.0691489 |
| 40.404 | 0.0744681 |
| 39.805 | 0.0797872 |
| 39.609 | 0.0851064 |
| 39.51 | 0.0904255 |
| 34.769 | 0.0957447 |
| 33.833 | 0.101064 |
| 33.632 | 0.106383 |
| 32.747 | 0.111702 |
| 31.915 | 0.117021 |
| 31.152 | 0.12234 |
| 29.918 | 0.12766 |
| 29.392 | 0.132979 |
| 28.503 | 0.138298 |
| 25.642 | 0.143617 |
| 24.187 | 0.148936 |
| 23.762 | 0.154255 |
| 22.184 | 0.159574 |
| 21.871 | 0.164894 |
| 21.391 | 0.170213 |
| 21.382 | 0.175532 |
| 20.761 | 0.180851 |
| 20.722 | 0.18617 |
| 20.682 | 0.191489 |
| 20.519 | 0.196809 |
| 19.942 | 0.202128 |
| 19.541 | 0.207447 |
| 18.117 | 0.212766 |
| 17.364 | 0.218085 |
| 17.361 | 0.223404 |
| 17.126 | 0.228723 |
| 16.979 | 0.234043 |
| 16.876 | 0.239362 |
| 16.406 | 0.244681 |
| 16.376 | 0.25 |
| 15.944 | 0.255319 |
| 15.812 | 0.260638 |
| 15.166 | 0.265957 |
| 15.142 | 0.271277 |
| 15.07 | 0.276596 |
| 15.017 | 0.281915 |
| 14.728 | 0.287234 |
| 14.534 | 0.292553 |
| 14.478 | 0.297872 |
| 14.451 | 0.303191 |
| 14.117 | 0.308511 |
| 13.325 | 0.31383 |
| 12.801 | 0.319149 |
| 12.705 | 0.324468 |
| 12.265 | 0.329787 |
| 12.2 | 0.335106 |
| 12.063 | 0.340426 |
| 11.653 | 0.345745 |
| 11.174 | 0.351064 |
| 11.151 | 0.356383 |
| 11.045 | 0.361702 |
| 10.022 | 0.367021 |
| 9.293 | 0.37234 |
| 9.15 | 0.37766 |
| 9.064 | 0.382979 |
| 9.039 | 0.388298 |
| 9.028 | 0.393617 |
| 8.654 | 0.398936 |
| 8.511 | 0.404255 |
| 8.211 | 0.409574 |
| 8.02 | 0.414894 |
| 8.005 | 0.420213 |
| 7.982 | 0.425532 |
| 7.935 | 0.430851 |
| 7.662 | 0.43617 |
| 7.576 | 0.441489 |
| 7.493 | 0.446809 |
| 7.457 | 0.452128 |
| 7.304 | 0.457447 |
| 7.265 | 0.462766 |
| 7.185 | 0.468085 |
| 7.075 | 0.473404 |
| 6.895 | 0.478723 |
| 6.86 | 0.484043 |
| 6.805 | 0.489362 |
| 6.701 | 0.494681 |
| 6.687 | 0.5 |
| 6.68 | 0.505319 |
| 6.519 | 0.510638 |
| 6.488 | 0.515957 |
| 6.402 | 0.521277 |
| 6.3 | 0.526596 |
| 6.203 | 0.531915 |
| 5.927 | 0.537234 |
| 5.681 | 0.542553 |
| 5.57 | 0.547872 |
| 5.547 | 0.553191 |
| 5.321 | 0.558511 |
| 5.301 | 0.56383 |
| 5.284 | 0.569149 |
| 5.181 | 0.574468 |
| 5.177 | 0.579787 |
| 5.037 | 0.585106 |
| 4.994 | 0.590426 |
| 4.857 | 0.595745 |
| 4.806 | 0.601064 |
| 4.78 | 0.606383 |
| 4.66 | 0.611702 |
| 4.65 | 0.617021 |
| 4.591 | 0.62234 |
| 4.453 | 0.62766 |
| 4.375 | 0.632979 |
| 4.243 | 0.638298 |
| 4.22 | 0.643617 |
| 4.117 | 0.648936 |
| 4.039 | 0.654255 |
| 3.971 | 0.659574 |
| 3.939 | 0.664894 |
| 3.824 | 0.670213 |
| 3.82 | 0.675532 |
| 3.767 | 0.680851 |
| 3.761 | 0.68617 |
| 3.727 | 0.691489 |
| 3.715 | 0.696809 |
| 3.706 | 0.702128 |
| 3.699 | 0.707447 |
| 3.667 | 0.712766 |
| 3.665 | 0.718085 |
| 3.422 | 0.723404 |
| 3.396 | 0.728723 |
| 3.216 | 0.734043 |
| 3.045 | 0.739362 |
| 2.997 | 0.744681 |
| 2.879 | 0.75 |
| 2.866 | 0.755319 |
| 2.855 | 0.760638 |
| 2.849 | 0.765957 |
| 2.837 | 0.771277 |
| 2.738 | 0.776596 |
| 2.665 | 0.781915 |
| 2.578 | 0.787234 |
| 2.546 | 0.792553 |
| 2.501 | 0.797872 |
| 2.464 | 0.803191 |
| 2.457 | 0.808511 |
| 2.377 | 0.81383 |
| 2.369 | 0.819149 |
| 2.338 | 0.824468 |
| 2.328 | 0.829787 |
| 2.319 | 0.835106 |
| 2.297 | 0.840426 |
| 2.294 | 0.845745 |
| 2.28 | 0.851064 |
| 2.251 | 0.856383 |
| 2.188 | 0.861702 |
| 2.18 | 0.867021 |
| 2.154 | 0.87234 |
| 2.148 | 0.87766 |
| 2.143 | 0.882979 |
| 2.098 | 0.888298 |
| 2.092 | 0.893617 |
| 2.081 | 0.898936 |
| 2.066 | 0.904255 |
| 1.951 | 0.909574 |
| 1.928 | 0.914894 |
| 1.9 | 0.920213 |
| 1.859 | 0.925532 |
| 1.846 | 0.930851 |
| 1.521 | 0.93617 |
| 1.187 | 0.941489 |
| 1.126 | 0.946809 |
| 1.073 | 0.952128 |
| 1.061 | 0.957447 |
| 0.767 | 0.962766 |
| 0.68 | 0.968085 |
| 0.637 | 0.973404 |
| 0.585 | 0.978723 |
| 0.417 | 0.984043 |
| 0.408 | 0.989362 |
| 0.327 | 0.994681 |
| 0.244 | 1 |

|  | ST 19A-Prevenar 13 (Day 300) |
| --- | --- |
| 142.963 | 0.00526316 |
| 114.225 | 0.0105263 |
| 100.164 | 0.0157895 |
| 99.973 | 0.0210526 |
| 97.394 | 0.0263158 |
| 69.566 | 0.0315789 |
| 68.635 | 0.0368421 |
| 61.344 | 0.0421053 |
| 58.4 | 0.0473684 |
| 49.797 | 0.0526316 |
| 48.685 | 0.0578947 |
| 48.179 | 0.0631579 |
| 45.751 | 0.0684211 |
| 45.21 | 0.0736842 |
| 45.033 | 0.0789474 |
| 44.625 | 0.0842105 |
| 43.882 | 0.0894737 |
| 43.283 | 0.0947368 |
| 42.608 | 0.1 |
| 41.01 | 0.105263 |
| 40.287 | 0.110526 |
| 38.652 | 0.115789 |
| 35.978 | 0.121053 |
| 32.995 | 0.126316 |
| 32.804 | 0.131579 |
| 30.797 | 0.136842 |
| 30.114 | 0.142105 |
| 27.733 | 0.147368 |
| 27.165 | 0.152632 |
| 25.767 | 0.157895 |
| 24.93 | 0.163158 |
| 24.214 | 0.168421 |
| 23.684 | 0.173684 |
| 23.665 | 0.178947 |
| 22.815 | 0.184211 |
| 22.554 | 0.189474 |
| 21.249 | 0.194737 |
| 20.629 | 0.2 |
| 20.517 | 0.205263 |
| 17.743 | 0.210526 |
| 17.619 | 0.215789 |
| 17.069 | 0.221053 |
| 16.946 | 0.226316 |
| 16.911 | 0.231579 |
| 16.874 | 0.236842 |
| 16.422 | 0.242105 |
| 16.35 | 0.247368 |
| 15.485 | 0.252632 |
| 15.473 | 0.257895 |
| 14.729 | 0.263158 |
| 14.37 | 0.268421 |
| 14.315 | 0.273684 |
| 14.284 | 0.278947 |
| 14.216 | 0.284211 |
| 13.763 | 0.289474 |
| 13.251 | 0.294737 |
| 12.947 | 0.3 |
| 12.94 | 0.305263 |
| 12.805 | 0.310526 |
| 12.375 | 0.315789 |
| 12.203 | 0.321053 |
| 11.22 | 0.326316 |
| 11.018 | 0.331579 |
| 10.929 | 0.336842 |
| 10.456 | 0.342105 |
| 10.372 | 0.347368 |
| 10.302 | 0.352632 |
| 10.28 | 0.357895 |
| 9.787 | 0.363158 |
| 9.531 | 0.368421 |
| 9.5 | 0.373684 |
| 9.475 | 0.378947 |
| 9.391 | 0.384211 |
| 9.285 | 0.389474 |
| 9.184 | 0.394737 |
| 9.179 | 0.4 |
| 8.953 | 0.405263 |
| 8.891 | 0.410526 |
| 8.851 | 0.415789 |
| 8.698 | 0.421053 |
| 8.686 | 0.426316 |
| 8.607 | 0.431579 |
| 8.604 | 0.436842 |
| 8.562 | 0.442105 |
| 8.545 | 0.447368 |
| 8.432 | 0.452632 |
| 8.316 | 0.457895 |
| 8.282 | 0.463158 |
| 8.275 | 0.468421 |
| 8.164 | 0.473684 |
| 7.916 | 0.478947 |
| 7.908 | 0.484211 |
| 7.885 | 0.489474 |
| 7.848 | 0.494737 |
| 7.785 | 0.5 |
| 7.703 | 0.505263 |
| 7.586 | 0.510526 |
| 7.571 | 0.515789 |
| 7.07 | 0.521053 |
| 6.742 | 0.526316 |
| 6.696 | 0.531579 |
| 6.635 | 0.536842 |
| 6.557 | 0.542105 |
| 6.5 | 0.547368 |
| 6.446 | 0.552632 |
| 6.162 | 0.557895 |
| 6.145 | 0.563158 |
| 6.083 | 0.568421 |
| 5.893 | 0.573684 |
| 5.89 | 0.578947 |
| 5.823 | 0.584211 |
| 5.695 | 0.589474 |
| 5.563 | 0.594737 |
| 5.187 | 0.6 |
| 5.08 | 0.605263 |
| 4.961 | 0.610526 |
| 4.96 | 0.615789 |
| 4.916 | 0.621053 |
| 4.877 | 0.626316 |
| 4.852 | 0.631579 |
| 4.829 | 0.636842 |
| 4.663 | 0.642105 |
| 4.583 | 0.647368 |
| 4.507 | 0.652632 |
| 4.506 | 0.657895 |
| 4.34 | 0.663158 |
| 4.218 | 0.668421 |
| 4.178 | 0.673684 |
| 4.101 | 0.678947 |
| 3.907 | 0.684211 |
| 3.905 | 0.689474 |
| 3.819 | 0.694737 |
| 3.818 | 0.7 |
| 3.762 | 0.705263 |
| 3.669 | 0.710526 |
| 3.656 | 0.715789 |
| 3.649 | 0.721053 |
| 3.597 | 0.726316 |
| 3.594 | 0.731579 |
| 3.551 | 0.736842 |
| 3.5 | 0.742105 |
| 3.452 | 0.747368 |
| 3.43 | 0.752632 |
| 3.369 | 0.757895 |
| 3.366 | 0.763158 |
| 3.355 | 0.768421 |
| 3.309 | 0.773684 |
| 3.303 | 0.778947 |
| 3.169 | 0.784211 |
| 3.059 | 0.789474 |
| 3.032 | 0.794737 |
| 3.032 | 0.8 |
| 2.893 | 0.805263 |
| 2.884 | 0.810526 |
| 2.87 | 0.815789 |
| 2.84 | 0.821053 |
| 2.779 | 0.826316 |
| 2.683 | 0.831579 |
| 2.37 | 0.836842 |
| 2.306 | 0.842105 |
| 2.21 | 0.847368 |
| 2.099 | 0.852632 |
| 2.088 | 0.857895 |
| 2.067 | 0.863158 |
| 1.952 | 0.868421 |
| 1.948 | 0.873684 |
| 1.853 | 0.878947 |
| 1.85 | 0.884211 |
| 1.841 | 0.889474 |
| 1.781 | 0.894737 |
| 1.631 | 0.9 |
| 1.613 | 0.905263 |
| 1.562 | 0.910526 |
| 1.458 | 0.915789 |
| 1.45 | 0.921053 |
| 1.342 | 0.926316 |
| 1.263 | 0.931579 |
| 1.151 | 0.936842 |
| 1.125 | 0.942105 |
| 1.121 | 0.947368 |
| 1.116 | 0.952632 |
| 0.998 | 0.957895 |
| 0.954 | 0.963158 |
| 0.705 | 0.968421 |
| 0.665 | 0.973684 |
| 0.628 | 0.978947 |
| 0.516 | 0.984211 |
| 0.374 | 0.989474 |
| 0.324 | 0.994737 |
| 0.32 | 1 |

BECT056 - RCDC_Serotype 19F

|  | ST 19F-BE-PCV14 (Day 84) |
| --- | --- |
| 147.562 | 0.0052356 |
| 126.856 | 0.0104712 |
| 117.525 | 0.0157068 |
| 113.806 | 0.0209424 |
| 112.415 | 0.026178 |
| 98.2 | 0.0314136 |
| 88.704 | 0.0366492 |
| 64.413 | 0.0418848 |
| 59.878 | 0.0471204 |
| 55.631 | 0.052356 |
| 49.536 | 0.0575916 |
| 49.037 | 0.0628272 |
| 45.174 | 0.0680628 |
| 44.849 | 0.0732984 |
| 44.291 | 0.078534 |
| 44.074 | 0.0837696 |
| 41.809 | 0.0890052 |
| 37.994 | 0.0942408 |
| 36.353 | 0.0994764 |
| 30.365 | 0.104712 |
| 30.298 | 0.109948 |
| 30.222 | 0.115183 |
| 26.321 | 0.120419 |
| 26.022 | 0.125654 |
| 23.811 | 0.13089 |
| 23.331 | 0.136126 |
| 23.093 | 0.141361 |
| 23.01 | 0.146597 |
| 22.795 | 0.151832 |
| 21.704 | 0.157068 |
| 21.221 | 0.162304 |
| 18.475 | 0.167539 |
| 17.601 | 0.172775 |
| 17.339 | 0.17801 |
| 16.888 | 0.183246 |
| 16.526 | 0.188482 |
| 15.755 | 0.193717 |
| 15.45 | 0.198953 |
| 15.105 | 0.204188 |
| 14.821 | 0.209424 |
| 14.238 | 0.21466 |
| 13.154 | 0.219895 |
| 13.001 | 0.225131 |
| 12.979 | 0.230366 |
| 12.77 | 0.235602 |
| 12.487 | 0.240838 |
| 11.641 | 0.246073 |
| 11.63 | 0.251309 |
| 11.448 | 0.256545 |
| 10.974 | 0.26178 |
| 10.824 | 0.267016 |
| 10.693 | 0.272251 |
| 10.519 | 0.277487 |
| 10.289 | 0.282723 |
| 10.006 | 0.287958 |
| 9.627 | 0.293194 |
| 9.302 | 0.298429 |
| 9.078 | 0.303665 |
| 9.067 | 0.308901 |
| 8.951 | 0.314136 |
| 8.899 | 0.319372 |
| 8.894 | 0.324607 |
| 8.874 | 0.329843 |
| 8.642 | 0.335079 |
| 8.59 | 0.340314 |
| 8.107 | 0.34555 |
| 8.058 | 0.350785 |
| 8.043 | 0.356021 |
| 8.009 | 0.361257 |
| 7.846 | 0.366492 |
| 7.799 | 0.371728 |
| 7.796 | 0.376963 |
| 7.659 | 0.382199 |
| 7.642 | 0.387435 |
| 6.837 | 0.39267 |
| 6.781 | 0.397906 |
| 6.629 | 0.403141 |
| 6.55 | 0.408377 |
| 6.538 | 0.413613 |
| 6.412 | 0.418848 |
| 6.322 | 0.424084 |
| 6.119 | 0.429319 |
| 6.111 | 0.434555 |
| 6.109 | 0.439791 |
| 6.037 | 0.445026 |
| 6.001 | 0.450262 |
| 5.954 | 0.455497 |
| 5.95 | 0.460733 |
| 5.942 | 0.465969 |
| 5.885 | 0.471204 |
| 5.832 | 0.47644 |
| 5.696 | 0.481675 |
| 5.62 | 0.486911 |
| 5.429 | 0.492147 |
| 5.413 | 0.497382 |
| 5.412 | 0.502618 |
| 5.286 | 0.507853 |
| 5.266 | 0.513089 |
| 5.127 | 0.518325 |
| 5.07 | 0.52356 |
| 5.068 | 0.528796 |
| 5.023 | 0.534031 |
| 4.942 | 0.539267 |
| 4.836 | 0.544503 |
| 4.641 | 0.549738 |
| 4.636 | 0.554974 |
| 4.614 | 0.560209 |
| 4.544 | 0.565445 |
| 4.531 | 0.570681 |
| 4.473 | 0.575916 |
| 4.426 | 0.581152 |
| 4.31 | 0.586387 |
| 4.275 | 0.591623 |
| 4.206 | 0.596859 |
| 4.137 | 0.602094 |
| 4.02 | 0.60733 |
| 3.979 | 0.612565 |
| 3.951 | 0.617801 |
| 3.862 | 0.623037 |
| 3.732 | 0.628272 |
| 3.713 | 0.633508 |
| 3.549 | 0.638743 |
| 3.459 | 0.643979 |
| 3.396 | 0.649215 |
| 3.347 | 0.65445 |
| 3.229 | 0.659686 |
| 3.221 | 0.664921 |
| 3.098 | 0.670157 |
| 3.093 | 0.675393 |
| 3.092 | 0.680628 |
| 3.084 | 0.685864 |
| 3.078 | 0.691099 |
| 3.072 | 0.696335 |
| 3.053 | 0.701571 |
| 3.039 | 0.706806 |
| 3.036 | 0.712042 |
| 2.98 | 0.717277 |
| 2.92 | 0.722513 |
| 2.903 | 0.727749 |
| 2.893 | 0.732984 |
| 2.873 | 0.73822 |
| 2.799 | 0.743455 |
| 2.663 | 0.748691 |
| 2.628 | 0.753927 |
| 2.622 | 0.759162 |
| 2.572 | 0.764398 |
| 2.484 | 0.769634 |
| 2.407 | 0.774869 |
| 2.292 | 0.780105 |
| 2.239 | 0.78534 |
| 2.177 | 0.790576 |
| 2.145 | 0.795812 |
| 2.1 | 0.801047 |
| 2.099 | 0.806283 |
| 2.065 | 0.811518 |
| 2.022 | 0.816754 |
| 1.97 | 0.82199 |
| 1.95 | 0.827225 |
| 1.919 | 0.832461 |
| 1.801 | 0.837696 |
| 1.716 | 0.842932 |
| 1.664 | 0.848168 |
| 1.649 | 0.853403 |
| 1.609 | 0.858639 |
| 1.607 | 0.863874 |
| 1.516 | 0.86911 |
| 1.495 | 0.874346 |
| 1.46 | 0.879581 |
| 1.448 | 0.884817 |
| 1.446 | 0.890052 |
| 1.299 | 0.895288 |
| 1.271 | 0.900524 |
| 1.25 | 0.905759 |
| 1.215 | 0.910995 |
| 1.181 | 0.91623 |
| 1.157 | 0.921466 |
| 1.124 | 0.926702 |
| 1.077 | 0.931937 |
| 1.012 | 0.937173 |
| 0.936 | 0.942408 |
| 0.909 | 0.947644 |
| 0.908 | 0.95288 |
| 0.872 | 0.958115 |
| 0.858 | 0.963351 |
| 0.798 | 0.968586 |
| 0.773 | 0.973822 |
| 0.719 | 0.979058 |
| 0.716 | 0.984293 |
| 0.637 | 0.989529 |
| 0.568 | 0.994764 |
| 0.458 | 1 |

|  | ST 19F-Prevenar 13 (Day 84) |
| --- | --- |
| 165.122 | 0.00505051 |
| 107.419 | 0.010101 |
| 89.93 | 0.0151515 |
| 66.445 | 0.020202 |
| 60.447 | 0.0252525 |
| 52.473 | 0.030303 |
| 49.811 | 0.0353535 |
| 46.085 | 0.040404 |
| 40.65 | 0.0454545 |
| 40.498 | 0.0505051 |
| 35.059 | 0.0555556 |
| 31.935 | 0.0606061 |
| 31.709 | 0.0656566 |
| 31.201 | 0.0707071 |
| 29.989 | 0.0757576 |
| 29.503 | 0.0808081 |
| 28.905 | 0.0858586 |
| 26.975 | 0.0909091 |
| 25.617 | 0.0959596 |
| 25.261 | 0.10101 |
| 24.805 | 0.106061 |
| 23.996 | 0.111111 |
| 23.551 | 0.116162 |
| 23.468 | 0.121212 |
| 22.657 | 0.126263 |
| 22.389 | 0.131313 |
| 22.343 | 0.136364 |
| 21.764 | 0.141414 |
| 21.447 | 0.146465 |
| 21.231 | 0.151515 |
| 20.757 | 0.156566 |
| 20.186 | 0.161616 |
| 18.755 | 0.166667 |
| 17.673 | 0.171717 |
| 17.633 | 0.176768 |
| 17.625 | 0.181818 |
| 16.65 | 0.186869 |
| 16.623 | 0.191919 |
| 15.819 | 0.19697 |
| 15.485 | 0.20202 |
| 15.454 | 0.207071 |
| 15.337 | 0.212121 |
| 15.26 | 0.217172 |
| 15.145 | 0.222222 |
| 14.992 | 0.227273 |
| 14.683 | 0.232323 |
| 14.648 | 0.237374 |
| 13.221 | 0.242424 |
| 13.02 | 0.247475 |
| 12.98 | 0.252525 |
| 12.903 | 0.257576 |
| 12.784 | 0.262626 |
| 12.7 | 0.267677 |
| 12.373 | 0.272727 |
| 12.299 | 0.277778 |
| 12.211 | 0.282828 |
| 11.81 | 0.287879 |
| 11.727 | 0.292929 |
| 11.64 | 0.29798 |
| 11.37 | 0.30303 |
| 10.961 | 0.308081 |
| 10.829 | 0.313131 |
| 10.791 | 0.318182 |
| 10.752 | 0.323232 |
| 10.496 | 0.328283 |
| 10.45 | 0.333333 |
| 10.445 | 0.338384 |
| 10.433 | 0.343434 |
| 10.365 | 0.348485 |
| 10.339 | 0.353535 |
| 10.311 | 0.358586 |
| 10.292 | 0.363636 |
| 10.192 | 0.368687 |
| 9.989 | 0.373737 |
| 9.823 | 0.378788 |
| 9.478 | 0.383838 |
| 9.448 | 0.388889 |
| 9.405 | 0.393939 |
| 9.321 | 0.39899 |
| 8.921 | 0.40404 |
| 8.836 | 0.409091 |
| 8.831 | 0.414141 |
| 8.811 | 0.419192 |
| 8.68 | 0.424242 |
| 8.646 | 0.429293 |
| 8.631 | 0.434343 |
| 8.497 | 0.439394 |
| 8.305 | 0.444444 |
| 8.277 | 0.449495 |
| 7.801 | 0.454545 |
| 7.762 | 0.459596 |
| 7.656 | 0.464646 |
| 7.655 | 0.469697 |
| 7.585 | 0.474747 |
| 7.514 | 0.479798 |
| 7.445 | 0.484848 |
| 7.434 | 0.489899 |
| 7.431 | 0.494949 |
| 7.065 | 0.5 |
| 6.838 | 0.505051 |
| 6.59 | 0.510101 |
| 6.541 | 0.515152 |
| 6.502 | 0.520202 |
| 6.428 | 0.525253 |
| 6.349 | 0.530303 |
| 6.273 | 0.535354 |
| 6.247 | 0.540404 |
| 6.139 | 0.545455 |
| 6.082 | 0.550505 |
| 5.99 | 0.555556 |
| 5.9 | 0.560606 |
| 5.808 | 0.565657 |
| 5.765 | 0.570707 |
| 5.739 | 0.575758 |
| 5.703 | 0.580808 |
| 5.68 | 0.585859 |
| 5.57 | 0.590909 |
| 5.534 | 0.59596 |
| 5.528 | 0.60101 |
| 5.498 | 0.606061 |
| 5.474 | 0.611111 |
| 5.403 | 0.616162 |
| 5.339 | 0.621212 |
| 5.294 | 0.626263 |
| 5.286 | 0.631313 |
| 5.27 | 0.636364 |
| 5.172 | 0.641414 |
| 5 | 0.646465 |
| 4.827 | 0.651515 |
| 4.792 | 0.656566 |
| 4.741 | 0.661616 |
| 4.685 | 0.666667 |
| 4.55 | 0.671717 |
| 4.412 | 0.676768 |
| 4.378 | 0.681818 |
| 4.245 | 0.686869 |
| 4.192 | 0.691919 |
| 4.113 | 0.69697 |
| 4.032 | 0.70202 |
| 3.919 | 0.707071 |
| 3.89 | 0.712121 |
| 3.876 | 0.717172 |
| 3.87 | 0.722222 |
| 3.865 | 0.727273 |
| 3.627 | 0.732323 |
| 3.619 | 0.737374 |
| 3.587 | 0.742424 |
| 3.583 | 0.747475 |
| 3.553 | 0.752525 |
| 3.52 | 0.757576 |
| 3.481 | 0.762626 |
| 3.373 | 0.767677 |
| 3.24 | 0.772727 |
| 3.207 | 0.777778 |
| 3.157 | 0.782828 |
| 3.12 | 0.787879 |
| 3.12 | 0.792929 |
| 3.117 | 0.79798 |
| 3.048 | 0.80303 |
| 3.042 | 0.808081 |
| 2.937 | 0.813131 |
| 2.919 | 0.818182 |
| 2.867 | 0.823232 |
| 2.84 | 0.828283 |
| 2.782 | 0.833333 |
| 2.51 | 0.838384 |
| 2.38 | 0.843434 |
| 2.357 | 0.848485 |
| 2.156 | 0.853535 |
| 2.117 | 0.858586 |
| 2.01 | 0.863636 |
| 2 | 0.868687 |
| 1.958 | 0.873737 |
| 1.866 | 0.878788 |
| 1.842 | 0.883838 |
| 1.791 | 0.888889 |
| 1.774 | 0.893939 |
| 1.718 | 0.89899 |
| 1.647 | 0.90404 |
| 1.617 | 0.909091 |
| 1.567 | 0.914141 |
| 1.549 | 0.919192 |
| 1.49 | 0.924242 |
| 1.43 | 0.929293 |
| 1.339 | 0.934343 |
| 1.338 | 0.939394 |
| 1.288 | 0.944444 |
| 1.288 | 0.949495 |
| 0.954 | 0.954545 |
| 0.827 | 0.959596 |
| 0.786 | 0.964646 |
| 0.747 | 0.969697 |
| 0.744 | 0.974747 |
| 0.715 | 0.979798 |
| 0.713 | 0.984848 |
| 0.669 | 0.989899 |
| 0.638 | 0.994949 |
| 0.629 | 1 |

|  | ST 19F-BE-PCV14 (Day 300) |
| --- | --- |
| 100.882 | 0.00531915 |
| 75.928 | 0.0106383 |
| 69.718 | 0.0159574 |
| 62.843 | 0.0212766 |
| 62.435 | 0.0265957 |
| 62.072 | 0.0319149 |
| 57.707 | 0.037234 |
| 52.422 | 0.0425532 |
| 49.427 | 0.0478723 |
| 45.885 | 0.0531915 |
| 45.851 | 0.0585106 |
| 45.129 | 0.0638298 |
| 43.478 | 0.0691489 |
| 43.264 | 0.0744681 |
| 43.155 | 0.0797872 |
| 42.888 | 0.0851064 |
| 42.77 | 0.0904255 |
| 42.149 | 0.0957447 |
| 41.616 | 0.101064 |
| 41.542 | 0.106383 |
| 40.762 | 0.111702 |
| 40.469 | 0.117021 |
| 39.855 | 0.12234 |
| 39.537 | 0.12766 |
| 39.496 | 0.132979 |
| 39.233 | 0.138298 |
| 38.853 | 0.143617 |
| 37.293 | 0.148936 |
| 37.293 | 0.154255 |
| 36.804 | 0.159574 |
| 36.385 | 0.164894 |
| 35.446 | 0.170213 |
| 34.577 | 0.175532 |
| 33.769 | 0.180851 |
| 30.086 | 0.18617 |
| 30.023 | 0.191489 |
| 28.72 | 0.196809 |
| 27.335 | 0.202128 |
| 26.995 | 0.207447 |
| 25.852 | 0.212766 |
| 25.544 | 0.218085 |
| 25.421 | 0.223404 |
| 25.327 | 0.228723 |
| 24.516 | 0.234043 |
| 24.138 | 0.239362 |
| 23.15 | 0.244681 |
| 22.385 | 0.25 |
| 21.448 | 0.255319 |
| 21.028 | 0.260638 |
| 20.885 | 0.265957 |
| 20.706 | 0.271277 |
| 20.641 | 0.276596 |
| 20.536 | 0.281915 |
| 20.482 | 0.287234 |
| 20.25 | 0.292553 |
| 20.007 | 0.297872 |
| 19.91 | 0.303191 |
| 19.878 | 0.308511 |
| 19.838 | 0.31383 |
| 19.176 | 0.319149 |
| 18.878 | 0.324468 |
| 18.159 | 0.329787 |
| 17.58 | 0.335106 |
| 17.527 | 0.340426 |
| 17.512 | 0.345745 |
| 17.453 | 0.351064 |
| 17.297 | 0.356383 |
| 17.14 | 0.361702 |
| 16.749 | 0.367021 |
| 16.384 | 0.37234 |
| 16.079 | 0.37766 |
| 15.669 | 0.382979 |
| 15.067 | 0.388298 |
| 14.778 | 0.393617 |
| 14.534 | 0.398936 |
| 14.527 | 0.404255 |
| 14.319 | 0.409574 |
| 14.299 | 0.414894 |
| 13.964 | 0.420213 |
| 13.952 | 0.425532 |
| 13.867 | 0.430851 |
| 13.244 | 0.43617 |
| 13.133 | 0.441489 |
| 12.965 | 0.446809 |
| 12.537 | 0.452128 |
| 12.35 | 0.457447 |
| 12.291 | 0.462766 |
| 12.097 | 0.468085 |
| 11.993 | 0.473404 |
| 11.989 | 0.478723 |
| 11.817 | 0.484043 |
| 11.138 | 0.489362 |
| 11.105 | 0.494681 |
| 11.094 | 0.5 |
| 10.828 | 0.505319 |
| 10.761 | 0.510638 |
| 10.701 | 0.515957 |
| 10.694 | 0.521277 |
| 10.158 | 0.526596 |
| 9.91 | 0.531915 |
| 9.578 | 0.537234 |
| 9.548 | 0.542553 |
| 9.342 | 0.547872 |
| 9.291 | 0.553191 |
| 9.253 | 0.558511 |
| 8.992 | 0.56383 |
| 8.917 | 0.569149 |
| 8.398 | 0.574468 |
| 8.231 | 0.579787 |
| 8.176 | 0.585106 |
| 8.127 | 0.590426 |
| 8.077 | 0.595745 |
| 7.657 | 0.601064 |
| 7.527 | 0.606383 |
| 7.504 | 0.611702 |
| 7.475 | 0.617021 |
| 7.469 | 0.62234 |
| 7.312 | 0.62766 |
| 7.277 | 0.632979 |
| 7.122 | 0.638298 |
| 6.912 | 0.643617 |
| 6.591 | 0.648936 |
| 6.492 | 0.654255 |
| 6.457 | 0.659574 |
| 6.27 | 0.664894 |
| 6.176 | 0.670213 |
| 5.992 | 0.675532 |
| 5.947 | 0.680851 |
| 5.815 | 0.68617 |
| 5.739 | 0.691489 |
| 5.531 | 0.696809 |
| 5.369 | 0.702128 |
| 5.357 | 0.707447 |
| 5.251 | 0.712766 |
| 5.034 | 0.718085 |
| 4.983 | 0.723404 |
| 4.974 | 0.728723 |
| 4.945 | 0.734043 |
| 4.62 | 0.739362 |
| 4.618 | 0.744681 |
| 4.562 | 0.75 |
| 4.541 | 0.755319 |
| 4.454 | 0.760638 |
| 4.312 | 0.765957 |
| 4.192 | 0.771277 |
| 3.967 | 0.776596 |
| 3.884 | 0.781915 |
| 3.87 | 0.787234 |
| 3.605 | 0.792553 |
| 3.584 | 0.797872 |
| 3.518 | 0.803191 |
| 3.423 | 0.808511 |
| 2.783 | 0.81383 |
| 2.743 | 0.819149 |
| 2.525 | 0.824468 |
| 2.425 | 0.829787 |
| 2.233 | 0.835106 |
| 2.141 | 0.840426 |
| 2.112 | 0.845745 |
| 2.088 | 0.851064 |
| 2.049 | 0.856383 |
| 1.883 | 0.861702 |
| 1.868 | 0.867021 |
| 1.861 | 0.87234 |
| 1.855 | 0.87766 |
| 1.836 | 0.882979 |
| 1.792 | 0.888298 |
| 1.791 | 0.893617 |
| 1.762 | 0.898936 |
| 1.755 | 0.904255 |
| 1.736 | 0.909574 |
| 1.665 | 0.914894 |
| 1.578 | 0.920213 |
| 1.559 | 0.925532 |
| 1.556 | 0.930851 |
| 1.471 | 0.93617 |
| 1.464 | 0.941489 |
| 1.408 | 0.946809 |
| 1.365 | 0.952128 |
| 1.34 | 0.957447 |
| 1.261 | 0.962766 |
| 1.032 | 0.968085 |
| 0.755 | 0.973404 |
| 0.747 | 0.978723 |
| 0.71 | 0.984043 |
| 0.638 | 0.989362 |
| 0.36 | 0.994681 |
| 0.25 | 1 |

|  | ST 19F-Prevenar 13 (Day 300) |
| --- | --- |
| 148.454 | 0.00526316 |
| 95.266 | 0.0105263 |
| 94.719 | 0.0157895 |
| 91.661 | 0.0210526 |
| 89.765 | 0.0263158 |
| 89.204 | 0.0315789 |
| 86.87 | 0.0368421 |
| 85.647 | 0.0421053 |
| 81.136 | 0.0473684 |
| 71.092 | 0.0526316 |
| 66.537 | 0.0578947 |
| 52.255 | 0.0631579 |
| 51.195 | 0.0684211 |
| 46.738 | 0.0736842 |
| 45.727 | 0.0789474 |
| 43.813 | 0.0842105 |
| 43.737 | 0.0894737 |
| 43.087 | 0.0947368 |
| 38.1 | 0.1 |
| 35.824 | 0.105263 |
| 34.362 | 0.110526 |
| 34.303 | 0.115789 |
| 34.21 | 0.121053 |
| 29.81 | 0.126316 |
| 25.828 | 0.131579 |
| 25.367 | 0.136842 |
| 24.383 | 0.142105 |
| 24.041 | 0.147368 |
| 23.469 | 0.152632 |
| 22.341 | 0.157895 |
| 21.964 | 0.163158 |
| 21.878 | 0.168421 |
| 21.595 | 0.173684 |
| 20.882 | 0.178947 |
| 20.749 | 0.184211 |
| 20.243 | 0.189474 |
| 20.235 | 0.194737 |
| 20.234 | 0.2 |
| 19.875 | 0.205263 |
| 19.846 | 0.210526 |
| 19.695 | 0.215789 |
| 19.692 | 0.221053 |
| 19.66 | 0.226316 |
| 19.206 | 0.231579 |
| 19.058 | 0.236842 |
| 18.937 | 0.242105 |
| 18.51 | 0.247368 |
| 17.875 | 0.252632 |
| 17.666 | 0.257895 |
| 17.171 | 0.263158 |
| 17.143 | 0.268421 |
| 17.123 | 0.273684 |
| 16.818 | 0.278947 |
| 16.042 | 0.284211 |
| 15.381 | 0.289474 |
| 14.849 | 0.294737 |
| 14.714 | 0.3 |
| 14.469 | 0.305263 |
| 14.422 | 0.310526 |
| 14.403 | 0.315789 |
| 14.322 | 0.321053 |
| 14.17 | 0.326316 |
| 13.881 | 0.331579 |
| 13.872 | 0.336842 |
| 13.573 | 0.342105 |
| 13.313 | 0.347368 |
| 13.167 | 0.352632 |
| 13.129 | 0.357895 |
| 13.023 | 0.363158 |
| 12.797 | 0.368421 |
| 12.211 | 0.373684 |
| 11.769 | 0.378947 |
| 10.758 | 0.384211 |
| 10.667 | 0.389474 |
| 10.407 | 0.394737 |
| 10.274 | 0.4 |
| 10.065 | 0.405263 |
| 10.043 | 0.410526 |
| 9.928 | 0.415789 |
| 9.867 | 0.421053 |
| 9.824 | 0.426316 |
| 9.807 | 0.431579 |
| 9.546 | 0.436842 |
| 9.448 | 0.442105 |
| 9.419 | 0.447368 |
| 9.286 | 0.452632 |
| 9.21 | 0.457895 |
| 9.101 | 0.463158 |
| 8.899 | 0.468421 |
| 8.728 | 0.473684 |
| 8.55 | 0.478947 |
| 8.362 | 0.484211 |
| 8.007 | 0.489474 |
| 7.896 | 0.494737 |
| 7.794 | 0.5 |
| 7.761 | 0.505263 |
| 7.571 | 0.510526 |
| 7.521 | 0.515789 |
| 7.44 | 0.521053 |
| 7.333 | 0.526316 |
| 7.223 | 0.531579 |
| 7.02 | 0.536842 |
| 7.02 | 0.542105 |
| 7.017 | 0.547368 |
| 6.919 | 0.552632 |
| 6.909 | 0.557895 |
| 6.842 | 0.563158 |
| 6.822 | 0.568421 |
| 6.771 | 0.573684 |
| 6.761 | 0.578947 |
| 6.671 | 0.584211 |
| 6.542 | 0.589474 |
| 6.519 | 0.594737 |
| 6.448 | 0.6 |
| 6.429 | 0.605263 |
| 6.348 | 0.610526 |
| 6.29 | 0.615789 |
| 6.261 | 0.621053 |
| 6.252 | 0.626316 |
| 6.179 | 0.631579 |
| 6.04 | 0.636842 |
| 6.012 | 0.642105 |
| 5.995 | 0.647368 |
| 5.934 | 0.652632 |
| 5.708 | 0.657895 |
| 5.701 | 0.663158 |
| 5.666 | 0.668421 |
| 5.485 | 0.673684 |
| 5.479 | 0.678947 |
| 5.438 | 0.684211 |
| 5.387 | 0.689474 |
| 5.322 | 0.694737 |
| 5.233 | 0.7 |
| 5.169 | 0.705263 |
| 5.152 | 0.710526 |
| 5.117 | 0.715789 |
| 5.085 | 0.721053 |
| 4.986 | 0.726316 |
| 4.947 | 0.731579 |
| 4.941 | 0.736842 |
| 4.845 | 0.742105 |
| 4.794 | 0.747368 |
| 4.733 | 0.752632 |
| 4.724 | 0.757895 |
| 4.677 | 0.763158 |
| 4.639 | 0.768421 |
| 4.525 | 0.773684 |
| 4.469 | 0.778947 |
| 4.388 | 0.784211 |
| 4.336 | 0.789474 |
| 4.292 | 0.794737 |
| 4.017 | 0.8 |
| 3.961 | 0.805263 |
| 3.507 | 0.810526 |
| 3.448 | 0.815789 |
| 3.374 | 0.821053 |
| 3.219 | 0.826316 |
| 3.205 | 0.831579 |
| 3.037 | 0.836842 |
| 2.857 | 0.842105 |
| 2.855 | 0.847368 |
| 2.766 | 0.852632 |
| 2.702 | 0.857895 |
| 2.642 | 0.863158 |
| 2.532 | 0.868421 |
| 2.474 | 0.873684 |
| 2.437 | 0.878947 |
| 2.417 | 0.884211 |
| 2.353 | 0.889474 |
| 2.103 | 0.894737 |
| 1.987 | 0.9 |
| 1.965 | 0.905263 |
| 1.946 | 0.910526 |
| 1.893 | 0.915789 |
| 1.778 | 0.921053 |
| 1.724 | 0.926316 |
| 1.6 | 0.931579 |
| 1.591 | 0.936842 |
| 1.478 | 0.942105 |
| 1.392 | 0.947368 |
| 1.361 | 0.952632 |
| 1.31 | 0.957895 |
| 1.115 | 0.963158 |
| 0.843 | 0.968421 |
| 0.841 | 0.973684 |
| 0.728 | 0.978947 |
| 0.657 | 0.984211 |
| 0.585 | 0.989474 |
| 0.536 | 0.994737 |
| 0.464 | 1 |

BECT056 - RCDC_Serotype 23F

|  | ST 23F-BE-PCV14 (Day 84) |
| --- | --- |
| 116.021 | 0.0052356 |
| 114.116 | 0.0104712 |
| 75.769 | 0.0157068 |
| 74.252 | 0.0209424 |
| 71.535 | 0.026178 |
| 63.616 | 0.0314136 |
| 51.262 | 0.0366492 |
| 41.64 | 0.0418848 |
| 38.365 | 0.0471204 |
| 37.613 | 0.052356 |
| 35.133 | 0.0575916 |
| 34.455 | 0.0628272 |
| 31.88 | 0.0680628 |
| 28.392 | 0.0732984 |
| 24.025 | 0.078534 |
| 21.555 | 0.0837696 |
| 20.84 | 0.0890052 |
| 18.899 | 0.0942408 |
| 18.75 | 0.0994764 |
| 17.537 | 0.104712 |
| 15.235 | 0.109948 |
| 14.026 | 0.115183 |
| 13.979 | 0.120419 |
| 13.478 | 0.125654 |
| 13.182 | 0.13089 |
| 11.259 | 0.136126 |
| 11.063 | 0.141361 |
| 10.689 | 0.146597 |
| 10.488 | 0.151832 |
| 10.175 | 0.157068 |
| 9.823 | 0.162304 |
| 9.71 | 0.167539 |
| 8.98 | 0.172775 |
| 8.456 | 0.17801 |
| 8.193 | 0.183246 |
| 7.929 | 0.188482 |
| 6.716 | 0.193717 |
| 6.709 | 0.198953 |
| 6.384 | 0.204188 |
| 5.799 | 0.209424 |
| 5.698 | 0.21466 |
| 5.429 | 0.219895 |
| 4.83 | 0.225131 |
| 4.806 | 0.230366 |
| 4.666 | 0.235602 |
| 4.484 | 0.240838 |
| 4.408 | 0.246073 |
| 4.393 | 0.251309 |
| 4.257 | 0.256545 |
| 4.119 | 0.26178 |
| 3.885 | 0.267016 |
| 3.801 | 0.272251 |
| 3.669 | 0.277487 |
| 3.651 | 0.282723 |
| 3.606 | 0.287958 |
| 3.495 | 0.293194 |
| 3.481 | 0.298429 |
| 3.454 | 0.303665 |
| 3.38 | 0.308901 |
| 3.229 | 0.314136 |
| 3.179 | 0.319372 |
| 3.125 | 0.324607 |
| 2.924 | 0.329843 |
| 2.815 | 0.335079 |
| 2.733 | 0.340314 |
| 2.705 | 0.34555 |
| 2.681 | 0.350785 |
| 2.631 | 0.356021 |
| 2.593 | 0.361257 |
| 2.575 | 0.366492 |
| 2.559 | 0.371728 |
| 2.488 | 0.376963 |
| 2.477 | 0.382199 |
| 2.449 | 0.387435 |
| 2.435 | 0.39267 |
| 2.428 | 0.397906 |
| 2.415 | 0.403141 |
| 2.399 | 0.408377 |
| 2.369 | 0.413613 |
| 2.328 | 0.418848 |
| 2.295 | 0.424084 |
| 2.294 | 0.429319 |
| 2.29 | 0.434555 |
| 2.277 | 0.439791 |
| 2.268 | 0.445026 |
| 2.265 | 0.450262 |
| 2.138 | 0.455497 |
| 2.085 | 0.460733 |
| 2.057 | 0.465969 |
| 2.017 | 0.471204 |
| 1.991 | 0.47644 |
| 1.98 | 0.481675 |
| 1.961 | 0.486911 |
| 1.932 | 0.492147 |
| 1.889 | 0.497382 |
| 1.81 | 0.502618 |
| 1.777 | 0.507853 |
| 1.705 | 0.513089 |
| 1.65 | 0.518325 |
| 1.619 | 0.52356 |
| 1.614 | 0.528796 |
| 1.577 | 0.534031 |
| 1.552 | 0.539267 |
| 1.519 | 0.544503 |
| 1.515 | 0.549738 |
| 1.5 | 0.554974 |
| 1.499 | 0.560209 |
| 1.447 | 0.565445 |
| 1.322 | 0.570681 |
| 1.307 | 0.575916 |
| 1.281 | 0.581152 |
| 1.271 | 0.586387 |
| 1.262 | 0.591623 |
| 1.166 | 0.596859 |
| 1.157 | 0.602094 |
| 1.146 | 0.60733 |
| 1.142 | 0.612565 |
| 1.125 | 0.617801 |
| 1.123 | 0.623037 |
| 1.117 | 0.628272 |
| 1.117 | 0.633508 |
| 1.106 | 0.638743 |
| 1.064 | 0.643979 |
| 1.059 | 0.649215 |
| 1.059 | 0.65445 |
| 0.989 | 0.659686 |
| 0.982 | 0.664921 |
| 0.961 | 0.670157 |
| 0.955 | 0.675393 |
| 0.922 | 0.680628 |
| 0.916 | 0.685864 |
| 0.87 | 0.691099 |
| 0.86 | 0.696335 |
| 0.841 | 0.701571 |
| 0.816 | 0.706806 |
| 0.815 | 0.712042 |
| 0.788 | 0.717277 |
| 0.776 | 0.722513 |
| 0.77 | 0.727749 |
| 0.758 | 0.732984 |
| 0.724 | 0.73822 |
| 0.652 | 0.743455 |
| 0.635 | 0.748691 |
| 0.63 | 0.753927 |
| 0.614 | 0.759162 |
| 0.6 | 0.764398 |
| 0.583 | 0.769634 |
| 0.575 | 0.774869 |
| 0.557 | 0.780105 |
| 0.555 | 0.78534 |
| 0.539 | 0.790576 |
| 0.518 | 0.795812 |
| 0.507 | 0.801047 |
| 0.49 | 0.806283 |
| 0.484 | 0.811518 |
| 0.483 | 0.816754 |
| 0.48 | 0.82199 |
| 0.472 | 0.827225 |
| 0.46 | 0.832461 |
| 0.455 | 0.837696 |
| 0.454 | 0.842932 |
| 0.435 | 0.848168 |
| 0.411 | 0.853403 |
| 0.404 | 0.858639 |
| 0.379 | 0.863874 |
| 0.377 | 0.86911 |
| 0.377 | 0.874346 |
| 0.375 | 0.879581 |
| 0.365 | 0.884817 |
| 0.349 | 0.890052 |
| 0.347 | 0.895288 |
| 0.321 | 0.900524 |
| 0.318 | 0.905759 |
| 0.316 | 0.910995 |
| 0.302 | 0.91623 |
| 0.282 | 0.921466 |
| 0.282 | 0.926702 |
| 0.263 | 0.931937 |
| 0.253 | 0.937173 |
| 0.244 | 0.942408 |
| 0.224 | 0.947644 |
| 0.218 | 0.95288 |
| 0.202 | 0.958115 |
| 0.18 | 0.963351 |
| 0.157 | 0.968586 |
| 0.155 | 0.973822 |
| 0.153 | 0.979058 |
| 0.147 | 0.984293 |
| 0.136 | 0.989529 |
| 0.108 | 0.994764 |
| 0.075 | 1 |

|  | ST 23F-Prevenar 13 (Day 84) |
| --- | --- |
| 177.287 | 0.00505051 |
| 147.579 | 0.010101 |
| 115.825 | 0.0151515 |
| 73.617 | 0.020202 |
| 72.26 | 0.0252525 |
| 68.409 | 0.030303 |
| 66.439 | 0.0353535 |
| 38.877 | 0.040404 |
| 38.835 | 0.0454545 |
| 38.522 | 0.0505051 |
| 38.244 | 0.0555556 |
| 34.389 | 0.0606061 |
| 34.257 | 0.0656566 |
| 30.89 | 0.0707071 |
| 28.556 | 0.0757576 |
| 28.541 | 0.0808081 |
| 23.539 | 0.0858586 |
| 17.982 | 0.0909091 |
| 17.011 | 0.0959596 |
| 15.229 | 0.10101 |
| 14.757 | 0.106061 |
| 13.118 | 0.111111 |
| 12.417 | 0.116162 |
| 11.817 | 0.121212 |
| 11.735 | 0.126263 |
| 11.498 | 0.131313 |
| 11.319 | 0.136364 |
| 10.7 | 0.141414 |
| 10.486 | 0.146465 |
| 10.396 | 0.151515 |
| 10.202 | 0.156566 |
| 9.576 | 0.161616 |
| 9.463 | 0.166667 |
| 8.883 | 0.171717 |
| 8.826 | 0.176768 |
| 7.073 | 0.181818 |
| 6.627 | 0.186869 |
| 5.864 | 0.191919 |
| 5.149 | 0.19697 |
| 5.104 | 0.20202 |
| 4.945 | 0.207071 |
| 4.786 | 0.212121 |
| 4.749 | 0.217172 |
| 4.64 | 0.222222 |
| 4.551 | 0.227273 |
| 4.26 | 0.232323 |
| 4.228 | 0.237374 |
| 4.16 | 0.242424 |
| 3.818 | 0.247475 |
| 3.767 | 0.252525 |
| 3.728 | 0.257576 |
| 3.63 | 0.262626 |
| 3.627 | 0.267677 |
| 3.548 | 0.272727 |
| 3.511 | 0.277778 |
| 3.51 | 0.282828 |
| 3.48 | 0.287879 |
| 3.471 | 0.292929 |
| 3.416 | 0.29798 |
| 3.198 | 0.30303 |
| 3.175 | 0.308081 |
| 2.991 | 0.313131 |
| 2.826 | 0.318182 |
| 2.777 | 0.323232 |
| 2.735 | 0.328283 |
| 2.632 | 0.333333 |
| 2.504 | 0.338384 |
| 2.484 | 0.343434 |
| 2.433 | 0.348485 |
| 2.352 | 0.353535 |
| 2.278 | 0.358586 |
| 2.256 | 0.363636 |
| 2.164 | 0.368687 |
| 2.105 | 0.373737 |
| 2.054 | 0.378788 |
| 1.97 | 0.383838 |
| 1.929 | 0.388889 |
| 1.882 | 0.393939 |
| 1.869 | 0.39899 |
| 1.858 | 0.40404 |
| 1.828 | 0.409091 |
| 1.765 | 0.414141 |
| 1.734 | 0.419192 |
| 1.668 | 0.424242 |
| 1.648 | 0.429293 |
| 1.636 | 0.434343 |
| 1.633 | 0.439394 |
| 1.623 | 0.444444 |
| 1.601 | 0.449495 |
| 1.581 | 0.454545 |
| 1.557 | 0.459596 |
| 1.515 | 0.464646 |
| 1.496 | 0.469697 |
| 1.494 | 0.474747 |
| 1.48 | 0.479798 |
| 1.476 | 0.484848 |
| 1.46 | 0.489899 |
| 1.455 | 0.494949 |
| 1.454 | 0.5 |
| 1.449 | 0.505051 |
| 1.422 | 0.510101 |
| 1.345 | 0.515152 |
| 1.328 | 0.520202 |
| 1.285 | 0.525253 |
| 1.285 | 0.530303 |
| 1.278 | 0.535354 |
| 1.234 | 0.540404 |
| 1.231 | 0.545455 |
| 1.227 | 0.550505 |
| 1.21 | 0.555556 |
| 1.178 | 0.560606 |
| 1.166 | 0.565657 |
| 1.158 | 0.570707 |
| 1.114 | 0.575758 |
| 1.112 | 0.580808 |
| 1.099 | 0.585859 |
| 1.083 | 0.590909 |
| 1.077 | 0.59596 |
| 1.056 | 0.60101 |
| 1.043 | 0.606061 |
| 1.014 | 0.611111 |
| 1.007 | 0.616162 |
| 0.919 | 0.621212 |
| 0.853 | 0.626263 |
| 0.843 | 0.631313 |
| 0.841 | 0.636364 |
| 0.798 | 0.641414 |
| 0.78 | 0.646465 |
| 0.751 | 0.651515 |
| 0.748 | 0.656566 |
| 0.743 | 0.661616 |
| 0.736 | 0.666667 |
| 0.733 | 0.671717 |
| 0.715 | 0.676768 |
| 0.713 | 0.681818 |
| 0.708 | 0.686869 |
| 0.705 | 0.691919 |
| 0.656 | 0.69697 |
| 0.647 | 0.70202 |
| 0.641 | 0.707071 |
| 0.64 | 0.712121 |
| 0.635 | 0.717172 |
| 0.629 | 0.722222 |
| 0.619 | 0.727273 |
| 0.615 | 0.732323 |
| 0.6 | 0.737374 |
| 0.593 | 0.742424 |
| 0.586 | 0.747475 |
| 0.584 | 0.752525 |
| 0.51 | 0.757576 |
| 0.508 | 0.762626 |
| 0.508 | 0.767677 |
| 0.508 | 0.772727 |
| 0.505 | 0.777778 |
| 0.505 | 0.782828 |
| 0.504 | 0.787879 |
| 0.496 | 0.792929 |
| 0.493 | 0.79798 |
| 0.491 | 0.80303 |
| 0.488 | 0.808081 |
| 0.485 | 0.813131 |
| 0.474 | 0.818182 |
| 0.47 | 0.823232 |
| 0.466 | 0.828283 |
| 0.455 | 0.833333 |
| 0.444 | 0.838384 |
| 0.435 | 0.843434 |
| 0.417 | 0.848485 |
| 0.41 | 0.853535 |
| 0.392 | 0.858586 |
| 0.392 | 0.863636 |
| 0.386 | 0.868687 |
| 0.384 | 0.873737 |
| 0.376 | 0.878788 |
| 0.369 | 0.883838 |
| 0.366 | 0.888889 |
| 0.36 | 0.893939 |
| 0.344 | 0.89899 |
| 0.342 | 0.90404 |
| 0.314 | 0.909091 |
| 0.302 | 0.914141 |
| 0.292 | 0.919192 |
| 0.253 | 0.924242 |
| 0.249 | 0.929293 |
| 0.235 | 0.934343 |
| 0.232 | 0.939394 |
| 0.228 | 0.944444 |
| 0.223 | 0.949495 |
| 0.214 | 0.954545 |
| 0.202 | 0.959596 |
| 0.178 | 0.964646 |
| 0.168 | 0.969697 |
| 0.168 | 0.974747 |
| 0.166 | 0.979798 |
| 0.164 | 0.984848 |
| 0.142 | 0.989899 |
| 0.134 | 0.994949 |
| 0.125 | 1 |

|  | ST 23F-BE-PCV14 (Day 300) |
| --- | --- |
| 33.191 | 0.00531915 |
| 28.297 | 0.0106383 |
| 24.768 | 0.0159574 |
| 23.679 | 0.0212766 |
| 21.375 | 0.0265957 |
| 20.434 | 0.0319149 |
| 19.728 | 0.037234 |
| 18.143 | 0.0425532 |
| 18.081 | 0.0478723 |
| 18.046 | 0.0531915 |
| 17.459 | 0.0585106 |
| 17.287 | 0.0638298 |
| 17.28 | 0.0691489 |
| 16.297 | 0.0744681 |
| 16.178 | 0.0797872 |
| 15.925 | 0.0851064 |
| 15.847 | 0.0904255 |
| 14.574 | 0.0957447 |
| 14.501 | 0.101064 |
| 12.488 | 0.106383 |
| 12.24 | 0.111702 |
| 12.065 | 0.117021 |
| 11.61 | 0.12234 |
| 11.273 | 0.12766 |
| 11.2 | 0.132979 |
| 11.197 | 0.138298 |
| 11.075 | 0.143617 |
| 10.76 | 0.148936 |
| 10.582 | 0.154255 |
| 10.48 | 0.159574 |
| 10.263 | 0.164894 |
| 10.152 | 0.170213 |
| 9.659 | 0.175532 |
| 9.646 | 0.180851 |
| 9.369 | 0.18617 |
| 9.358 | 0.191489 |
| 9.269 | 0.196809 |
| 9.21 | 0.202128 |
| 8.496 | 0.207447 |
| 8.325 | 0.212766 |
| 8.266 | 0.218085 |
| 8.21 | 0.223404 |
| 8.153 | 0.228723 |
| 8.109 | 0.234043 |
| 8.048 | 0.239362 |
| 8.007 | 0.244681 |
| 7.995 | 0.25 |
| 7.831 | 0.255319 |
| 7.786 | 0.260638 |
| 7.667 | 0.265957 |
| 7.533 | 0.271277 |
| 7.486 | 0.276596 |
| 7.364 | 0.281915 |
| 7.016 | 0.287234 |
| 6.575 | 0.292553 |
| 6.557 | 0.297872 |
| 6.525 | 0.303191 |
| 6.513 | 0.308511 |
| 6.469 | 0.31383 |
| 6.324 | 0.319149 |
| 6.297 | 0.324468 |
| 6.154 | 0.329787 |
| 6.046 | 0.335106 |
| 5.85 | 0.340426 |
| 5.616 | 0.345745 |
| 5.239 | 0.351064 |
| 5.181 | 0.356383 |
| 5.066 | 0.361702 |
| 5.034 | 0.367021 |
| 4.997 | 0.37234 |
| 4.966 | 0.37766 |
| 4.902 | 0.382979 |
| 4.764 | 0.388298 |
| 4.629 | 0.393617 |
| 4.608 | 0.398936 |
| 4.58 | 0.404255 |
| 4.494 | 0.409574 |
| 4.379 | 0.414894 |
| 4.301 | 0.420213 |
| 4.255 | 0.425532 |
| 4.252 | 0.430851 |
| 4.244 | 0.43617 |
| 4.182 | 0.441489 |
| 4.134 | 0.446809 |
| 3.996 | 0.452128 |
| 3.88 | 0.457447 |
| 3.846 | 0.462766 |
| 3.776 | 0.468085 |
| 3.776 | 0.473404 |
| 3.721 | 0.478723 |
| 3.448 | 0.484043 |
| 3.427 | 0.489362 |
| 3.304 | 0.494681 |
| 3.21 | 0.5 |
| 3.083 | 0.505319 |
| 3.076 | 0.510638 |
| 2.924 | 0.515957 |
| 2.859 | 0.521277 |
| 2.85 | 0.526596 |
| 2.835 | 0.531915 |
| 2.801 | 0.537234 |
| 2.719 | 0.542553 |
| 2.715 | 0.547872 |
| 2.703 | 0.553191 |
| 2.585 | 0.558511 |
| 2.565 | 0.56383 |
| 2.529 | 0.569149 |
| 2.474 | 0.574468 |
| 2.366 | 0.579787 |
| 2.163 | 0.585106 |
| 2.132 | 0.590426 |
| 2.031 | 0.595745 |
| 2.023 | 0.601064 |
| 2.01 | 0.606383 |
| 2.009 | 0.611702 |
| 1.994 | 0.617021 |
| 1.969 | 0.62234 |
| 1.956 | 0.62766 |
| 1.912 | 0.632979 |
| 1.907 | 0.638298 |
| 1.877 | 0.643617 |
| 1.876 | 0.648936 |
| 1.86 | 0.654255 |
| 1.819 | 0.659574 |
| 1.815 | 0.664894 |
| 1.809 | 0.670213 |
| 1.752 | 0.675532 |
| 1.747 | 0.680851 |
| 1.74 | 0.68617 |
| 1.718 | 0.691489 |
| 1.71 | 0.696809 |
| 1.703 | 0.702128 |
| 1.693 | 0.707447 |
| 1.656 | 0.712766 |
| 1.649 | 0.718085 |
| 1.617 | 0.723404 |
| 1.588 | 0.728723 |
| 1.588 | 0.734043 |
| 1.555 | 0.739362 |
| 1.509 | 0.744681 |
| 1.503 | 0.75 |
| 1.465 | 0.755319 |
| 1.459 | 0.760638 |
| 1.427 | 0.765957 |
| 1.41 | 0.771277 |
| 1.356 | 0.776596 |
| 1.346 | 0.781915 |
| 1.238 | 0.787234 |
| 1.203 | 0.792553 |
| 1.182 | 0.797872 |
| 1.134 | 0.803191 |
| 1.129 | 0.808511 |
| 1.1 | 0.81383 |
| 1.073 | 0.819149 |
| 1.048 | 0.824468 |
| 0.97 | 0.829787 |
| 0.962 | 0.835106 |
| 0.946 | 0.840426 |
| 0.942 | 0.845745 |
| 0.906 | 0.851064 |
| 0.857 | 0.856383 |
| 0.647 | 0.861702 |
| 0.584 | 0.867021 |
| 0.581 | 0.87234 |
| 0.567 | 0.87766 |
| 0.552 | 0.882979 |
| 0.534 | 0.888298 |
| 0.524 | 0.893617 |
| 0.518 | 0.898936 |
| 0.482 | 0.904255 |
| 0.424 | 0.909574 |
| 0.399 | 0.914894 |
| 0.397 | 0.920213 |
| 0.365 | 0.925532 |
| 0.358 | 0.930851 |
| 0.355 | 0.93617 |
| 0.349 | 0.941489 |
| 0.279 | 0.946809 |
| 0.254 | 0.952128 |
| 0.23 | 0.957447 |
| 0.228 | 0.962766 |
| 0.228 | 0.968085 |
| 0.169 | 0.973404 |
| 0.169 | 0.978723 |
| 0.16 | 0.984043 |
| 0.114 | 0.989362 |
| 0.101 | 0.994681 |
| 0.079 | 1 |

|  | ST 23F-Prevenar 13 (Day 300) |
| --- | --- |
| 97.084 | 0.00526316 |
| 72.286 | 0.0105263 |
| 68.286 | 0.0157895 |
| 62.251 | 0.0210526 |
| 62.18 | 0.0263158 |
| 61.656 | 0.0315789 |
| 56.876 | 0.0368421 |
| 43.072 | 0.0421053 |
| 36.092 | 0.0473684 |
| 34.797 | 0.0526316 |
| 34.473 | 0.0578947 |
| 33.673 | 0.0631579 |
| 28.776 | 0.0684211 |
| 27.642 | 0.0736842 |
| 26.17 | 0.0789474 |
| 22.387 | 0.0842105 |
| 22.323 | 0.0894737 |
| 21.902 | 0.0947368 |
| 21.324 | 0.1 |
| 21.142 | 0.105263 |
| 20.547 | 0.110526 |
| 18.719 | 0.115789 |
| 17.656 | 0.121053 |
| 16.944 | 0.126316 |
| 15.934 | 0.131579 |
| 15.905 | 0.136842 |
| 15.516 | 0.142105 |
| 15.2 | 0.147368 |
| 15.131 | 0.152632 |
| 14.369 | 0.157895 |
| 13.796 | 0.163158 |
| 12.749 | 0.168421 |
| 11.971 | 0.173684 |
| 11.472 | 0.178947 |
| 9.789 | 0.184211 |
| 8.987 | 0.189474 |
| 8.857 | 0.194737 |
| 8.099 | 0.2 |
| 8.093 | 0.205263 |
| 7.502 | 0.210526 |
| 7.446 | 0.215789 |
| 7.365 | 0.221053 |
| 6.918 | 0.226316 |
| 6.85 | 0.231579 |
| 6.485 | 0.236842 |
| 6.124 | 0.242105 |
| 5.944 | 0.247368 |
| 5.866 | 0.252632 |
| 5.702 | 0.257895 |
| 5.614 | 0.263158 |
| 5.597 | 0.268421 |
| 5.596 | 0.273684 |
| 5.526 | 0.278947 |
| 5.483 | 0.284211 |
| 5.355 | 0.289474 |
| 4.905 | 0.294737 |
| 4.885 | 0.3 |
| 4.873 | 0.305263 |
| 4.858 | 0.310526 |
| 4.783 | 0.315789 |
| 4.708 | 0.321053 |
| 4.524 | 0.326316 |
| 4.495 | 0.331579 |
| 4.486 | 0.336842 |
| 4.479 | 0.342105 |
| 4.471 | 0.347368 |
| 4.425 | 0.352632 |
| 4.41 | 0.357895 |
| 4.397 | 0.363158 |
| 4.297 | 0.368421 |
| 4.193 | 0.373684 |
| 4.143 | 0.378947 |
| 4.096 | 0.384211 |
| 4.086 | 0.389474 |
| 4.072 | 0.394737 |
| 4.052 | 0.4 |
| 3.994 | 0.405263 |
| 3.929 | 0.410526 |
| 3.838 | 0.415789 |
| 3.703 | 0.421053 |
| 3.687 | 0.426316 |
| 3.534 | 0.431579 |
| 3.531 | 0.436842 |
| 3.482 | 0.442105 |
| 3.418 | 0.447368 |
| 3.405 | 0.452632 |
| 3.357 | 0.457895 |
| 3.349 | 0.463158 |
| 3.292 | 0.468421 |
| 3.246 | 0.473684 |
| 3.244 | 0.478947 |
| 3.154 | 0.484211 |
| 3.109 | 0.489474 |
| 3.052 | 0.494737 |
| 3.047 | 0.5 |
| 3.042 | 0.505263 |
| 3.019 | 0.510526 |
| 2.934 | 0.515789 |
| 2.929 | 0.521053 |
| 2.924 | 0.526316 |
| 2.914 | 0.531579 |
| 2.911 | 0.536842 |
| 2.902 | 0.542105 |
| 2.899 | 0.547368 |
| 2.848 | 0.552632 |
| 2.844 | 0.557895 |
| 2.841 | 0.563158 |
| 2.795 | 0.568421 |
| 2.766 | 0.573684 |
| 2.765 | 0.578947 |
| 2.765 | 0.584211 |
| 2.764 | 0.589474 |
| 2.652 | 0.594737 |
| 2.594 | 0.6 |
| 2.581 | 0.605263 |
| 2.576 | 0.610526 |
| 2.561 | 0.615789 |
| 2.537 | 0.621053 |
| 2.496 | 0.626316 |
| 2.467 | 0.631579 |
| 2.462 | 0.636842 |
| 2.46 | 0.642105 |
| 2.411 | 0.647368 |
| 2.409 | 0.652632 |
| 2.399 | 0.657895 |
| 2.396 | 0.663158 |
| 2.335 | 0.668421 |
| 2.322 | 0.673684 |
| 2.29 | 0.678947 |
| 2.286 | 0.684211 |
| 2.279 | 0.689474 |
| 2.261 | 0.694737 |
| 2.227 | 0.7 |
| 2.174 | 0.705263 |
| 2.146 | 0.710526 |
| 2.125 | 0.715789 |
| 2.014 | 0.721053 |
| 1.99 | 0.726316 |
| 1.944 | 0.731579 |
| 1.904 | 0.736842 |
| 1.903 | 0.742105 |
| 1.844 | 0.747368 |
| 1.818 | 0.752632 |
| 1.765 | 0.757895 |
| 1.755 | 0.763158 |
| 1.652 | 0.768421 |
| 1.613 | 0.773684 |
| 1.605 | 0.778947 |
| 1.54 | 0.784211 |
| 1.529 | 0.789474 |
| 1.497 | 0.794737 |
| 1.496 | 0.8 |
| 1.493 | 0.805263 |
| 1.451 | 0.810526 |
| 1.437 | 0.815789 |
| 1.413 | 0.821053 |
| 1.398 | 0.826316 |
| 1.374 | 0.831579 |
| 1.3 | 0.836842 |
| 1.27 | 0.842105 |
| 1.264 | 0.847368 |
| 1.186 | 0.852632 |
| 1.066 | 0.857895 |
| 0.775 | 0.863158 |
| 0.707 | 0.868421 |
| 0.693 | 0.873684 |
| 0.684 | 0.878947 |
| 0.666 | 0.884211 |
| 0.65 | 0.889474 |
| 0.555 | 0.894737 |
| 0.511 | 0.9 |
| 0.48 | 0.905263 |
| 0.477 | 0.910526 |
| 0.471 | 0.915789 |
| 0.464 | 0.921053 |
| 0.457 | 0.926316 |
| 0.433 | 0.931579 |
| 0.353 | 0.936842 |
| 0.249 | 0.942105 |
| 0.241 | 0.947368 |
| 0.231 | 0.952632 |
| 0.225 | 0.957895 |
| 0.202 | 0.963158 |
| 0.174 | 0.968421 |
| 0.159 | 0.973684 |
| 0.119 | 0.978947 |
| 0.112 | 0.984211 |
| 0.109 | 0.989474 |
| 0.069 | 0.994737 |
| 0.06 | 1 |

BECT056 - RCDC_Serotype 23F

|  | ST 23F-BE-PCV14 (Day 84) |
| --- | --- |
| 116.021 | 0.0052356 |
| 114.116 | 0.0104712 |
| 75.769 | 0.0157068 |
| 74.252 | 0.0209424 |
| 71.535 | 0.026178 |
| 63.616 | 0.0314136 |
| 51.262 | 0.0366492 |
| 41.64 | 0.0418848 |
| 38.365 | 0.0471204 |
| 37.613 | 0.052356 |
| 35.133 | 0.0575916 |
| 34.455 | 0.0628272 |
| 31.88 | 0.0680628 |
| 28.392 | 0.0732984 |
| 24.025 | 0.078534 |
| 21.555 | 0.0837696 |
| 20.84 | 0.0890052 |
| 18.899 | 0.0942408 |
| 18.75 | 0.0994764 |
| 17.537 | 0.104712 |
| 15.235 | 0.109948 |
| 14.026 | 0.115183 |
| 13.979 | 0.120419 |
| 13.478 | 0.125654 |
| 13.182 | 0.13089 |
| 11.259 | 0.136126 |
| 11.063 | 0.141361 |
| 10.689 | 0.146597 |
| 10.488 | 0.151832 |
| 10.175 | 0.157068 |
| 9.823 | 0.162304 |
| 9.71 | 0.167539 |
| 8.98 | 0.172775 |
| 8.456 | 0.17801 |
| 8.193 | 0.183246 |
| 7.929 | 0.188482 |
| 6.716 | 0.193717 |
| 6.709 | 0.198953 |
| 6.384 | 0.204188 |
| 5.799 | 0.209424 |
| 5.698 | 0.21466 |
| 5.429 | 0.219895 |
| 4.83 | 0.225131 |
| 4.806 | 0.230366 |
| 4.666 | 0.235602 |
| 4.484 | 0.240838 |
| 4.408 | 0.246073 |
| 4.393 | 0.251309 |
| 4.257 | 0.256545 |
| 4.119 | 0.26178 |
| 3.885 | 0.267016 |
| 3.801 | 0.272251 |
| 3.669 | 0.277487 |
| 3.651 | 0.282723 |
| 3.606 | 0.287958 |
| 3.495 | 0.293194 |
| 3.481 | 0.298429 |
| 3.454 | 0.303665 |
| 3.38 | 0.308901 |
| 3.229 | 0.314136 |
| 3.179 | 0.319372 |
| 3.125 | 0.324607 |
| 2.924 | 0.329843 |
| 2.815 | 0.335079 |
| 2.733 | 0.340314 |
| 2.705 | 0.34555 |
| 2.681 | 0.350785 |
| 2.631 | 0.356021 |
| 2.593 | 0.361257 |
| 2.575 | 0.366492 |
| 2.559 | 0.371728 |
| 2.488 | 0.376963 |
| 2.477 | 0.382199 |
| 2.449 | 0.387435 |
| 2.435 | 0.39267 |
| 2.428 | 0.397906 |
| 2.415 | 0.403141 |
| 2.399 | 0.408377 |
| 2.369 | 0.413613 |
| 2.328 | 0.418848 |
| 2.295 | 0.424084 |
| 2.294 | 0.429319 |
| 2.29 | 0.434555 |
| 2.277 | 0.439791 |
| 2.268 | 0.445026 |
| 2.265 | 0.450262 |
| 2.138 | 0.455497 |
| 2.085 | 0.460733 |
| 2.057 | 0.465969 |
| 2.017 | 0.471204 |
| 1.991 | 0.47644 |
| 1.98 | 0.481675 |
| 1.961 | 0.486911 |
| 1.932 | 0.492147 |
| 1.889 | 0.497382 |
| 1.81 | 0.502618 |
| 1.777 | 0.507853 |
| 1.705 | 0.513089 |
| 1.65 | 0.518325 |
| 1.619 | 0.52356 |
| 1.614 | 0.528796 |
| 1.577 | 0.534031 |
| 1.552 | 0.539267 |
| 1.519 | 0.544503 |
| 1.515 | 0.549738 |
| 1.5 | 0.554974 |
| 1.499 | 0.560209 |
| 1.447 | 0.565445 |
| 1.322 | 0.570681 |
| 1.307 | 0.575916 |
| 1.281 | 0.581152 |
| 1.271 | 0.586387 |
| 1.262 | 0.591623 |
| 1.166 | 0.596859 |
| 1.157 | 0.602094 |
| 1.146 | 0.60733 |
| 1.142 | 0.612565 |
| 1.125 | 0.617801 |
| 1.123 | 0.623037 |
| 1.117 | 0.628272 |
| 1.117 | 0.633508 |
| 1.106 | 0.638743 |
| 1.064 | 0.643979 |
| 1.059 | 0.649215 |
| 1.059 | 0.65445 |
| 0.989 | 0.659686 |
| 0.982 | 0.664921 |
| 0.961 | 0.670157 |
| 0.955 | 0.675393 |
| 0.922 | 0.680628 |
| 0.916 | 0.685864 |
| 0.87 | 0.691099 |
| 0.86 | 0.696335 |
| 0.841 | 0.701571 |
| 0.816 | 0.706806 |
| 0.815 | 0.712042 |
| 0.788 | 0.717277 |
| 0.776 | 0.722513 |
| 0.77 | 0.727749 |
| 0.758 | 0.732984 |
| 0.724 | 0.73822 |
| 0.652 | 0.743455 |
| 0.635 | 0.748691 |
| 0.63 | 0.753927 |
| 0.614 | 0.759162 |
| 0.6 | 0.764398 |
| 0.583 | 0.769634 |
| 0.575 | 0.774869 |
| 0.557 | 0.780105 |
| 0.555 | 0.78534 |
| 0.539 | 0.790576 |
| 0.518 | 0.795812 |
| 0.507 | 0.801047 |
| 0.49 | 0.806283 |
| 0.484 | 0.811518 |
| 0.483 | 0.816754 |
| 0.48 | 0.82199 |
| 0.472 | 0.827225 |
| 0.46 | 0.832461 |
| 0.455 | 0.837696 |
| 0.454 | 0.842932 |
| 0.435 | 0.848168 |
| 0.411 | 0.853403 |
| 0.404 | 0.858639 |
| 0.379 | 0.863874 |
| 0.377 | 0.86911 |
| 0.377 | 0.874346 |
| 0.375 | 0.879581 |
| 0.365 | 0.884817 |
| 0.349 | 0.890052 |
| 0.347 | 0.895288 |
| 0.321 | 0.900524 |
| 0.318 | 0.905759 |
| 0.316 | 0.910995 |
| 0.302 | 0.91623 |
| 0.282 | 0.921466 |
| 0.282 | 0.926702 |
| 0.263 | 0.931937 |
| 0.253 | 0.937173 |
| 0.244 | 0.942408 |
| 0.224 | 0.947644 |
| 0.218 | 0.95288 |
| 0.202 | 0.958115 |
| 0.18 | 0.963351 |
| 0.157 | 0.968586 |
| 0.155 | 0.973822 |
| 0.153 | 0.979058 |
| 0.147 | 0.984293 |
| 0.136 | 0.989529 |
| 0.108 | 0.994764 |
| 0.075 | 1 |

|  | ST 23F-Prevenar 13 (Day 84) |
| --- | --- |
| 177.287 | 0.00505051 |
| 147.579 | 0.010101 |
| 115.825 | 0.0151515 |
| 73.617 | 0.020202 |
| 72.26 | 0.0252525 |
| 68.409 | 0.030303 |
| 66.439 | 0.0353535 |
| 38.877 | 0.040404 |
| 38.835 | 0.0454545 |
| 38.522 | 0.0505051 |
| 38.244 | 0.0555556 |
| 34.389 | 0.0606061 |
| 34.257 | 0.0656566 |
| 30.89 | 0.0707071 |
| 28.556 | 0.0757576 |
| 28.541 | 0.0808081 |
| 23.539 | 0.0858586 |
| 17.982 | 0.0909091 |
| 17.011 | 0.0959596 |
| 15.229 | 0.10101 |
| 14.757 | 0.106061 |
| 13.118 | 0.111111 |
| 12.417 | 0.116162 |
| 11.817 | 0.121212 |
| 11.735 | 0.126263 |
| 11.498 | 0.131313 |
| 11.319 | 0.136364 |
| 10.7 | 0.141414 |
| 10.486 | 0.146465 |
| 10.396 | 0.151515 |
| 10.202 | 0.156566 |
| 9.576 | 0.161616 |
| 9.463 | 0.166667 |
| 8.883 | 0.171717 |
| 8.826 | 0.176768 |
| 7.073 | 0.181818 |
| 6.627 | 0.186869 |
| 5.864 | 0.191919 |
| 5.149 | 0.19697 |
| 5.104 | 0.20202 |
| 4.945 | 0.207071 |
| 4.786 | 0.212121 |
| 4.749 | 0.217172 |
| 4.64 | 0.222222 |
| 4.551 | 0.227273 |
| 4.26 | 0.232323 |
| 4.228 | 0.237374 |
| 4.16 | 0.242424 |
| 3.818 | 0.247475 |
| 3.767 | 0.252525 |
| 3.728 | 0.257576 |
| 3.63 | 0.262626 |
| 3.627 | 0.267677 |
| 3.548 | 0.272727 |
| 3.511 | 0.277778 |
| 3.51 | 0.282828 |
| 3.48 | 0.287879 |
| 3.471 | 0.292929 |
| 3.416 | 0.29798 |
| 3.198 | 0.30303 |
| 3.175 | 0.308081 |
| 2.991 | 0.313131 |
| 2.826 | 0.318182 |
| 2.777 | 0.323232 |
| 2.735 | 0.328283 |
| 2.632 | 0.333333 |
| 2.504 | 0.338384 |
| 2.484 | 0.343434 |
| 2.433 | 0.348485 |
| 2.352 | 0.353535 |
| 2.278 | 0.358586 |
| 2.256 | 0.363636 |
| 2.164 | 0.368687 |
| 2.105 | 0.373737 |
| 2.054 | 0.378788 |
| 1.97 | 0.383838 |
| 1.929 | 0.388889 |
| 1.882 | 0.393939 |
| 1.869 | 0.39899 |
| 1.858 | 0.40404 |
| 1.828 | 0.409091 |
| 1.765 | 0.414141 |
| 1.734 | 0.419192 |
| 1.668 | 0.424242 |
| 1.648 | 0.429293 |
| 1.636 | 0.434343 |
| 1.633 | 0.439394 |
| 1.623 | 0.444444 |
| 1.601 | 0.449495 |
| 1.581 | 0.454545 |
| 1.557 | 0.459596 |
| 1.515 | 0.464646 |
| 1.496 | 0.469697 |
| 1.494 | 0.474747 |
| 1.48 | 0.479798 |
| 1.476 | 0.484848 |
| 1.46 | 0.489899 |
| 1.455 | 0.494949 |
| 1.454 | 0.5 |
| 1.449 | 0.505051 |
| 1.422 | 0.510101 |
| 1.345 | 0.515152 |
| 1.328 | 0.520202 |
| 1.285 | 0.525253 |
| 1.285 | 0.530303 |
| 1.278 | 0.535354 |
| 1.234 | 0.540404 |
| 1.231 | 0.545455 |
| 1.227 | 0.550505 |
| 1.21 | 0.555556 |
| 1.178 | 0.560606 |
| 1.166 | 0.565657 |
| 1.158 | 0.570707 |
| 1.114 | 0.575758 |
| 1.112 | 0.580808 |
| 1.099 | 0.585859 |
| 1.083 | 0.590909 |
| 1.077 | 0.59596 |
| 1.056 | 0.60101 |
| 1.043 | 0.606061 |
| 1.014 | 0.611111 |
| 1.007 | 0.616162 |
| 0.919 | 0.621212 |
| 0.853 | 0.626263 |
| 0.843 | 0.631313 |
| 0.841 | 0.636364 |
| 0.798 | 0.641414 |
| 0.78 | 0.646465 |
| 0.751 | 0.651515 |
| 0.748 | 0.656566 |
| 0.743 | 0.661616 |
| 0.736 | 0.666667 |
| 0.733 | 0.671717 |
| 0.715 | 0.676768 |
| 0.713 | 0.681818 |
| 0.708 | 0.686869 |
| 0.705 | 0.691919 |
| 0.656 | 0.69697 |
| 0.647 | 0.70202 |
| 0.641 | 0.707071 |
| 0.64 | 0.712121 |
| 0.635 | 0.717172 |
| 0.629 | 0.722222 |
| 0.619 | 0.727273 |
| 0.615 | 0.732323 |
| 0.6 | 0.737374 |
| 0.593 | 0.742424 |
| 0.586 | 0.747475 |
| 0.584 | 0.752525 |
| 0.51 | 0.757576 |
| 0.508 | 0.762626 |
| 0.508 | 0.767677 |
| 0.508 | 0.772727 |
| 0.505 | 0.777778 |
| 0.505 | 0.782828 |
| 0.504 | 0.787879 |
| 0.496 | 0.792929 |
| 0.493 | 0.79798 |
| 0.491 | 0.80303 |
| 0.488 | 0.808081 |
| 0.485 | 0.813131 |
| 0.474 | 0.818182 |
| 0.47 | 0.823232 |
| 0.466 | 0.828283 |
| 0.455 | 0.833333 |
| 0.444 | 0.838384 |
| 0.435 | 0.843434 |
| 0.417 | 0.848485 |
| 0.41 | 0.853535 |
| 0.392 | 0.858586 |
| 0.392 | 0.863636 |
| 0.386 | 0.868687 |
| 0.384 | 0.873737 |
| 0.376 | 0.878788 |
| 0.369 | 0.883838 |
| 0.366 | 0.888889 |
| 0.36 | 0.893939 |
| 0.344 | 0.89899 |
| 0.342 | 0.90404 |
| 0.314 | 0.909091 |
| 0.302 | 0.914141 |
| 0.292 | 0.919192 |
| 0.253 | 0.924242 |
| 0.249 | 0.929293 |
| 0.235 | 0.934343 |
| 0.232 | 0.939394 |
| 0.228 | 0.944444 |
| 0.223 | 0.949495 |
| 0.214 | 0.954545 |
| 0.202 | 0.959596 |
| 0.178 | 0.964646 |
| 0.168 | 0.969697 |
| 0.168 | 0.974747 |
| 0.166 | 0.979798 |
| 0.164 | 0.984848 |
| 0.142 | 0.989899 |
| 0.134 | 0.994949 |
| 0.125 | 1 |

|  | ST 23F-BE-PCV14 (Day 300) |
| --- | --- |
| 33.191 | 0.00531915 |
| 28.297 | 0.0106383 |
| 24.768 | 0.0159574 |
| 23.679 | 0.0212766 |
| 21.375 | 0.0265957 |
| 20.434 | 0.0319149 |
| 19.728 | 0.037234 |
| 18.143 | 0.0425532 |
| 18.081 | 0.0478723 |
| 18.046 | 0.0531915 |
| 17.459 | 0.0585106 |
| 17.287 | 0.0638298 |
| 17.28 | 0.0691489 |
| 16.297 | 0.0744681 |
| 16.178 | 0.0797872 |
| 15.925 | 0.0851064 |
| 15.847 | 0.0904255 |
| 14.574 | 0.0957447 |
| 14.501 | 0.101064 |
| 12.488 | 0.106383 |
| 12.24 | 0.111702 |
| 12.065 | 0.117021 |
| 11.61 | 0.12234 |
| 11.273 | 0.12766 |
| 11.2 | 0.132979 |
| 11.197 | 0.138298 |
| 11.075 | 0.143617 |
| 10.76 | 0.148936 |
| 10.582 | 0.154255 |
| 10.48 | 0.159574 |
| 10.263 | 0.164894 |
| 10.152 | 0.170213 |
| 9.659 | 0.175532 |
| 9.646 | 0.180851 |
| 9.369 | 0.18617 |
| 9.358 | 0.191489 |
| 9.269 | 0.196809 |
| 9.21 | 0.202128 |
| 8.496 | 0.207447 |
| 8.325 | 0.212766 |
| 8.266 | 0.218085 |
| 8.21 | 0.223404 |
| 8.153 | 0.228723 |
| 8.109 | 0.234043 |
| 8.048 | 0.239362 |
| 8.007 | 0.244681 |
| 7.995 | 0.25 |
| 7.831 | 0.255319 |
| 7.786 | 0.260638 |
| 7.667 | 0.265957 |
| 7.533 | 0.271277 |
| 7.486 | 0.276596 |
| 7.364 | 0.281915 |
| 7.016 | 0.287234 |
| 6.575 | 0.292553 |
| 6.557 | 0.297872 |
| 6.525 | 0.303191 |
| 6.513 | 0.308511 |
| 6.469 | 0.31383 |
| 6.324 | 0.319149 |
| 6.297 | 0.324468 |
| 6.154 | 0.329787 |
| 6.046 | 0.335106 |
| 5.85 | 0.340426 |
| 5.616 | 0.345745 |
| 5.239 | 0.351064 |
| 5.181 | 0.356383 |
| 5.066 | 0.361702 |
| 5.034 | 0.367021 |
| 4.997 | 0.37234 |
| 4.966 | 0.37766 |
| 4.902 | 0.382979 |
| 4.764 | 0.388298 |
| 4.629 | 0.393617 |
| 4.608 | 0.398936 |
| 4.58 | 0.404255 |
| 4.494 | 0.409574 |
| 4.379 | 0.414894 |
| 4.301 | 0.420213 |
| 4.255 | 0.425532 |
| 4.252 | 0.430851 |
| 4.244 | 0.43617 |
| 4.182 | 0.441489 |
| 4.134 | 0.446809 |
| 3.996 | 0.452128 |
| 3.88 | 0.457447 |
| 3.846 | 0.462766 |
| 3.776 | 0.468085 |
| 3.776 | 0.473404 |
| 3.721 | 0.478723 |
| 3.448 | 0.484043 |
| 3.427 | 0.489362 |
| 3.304 | 0.494681 |
| 3.21 | 0.5 |
| 3.083 | 0.505319 |
| 3.076 | 0.510638 |
| 2.924 | 0.515957 |
| 2.859 | 0.521277 |
| 2.85 | 0.526596 |
| 2.835 | 0.531915 |
| 2.801 | 0.537234 |
| 2.719 | 0.542553 |
| 2.715 | 0.547872 |
| 2.703 | 0.553191 |
| 2.585 | 0.558511 |
| 2.565 | 0.56383 |
| 2.529 | 0.569149 |
| 2.474 | 0.574468 |
| 2.366 | 0.579787 |
| 2.163 | 0.585106 |
| 2.132 | 0.590426 |
| 2.031 | 0.595745 |
| 2.023 | 0.601064 |
| 2.01 | 0.606383 |
| 2.009 | 0.611702 |
| 1.994 | 0.617021 |
| 1.969 | 0.62234 |
| 1.956 | 0.62766 |
| 1.912 | 0.632979 |
| 1.907 | 0.638298 |
| 1.877 | 0.643617 |
| 1.876 | 0.648936 |
| 1.86 | 0.654255 |
| 1.819 | 0.659574 |
| 1.815 | 0.664894 |
| 1.809 | 0.670213 |
| 1.752 | 0.675532 |
| 1.747 | 0.680851 |
| 1.74 | 0.68617 |
| 1.718 | 0.691489 |
| 1.71 | 0.696809 |
| 1.703 | 0.702128 |
| 1.693 | 0.707447 |
| 1.656 | 0.712766 |
| 1.649 | 0.718085 |
| 1.617 | 0.723404 |
| 1.588 | 0.728723 |
| 1.588 | 0.734043 |
| 1.555 | 0.739362 |
| 1.509 | 0.744681 |
| 1.503 | 0.75 |
| 1.465 | 0.755319 |
| 1.459 | 0.760638 |
| 1.427 | 0.765957 |
| 1.41 | 0.771277 |
| 1.356 | 0.776596 |
| 1.346 | 0.781915 |
| 1.238 | 0.787234 |
| 1.203 | 0.792553 |
| 1.182 | 0.797872 |
| 1.134 | 0.803191 |
| 1.129 | 0.808511 |
| 1.1 | 0.81383 |
| 1.073 | 0.819149 |
| 1.048 | 0.824468 |
| 0.97 | 0.829787 |
| 0.962 | 0.835106 |
| 0.946 | 0.840426 |
| 0.942 | 0.845745 |
| 0.906 | 0.851064 |
| 0.857 | 0.856383 |
| 0.647 | 0.861702 |
| 0.584 | 0.867021 |
| 0.581 | 0.87234 |
| 0.567 | 0.87766 |
| 0.552 | 0.882979 |
| 0.534 | 0.888298 |
| 0.524 | 0.893617 |
| 0.518 | 0.898936 |
| 0.482 | 0.904255 |
| 0.424 | 0.909574 |
| 0.399 | 0.914894 |
| 0.397 | 0.920213 |
| 0.365 | 0.925532 |
| 0.358 | 0.930851 |
| 0.355 | 0.93617 |
| 0.349 | 0.941489 |
| 0.279 | 0.946809 |
| 0.254 | 0.952128 |
| 0.23 | 0.957447 |
| 0.228 | 0.962766 |
| 0.228 | 0.968085 |
| 0.169 | 0.973404 |
| 0.169 | 0.978723 |
| 0.16 | 0.984043 |
| 0.114 | 0.989362 |
| 0.101 | 0.994681 |
| 0.079 | 1 |

|  | ST 23F-Prevenar 13 (Day 300) |
| --- | --- |
| 97.084 | 0.00526316 |
| 72.286 | 0.0105263 |
| 68.286 | 0.0157895 |
| 62.251 | 0.0210526 |
| 62.18 | 0.0263158 |
| 61.656 | 0.0315789 |
| 56.876 | 0.0368421 |
| 43.072 | 0.0421053 |
| 36.092 | 0.0473684 |
| 34.797 | 0.0526316 |
| 34.473 | 0.0578947 |
| 33.673 | 0.0631579 |
| 28.776 | 0.0684211 |
| 27.642 | 0.0736842 |
| 26.17 | 0.0789474 |
| 22.387 | 0.0842105 |
| 22.323 | 0.0894737 |
| 21.902 | 0.0947368 |
| 21.324 | 0.1 |
| 21.142 | 0.105263 |
| 20.547 | 0.110526 |
| 18.719 | 0.115789 |
| 17.656 | 0.121053 |
| 16.944 | 0.126316 |
| 15.934 | 0.131579 |
| 15.905 | 0.136842 |
| 15.516 | 0.142105 |
| 15.2 | 0.147368 |
| 15.131 | 0.152632 |
| 14.369 | 0.157895 |
| 13.796 | 0.163158 |
| 12.749 | 0.168421 |
| 11.971 | 0.173684 |
| 11.472 | 0.178947 |
| 9.789 | 0.184211 |
| 8.987 | 0.189474 |
| 8.857 | 0.194737 |
| 8.099 | 0.2 |
| 8.093 | 0.205263 |
| 7.502 | 0.210526 |
| 7.446 | 0.215789 |
| 7.365 | 0.221053 |
| 6.918 | 0.226316 |
| 6.85 | 0.231579 |
| 6.485 | 0.236842 |
| 6.124 | 0.242105 |
| 5.944 | 0.247368 |
| 5.866 | 0.252632 |
| 5.702 | 0.257895 |
| 5.614 | 0.263158 |
| 5.597 | 0.268421 |
| 5.596 | 0.273684 |
| 5.526 | 0.278947 |
| 5.483 | 0.284211 |
| 5.355 | 0.289474 |
| 4.905 | 0.294737 |
| 4.885 | 0.3 |
| 4.873 | 0.305263 |
| 4.858 | 0.310526 |
| 4.783 | 0.315789 |
| 4.708 | 0.321053 |
| 4.524 | 0.326316 |
| 4.495 | 0.331579 |
| 4.486 | 0.336842 |
| 4.479 | 0.342105 |
| 4.471 | 0.347368 |
| 4.425 | 0.352632 |
| 4.41 | 0.357895 |
| 4.397 | 0.363158 |
| 4.297 | 0.368421 |
| 4.193 | 0.373684 |
| 4.143 | 0.378947 |
| 4.096 | 0.384211 |
| 4.086 | 0.389474 |
| 4.072 | 0.394737 |
| 4.052 | 0.4 |
| 3.994 | 0.405263 |
| 3.929 | 0.410526 |
| 3.838 | 0.415789 |
| 3.703 | 0.421053 |
| 3.687 | 0.426316 |
| 3.534 | 0.431579 |
| 3.531 | 0.436842 |
| 3.482 | 0.442105 |
| 3.418 | 0.447368 |
| 3.405 | 0.452632 |
| 3.357 | 0.457895 |
| 3.349 | 0.463158 |
| 3.292 | 0.468421 |
| 3.246 | 0.473684 |
| 3.244 | 0.478947 |
| 3.154 | 0.484211 |
| 3.109 | 0.489474 |
| 3.052 | 0.494737 |
| 3.047 | 0.5 |
| 3.042 | 0.505263 |
| 3.019 | 0.510526 |
| 2.934 | 0.515789 |
| 2.929 | 0.521053 |
| 2.924 | 0.526316 |
| 2.914 | 0.531579 |
| 2.911 | 0.536842 |
| 2.902 | 0.542105 |
| 2.899 | 0.547368 |
| 2.848 | 0.552632 |
| 2.844 | 0.557895 |
| 2.841 | 0.563158 |
| 2.795 | 0.568421 |
| 2.766 | 0.573684 |
| 2.765 | 0.578947 |
| 2.765 | 0.584211 |
| 2.764 | 0.589474 |
| 2.652 | 0.594737 |
| 2.594 | 0.6 |
| 2.581 | 0.605263 |
| 2.576 | 0.610526 |
| 2.561 | 0.615789 |
| 2.537 | 0.621053 |
| 2.496 | 0.626316 |
| 2.467 | 0.631579 |
| 2.462 | 0.636842 |
| 2.46 | 0.642105 |
| 2.411 | 0.647368 |
| 2.409 | 0.652632 |
| 2.399 | 0.657895 |
| 2.396 | 0.663158 |
| 2.335 | 0.668421 |
| 2.322 | 0.673684 |
| 2.29 | 0.678947 |
| 2.286 | 0.684211 |
| 2.279 | 0.689474 |
| 2.261 | 0.694737 |
| 2.227 | 0.7 |
| 2.174 | 0.705263 |
| 2.146 | 0.710526 |
| 2.125 | 0.715789 |
| 2.014 | 0.721053 |
| 1.99 | 0.726316 |
| 1.944 | 0.731579 |
| 1.904 | 0.736842 |
| 1.903 | 0.742105 |
| 1.844 | 0.747368 |
| 1.818 | 0.752632 |
| 1.765 | 0.757895 |
| 1.755 | 0.763158 |
| 1.652 | 0.768421 |
| 1.613 | 0.773684 |
| 1.605 | 0.778947 |
| 1.54 | 0.784211 |
| 1.529 | 0.789474 |
| 1.497 | 0.794737 |
| 1.496 | 0.8 |
| 1.493 | 0.805263 |
| 1.451 | 0.810526 |
| 1.437 | 0.815789 |
| 1.413 | 0.821053 |
| 1.398 | 0.826316 |
| 1.374 | 0.831579 |
| 1.3 | 0.836842 |
| 1.27 | 0.842105 |
| 1.264 | 0.847368 |
| 1.186 | 0.852632 |
| 1.066 | 0.857895 |
| 0.775 | 0.863158 |
| 0.707 | 0.868421 |
| 0.693 | 0.873684 |
| 0.684 | 0.878947 |
| 0.666 | 0.884211 |
| 0.65 | 0.889474 |
| 0.555 | 0.894737 |
| 0.511 | 0.9 |
| 0.48 | 0.905263 |
| 0.477 | 0.910526 |
| 0.471 | 0.915789 |
| 0.464 | 0.921053 |
| 0.457 | 0.926316 |
| 0.433 | 0.931579 |
| 0.353 | 0.936842 |
| 0.249 | 0.942105 |
| 0.241 | 0.947368 |
| 0.231 | 0.952632 |
| 0.225 | 0.957895 |
| 0.202 | 0.963158 |
| 0.174 | 0.968421 |
| 0.159 | 0.973684 |
| 0.119 | 0.978947 |
| 0.112 | 0.984211 |
| 0.109 | 0.989474 |
| 0.069 | 0.994737 |
| 0.06 | 1 |
